# Cohort profile for the creation of the SAIL MELD-B e-cohort (SMC) and SAIL MELD-B children and Young adult e-cohort (SMYC)

**DOI:** 10.1101/2024.04.22.24306168

**Authors:** Roberta Chiovoloni, Jakub Dylag, Nisreen A Alwan, Ann Berrington, Michael Boniface, Nic Fair, Emilia Holland, Rebecca B Hoyle, Mozhdeh Shiranirad, Sebastian Stannard, Zlatko Zlatev, Rhiannon K Owen, Simon DS Fraser, Ashley Akbari

## Abstract

**Purpose:** We have established the SAIL MELD-B electronic cohort (e-cohort SMC) and the SAIL MELD-B children and Young adults e-cohort (SMYC) as a part of the Multidisciplinary Ecosystem to study Lifecourse Determinants and Prevention of Early-onset Burdensome Multimorbidity (MELD-B) project. Each cohort has been created to investigate and develop a deeper understanding of the lived experience of the ‘burdensomeness’ of multimorbidity by identifying new clusters of burdensomeness indicators, exploring early life risk factors of multimorbidity and modelling hypothetical prevention scenarios.

**Participants:** The SMC and SMYC are longitudinal e-cohorts created from routinely-collected individual-level population-scale anonymised data sources available within the Secure Anonymised Information Linkage (SAIL) Databank. They include individuals with available records from linked health and demographic data sources in SAIL at any time between 1^st^ January 2000 and 31^st^ December 2022. The SMYC e-cohort is a subset of the SMC, including only individuals born on or after the cohort start date.

**Findings to date:** The SMC and SMYC cohorts include 5,180,602 (50.3% female and 49.7% male) and 896,155 (48.7% female and 51.3% male) individuals respectively. Considering both primary and secondary care health data, the five most common long-term conditions for individuals in SMC are ‘Depression’, affecting 21.6% of the cohort, ‘Anxiety’ (21.1%), ‘Asthma’ (17.5%), ‘Hypertension’ (16.2%) and ‘Atopic Eczema’ (14.1%), and the five most common conditions for individuals in SMYC are ‘Atopic Eczema’ (21.2%), ‘Asthma’ (11.6%), ‘Anxiety’ (6.0%), ‘Deafness’ (4.6%) and ‘Depression’ (4.3%).

**Future plans:** The SMC and SMYC e-cohorts have been developed using a reproducible, maintainable concept curation pipeline, which allows for the cohorts to be updated dynamically over time and manages for the request and processing of further approved long-term conditions and burdensomeness indicators extraction. Best practices from the MELD-B project can be utilised across other projects, accessing similar data with population-scale data sources and trusted research environments.

**STRENGTHS AND LIMITATIONS OF THIS STUDY:**

- SMC and SMYC are representative of the Welsh population.
- Anonymised cohorts serve as an effective strategy for overcoming consent-related barriers, enabling seamless data aggregation and analysis.
- The creation of a reproducible concept curation pipeline to manage and process data extraction for the e-cohorts enables efficient delivery of datasets in support of multiple research questions and outcomes.
- Routine data does not capture data on important aspects such as quality of life.
- Routine data can be subject to missing data or errors.
- Lack of coverage of burdensomeness indicators in routine data.

## 1. INTRODUCTION

The prevalence of Multiple Long-Term Condition Multimorbidity (MLTC-M), commonly defined as the co-occurrence of two or more chronic conditions in an individual, has increased in many regions of the world as a result of many factors, including changes in lifestyles, the ageing population and increasing diagnosis of long-term conditions (LTCs).[1]

In the United Kingdom (UK), it is estimated that more than half of the population aged 65 and above suffers from two or more LTCs, and it is predicted that by 2035 two thirds of people aged over 65 will experience MLTC-M [2].

MLTC-M is often a burden for patients, their carers and their health service providers. It is associated with reduced quality of life [3], fragmented and costly care [4,5], polypharmacy [6–8], physiological distress, extended hospital stays [9,10], increased mortality [11] and it substantially contributes to healthcare inefficiency and cost in both primary and secondary care settings [12–15].

However, to date, different aspects of MLTC-M are not well understood [16]. For example, most MLTC-M studies have focused only on a selected subset of the population, specifically older individuals in high income countries [17], a small number of conditions [18], and the analysis of clustering of conditions in repeated cross-sectional studies [19–24].

There is limited research examining the association between MLTC-M, socioeconomic status and longitudinal trends [25–27], and limited evidence regarding other social and behavioural determinants that could be fundamental in the emergence and evolution of less common MLTC-M patterns [28]. Additionally, few studies investigate how the timing and nature of exposure to risk factors influence the accrual of LTCs [27,29–33], and little research focuses on how to prevent MLTC-M development [34].

The Multidisciplinary Ecosystem to study Lifecourse Determinants and Prevention of Early-onset Burdensome Multimorbidity (MELD-B) collaboration aims to address some of these key gaps in the evidence in MLTC-M research by developing a deeper understanding of the lived experience of ‘burdensomeness’ of multimorbidity, identifying new clusters of burdensome MLTC-M and their key early-life risk factors, mapping trajectories across the lifecourse towards burdensome clusters in those under 65, and modelling prevention scenarios to inform policy, see [35]. These will be achieved through the analysis of birth cohorts and routinely-collected electronic health record (EHR) data sources, using a combination of Artificial-Intelligence (AI)-enhanced epidemiological analysis and statistical methods. For a more detailed description of the MELD-B objectives and structure, see [35–37].

The use of routine-collected EHR data presents a number of challenges: i) routinely-collected EHR data are primarily collected for clinical and administrative purposes rather than supporting research, ii) there is often incomplete and/or inaccurate data, which may not be harmonised and standardised across data sources. However the MELD-B project recognises the importance of using large-scale EHR data sources, such as the Secure Anonymised Information Linkage (SAIL) Databank and the Clinical Practice Research Datalink (CPRD). These data sources offer large sample sizes, long follow-up periods and include a wide range of study variables and generalizable populations.

To support the MELD-B project, we have created the SAIL MELD-B e-cohort (SMC) and the SAIL MELD-B children and Young adult e-cohort (SMYC), longitudinal population-based e-cohorts based in Wales. The e-cohorts are representative of the wider population in terms of sex, age and socioeconomic deprivation.

The e-cohorts are developed to support multiple research questions within the MELD-B work packages and collaboration. They will be used as maintainable research ready data assets (RRDA) enabling the MELD-B collaboration to perform clustering, sequencing and statistical analyses to identify the critical time-points for public health intervention [38].

## 2. COHORT DESCRIPTION

The SMC is a longitudinal e-cohort defined using routinely collected anonymised linked demographic, administrative and EHR data sources available within the SAIL Databank [39]. The SMYC is a subset of the SMC, including only individuals born after the study start date with demographic data available before 18 years of age, and with consistent maternal records.

Using longitudinal e-cohorts as available RRDAs to study MLTC-M will not only allow the evaluation of the burden of MLTC-M on individuals but also provide insights into the wider determinants of MLTC-M, the temporal dynamics of disease and burden progression, and potential effects of intervention and prevention.

The inclusion of newborns and young individuals in the e-cohort will allow us to better understand how social, biological and environmental factors in early life contribute to the risk of developing MLTC-M, as there is substantial evidence indicating the critical role of early life in determining health during childhood and adulthood [40–44].

The MELD-B co-investigators derived burdensomeness concepts from a qualitative evidence synthesis with extensive patient and public involvement. Extracting health service interactions and records from routine data can provide measurable observations for the derived concepts for individuals in SMC and SMYC. This will offer fundamental insights into measuring and conceptualising burdensome MLTC-M.

All codes and scripts used in this study are available for others to access here: https://github.com/SwanseaUniversityDataScience/1377-MELD_B-CohortCuration

### 2.1 SAIL Databank and data sources

The SAIL Databank (www.saildatabank.com) contains anonymised, encrypted, routinely collected individual-level population-scale linkable data sources for all Welsh residents using any National Health Service (NHS) UK-wide services, and any individuals residing outside of Wales using NHS Wales services. To ensure anonymity and confidentiality, each individual is assigned a unique identifier (Anonymised Linking Field, ALF), used to link together different data sources at the individual level. The ALF is generated through a double encryption process: Digital Health and Care Wales (DHCW) uses NHS number or a combination of unique demographic information (such as sex, name, date of birth) to generate a unique identifier, which is then further encrypted within the SAIL Databank. This process ensures that no single organisation can decrypt the records, making SAIL a TRE for record-linkage studies [45–47].

To build the SMC and SMYC, we linked demographic and mortality data sources: the Welsh Demographic Service Dataset (WDSD), the Annual District Death Extract (ADDE) from the Office for National Statistics (ONS) mortality register, and the Annual District Birth Extract (ADBE) from the ONS birth register, the National Community Child Health database (NCCH) and the Maternal Indicators DataSet (MIDS), see Table 1.

**Table 1:**
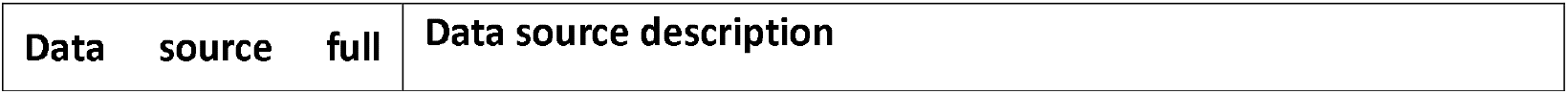
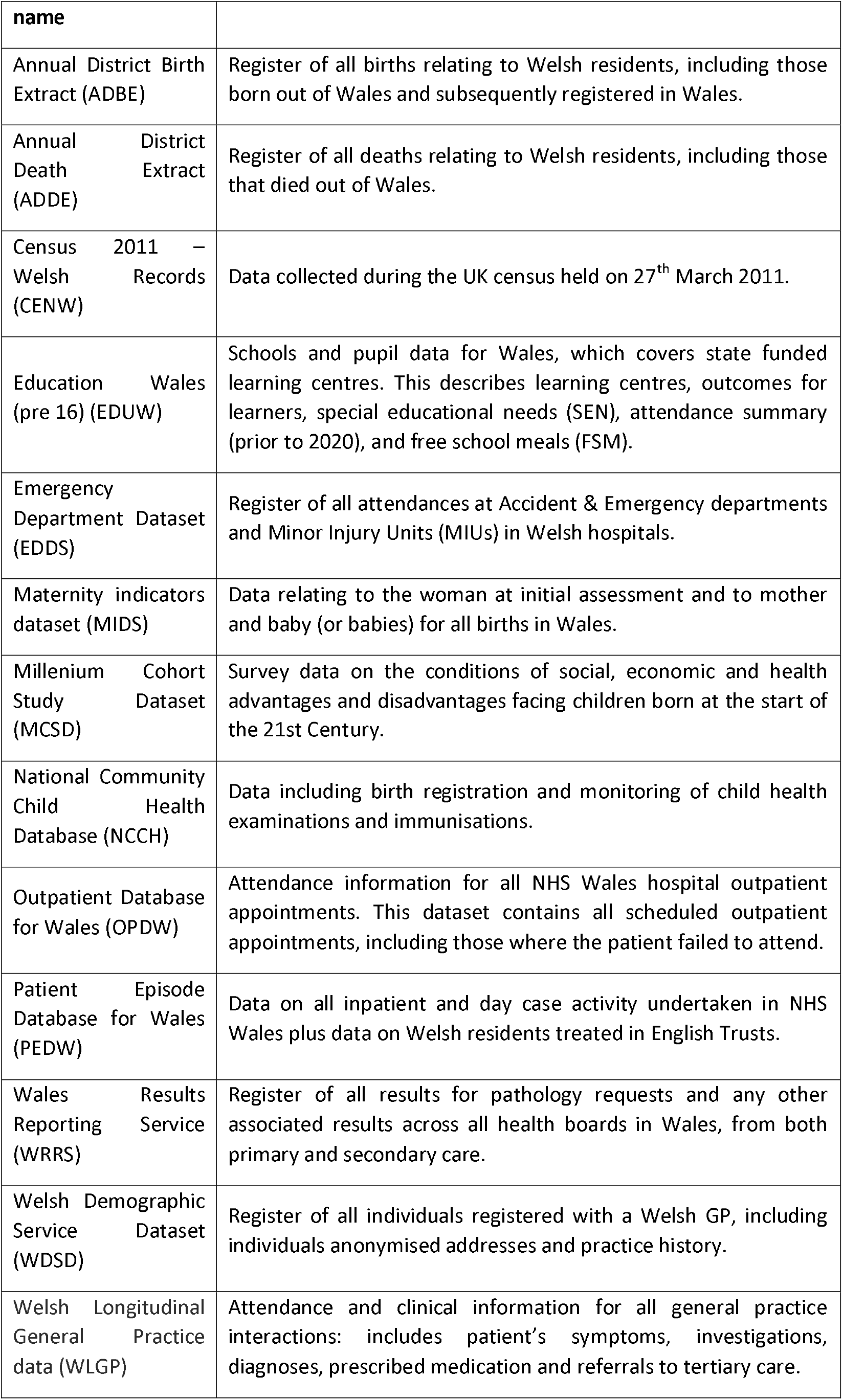
Data sources available to the MELD-B project. For more detailed information on the data sources, see [51].

The baseline demographic characteristics of the e-cohorts include: ALF, Sex (male or female), Week of Birth (WOB), Date of Death (DOD) where applicable, Ethnic group [48], Lower-layer Super Output Area, 2011 version (LSOA 2011) and Welsh Index of Multiple Deprivation, 2019 version (WIMD 2019). The last two provide insights on the socioeconomic status of the individuals at an area level: LSOAs are small areas containing around 1,500 individuals used to link individual records to the WIMD 2019 to derive deprivation status.

Ethnic groups have been classified using two different classifications, the ONS and the New and Emerging Respiratory Virus Threats Advisory Group (NER) classifications, which have five and nine ethnicity categories respectively.

The health data sources available to the MELD-B project include, the Welsh Longitudinal General Practice (WLGP) data, the Patient Episode Database for Wales (PEDW), the Emergency Department Dataset (EDDS), the Outpatient Database for Wales (OPDW) and the Welsh Results Reports Service (WRRS), the National Community Child Health Database (NCCH) and the Maternity indicators dataset (MIDS) see Table 1.

Currently WLGP contains primary care data for 86% of the Welsh population registered with a General Practice (GP) and 80% of GP practices covering all local authorities in Wales [49]. In Wales, primary care general practice data are recorded using Read V2 codes, while data for secondary care episodes, such as hospital admissions, are recorded using the International Classification of Disease v10 (ICD-10) and the Office of Population Censuses and Surveys codes v4 (OPCS-4). Emergency department data has its own coding system (see [50]).

Data are available from different data sources at different times, and their quality improves over time. Given the requirements of the study and the completeness of the data sources, 1^st^ January 2000 was chosen as the start date of the study and 1^st^ January 1990 as the start date of data collection. Note where data sources start after these dates, their coverage begins from the respective data source’s start date, see [51].

### 2.2 Cohorts Design

The SMC is a longitudinal population e-cohort including all people residing in Wales and registered with a Welsh GP between 1^st^ January 2000 (identified as *cohort start date*) and 31^st^ December 2022^1^ (identified as *cohort end date*), and it provides a generalisable population sample to the population of Wales with respect to sex, age and socioeconomic deprivation [52].

The primary data source used to build the SMC is WDSD.

As a longitudinal e-cohort, the number of individuals will change throughout the study as they can leave or join the e-cohort at any time during the study period. SMC entries include all residents in Wales who meet *all* of the following conditions:

a. They have a recorded sex at birth (Male/Female) in either ADBE or WDSD.
b. They have a recorded date of death after the 1^st^ January 2000, or without a recorded date of death.
c. They have a recorded date of birth before 31^st^ December 2022.
d. They are less than 105 years of age on 1^st^ January 2000.
e. They have WDSD and WLGP data available over the same period of time. The SMYC cohort is a subset of the SMC cohort, including only:
f. Individuals born between 1^st^ January 2000 and 31^st^ December 2021^2^
g. Individuals with both demographic and healthcare data available before they turn 18 years of age.
h. Individuals with *consistent* maternal records (an individual has a *consistent* maternal record if they can be linked to *at most* one mother, see Sec 2.2.1).

The cohort entry date is defined as the date an individual enters the cohort and is identified by the first date the individual is registered in the WDSD.

Cohort censorship was defined by the earliest of:

a. Death.
b. Migration outside of Wales or break in their residency data.
c. Study endpoint on 31^st^ December 2022.

Note that once an individual meets one of the censorship criteria, they are not allowed to re-enter the e-cohort, see Appendix A for further details.

To identify all relevant health events recorded in the routinely collected EHR data, we linked individuals in both cohorts to the available data sources. While primary care data are available for all cohort participants, as it is a prerequisite for cohort membership, secondary care and pathology data might not be available for everyone if they have not utilised these services during the study period. The Upset plots in Figure 4 and Figure 5 quantitatively represent individuals within the SMC and SMYC and their interactions with various healthcare settings throughout their cohort membership.

#### 2.2.1 Maternal records study to identify SMYC

A necessary condition for being part of the SMYC is to have a consistent maternal record (MAT_ALF) within the NCCH and MIDS data sources.

An individual has a consistent maternal record if he or she can be linked to at most one mother, i.e. if:

1. The individual has no MAT_ALF in either NCCH or MIDS.
2. The individual has one MAT_ALF either in NCCH or MIDS.
3. The individual has two or more MAT_ALF in both NCCH and MIDS, and they match each other.

When none of the above conditions apply, then the individual is excluded from the SMYC.

Note that if a person has an available maternal record but the mother is not included in SMC, we considered this individual as *not* having a maternal record (n = 1248, 0.1%) thus, all the MAT_ALF linked to the SMYC form a subset of the SMC.

After selecting individuals from SMC who meet the SMYC conditions stated above (896,763), we extracted the maternal record(s) available in multiple data classes within NCCH and MIDS^3^: 809,616 (90.3%) individuals have at least 1 maternal record, with 223,310 (24.9%) of them having records in both NCCH and MIDS. 608 (< 0.001%) among these are linked to more than one MAT_ALF, i.e. have inconsistent maternal records. These individuals are excluded from the SMYC cohort.

## 2.3 Concept curation pipeline

MELD-B has defined a set of burdensomeness concepts, which can be identified and characterised in routinely collected EHR data^4^ to better understand how living with MLTC-M affects people’s lives and to apply this knowledge to inform data curation and extraction. These concepts include long-term conditions and indicators of the work associated with them such as symptoms, emotions, indicators of financial stress, and observable and measurable information relevant to health or healthcare, such as medical diagnoses, blood tests, appointments, hospital admissions and number of medications.

We implemented a reproducible concept curation pipeline to define, approve, process and import the identified burdensomeness concepts inside SAIL. This pipeline facilitates the extraction of relevant data associated with the various concepts identified by the MELD-B clinical group from the available linked data sources in SAIL. The pipeline and its outcomes are documented and managed outside of SAIL. It is accessible to all team members to ensure transparency of the process and facilitate collaboration. In Figure 1, we provide a visualisation of the pipeline.

**Figure 1:**
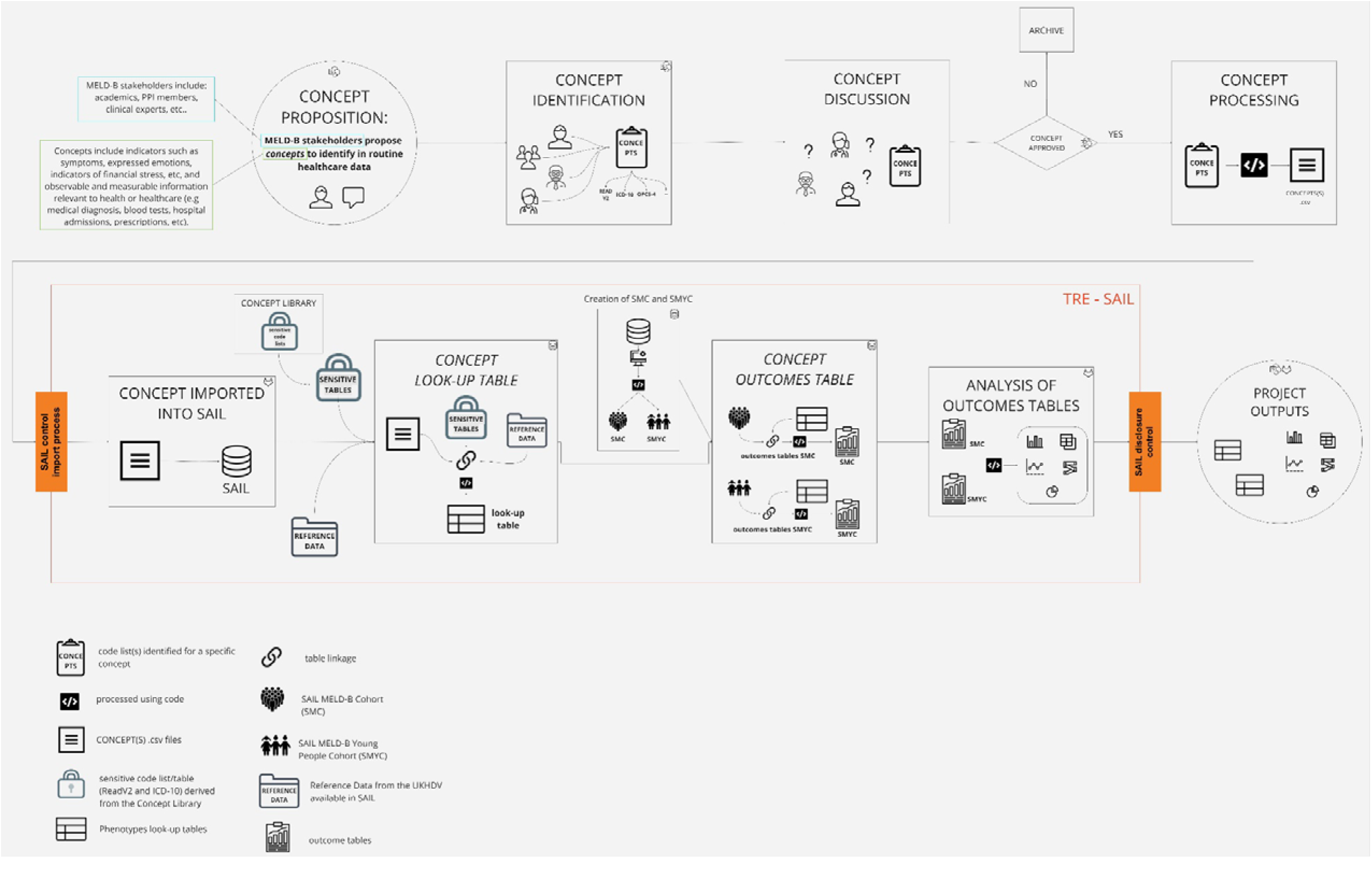
Concept curation pipeline to extract relevant data for SMC and SMYC.

The first step of the pipeline is the proposition of each concept: clinicians, with a deep understanding of the phenomena of burden and their representation in routine data, can propose a concept that they believe embodies the idea of burden.

For the proposed concept to be considered and approved, the proposal must be accompanied by a published or open-source code list to review and agree on or a list of requirements that defines the concept, which can be used to derive a code list. A code list is a collection of classification codes associated with a specific concept of interest. Each code list includes only codes associated with a specific classification (e.g. SNOMED, Read V2, ICD-10, OPCS-4) therefore, it is possible to identify one or more code lists for each concept.

All proposed concepts and their associated code lists are then reviewed and discussed by the clinical group and, if approved, included in the study. If the concept proposed is not deemed relevant, it is archived. The approved concepts and their associated code lists are first processed to ensure their formatting, structure and content are available in a machine-readable output file, and then processed into a standardised format [53]Subsequently the output code list can be utilised with routinely collected data sources and imported into the SAIL Databank.^5^

As part of the import and implementation process of the pipeline, all concepts and their associated code lists are cross-referenced with the SAIL sensitive code lists. Any sensitive codes are excluded from the concepts, so they will not be used or included for data extraction or used in project outputs. The current lists of all known sensitive ICD-10 and ReadV2 codes, which are based on a combination of sensitive codes provided by Digital Health and Care Wales (DHCW), a published list of sensitive codes for England and any other code flagged as sensitive by the SAIL team^6^, are downloaded from the Concept Library [54] and processed as part of the pipeline.

Linking the imported concepts’ code lists with the sensitive code tables and the relevant tables available in SAIL^7^ we create look-up tables for each concept and each concept coding classification, see Suppl. Material 2 for an example. Linking the *look-up* tables to SMC and SMYC and the relevant data sources available in SAIL, we extract the relevant records for all the individuals in the cohort in *outcome tables*. Each of these tables contains *all* available records for a specific concept and each applicable concept-specific coding classification for *all* the individuals in SMC and SMYC. The outcome tables can then be used for analysis.

The pipeline allows us to re-run concepts efficiently and reproducibly, facilitating the data extraction and the data linkage with SMC and SMYC. In particular, this involves creating newer versions of look-up and outcomes tables with version control when existing concepts are updated, new concepts are provided and approved, and new and updated versions of the routinely collected data are available. Given the broad applicability of this pipeline, this approach can be shared with other projects, both in SAIL and in other TREs, as a transferable and reproducible method to be implemented.

### 2.3.1. MELD-B initial set of MLTC-M indicators

The MELD-B clinical domain expert group proposed an initial set of long-term conditions based on the list agreed upon by the NIHR AIM Research Consortia [55], existing literature and project requirements [56–59].

The initial set of concepts proposed by the group consists of 84 long-term conditions, and the full list can be found in Suppl. Table 1^8^ .

As an objective of the MELD-B Research Collaboration is to characterise the ‘burden’ of living with MLTC-M, additional long-term conditions and a set of burden concepts are being proposed by the clinical team and once reviewed, will be processed through the concept pipeline. The identification of burden concepts to be included in the study is ongoing work, and may be limited in certain concepts due to the capture and availability of information related to lived experience observations in EHR data. The concepts version used as the basis for cohort analysis in this paper is v2.2.4 [60].

The ReadV2 and ICD-10 code lists associated with the initial 83 concepts proposed have been extracted from [59] and the supplementary file provided by the authors in [58]^9^. The MELD-B clinical group reviewed and approved all 82 ReadV2 and 69 (84.1%) of the ICD-10 code lists identified for the 83 concepts. The total number of medical codes identified andapproved is 7,503: 5,987 (81.7%) ReadV2 and 1,516 (18.3%) ICD-10 codes. 109 ReadV2 and 16 ICD-10 codes are flagged as sensitive and are therefore not included in the outcomes tables created in SAIL or any of the descriptive analyses performed, see Sec.3.

The look-up tables created in the first stage of the MELD-B project are available in Suppl.Material 2.

## 3. Findings to date

### 3.1. Sociodemographic characteristics

The SMC and SMYC e-cohorts have been designed to provide a generalisable population sample to be used to answer different research questions. From the 5,475,154 individuals available longitudinally within the WDSD, 5,180,610 individuals met the SMC inclusion criteria described in Sec. 2.2, and 896,155 individuals met the inclusion criteria to be included in the SMYC, see Figure 2.

**Figure 2:**
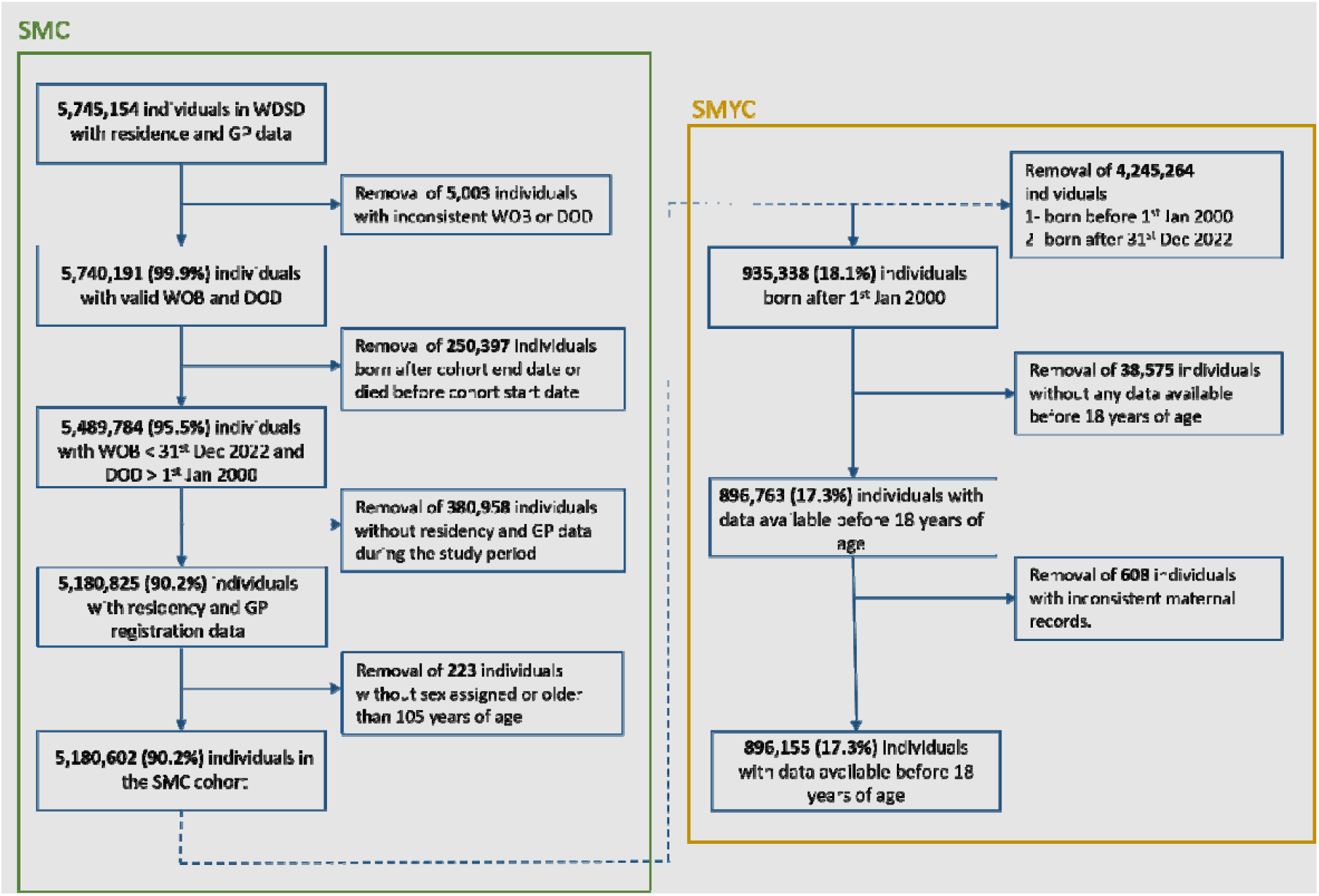
SAIL MELD-B consort diagram based on inclusion criteria. SMC = SAIL MELD-B e-cohort, SMYC = SAIL MELD-B Young cohort.

The follow-up period is defined as the time an individual spends in the e-cohort, with a minimum follow-up of one day and a maximum of 23 years. The number of individuals with full coverage is 1,731,280 (33.4%) for SMC and 47,500 (5.3%)^10^for SMYC, see Table 2.

**Table 2:** SMC and SMYC baseline demographic information. The percentage refers to the total number of individuals in each cohort.Note that WIMDs are not available for SMYC as they are computed on the 1^st^ January 2000, and SMYC only includes individuals born on or after this date.

| **<u>Baseline demographics</u>** | **<u>SMC</u>** | **<u>SMYC</u>** |
| --- | --- | --- |
| *Cohort size* | **5,180,602 (100%)** | **896,155 (100%)** |
| *Male (%)* | 2,575,867 (49.7%) | 459,644 (51.3%) |
| *Female (%)* | 2,604,735 (50.3%) | 436,511 (48.7%) |
| *Cohort size at cohort start* | 2,990,123 (57.8%) | N/A |
| *Mean age in years at cohort start* | 39.7 years | N/A |
| **WIMD quintile at cohort start** |  |  |
| - WIMD = 1 (most deprived) | 615,012 (11.9%) | N/A |
| - WIMD = 2 | 600,128 (11.6%) | N/A |
| - WIMD = 3 | 608,952 (11.8%) | N/A |
| - WIMD = 4 | 589,163 (11.4%) | N/A |
| - WIMD = 5 (least deprived) | 575,782 (11.1%) | N/A |
| **Cohort exit reason** |  |  |
| - Death | 747,927 (14.4%) | 1,614 (0.2%) |
| - Loss to follow-up | 1,420,930 (27.4%) | 143,836 (16.1%) |
| - Study end point | 3,011,745 (58.1%) | 750,705 (83.7%) |
| **Ethnic group (NER code)** |  |  |
| 1. White | 3,654,965 (70.6%) | 715,839 (79.9%) |
| 2. Mixed | 55,150 (1.1%) | 27,434 (3.1%) |
| 3. Indian | 37,543 (0.7%) | 8,525 (1.0%) |
| 4. Pakistani | 20,404 (0.4%) | 6,022 (0.7%) |
| 5. Bangladeshi | 107,146 (2.1%) | 28,849 (3.2%) |
| 6. Chinese | 29,316 (0.6%) | 2,549 (0.3%) |
| 7. Black Caribbean | 7,803 (0.2%) | 914 (0.1%) |
| 8. Black African | 28,372 (0.5%) | 8,932 (1.0%) |
| 9. Other | 99,677 (1.9%) | 26,252 (2.9%) |
| 10. Unknown ethnicity | 1,140,226 (22%) | 70,839 (7.8%) |

| Follow-up period |  |  |
| --- | --- | --- |
| - 1 year | 354,157 (6.8%) | 72,301 (8.1%) |
| - 1 – 3 years | 670,798 (12.9%) | 115,148 (12.8%) |
| - 3-5 years | 387,791 (7.5%) | 90,087 (10.1%) |
| - 5 – 10 years | 663,472 (12.8%) | 200,183 (22.3%) |
| - 10 – 15 years | 567,058 (10.9%) | 183,958 (20.5%) |
| - 15-20 years | 532,310 (10.3%) | 168,030 (18.7%) |
| - 20 years | 2,005,016 (38.7%) | 66,448 (7.4%) |

**Table 3:** Number of individuals that join the cohort 1, 2, 3 and 4+ times.

| N periods | N individuals |
| --- | --- |
| 1 | 4,828,060 (93.2%) |
| 2 | 323,630 (6.3%) |
| 3 | 25,860 (0.5%) |
| 4+ | 3,060 (<0.1%) |

Ethnic group records are available for 78% of the individuals in SMC and 92.2% of individuals in SMYC. In both cohorts, the predominant ethnicities are “White” (70.6% and 79.9%), followed by “Bangladeshi” (2.1% and 3.2%) and “Mixed” (1.1% and 3.1%).

In Table 2, we summarise the demographic information for SMC and SMYC.

The distribution of SMC at cohort inception by age groups and sex is visualised in Figure 3.

**Figure 3:**
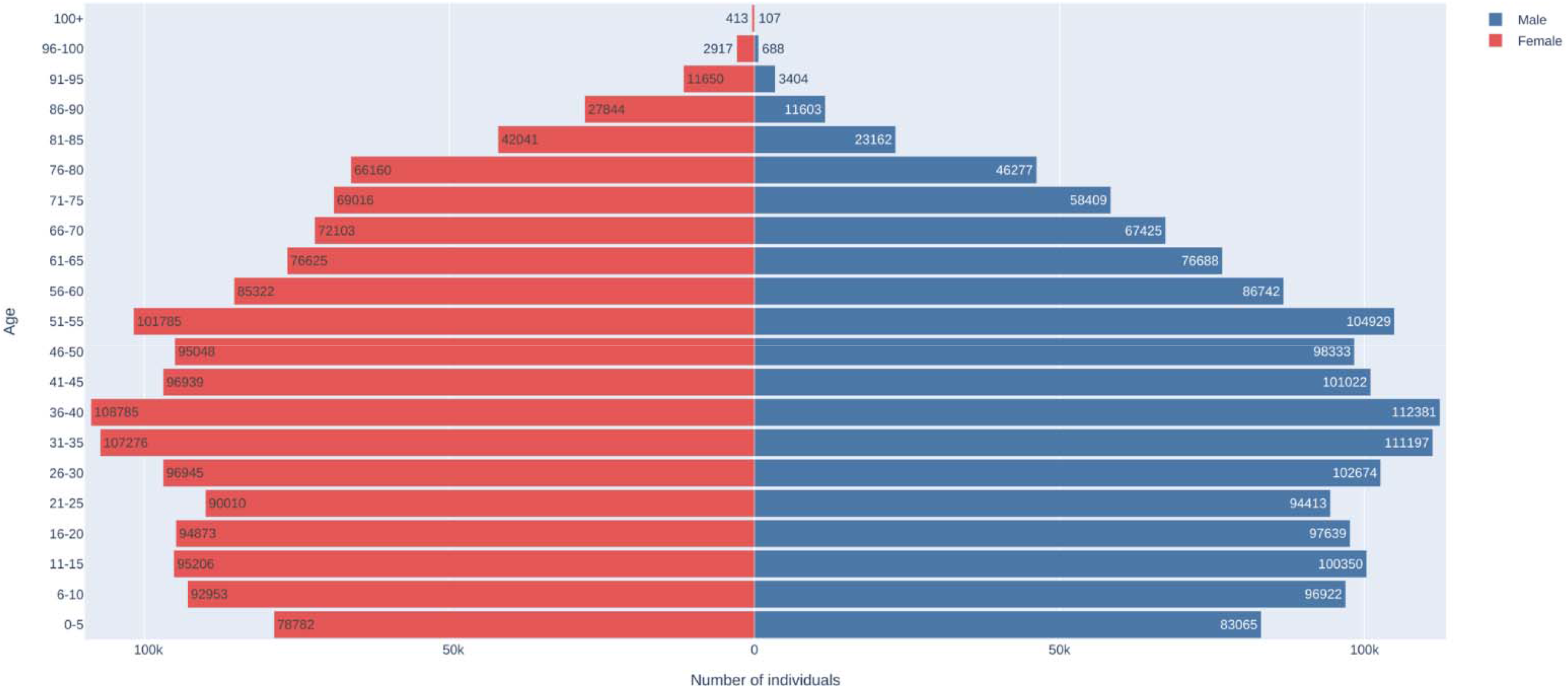
Pyramid plot of SMC at cohort start date.

To provide a quantitative representation of individuals within the cohorts and their interactions with various healthcare settings throughout the cohort membership, we produced the Upset plots in Figure 4 and Figure 5. Almost 37% of the individuals in the SMC have records in all the routinely collected EHR data sources available to the MELD-B project. In total, 66.4% and 67.9% of individuals used inpatient and outpatient services, respectively, while only 48% of SMC used emergency department services.

**Figure 4:**
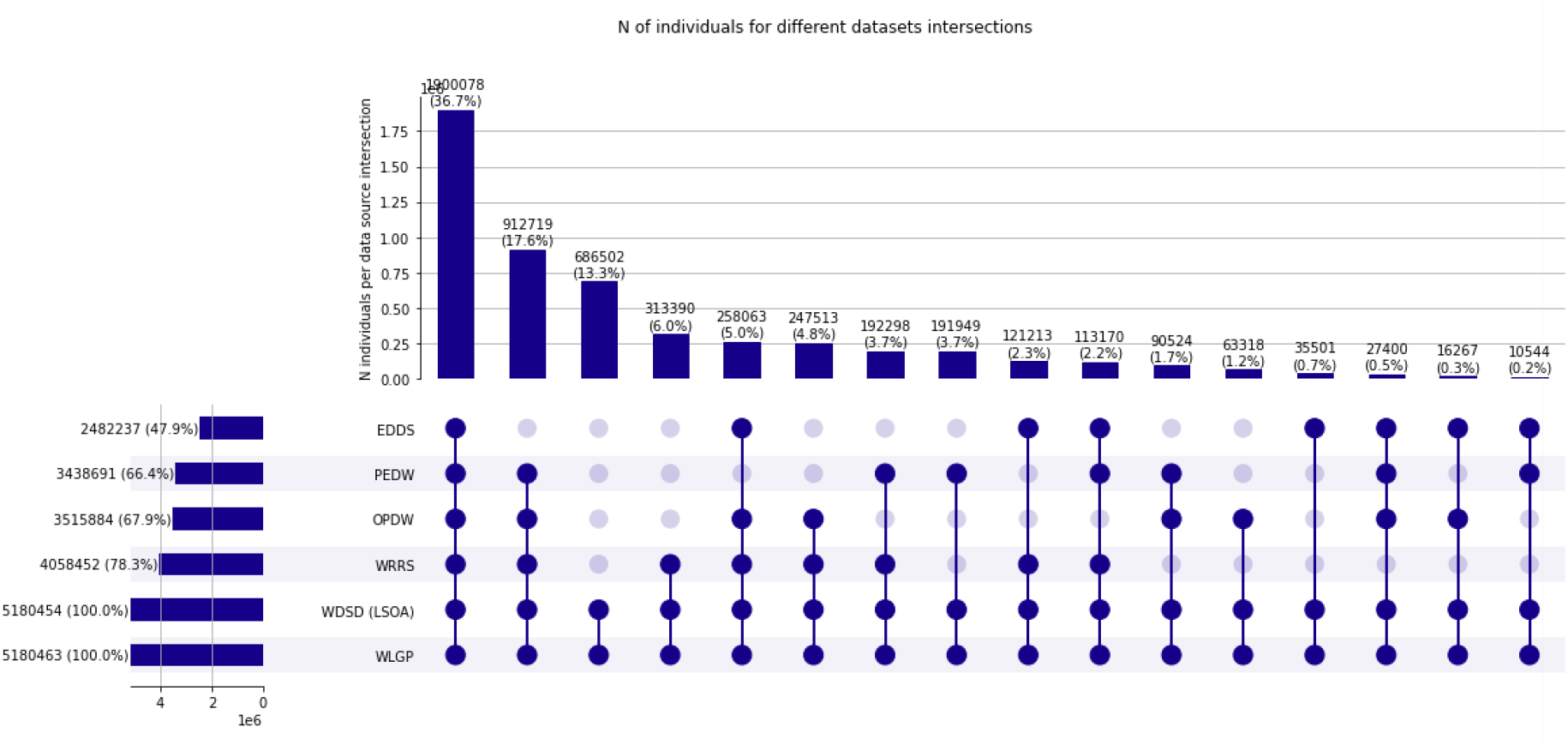
Number of SMC individuals using healthcare services recorded in multiple datasources.

**Figure 5:**
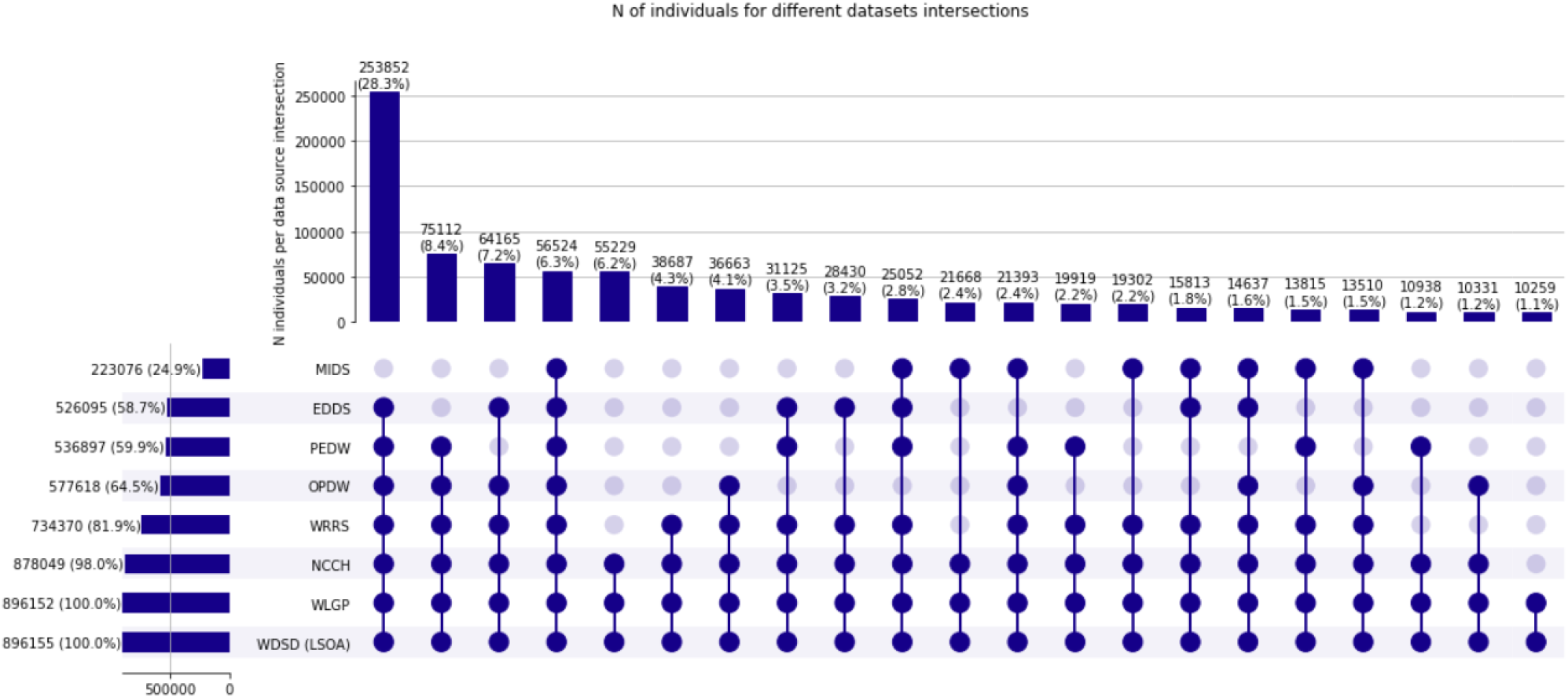
Number of SMYC individuals using healthcare services recorded in multiple datasources.

The SMYC upset plot includes NCCH and MIDS in addition to the routinely collected EHR data sources. Almost every individual (98%) has at least one record in the NCCH data source, see also Sec 2.2.1, and 59.9%, 64.5% and 58.7% of individuals can be linked to inpatient secondary care, outpatient secondary care and emergency data sources respectively.

#### 3.1.1 Cohorts evolution over the cohort period

To better understand how SMC and SMYC evolved over the study period, we collected demographic information on the 1^st^ January of every year during the cohort study period (e.g. total number of individuals, sex ratio, number of individuals leaving and/or joining the e-cohort, etc.), see Suppl.Table 1 and Suppl.Table 2 for more details ^11^.

The total number of individuals in the SMC increases until 2010, when the e-cohort includes 3,089,310 individuals. It decreases after this year and reaches its new minimum in 2022 (3,013,498). The female/male ratio steadily reduces from 2000 to 2017, reaching a minimum value of 0.995, and then increases again from 2016 to 2022. In absolute terms, women outnumber men from 2000 to 2011. The number of individuals in SMYC increases over the cohort period, reaching its maximum in 2022 with 720,500 people. Differently from SMC, the ratio of female/male is always less than one, see Figure 6.

**Figure 6:**
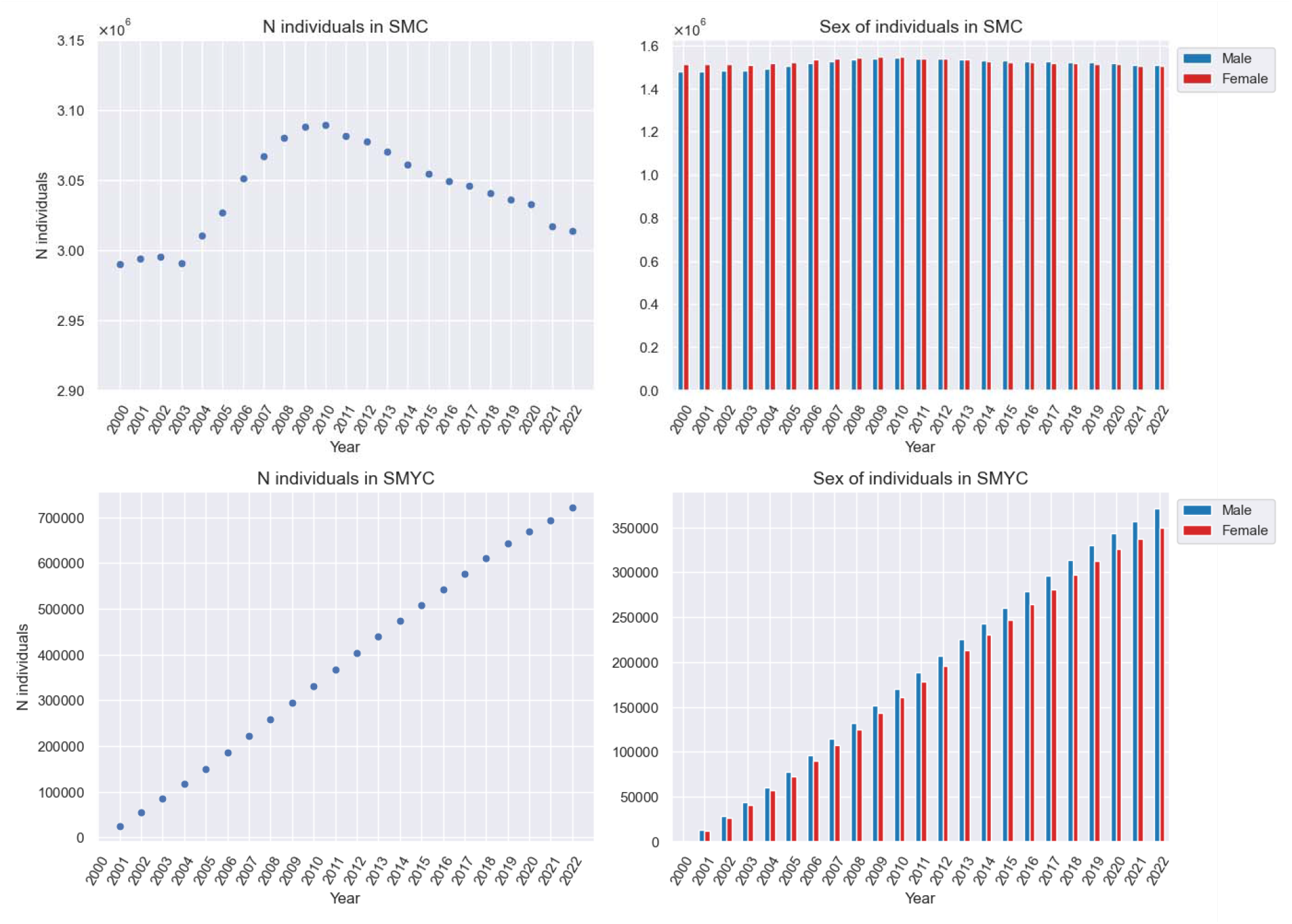
On the left total number of individuals on the 1st January of each year during the study period. On the right, the number of females/males on the 1st January of every year during the study period for SMC and SMYC.

Between 2002 and 2009, the number of individuals joining SMC is larger than the number of people leaving it, see Figure 7. This trend is reversed from 2010 to 2022 when, for the first time in ten years, the number of people leaving SMC is again greater than the number joining it. The year with the biggest gap between cohort joiners and cohort leavers is 2005 (104,258 vs 79,923). The year with the smallest number of joiners is 2020 (74,609). This dip is likely attributed to the impact of the COVID-19 pandemic and the resulting decline in the university student population.

**Figure 7:**
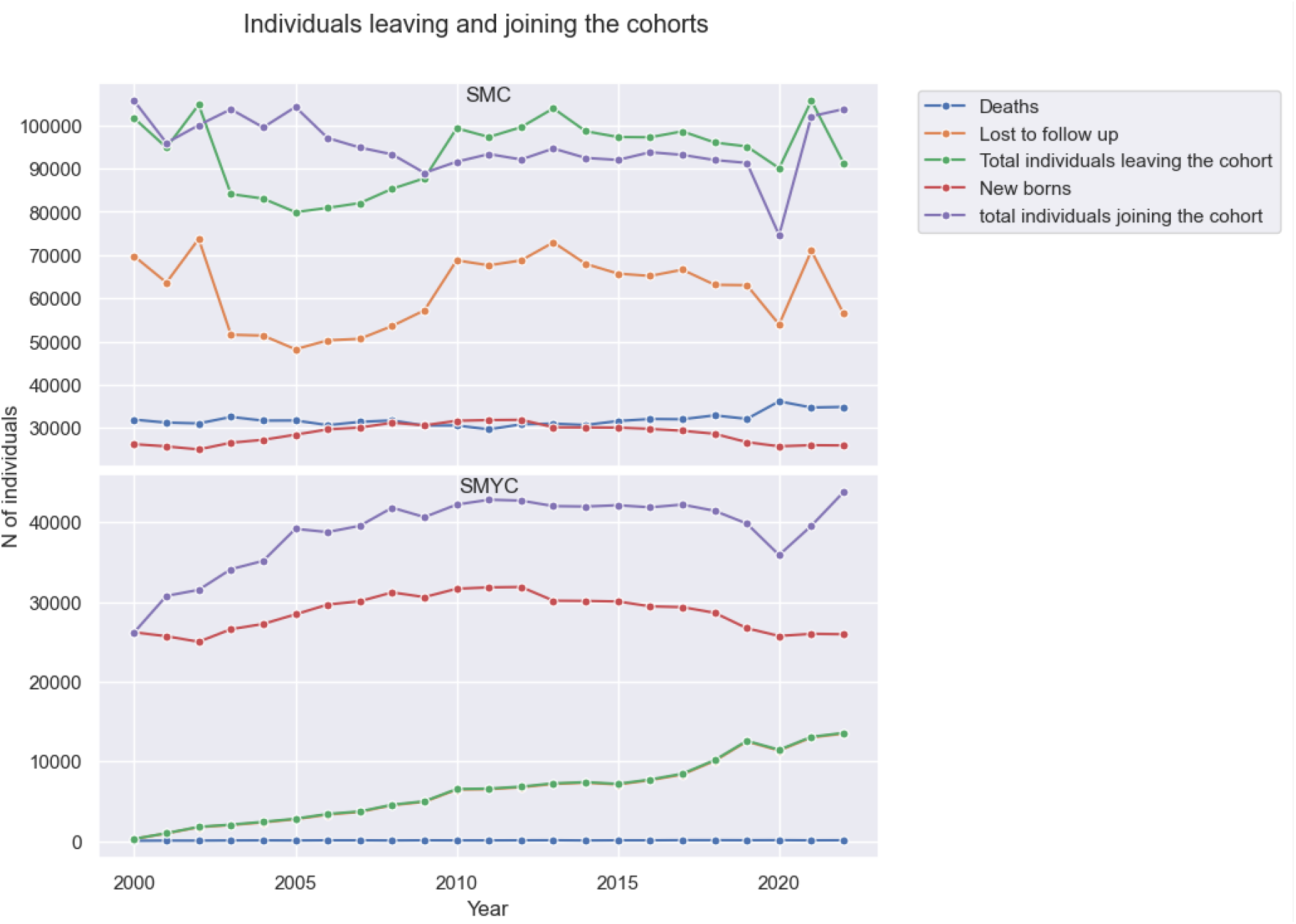
Individuals leaving and joining SMC and SMYC each year. Note that in the SMYC plot, the “Lost to follow-up” line almost coincides with the “total individuals leaving the cohort” line.

In SMYC, the number of individuals joining the cohort is always larger than the number of individuals leaving it, see Figure 7. The year with the smallest gap between cohort joiners and cohort leavers is 2020 (35,890 joiners vs 11,440 leavers), where it is possible to see a clear decrease in the number of young individuals registering as Welsh residents compared to the previous years. However, in 2021 and 2022, this number increases again, returning to pre-2020 values.

For SMC, the average number of deaths every year during the e-cohort study was 31,900, which accounts for 30%-40% of people leaving the cohort each year. The newborn, on average 28,640, accounts for 24%-34% of people joining SMC. For SMYC, the average number of deaths per year during is 73, which accounts for 1-2% of people leaving the cohort each year. As expected, more than 95% of people leave the cohort because they relocate outside of Wales. Between 2005 and 2020, newborns account for 70-75% of individuals joining the cohort, while in 2021 and 2022, they account only for 65% and 59% respectively, see Figure 7.

Within SMC, there is a noticeable decline in the percentage of people residing in areas with the lowest WIMD quintile (WIMD = 1 and WIMD = 2) between 2000 and 2005, as a growing number of individuals relocate to less deprived LSOAs (WIMD = 4, WIMD = 5). However, post 2014, there is a noticeable uptick in the population residing in more deprived areas (WIMD = 1) and a consequential decrease of those living in less deprived areas (WIMD = 5 and WIMD = 3), see Figure 8.

**Figure 8:**
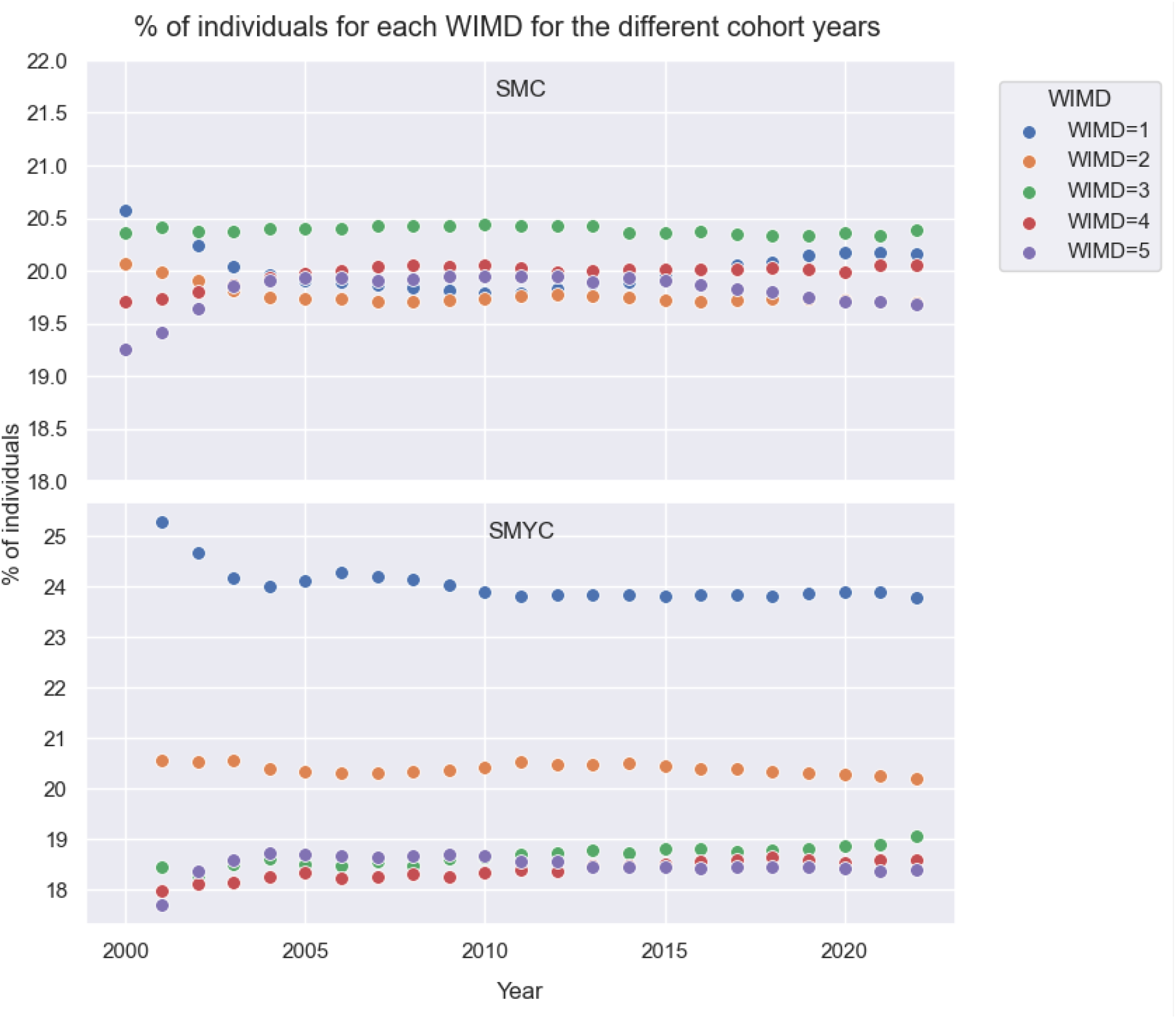
% of individuals living in areas with WIMD = 1,2,3,4,5 on the 1st January of each year of the e-cohort study for SMC and SMYC.

The majority of individuals in SMYC, between 24% and 26%, reside in an area with the lowest WIMD quintile (WIMD = 1). Approximately 20% of individuals reside in an area with WIMD = 2, while 18% of people live in less deprived areas (WIMD = 3,4,5). These percentages remain consistent during the cohort period.

### 3.2 SMC and SMYC concept curation pipeline

From the outcomes tables, created through the concept curation pipeline by linking SMC and SMYC to the look-up tables and the relevant data source inside SAIL, it is possible to extract descriptive analysis for all the concepts identified. Considering both primary and secondary care data (WLGP and PEDW), in Figure 9, we present the 20 most common condition concepts identified in SMC and SMYC. The five most common conditions for individuals in SMC are depression, affecting 21.6% of the total cohort, anxiety (21.1%), asthma (17.5%), hypertension (16.2%) and atopic eczema (14.1%). Notably, females are subject to a substantially higher incidence of anxiety and depression compared to males, with prevalence rates of 26.4% vs 16.6% and 26.1% vs 17.0%.

**Figure 9:**
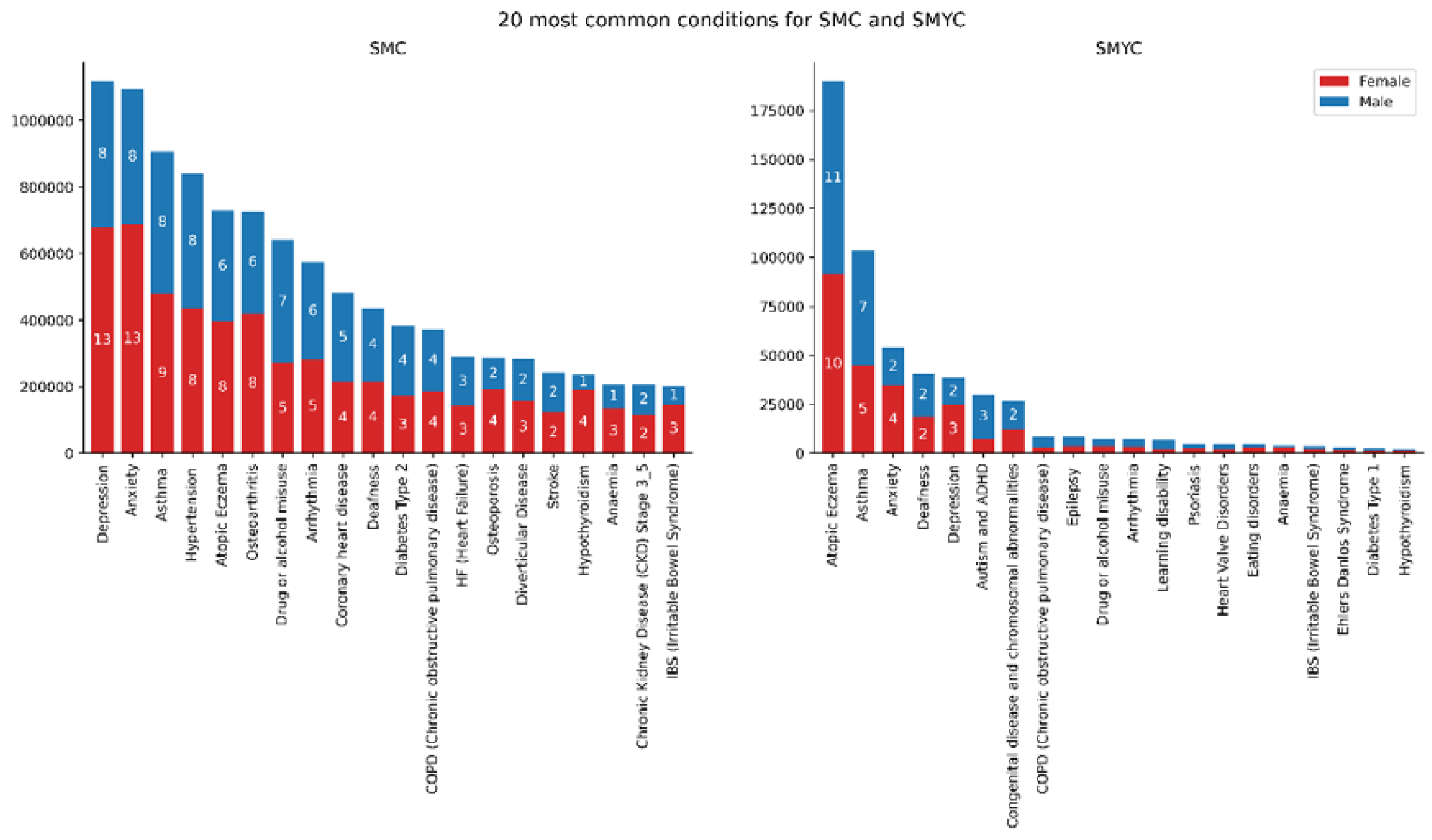
20 most common concepts for SMC and SMYC. The number on the bars represents the % of individuals for each sex with records of each concept compared to the complete SMC.

The five most common conditions for individuals in SMYC are atopic eczema (21.2%), asthma (11.6%), anxiety (6.0%), deafness (4.6%) and depression (4.3%). Similarly, within the young cohort, females demonstrate a higher prevalence of anxiety (7.5%) and depression (5.4%) compared to males (4.4% and 3.1%). However, for males, the prevalence of autism and ADHD is notably higher compared to females, with rates of 5.2% and 1.6% respectively.

Looking at the most common 20 concepts for SMC and their age onset Figure 10 it is clear that certain concepts have distinct patterns of onset across different age groups. Atopic eczema and asthma show a significant proportion of initial diagnoses occurring between the ages of 1 and 10, which account for 35.3% and 23.3% of the total diagnoses respectively. Depression, anxiety and IBS typically begin to be diagnosed during the teenage years, peaking between the ages of 20 and 30. The first records of concepts such as hypertension, diabetes type 2, coronary heart disease, osteoporosis etc., are more frequently expected in older individuals, and are typically recorded in individuals after the age of 50, see Figure 10.

**Figure 10:**
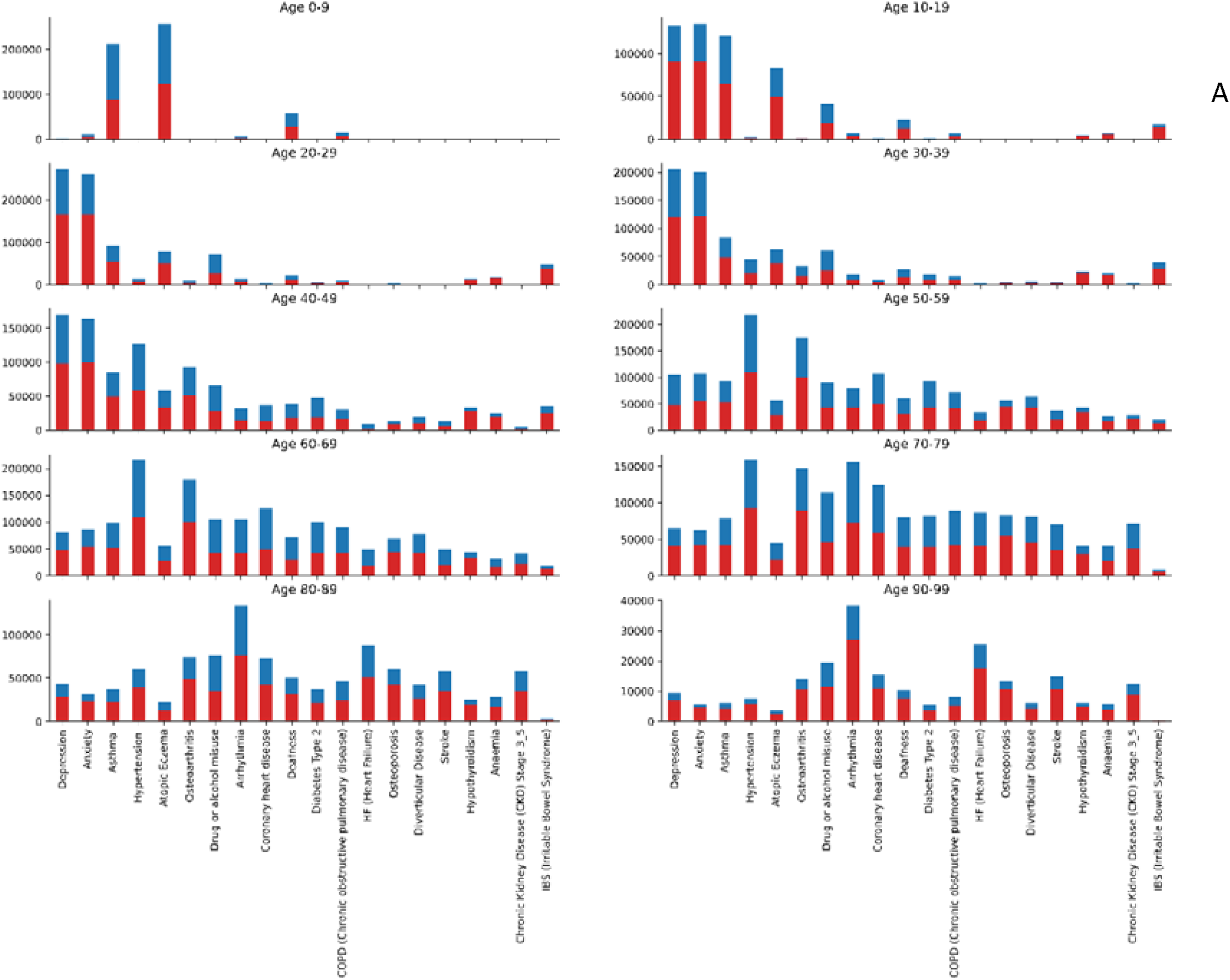
Number of individuals for each age onset group for each of the 20 most common concepts in SMC.

similar analysis for the most common 20 concepts in the SMYC Figure 11 indicates that atopic eczema, deafness, COPD, epilepsy, congenital disease and chromosomal abnormalities, arrhythmia and heart valve disorders peak between the ages of 0 and 4. Autism and ADHD and learning disabilities are mostly recorded for children in primary school age (age 4-11), while anxiety, depression and IBS have a higher incidence rate in teenagers, from the age of 16 years onwards, see Figure 11.

**Figure 11:**
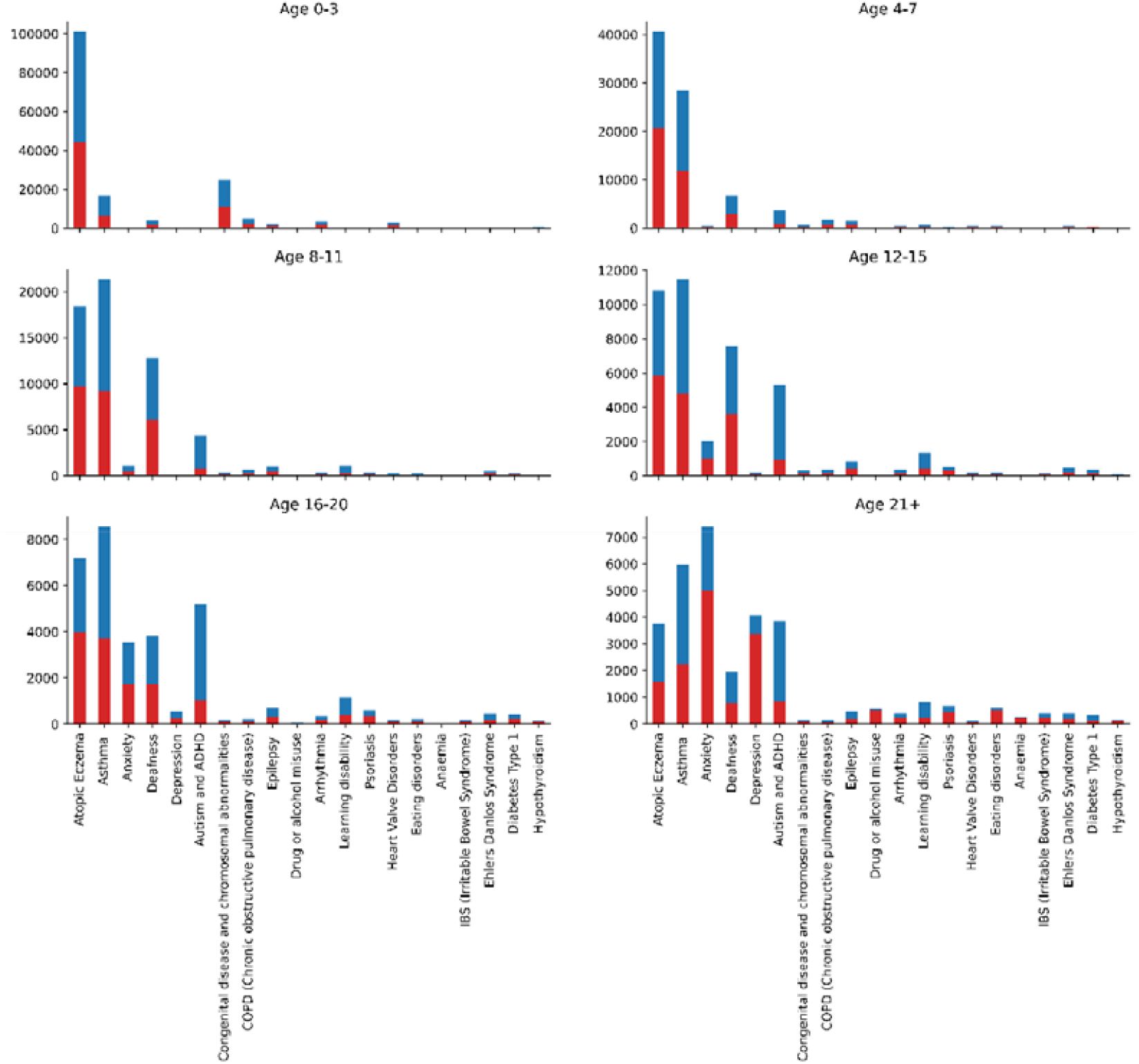
Number of individuals for each age onset group for each of the 20 most common concepts in SMYC.

For a more complete and detailed analysis for each concept included in the study, see Table 4 in Suppl.Material.

## 4. MAIN STRENGTHS AND LIMITATIONS

The main strengths of the two prospective longitudinal e-cohorts we built is the nationwide coverage of the individuals, making these e-cohorts representative of the comprehensive Welsh population over 22 years of coverage. Having the possibility to link these individuals to demographic, primary and secondary healthcare data facilitates and supports a wide range of approaches to address research questions and deliverables for the MELD-B project and future research. Moreover, the utilisation of anonymised e-cohorts serves as an effective strategy for overcoming consent-related barriers, enabling seamless data aggregation and analysis.

In this paper, we also defined a reproducible concept curation pipeline to manage and process data extraction for the e-cohorts. This pipeline ensures that whenever there is a new data release, updates to the cohorts, or modifications to the concepts or their code lists, the relevant tables containing data for the cohorts can be immediately updated, provenance of changes tracked and new dataset versions published. This enables support for multiple research questions and outcomes across the range of data analysis in SAIL. Moreover, the adaptability of this pipeline makes it a reusable tool for data preparation or initial data analysis in other research projects.

The use of electronic health record (EHR) data in cohort studies is limited by missing data and errors in routine records. In addition, EHRs often lack observations related to lived experiences, which are important considerations in multimorbidity studies.

## Data Availability

The data used in this study are available in the SAIL Databank at Swansea University, Swansea, UK. All proposals to use SAIL data are subject to review by an independent Information Governance Review Panel (IGRP). Before any data can be accessed, approval must be given by the IGRP. The IGRP carefully considers each project to ensure the proper and appropriate use of SAIL data. When approved, access is gained through a privacy-protecting trusted research environment (TRE) and remote access system referred to as the SAIL Gateway. SAIL has established an application process to be followed by anyone who would like to access data via SAIL https://www.saildatabank.com/application-process.

## ACKNOWLEDGMENTS

We would like to acknowledge all other members of the MELD-B Consortium. This work uses data provided by patients and collected by the NHS as part of their care and support. We would also like to acknowledge all data providers who make anonymised data available for research.

## FOOTNOTES

### Contributors

Conceptualisation of the study RC, AA, NA, MB, SF, RH, ZZ. Data curation and analysis RC, JD, MS: original draft writing RC, review and editing of the manuscript RC, AA, AB, MB, JD, NF, SF, EH, RH, RO, MS, SS, ZZ. All authors have read and agreed to the published version of the manuscript.

## Ethical approval

The SAIL Databank independent Information Governance Review Panel has approved this study (SAIL Project: 1377).

## Patient and public involvement

Patients and/or the public were not involved in the design of the e-cohorts.

## APPENDIX A – How to treat lost to follow-up

*Loss to follow-up* is one of the cohort censorship criteria adopted in this study. An individual is considered lost to *follow up* if their participation in the study is discontinued, which in the specific case of the MELD-B project, means that the participant moves outside of Wales. Although it is possible for an individual to re-join the cohort if they return to Wales, the MELD-B has taken a collective decision not to allow this situation following an analysis performed on the WDSD.

From WDSD we calculated that 352,550 (6.8%) individuals in SMC have a break in their residence data as they leave Wales before returning to the country after a period of at least 30 days. They account for the 24.8% of the total number of individuals censored because lost to follow up. Referring to *N periods* as the number of time an individual enters the cohort, assuming we have allowed people to re-join once they return to Wales, in the following table we present the number of individuals that join the cohort one, two, three and four or more times:

Most individuals (93.2%) join the cohort only once: once they are included in the cohort they die, move permanently outside Wales or remain in the country until the end of the study period. The vast majority of individuals with a break in their residency data leave and return to Wales only once (N = 2), and only the 0.6% of SMC have more than one break in their residency data. This result suggests to consider individuals with residency breaks as lost to follow up on the date they firstly move outside of Wales and to *not allow them to be re-included in the cohort*.

## Footnotes

1 Note that this is the cohort end date at the point of publication, however follow up data will be included once available in the SAIL Databank.

2 This allows for at least one year of follow up after entering the cohort

3 NCCH has two data classes which include maternal record: NCCH_CHILD_BIRTHS and NCCH_CHILD_TRUST

4 If these concepts are not available in routine data, proxy or derived phenotypes will be derived.

5 A controlled process is in place for importing files into SAIL to ensure they comply with SAIL policies and processes. Any files brought into SAIL must be within the scope of the project and approved by the IGRP since they could, directly or indirectly, impact the privacy protection of any data held within the TRE.

6 The current set of sensitive codes covers: miscarriage, HIV/AIDS, pregnancy termination and sexually transmitted diseases. The sensitive code lists are processed as part of the pipeline.

7 These tables are derived by the UK Health dimensions Database, which groups reference data for coding information

8 Note that, as some concepts are derived from more sources, they appear only one in the final list.

9 Each condition derived from [57] has been mapped to a concept (or more than one) available in [59] by the clinical group.

10 We considered that there is a delay between an individual’s WOB and his GP and its registration. The average delay is 23 days, but here we considered a delay of 30 days to be more inclusive.

11 For the SMYC we collect information starting on the 1^st^ January 2001.

## BIBLIOGRAPHY

[1] M. Fortin, J. Haggerty, J. Almirall, T. Bouhali, M. Sasseville, M. Lemieux, Lifestyle factors and multimorbidity: a cross sectional study, BMC Public Health 14 (2014) 686. 10.1186/1471-2458-14-686.

[2] A. Kingston, L. Robinson, H. Booth, M. Knapp, C. Jagger, B. Adelaja, M. Avendano, S.M. Bamford, S. Banerjee, S. Berwald, A. Bowling, C. Burgon, E. Bustard, A. Comas-Herrera, M. Dangoor, J. Dixon, N. Farina, S. Greengross, E. Grundy, B. Hu, D. King, D. Lombard, K. Lorenz, D. McDaid, A. La Park, J. Pikhartova, A. Rehill, R. Wittenberg, Projections of multi-morbidity in the older population in England to 2035: Estimates from the Population Ageing and Care Simulation (PACSim) model, Age Ageing 47 (2018). 10.1093/ageing/afx201.

[3] K.D. Lawson, S.W. Mercer, S. Wyke, E. Grieve, B. Guthrie, G.C. Watt, E.A. Fenwick, Double trouble: the impact of multimorbidity and deprivation on preference-weighted health related quality of life a cross sectional analysis of the Scottish Health Survey, Int J Equity Health 12 (2013). 10.1186/1475-9276-12-67.

[4] G. Berntsen, A. Høyem, I. Lettrem, C. Ruland, M. Rumpsfeld, D. Gammon, A person-centered integrated care quality framework, based on a qualitative study of patients’ evaluation of care in light of chronic care ideals, BMC Health Serv Res 18 (2018) 1–15. 10.1186/S12913-018-3246-Z/TABLES/2.

[5] C. Salisbury, L. Johnson, S. Purdy, J.M. Valderas, A.A. Montgomery, Epidemiology and impact of multimorbidity in primary care: a retrospective cohort study, British Journal of General Practice 61 (2011) e12–e21. 10.3399/BJGP11X548929.

[6] X. Feng, X. Tan, B. Riley, T. Zheng, T. Bias, U. Sambamoorthi, Polypharmacy and Multimorbidity Among Medicaid Enrollees: A Multistate Analysis, Popul Health Manag 21 (2018) 123–129. 10.1089/POP.2017.0065.

[7] M. Khezrian, C.J. McNeil, A.D. Murray, P.K. Myint, An overview of prevalence, determinants and health outcomes of polypharmacy, Ther Adv Drug Saf 11 (2020). 10.1177/2042098620933741.

[8] A. Cassell, D. Edwards, A. Harshfield, K. Rhodes, J. Brimicombe, R. Payne, S. Griffin, The epidemiology of multimorbidity in primary care: a retrospective cohort study, British Journal of General Practice 68 (2018) e245–e251. 10.3399/BJGP18X695465.

[9] D. Koller, G. Schön, I. Schäfer, G. Glaeske, H. Van Den Bussche, H. Hansen, Multimorbidity and long-term care dependency - A five-year follow-up, BMC Geriatr 14 (2014) 1–9. 10.1186/1471-2318-14-70/TABLES/3.

[10] C.E. Aubert, J.L. Schnipper, N. Fankhauser, P. Marques-Vidal, J. Stirnemann, A.D. Auerbach, E. Zimlichman, S. Kripalani, E.E. Vasilevskis, E. Robinson, J. Metlay, G.S. Fletcher, A. Limacher, J. Donzé, Association of patterns of multimorbidity with length of stay: A multinational observational study, Medicine 99 (2020) e21650. 10.1097/MD.0000000000021650.

[11] F. Eto, M. Samuel, R. Henkin, M. Mahesh, T. Ahmad, A. Angdembe, R. Hamish McAllister-Williams, P. Missier, N.J. Reynolds, M.R. Barnes, S. Hull, S. Finer, R. Mathur, Ethnic differences in early onset multimorbidity and associations with health service use, long-term prescribing, years of life lost, and mortality: A cross-sectional study using clustering in the UK Clinical Practice Research Datalink, PLoS Med 20 (2023). 10.1371/JOURNAL.PMED.1004300.

[12] T. Lehnert, D. Heider, H. Leicht, S. Heinrich, S. Corrieri, M. Luppa, S. Riedel-Heller, H.H. König, Review: health care utilization and costs of elderly persons with multiple chronic conditions, Med Care Res Rev 68 (2011) 387–420. 10.1177/1077558711399580.

[13] L.G. Glynn, J.M. Valderas, P. Healy, E. Burke, J. Newell, P. Gillespie, A.W. Murphy, The prevalence of multimorbidity in primary care and its effect on health care utilization and cost, Fam Pract 28 (2011) 516–523. 10.1093/fampra/cmr013.

[14] R. Palladino, J.T. Lee, M. Ashworth, M. Triassi, C. Millett, Associations between multimorbidity, healthcare utilisation and health status: evidence from 16 European countries, Age Ageing 45 (2016) 431–435. 10.1093/ageing/afw044.

[15] C. Bähler, C.A. Huber, B. Brüngger, O. Reich, Multimorbidity, health care utilization and costs in an elderly community-dwelling population: A claims data based observational study, BMC Health Serv Res 15 (2015) 1–12. 10.1186/S12913-015-0698-2/TABLES/6.

[16] S. MacMahon, Multiple Long-Term Conditions (Multimorbidity): a priority for global health research, London, 2018. https://acmedsci.ac.uk/policy/policy-projects/multimorbidity (accessed June 1, 2023).

[17] X. Xu, G.D. Mishra, M. Jones, Mapping the global research landscape and knowledge gaps on multimorbidity: a bibliometric study, J Glob Health 7 (2017). 10.7189/JOGH.07.010414.

[18] I.S.S. Ho, A. Azcoaga-Lorenzo, A. Akbari, C. Black, J. Davies, P. Hodgins, K. Khunti, U. Kadam, R.A. Lyons, C. McCowan, S. Mercer, K. Nirantharakumar, B. Guthrie, Examining variation in the measurement of multimorbidity in research: a systematic review of 566 studies, Lancet Public Health 6 (2021) e587–e597. 10.1016/S2468-2667(21)00107-9.

[19] F. Prazeres, L. Santiago, Prevalence of multimorbidity in the adult population attending primary care in Portugal: a cross-sectional study, BMJ Open 5 (2015) e009287. 10.1136/BMJOPEN-2015-009287.

[20] Q. Foguet-Boreu, C. Violán, T. Rodriguez-Blanco, A. Roso-Llorach, M. Pons-Vigués, E. Pujol-Ribera, Y.C. Gil, J.M. Valderas, Multimorbidity Patterns in Elderly Primary Health Care Patients in a South Mediterranean European Region: A Cluster Analysis, PLoS One 10 (2015). 10.1371/JOURNAL.PONE.0141155.

[21] A. Prados-Torres, A. Calderón-Larrañaga, J. Hancco-Saavedra, B. Poblador-Plou, M. Van Den Akker, Multimorbidity patterns: a systematic review, J Clin Epidemiol 67 (2014) 254–266. 10.1016/J.JCLINEPI.2013.09.021.

[22] N. Garin, A. Koyanagi, S. Chatterji, S. Tyrovolas, B. Olaya, M. Leonardi, E. Lara, S. Koskinen, B. Tobiasz-Adamczyk, J.L. Ayuso-Mateos, J.M. Haro, Global Multimorbidity Patterns: A Cross-Sectional, Population-Based, Multi-Country Study, J Gerontol A Biol Sci Med Sci 71 (2016) 205–214. 10.1093/GERONA/GLV128.

[23] R.A. Goodman, S.M. Ling, P.A. Briss, R.G. Parrish, M.E. Salive, B.S. Finke, Multimorbidity Patterns in the United States: Implications for Research and Clinical Practice, The Journals of Gerontology: Series A 71 (2016) 215–220. 10.1093/GERONA/GLV199.

[24] I. Kirchberger, C. Meisinger, M. Heier, A.K. Zimmermann, B. Thorand, C.S. Autenrieth, A. Peters, K.H. Ladwig, A. Döring, Patterns of multimorbidity in the aged population. Results from the KORA-Age study, PLoS One 7 (2012). 10.1371/JOURNAL.PONE.0030556.

[25] J. Lyons, A. Akbari, K.R. Abrams, A. Azcoaga Lorenzo, T. Ba Dhafari, J. Chess, S. Denaxas, R. Fry, C.P. Gale, J. Gallacher, L.J. Griffiths, B. Guthrie, M. Hall, F. Jalalinajafabadi, A. John, C. MacRae, C. McCowan, N. Peek, D. O’Reilly, J. Rafferty, R.A. Lyons, R.K. Owen, Trajectories in chronic disease accrual and mortality across the lifespan in Wales, UK (2005–2019), by area deprivation profile: linked electronic health records cohort study on 965,905 individuals, The Lancet Regional Health - Europe 32 (2023) 100687. 10.1016/j.lanepe.2023.100687.

[26] E.F. France, S. Wyke, J.M. Gunn, F.S. Mair, G. McLean, S.W. Mercer, Multimorbidity in primary care: a systematic review of prospective cohort studies, Br J Gen Pract 62 (2012). 10.3399/BJGP12X636146.

[27] M. Ashworth, S. Durbaba, D. Whitney, J. Crompton, M. Wright, H. Dodhia, Journey to multimorbidity: longitudinal analysis exploring cardiovascular risk factors and sociodemographic determinants in an urban setting, BMJ Open 9 (2019) e031649. 10.1136/BMJOPEN-2019-031649.

[28] R. López-Bueno, Z. Feng, E. Ortega-Martín, Social determinants of multimorbidity patterns: A systematic review, n.d.

[29] X. Xu, G.D. Mishra, A.J. Dobson, M. Jones, Progression of diabetes, heart disease, and stroke multimorbidity in middle-aged women: A 20-year cohort study, PLoS Med 15 (2018) e1002516. 10.1371/JOURNAL.PMED.1002516.

[30] G. Ruel, J.F. Lévesque, N. Stocks, C. Sirois, E. Kroger, R.J. Adams, M. Doucet, A.W. Taylor, Understanding the Evolution of Multimorbidity: Evidences from the North West Adelaide Health Longitudinal Study (NWAHS), PLoS One 9 (2014) e96291. 10.1371/JOURNAL.PONE.0096291.

[31] S. Stannard, E. Holland, S.R. Crozier, R. Hoyle, M. Boniface, M. Ahmed, J. McMahon, W. Ware, Z. Zlatev, N.A. Alwan, S.D.S. Fraser, Early-onset burdensome multimorbidity: an exploratory analysis of sentinel conditions, condition accrual sequence and duration of three long-term conditions using the 1970 British Cohort Study, BMJ Open 12 (2022). 10.1136/BMJOPEN-2021-059587.

[32] G. Cezard, C.T. McHale, F. Sullivan, J.K.F. Bowles, K. Keenan, Studying trajectories of multimorbidity: a systematic scoping review of longitudinal approaches and evidence, BMJ Open 11 (2021). 10.1136/BMJOPEN-2020-048485.

[33] R. Owen, J. Lyons, A. Akbari, B. Guthrie, D. Alexander, A. Azcoaga-Lorenzo, A. Brookes, S. Denaxas, C. Dezateux, A. Fagbamigbe, G. Harper, P. Kirk, E. Özyiğit, S. Richardson, S. Staniszewska, C. Mccowan, R. Lyons, K. Abrams, Temporal sequencing in multimorbidity using population-scale linked data for 1.7 million individuals with 20-year follow-up, (2022). 10.21203/RS.3.RS-1537576/V1.

[34] A. Head, K. Fleming, C. Kypridemos, J. Pearson-Stuttard, M. O’Flaherty, Multimorbidity: the case for prevention, J Epidemiol Community Health (1978) 75 (2021) 242–244. 10.1136/JECH-2020-214301.

[35] S.D. Fraser, S. Stannard, E. Holland, M. Boniface, R.B. Hoyle, R. Wilkinson, A. Akbari, M. Ashworth, A. Berrington, R. Chiovoloni, J. Enright, N.A. Francis, G. Giles, M. Gulliford, S. Macdonald, F.S. Mair, R.K. Owen, S. Paranjothy, H. Parsons, R.J. Sanchez-Garcia, M. Shiranirad, Z. Zlatev, N. Alwan, Multidisciplinary ecosystem to study lifecourse determinants and prevention of early-onset burdensome multimorbidity (MELD-B) – protocol for a research collaboration, 10.1177/26335565231204544 13 (2023). 10.1177/26335565231204544.

[36] S. Stannard, A. Berrington, S. Fraser, S. Paranjothy, R. Hoyle, R. Owen, A. Akbari, M. Shiranirad, R. Chiovoloni, N. Alwan, Mapping domains of early-life determinants of future multimorbidity across three UK longitudinal cohort studies, MedRxiv (2024) 2024.02.01.24301771. 10.1101/2024.02.01.24301771.

[37] S. Stannard, A. Berrington, S. Paranjothy, R. Owen, S. Fraser, R. Hoyle, M. Boniface, B. Wilkinson, A. Akbari, S. Batchelor, W. Jones, M. Ashworth, J. Welch, F.S. Mair, N.A. Alwan, A conceptual framework for characterising lifecourse determinants of multiple long-term condition multimorbidity, Journal of Multimorbidity and Comorbidity 13 (2023). 10.1177/26335565231193951.

[38] R.K. Owen, J. Lyons, A. Akbari, B. Guthrie, U. Agrawal, D.C. Alexander, A. Azcoaga-Lorenzo, A.J. Brookes, S. Denaxas, C. Dezateux, A. Francis Fagbamigbe, G. Harper, P.D. W Kirk, E. Bilici Özyiğit, S. Richardson, S. Staniszewska, C. McCowan, R.A. Lyons, K.R. Abrams, Effect on life expectancy of temporal sequence in a multimorbidity cluster of psychosis, diabetes, and congestive heart failure among 1Â·7 million individuals in Wales with 20-year follow-up: a retrospective cohort study using linked data, Lancet Public Health 8 (2023) e535–e545. 10.1016/S2468-2667(23)00098-1.

[39] Home - SAIL Databank, (n.d.). https://saildatabank.com/ (accessed June 5, 2023).

[40] P.D. Gluckman, T. Buklijas, M.A. Hanson, The Developmental Origins of Health and Disease (DOHaD) Concept: Past, Present, and Future, The Epigenome and Developmental Origins of Health and Disease (2016) 1–15. 10.1016/B978-0-12-801383-0.00001-3.

[41] J. Humphreys, K. Jameson, C. Cooper, E. Dennison, Early-life predictors of future multi-morbidity: results from the Hertfordshire Cohort, Age Ageing 47 (2018) 474–478. 10.1093/AGEING/AFY005.

[42] D. Gondek, D. Bann, M. Brown, M. Hamer, A. Sullivan, G.B. Ploubidis, Prevalence and early-life determinants of mid-life multimorbidity: evidence from the 1970 British birth cohort, BMC Public Health 21 (2021). 10.1186/S12889-021-11291-W.

[43] S. Wilding, N. Ziauddeen, D. Smith, P. Roderick, D. Chase, N.A. Alwan, Are environmental area characteristics at birth associated with overweight and obesity in school-aged children? Findings from the SLOPE (Studying Lifecourse Obesity PrEdictors) population-based cohort in the south of England, BMC Med 18 (2020). 10.1186/S12916-020-01513-0.

[44] T.P. Fleming, A.J. Watkins, M.A. Velazquez, J.C. Mathers, A.M. Prentice, J. Stephenson, M. Barker, R. Saffery, C.S. Yajnik, J.J. Eckert, M.A. Hanson, T. Forrester, P.D. Gluckman, K.M. Godfrey, Origins of lifetime health around the time of conception: causes and consequences, Lancet 391 (2018) 1842–1852. 10.1016/S0140-6736(18)30312-X.

[45] R.A. Lyons, K.H. Jones, G. John, C.J. Brooks, J.P. Verplancke, D. V. Ford, G. Brown, K. Leake, The SAIL databank: Linking multiple health and social care datasets, BMC Med Inform Decis Mak 9 (2009). 10.1186/1472-6947-9-3.

[46] D. V. Ford, K.H. Jones, J.P. Verplancke, R.A. Lyons, G. John, G. Brown, C.J. Brooks, S. Thompson, O. Bodger, T. Couch, K. Leake, The SAIL Databank: Building a national architecture for e-health research and evaluation, BMC Health Serv Res 9 (2009) 1–12. 10.1186/1472-6963-9-157/TABLES/1.

[47] K.H. Jones, D. V. Ford, C. Jones, R. Dsilva, S. Thompson, C.J. Brooks, M.L. Heaven, D.S. Thayer, C.L. McNerney, R.A. Lyons, A case study of the Secure Anonymous Information Linkage (SAIL) Gateway: A privacy-protecting remote access system for health-related research and evaluation, J Biomed Inform 50 (2014) 196–204. 10.1016/J.JBI.2014.01.003.

[48] A. Akbari, F. Torabi, S. Bedston, E. Lowthian, H. Abbasizanjani, R. Fry, M. Jane Lyons, R.K. Owen, P. Kamlesh Khunti, P.A. Ronan Lyons, Developing a research ready population-scale linked data ethnicity-spine in Wales, MedRxiv (2022) 2022.11.28.22282810. 10.1101/2022.11.28.22282810.

[49] WLGP coverage reports - Analytical Services Public - Swansea University Medical School Confluence Site, (n.d.). https://docs.hiru.swan.ac.uk/display/SATP/WLGP+coverage+reports (accessed April 4, 2024).

[50] H. Abbasizanjani, F. Torabi, S. Bedston, T. Bolton, G. Davies, S. Denaxas, R. Griffiths, L. Herbert, S. Hollings, S. Keene, K. Khunti, E. Lowthian, J. Lyons, M.A. Mizani, J. Nolan, C. Sudlow, V. Walker, W. Whiteley, A. Wood, A. Akbari, Harmonising electronic health records for reproducible research: challenges, solutions and recommendations from a UK-wide COVID-19 research collaboration, BMC Med Inform Decis Mak 23 (2023) 1–15. 10.1186/S12911-022-02093-0/FIGURES/4.

[51] Health Data Research Innovation Gateway, (n.d.). https://web.www.healthdatagateway.org/search?search=&datasetSort=latest&tab=Datasets (accessed July 13, 2023).

[52] Welsh Longitudinal General Practice Dataset (WLGP) - Welsh Primary Care, (n.d.). https://web.www.healthdatagateway.org/dataset/33fc3ffd-aa4c-4a16-a32f-0c900aaea3d2 (accessed May 30, 2023).

[53] J. Dylag, R. Chiovoloni, A. Akbari, S. Fraser, M. Boniface, A Tool for Automating the Curation of Medical Concepts derived from Coding Lists, 2024 (n.d.). https://git.soton.ac.uk/meld/meldb/concepts-processing (accessed April 22, 2024).

[54] Concept Library, (n.d.). https://conceptlibrary.saildatabank.com/ (accessed September 21, 2023).

[55] H. Dambha-Miller, A. Farmer, K. Nirantharakumar, T. Jackson, C. Yau, L. Walker, I. Buchan, S. Finer, M.R. Barnes, N.J. Reynolds, G.T. Jun, S. Gangadharan, S. Fraser, B. Guthrie, Artificial Intelligence for Multiple Long-term conditions (AIM): A consensus statement from the NIHR AIM consortia, 2023. 10.3310/nihropenres.1115210.1.

[56] K. Barnett, S.W. Mercer, M. Norbury, G. Watt, S. Wyke, B. Guthrie, Epidemiology of multimorbidity and implications for health care, research, and medical education: A cross-sectional study, The Lancet 380 (2012). 10.1016/S0140-6736(12)60240-2.

[57] I.S.S. Ho, A. Azcoaga-Lorenzo, A. Akbari, J. Davies, K. Khunti, U.T. Kadam, R.A. Lyons, C. McCowan, S.W. Mercer, K. Nirantharakumar, S. Staniszewska, B. Guthrie, Measuring multimorbidity in research: Delphi consensus study, BMJ Medicine 1 (2022). 10.1136/bmjmed-2022-000247.

[58] P. Hanlon, B.D. Jani, B. Nicholl, J. Lewsey, D.A. McAllister, F.S. Mair, Associations between multimorbidity and adverse health outcomes in UK Biobank and the SAIL Databank: A comparison of longitudinal cohort studies, PLoS Med 19 (2022). 10.1371/JOURNAL.PMED.1003931.

[59] GitHub - THINKINGGroup/phenotypes, (n.d.). https://github.com/THINKINGGroup/phenotypes (accessed May 30, 2023).

[60] MELD-B Concepts Release, (2024). https://git.soton.ac.uk/meldb/concepts/-/tree/v2.2.4 (accessed April 22, 2024).

