## Supplementary material for "Cohort profile for the creation of the SAIL MELD-B e-cohort (SMC) and SAIL MELD-B children and Young adult e-cohort (SMYC)": Suppl.Material1

**Suppl. Table 1**

**List of long-term conditions concepts approved and imported into SAIL at the initial stage**

In the following table we report the provisional conditions selected by the MELD-B clinical group, their source, and their code lists sources. These concepts have gone through the full concept pipeline. All the concepts and associated code lists in this table are derived from [50] [51] and [52].

| ***CONCEPT*** | ***Code list source^[1]^*** |
| --- | --- |
| Addison's disease | [52]: Addison_Disease_birm_cam |
| Anaemia | [52] : B12deficiency_birm_cam,; PerniciousAnaemia_birm_cam,; Folate%20deficiency_birm_cam |
| Aneurysm | [52] : AorticAneurysm_Bham_CAM |
| Ankylosing Spondylitis | [52] : AnkylosingSpondylitis_MM_birm_cam |
| Anxiety | [51]  [52]: Anxiety_birm_cam |
| Arrhythmia | [51]  [52]: Arrhythmia_Bham_CAM |
| Asthma | [51]  [52]: Asthma_PUSHAsthma |
| Atopic Eczema | [52]: AtopicEczema_birm_cam |
| Autism and ADHD | [52]: ADHD_mm_birm_cam, Autism_birm_cam |
| Bipolar Disorder | [52]: Bipolar_birm_cam |
| Blindness and low vision | [51]  [52]: Visual_Impairment_Birm_Cam |
| Breast Cancer | [52]: BreastCancer_birm_cam |
| Bronchiectasis | [51]  [52]: Bronchiectasis_birm_cam |
| Chronic Back Pain | [52]: Chronicbackpain_MM_birm_cam |
| Chronic Fatigue Syndrome | [52]: ChronicFatigueSyndromeMM_birm_cam |
| Chronic Liver Disease | [51]  [52]:ChronicLiverDisease_MM_birm_cam, Chronic_liver_disease_alcohol_birm_cam, Autoimmune_liver_disease_birm_cam, Hepatitis_B_birm_cam, Hepatitis_C_birm_cam, NAFLD_birm_cam |
| Chronic Pain | [52]: ComplexPainMM_birm_cam, Chronicbackpain_MM_birm_cam |
| Chronic Sinusitis | [51]  [52]: ChronicSinusitis_birm_cam |
| Chronic Kidney Disease (CKD) Stage 3_5 | [52]: CKDstage3to5_Bham_CAM |
| Coeliac Disease | [52]: Coeliac_disease_birm_cam |
| Colon Cancer | [52]: Colon_cancer__bham_cam |
| Congenital disease and chromosomal abnormalities | [52]: CongenitalHrtDx_NoSurg_Bham_CAM |
| COPD (Chronic obstructive pulmonary disease) | [51]  [52]: COPD_birm_cam |
| Coronary heart disease | [51]  [52]: IHD_noMI_Bham_CAM, MInfarction_Bham_CAM |
| Cystic fibrosis | [52]: CysticFibrosis_birm_cam |
| Deafness | [51]  [52]: SevereDeafHearingLoss_birm_cam, Any_Deafness_Hearing_Loss_birm_cam |
| Dementia alzheimer | [51]  [52]: DementiaOther_birm_cam, Alzheimers_birm_cam |
| Depression | [51]  [52]: Depression_birm_cam |
| Diabetes Type 1 | [52]: Type1DM_11_3_21_birm_cam |
| Diabetes Type 2 | [52]: Type2Diabetes_11_3_21_birm_cam |
| Diabetic retinopathy | [52]: diabetic_retinopathy_090121_birm_cam |
| Dialysis | [52] Dialysis MM_birm_cam |
| Diverticular Disease | [51]  [52]: diverticular_disease_birm_cam |
| Drug or alcohol misuse | [51]  [52]:AlcoholMisuse_birm_cam, SubstanceMisuse_birm_cam |
| Eating disorders | [51]  [52]: EatingDisorders_birm_cam |
| Ehlers Danlos Syndrome | [52]: EhlersDanlosSyndrome3_birm_cam |
| Endometriosis | [52]: Endometriosis_Adenomyosis_birm_cam |
| Epilepsy | [51]  [52]: Epilepsy_birm_cam |
| Fibromyalgia | [52]: Fibromyalgia_Bham_CAM |
| Glaucoma | [51]  [52]: Glaucoma_Prevalence_Birm_Cam |
| Gout | [52]: Gout_MM_birm_cam |
| Heart Valve Disorders | [52]: ValvularDiseases_BhaM_CAM |
| HF (Heart Failure) | [51]  [52]: HF_Bham_CAM_Final |
| HIV/AIDS | [52]: HIVaids_birm_cam |
| Hypertension | [51]  [52]: Hypertension_BhaM_CAM |
| Hyperthyroidism | [52]: Hyperthyroidism V2_birm_cam |
| Hypothyroidism | [52]: Hypothyroidism draft V1_birm_cam |
| IBD (Inflammatory bowel disease) | [51]  [52] : Crohns_disease_birm_cam, Ulcerative_colitis_birm_cam |
| IBS (Irritable Bowel Syndrome) | [51] |
| ILD (Inflammatory Lung Disease) | [52] : ILD_sh_20092020_birm_cam |
| Learning disability | [51]  [52]: Learningdisability_birm_cam |
| Leukaemia | [52]: leukaemia_prevalence_birm_cam |
| Lymphoma | [52]: lymphoma_prevalence_birm_cam |
| Marfan Syndrome | [52]: MarfanSyndrome_birm_cam |
| Meniere's disease | [52]: Menieresdisease_birm_cam |
| Metastatic cancers | [52]: MetastaticCancer_birm_cam |
| Multiple sclerosis | [51]  [52]: MS_120421_birm_cam |
| Osteoarthritis | [52]: Osteoarthritis_MM_birm_cam |
| Osteoporosis | [52]: Osteoporosis_birm_cam |
| Pancreatic disease | [52]: ChronicPancreatitis_MM_birm_cam |
| Paralysis | [52]: Hemi_Para_Quadriplegia_birm_cam |
| Parkinson's disease | [51]  [52]: Parkinsons_birm_cam |
| PCOS (Polycystic ovary syndrome) | [52]: Polycystic_ovarian_syndrome_PCOS_birm_cam_v2 |
| Plasmacell | [52]: plasmacell_neoplasm_birm_cam |
| PMR (Polymyalgia reheumatica) | [52]: Polymyalgiarheumatica_MM |
| Primary Lung Cancer | [52]: Primary lung cancer_birm_cam |
| Prostate Cancer | [52]: PrimaryProstateCa_prevalence_120421_birm_cam |
| Prostate disorders | [51]  [52]: Benign Prostate Hyperplasia_birm_cam |
| Psoriasis | [52]: Psoriasis_birm_cam |
| Psoriasis Arthritis | [52]: PsoriaticArthritis2021_MM_birm_cam |
| PTSD (Post-traumatic stress disorders) | [52]: PTSD_TLC_V2 |
| PVD (Pheripheral vascular disease) | [51]  [52] : PVD_Bham_CAM |
| Renal Transplant | [52] : Renal transplant_MM_birm_cam |
| Rheumatoid arthritis | [52]: RheumatoidArthritis_MM_birm_cam |
| Schizophrenia | [51]  [52]: SchizophreniaMM |
| Skin Cancer | [52]: SkinCancer_200421_birm_cam |
| Stroke | [52]: StrokeUnspecified_Bham_CAM, Stroke_Haemrgic_Bham_CAM |
| Systemic Lupus Erythematosus | [52]: Systemic_lupus_erythematosus_MM_birm_cam |
| Systemic sclerosis | [52]: Systemic_Sclerosis_MM_birm_cam |
| Thalassaemia | [52]: Thalassaemia_birm_cam |
| TIA (Transient ischaemic attack) | [52]: TIA_Bham_CAM |
| Viral Hepatitis | [51]  [52]: Hepatitis_B_birm_cam, Hepatitis_C_birm_cam |
| VTD (Venous thrombotic disease) | [52]: VTEnoPE_birm_cam |

**Suppl.table 2**

**SMC Evolution over the e-cohort study period**

| **Year** | **Total (N)** | **Male**  **(N, %)** | **Female**  **(N ,%)** | **WIMD = 1 (%)** | **WIMD = 2 %** | **WIMD = 3 (%)** | **WIMD = 4 (%)** | **WIMD = 5 (%)** | **People leaving (N)** | **Deaths**  **(N, %)** | **Lost to follow-up**  **(N, %)** | **People joining (N)** | **New born**  **(N , %)** |
| --- | --- | --- | --- | --- | --- | --- | --- | --- | --- | --- | --- | --- | --- |
| **2000** | 2,990,123 | 1,477,430 (49.41) | 1,512,693 (50.59) | 20.57 | 20.07 | 20.37 | 19.70 | 19.26 | 101,648 | 31,919 (31.4) | 69,729  (68.6) | 105,654 | 26,248 (24.84) |
| **2001** | 2,994,184 | 1,481,187 (49.47) | 1,512,997 (50.53) | 20.41 | 19.99 | 20.41 | 19.73 | 19.41 | 94,869 | 31,236 (32.93) | 63,633 (67.07) | 95,940 | 25,733 (26.82) |
| **2002** | 2,995,269 | 1,483,212 (49.52) | 1,512,057 (50.48) | 20.24 | 19.91 | 20.37 | 19.80 | 19.64 | 104,774 | 31,041 (29.63) | 73,733 (70.37) | 100,048 | 25,022 (25.01) |
| **2003** | 2,990,558 | 1,481,707 (49.55) | 1,508,851 (50.45) | 20.05 | 19.82 | 20.37 | 19.87 | 19.85 | 84,111 | 32,548 (38.7) | 51,563  (61.3) | 103,729 | 26,586 (25.63) |
| **2004** | 3,010,192 | 1,493,705 (49.62) | 1,516,487 (50.38) | 19.96 | 19.75 | 20.41 | 19.93 | 19.91 | 83,055 | 31,695 (38.16) | 51,360 (61.84) | 99,527 | 27,245 (27.37) |
| **2005** | 3,026,683 | 1,503,414 (49.67) | 1,523,269 (50.33) | 19.91 | 19.74 | 20.41 | 19.97 | 19.93 | 79,923 | 31,723 (39.69) | 48,200 (60.31) | 104,258 | 28,439 (27.28) |
| **2006** | 3,0510,44 | 1,516,967 (49.72) | 1,534,077 (50.28) | 19.89 | 19.74 | 20.40 | 20.00 | 19.93 | 80,919 | 30,653 (37.88) | 50,266 (62.12) | 97,075 | 29,670 (30.56) |
| **2007** | 3,067,222 | 1,526,490 (49.77) | 1,540,732 (50.23) | 19.87 | 19.72 | 20.43 | 20.04 | 19.91 | 82,019 | 31,401 (38.29) | 50,618 (61.71) | 94,865 | 30,081 (31.71) |
| **2008** | 3,080,096 | 1,534,652 (49.82) | 1,545,444 (50.18) | 19.84 | 19.70 | 20.43 | 20.06 | 19.92 | 85,327 | 31,741 (37.2) | 53,586  (62.8) | 93,286 | 31,194 (33.44) |
| **2009** | 3,088,059 | 1,540,893 (49.9) | 1,547,166 (50.1) | 19.81 | 19.72 | 20.43 | 20.05 | 19.95 | 87,761 | 30,555 (34.82) | 57,206 (65.18) | 89,007 | 30,610 (34.39) |
| **2010** | 3,089,310 | 1,543,193 (49.95) | 1,546,117 (50.05) | 19.80 | 19.73 | 20.44 | 20.05 | 19.95 | 99,335 | 30,561 (30.77) | 68,774 (69.23) | 91,584 | 31,657 (34.57) |
| **2011** | 3,081,559 | 1,540,737 (50) | 1,540,822 (50) | 19.80 | 19.77 | 20.43 | 20.03 | 19.95 | 97,299 | 29,682 (30.51) | 67,617 (69.49) | 93,357 | 31,821 (34.09) |
| **2012** | 3,077,617 | 1,539,702 (50.03) | 1,5379,15 (49.97) | 19.83 | 19.77 | 20.43 | 19.99 | 19.95 | 99,588 | 30,825 (30.95) | 68,763 (69.05) | 92,103 | 31,875 (34.61) |
| **2013** | 3,070,132 | 1,536,640 (50.05) | 1,533,492 (49.95) | 19.88 | 19.77 | 20.43 | 20.01 | 19.90 | 103,938 | 31,005 (29.83) | 72,933 (70.17) | 94,646 | 30,156 (31.86) |
| **2014** | 3,060,841 | 1,532,653 (50.07) | 1,528,188 (49.93) | 19.90 | 19.75 | 20.36 | 20.01 | 19.94 | 98,647 | 30,687 (31.11) | 67,960 (68.89) | 92,464 | 30,124 (32.58) |
| **2015** | 3,054,662 | 1,530,274 (50.1) | 1,524,388 (49.9) | 19.95 | 19.73 | 20.37 | 20.01 | 19.91 | 97,303 | 31,597 (32.47) | 65,706 (67.53) | 92,003 | 30,082  (32.7) |
| **2016** | 3,049,364 | 1,528,550 (50.13) | 1,520,814 (49.87) | 20.02 | 19.71 | 20.37 | 20.01 | 19.87 | 97,241 | 32,073 (32.98) | 65,168 (67.02) | 93,787 | 29,778 (31.75) |
| **2017** | 3,045,910 | 1,526,866 (50.13) | 1,519,044 (49.87) | 20.06 | 19.73 | 20.35 | 20.01 | 19.83 | 98,619 | 32,007 (32.46) | 66,612 (67.54) | 93,177 | 29,352  (31.5) |
| **2018** | 3,040,468 | 1,523,050 (50.09) | 1,517,418 (49.91) | 20.09 | 19.73 | 20.33 | 20.02 | 19.80 | 96,033 | 32,917 (34.28) | 63,116 (65.72) | 91,973 | 28,644 (31.14) |
| **2019** | 3,036,408 | 1,520,715 (50.08) | 1,515,693 (49.92) | 20.15 | 19.73 | 20.33 | 20.01 | 19.75 | 95,119 | 32,089 (33.74) | 63,030 (66.26) | 91,317 | 26,706 (29.25) |
| **2020** | 3,032,606 | 1,518,740 (50.08) | 1,513,866 (49.92) | 20.18 | 19.72 | 20.37 | 19.99 | 19.72 | 90,097 | 36,157 (40.13) | 53,940 (59.87) | 74,609 | 25,750 (34.51) |
| **2021** | 3,017,118 | 1,511,147 (50.09) | 1,505,971 (49.91) | 20.17 | 19.70 | 20.34 | 20.05 | 19.71 | 105,720 | 34,727 (32.85) | 70,993 (67.15) | 102,100 | 26,006 (25.47) |

**Suppl.table 3**

SMYC evolution over the e-cohort study period.

| **Year** | **Total (N)** | **Male**  **(N, %)** | **Female**  **(N ,%)** | **WIMD = 1 (%)** | **WIMD = 2 %** | **WIMD = 3 (%)** | **WIMD = 4 (%)** | **WIMD = 5 (%)** | **People leaving (N)** | **Deaths**  **(N, %)** | **Lost to follow-up**  **(N, %)** | **People joining (N)** | **New born**  **(N , %)** |
| --- | --- | --- | --- | --- | --- | --- | --- | --- | --- | --- | --- | --- | --- |
| **2000** | NA | NA | NA | NA | NA | NA | NA | NA | 259 | 16  (6.17) | 243 (93.82) | 26,184 | 26,184 (100) |
| **2001** | 25,926 | 13,453 (51.89) | 12,473 (48.11) | 25.28 | 20.57 | 18.44 | 17.97 | 25.28 | 980 | 35 (3.57) | 945 (96.43) | 30,754 | 25,693 (83.54) |
| **2002** | 55,700 | 28,779 (51.67) | 26,921 (48.33) | 24.68 | 20.53 | 18.27 | 18.11 | 24.68 | 1,765 | 40  (2.26) | 1,725 (97.74) | 31,514 | 24,994 (79.31) |
| **2003** | 85,451 | 44,226 (51.76) | 41,225 (48.24) | 24.16 | 20.57 | 18.51 | 18.15 | 24.16 | 2,049 | 51 (2.48) | 1,998 (97.51) | 34,108 | 26,573 (77.91) |
| **2004** | 117,510 | 60,651 (51.61) | 56,859 (48.39) | 23.99 | 20.40 | 18.62 | 18.24 | 23.99 | 2,392 | 63 (2.63) | 2,329 (97.37) | 35,153 | 27,237 (77.48) |
| **2005** | 150,274 | 77,416 (51.52) | 72,858 (48.48) | 24.12 | 20.33 | 18.49 | 18.33 | 24.12 | 2,789 | 59 (2.11) | 2,730 (97.89) | 39,170 | 28,438 (72.60) |
| **2006** | 186,660 | 96,131 (51.50) | 90,529 (48.50) | 24.29 | 20.30 | 18.48 | 18.21 | 24.29 | 3,386 | 76 (2.24) | 3,310 (97.76) | 38,766 | 29,670 (76.54) |
| **2007** | 222,032 | 114,254 (51.46) | 107,778 (48.54) | 24.19 | 20.31 | 18.55 | 18.26 | 24.19 | 3,706 | 71 (1.91) | 3,635 (98.09) | 39,558 | 30,081 (76.04) |
| **2008** | 257,893 | 132,625 (51.43) | 125,268 (48.57) | 24.15 | 20.33 | 18.49 | 18.32 | 24.15 | 4,550 | 60 (1.31) | 4,490 (98.69) | 41,829 | 31,194 (74.58) |
| **2009** | 295,161 | 151,832 (51.44) | 143,329 (46.56) | 24.04 | 20.37 | 18.62 | 18.25 | 24.04 | 4,977 | 76 (1.52) | 4,901 (98.48) | 40,648 | 30,610 (75.31) |
| **2010** | 330,831 | 170,056 (51.40) | 160,775 (48.60) | 23.90 | 20.42 | 18.65 | 18.34 | 23.90 | 6,516 | 69 (1.05) | 6,447 (98.95) | 42,213 | 31,656 (74.99) |
| **2011** | 366,528 | 188,301 (51.37) | 178,227 (48.63) | 23.82 | 20.52 | 18.70 | 18.39 | 23.82 | 6,561 | 62 (0.94) | 6,499 (99.06) | 42,826 | 31,821 (74.30) |
| **2012** | 402,793 | 206,930 (51.37) | 195,863 (48.63) | 23.84 | 20.48 | 18.73 | 18.36 | 23.84 | 6,816 | 79 (1.15) | 6,737 (99.85) | 42,714 | 31,875 (74.62) |
| **2013** | 438,691 | 225,158 (51.32) | 213,533 (48.68) | 23.84 | 20.46 | 18.77 | 18.46 | 23.84 | 7,230 | 84 (1.16) | 7,146 (98.84) | 42,027 | 30,156 (74.75) |
| **2014** | 473,488 | 243,107 (51.34) | 230,381 (48.66) | 23.83 | 20.51 | 18.73 | 18.47 | 23.83 | 7,376 | 59 (0.79) | 7,317 (99.21) | 41,976 | 30,124 (71.76) |
| **2015** | 508,090 | 260,838 (51.34) | 247,252 (48.66) | 23.81 | 20.44 | 18.80 | 18.50 | 23.81 | 7,162 | 75 (1.04) | 7,087 (99.96) | 42,137 | 30,054 (71.32) |
| **2016** | 543,065 | 278,678 (51.32) | 264,387 (48.68) | 23.83 | 20.38 | 18.81 | 18.55 | 23.83 | 7,707 | 80 (1.03) | 7,627 (98.97) | 41,875 | 29,443 (70.31) |
| **2017** | 577,231 | 296,134 (51.30) | 281,097 (48.70) | 23.83 | 20.39 | 18.75 | 18.59 | 23.83 | 8,433 | 97 (1.15) | 8,336 (98.85) | 42,211 | 29,342 (69.51) |
| **2018** | 611,009 | 313,493 (51.31) | 297,516 (48.69) | 23.81 | 20.34 | 18.77 | 18.64 | 23.81 | 10,150 | 100 (0.98) | 10,050 (99.02) | 41,433 | 28,639 (69.12) |
| **2019** | 642,292 | 329,670 (51.33) | 312,622 (46.67) | 23.86 | 20.30 | 18.80 | 18.59 | 23.86 | 12,562 | 81 (0.64) | 12,481 (99.36) | 39,830 | 26,696 (67.02) |
| **2020** | 669,560 | 343,844 (51.35) | 325,716 (48.65) | 23.89 | 20.28 | 18.86 | 18.54 | 23.89 | 11,440 | 91 (0.79) | 11,349 (99.21) | 35,881 | 25,727 (71.70) |
| **2021** | 694,001 | 356,678 (51.39) | 337,323 (48.61) | 23.89 | 20.24 | 18.90 | 18.58 | 23.89 | 13,062 | 79 (0.60) | 12,983 (99.40) | 39,552 | 25,993 (65.72) |
| **2022** | 720,491 | 370,805 (51.47) | 349,686 (48.53) | 23.78 | 20.19 | 19.05 | 18.58 | 23.78 | 13,553 | 82 (0.61) | 13,471 (99.39) | 43,796 | 25,948 (59.25) |

**Suppl.Table 4**

Number of male / female individuals and number of admissions for each concept.

|  | **SMC** | | | | **SMYC** | | | |
| --- | --- | --- | --- | --- | --- | --- | --- | --- |
| **Concept** | **N of male individuals (WLGP or PEDW)** | **N of female individuals (PEDW or WLGP)** | **N of admissions for concept (WLGP)** | **N of admissions for concept (PEDW)** | **N of male individuals (WLGP or PEDW)** | **N of female individuals (WLGP or PEDW)** | **N of admissions for concept (WLGP)** | **N of admissions for concept (PEDW)** |
| Addison's disease | 1,819 | 2,485 | 5,288 | 13,883 | 110 | 80 | 210 | 260 |
| Anaemia | 75,178 | 136,276 | 2,854,711 | 43,691 | 860 | 3,020 | 20,720 | 70 |
| Aneurysm | 30,231 | 11,975 | 43,718 | 83,306 | 40 | 30 | 40 | 60 |
| Ankylosing Spondylitis | 5,876 | 2,818 | 11,596 | 17,459 | 10 | 10 | 30 | 10 |
| Anxiety | 421,188 | 711,418 | 2,761,797 | 393,537 | 19,380 | 34,510 | 85,470 | 10,690 |
| Arrhythmia | 294,364 | 285,688 | 899,258 | 1,365,715 | 3,840 | 3,430 | 6,640 | 5,080 |
| Asthma | 436,608 | 491,065 | 7,657,263 | 1,243,152 | 59,010 | 44,830 | 618,420 | 68,940 |
| Atopic Eczema | 352,562 | 421,954 | 1,575,992 | 5,486 | 98,710 | 91,260 | 403,530 | 1,130 |
| Autism and ADHD | 46,209 | 17,825 | 92,767 | 42,716 | 22,680 | 7,460 | 40,690 | 17,920 |
| Bipolar Disorder | 12,190 | 19,209 | 45,588 | 64,734 | 70 | 210 | 200 | 240 |
| Blindness and low vision | 29,722 | 39,555 | 100,881 | 22,806 | 1,000 | 860 | 2,590 | 1,260 |
| Breast Cancer | 1,224 | 113,038 | 121,975 | 965,366 | 10 | 460 | 560 | 20 |
| Bronchiectasis | 14,744 | 16,377 | 38,469 | 72,452 | 190 | 130 | 380 | 70 |
| Chronic Back Pain | 27,762 | 37,263 | 135,174 | // | 90 | 110 | 220 | // |
| Chronic Fatigue Syndrome | 24,647 | 30,621 | 78,743 | // | 380 | 490 | 1,150 | // |
| Chronic Liver Disease | 46,417 | 35,886 | 86,452 | 174,139 | 360 | 300 | 620 | 1,660 |
| Chronic Pain | 35,937 | 51,797 | 165,087 | 17,724 | 180 | 350 | 600 | 330 |
| Chronic Sinusitis | 29,457 | 46,726 | 105,900 | // | 540 | 580 | 1,320 | // |
| Chronic Kidney Disease (CKD) Stage 3_5 | 101,539 | 129,334 | 545,909 | 345,884 | 160 | 80 | 360 | 5,200 |
| Coeliac Disease | 6,680 | 13,505 | 26,577 | 38,808 | 780 | 1,310 | 2,600 | 1,930 |
| Colon Cancer | 39,596 | 32,318 | 62,307 | 416,141 | 30 | 40 | 10 | 100 |
| Congenital disease and chromosomal abnormalities | 26,786 | 24,591 | 42,443 | 70,887 | 14,870 | 11,990 | 16,810 | 42,760 |
| COPD (Chronic obstructive pulmonary disease) | 192,803 | 189,528 | 1,939,227 | 1,008,129 | 5,090 | 3,320 | 9,530 | 890 |
| Coronary heart disease | 274,050 | 218,947 | 1,392,422 | 1,732,122 | 390 | 330 | 690 | 390 |
| Cystic fibrosis | 689 | 1,106 | 5,181 | 19,498 | 220 | 180 | 1,020 | 3,850 |
| Deafness | 233,517 | 229,120 | 718,602 | // | 21,950 | 18,850 | 56,120 | // |
| Dementia alzheimer | 61,089 | 104,528 | 370,160 | 314,632 | 10 | 20 | 10 | 10 |
| Depression | 451,108 | 698,921 | 4,006,408 | 621,124 | 13,500 | 24,760 | 75,250 | 7,640 |
| Diabetes Type 1 | 30,827 | 25,663 | 79,215 | 216,754 | 1,480 | 1,340 | 3,950 | 9,670 |
| Diabetes Type 2 | 211,979 | 173,817 | 1,332,350 | 1,603,897 | 130 | 160 | 250 | 270 |
| Diabetic retinopathy | 76,158 | 57,434 | 351,522 | 120,183 | 200 | 210 | 620 | 70 |
| Dialysis | 8,082 | 4,931 | 14,090 | 319,702 | 80 | 70 | 170 | 4,240 |
| Diverticular Disease | 127,107 | 159,618 | 291,313 | 491,128 | 245,360 | 238,390 | 1,554,550 | // |
| Drug or alcohol misuse | 375,499 | 272,349 | 769,385 | 1,468,796 | 70 | 50 | 100 | 60 |
| Eating disorders | 4,110 | 24,927 | 45,481 | 12,652 | 3,690 | 3,820 | 6,060 | 4,830 |
| Ehlers Danlos Syndrome | 2,823 | 6,741 | 11,627 | // | 1,220 | 3,310 | 5,740 | 2,490 |
| Endometriosis | 39 | 30,451 | 51,833 | No PEDW data available | 1,620 | 1,720 | 3,860 | // |
| Epilepsy | 52,461 | 50,490 | 526,112 | 311,820 | 10 | 290 | 460 | // |
| Fibromyalgia | 5,514 | 39,935 | 75,012 | 55,132 | 4,440 | 3,730 | 17,630 | 26,050 |
| Glaucoma | 49,018 | 55,239 | 157,411 | 152,108 | 30 | 210 | 290 | 120 |
| Gout | 133,558 | 47,001 | 408,496 | 123,877 | 130 | 100 | 360 | 470 |
| Heart Valve Disorders | 98,338 | 99,546 | 218,429 | 416,839 | 40 | 20 | 90 | 10 |
| HF (Heart Failure) | 149,378 | 143,531 | 383,330 | 590,623 | 2,640 | 2,100 | 5,820 | 9,460 |
| HIV/AIDS | 0 | 0 | 0 | 0 | 640 | 510 | 870 | 1,790 |
| Hypertension | 536,164 | 566,459 | 3,389,947 | 3,531,075 | 430 | 370 | 970 | 1,100 |
| Hyperthyroidism | 11,379 | 44,722 | 81,067 | 57,427 | 110 | 290 | 540 | 370 |
| Hypothyroidism | 50,268 | 192,504 | 428,688 | 609,098 | 780 | 1,380 | 3,180 | 4,000 |
| IBD (Inflammatory bowel disease) | 26,638 | 30,031 | 116,043 | 234,526 | 670 | 500 | 1,500 | 8,150 |
| IBS (Irritable Bowel Syndrome) | 58,948 | 157,163 | 336,107 | // | 1,380 | 2,450 | 4,650 | // |
| ILD (Inflammatory Lung Disease) | 19,190 | 11,291 | 28,097 | 55,374 | 50 | 50 | 80 | 50 |
| Learning disability | 22,309 | 13,467 | 145,763 | // | 4,630 | 2,220 | 10,700 | // |
| Leukaemia | 15,596 | 12,028 | 22,352 | 291,177 | 360 | 240 | 620 | 27,590 |
| Lymphoma | 8,713 | 6,974 | 28,942 | // | 120 | 70 | 260 | // |
| Marfan Syndrome | 508 | 254 | 1,281 | // | 70 | 40 | 170 | // |
| Meniere's disease | 13,239 | 26,571 | 62,201 | // | 30 | 40 | 70 | // |
| Metastatic cancers | 81,795 | 82,920 | 58,810 | 1,354,577 | 110 | 100 | 60 | 6,090 |
| Multiple sclerosis | 3,473 | 7,967 | 26,555 | 76,537 | 20 | 40 | 60 | 0 |
| Osteoarthritis | 314,975 | 430,019 | 1,160,525 | 949,936 | 190 | 200 | 350 | 140 |
| Osteoporosis | 93,772 | 195,569 | 214,880 | 812,551 | 300 | 290 | 110 | 890 |
| Pancreatic disease | 6,206 | 3,886 | 9,087 | 29,866 | 20 | 30 | 30 | 230 |
| Paralysis | 32,652 | 32,684 | 22,914 | 136,276 | 980 | 690 | 1,330 | 5,730 |
| Parkinson's disease | 20,529 | 15,187 | 49,600 | 115,684 | 10 | 10 | 10 | 0 |
| PCOS (Polycystic ovary syndrome) | 51 | 36,678 | 55,874 | // | 0 | 1,240 | 1,520 | // |
| Plasmacell | 4,029 | 3,488 | 9,423 | 134,540 | 10 | 10 | 10 | 10 |
| PMR (Polymyalgia reheumatica) | 16,730 | 34,542 | 116,063 | 79,353 | 10 | 10 | 10 | 0 |
| Primary Lung Cancer | 30,387 | 24,748 | 42,979 | 280,280 | 30 | 10 | 30 | 100 |
| Prostate Cancer | 63,638 | 43 | 67,667 | 398,883 | 10 | 10 | 10 | 0 |
| Prostate disorders | 151,471 | 129 | 322,084 | // | 30 | 10 | 40 | // |
| Psoriasis | 70,848 | 80,034 | 354,750 | 237,866 | 2,040 | 2,750 | 7,580 | 3,040 |
| Psoriasis Arthritis | 5,820 | 6,578 | 20,335 | 22,165 | 20 | 40 | 50 | 110 |
| PTSD (Post-traumatic stress disorders) | 15,780 | 16,695 | 38,886 | 21,040 | 300 | 830 | 1,020 | 900 |
| Pheripheral vascular disease | 122,079 | 128,337 | 411,730 | 225,698 | 70 | 40 | 180 | 2,290 |
| Renal transplant | 2,716 | 1,665 | 21,562 | 41,356 | 50 | 80 | 120 | 60 |
| Rheumatoid Arthritis | 22,446 | 48,748 | 199,980 | 253,548 | 160 | 120 | 260 | 170 |
| Schizophrenia | 16,676 | 12,195 | 49,896 | 84,754 | 160 | 190 | 350 | 120 |
| Skin Cancer | 127,493 | 136,543 | 482,073 | 145,245 | 580 | 350 | 980 | 360 |
| Stroke | 1,107 | 5,325 | 9,068 | 18,840 | 400 | 270 | 600 | 390 |
| Systemic Lupus Erythematosus | 543 | 2,114 | 3,903 | 11,474 | 20 | 50 | 70 | 240 |
| Systemic sclerosis | 1,485 | 2,446 | 2,898 | 9,496 | 20 | 30 | 50 | 70 |
| Thalassaemia | 55,324 | 59,117 | 138,729 | 52,448 | 180 | 170 | 300 | 1,210 |
| TIA (Transient ischaemic attack) | 6,876 | 6,510 | 15,049 | 5,528 | 60 | 40 | 90 | 30 |
| Viral Hepatitis | 0 | 0 | 0 | 0 | 180 | 170 | 330 | 270 |
| VTD (Venous thrombotic disease) | 46,620 | 60,261 | 140,423 | 56,167 | 6,330 | 5,040 | 25,880 | // |

Note that the number related to SMYC have been rounded up to the closest 10 to avoid disclosure control.

1. If the code list is derived from [52], we add the folder(s) the code lists are extracted from.
