## Supplementary material for "Cohort profile for the creation of the SAIL MELD-B e-cohort (SMC) and SAIL MELD-B children and Young adult e-cohort (SMYC)": Suppl.Material2: Suppl.Material2 - Copy.html

a0.a.CONC\_tables\_draft1


### **ReadV2 codes**¶

---

#### CONCEPT: Addison's disease¶

| READ\_CODE | DESCRIPTION\_SAIL | SENSITIVE\_CODE |
| --- | --- | --- |
| A176. | Tuberculosis of adrenal glands - Addison's disease | 0 |
| C153. | Other corticoadrenal overactivity | 0 |
| C154. | Corticoadrenal insufficiency | 0 |
| C1540 | Acute adrenal insufficiency | 0 |
| C1541 | Addison's disease | 0 |
| C1545 | Postprocedural adrenocortical(-medullary) hypofunction | 0 |
| C1546 | Addisonian crisis | 0 |
| C154z | Corticoadrenal insufficiency NOS | 0 |
| C155. | Other adrenal hypofunction | 0 |
| C1550 | Adrenal medullary insufficiency | 0 |
| C155z | Other adrenal hypofunction NOS | 0 |
| F3950 | Myopathy due to Addison's disease | 0 |
| PK13. | Hypoplasia of adrenal gland | 0 |

---

#### CONCEPT: Anaemia¶

| READ\_CODE | DESCRIPTION\_SAIL | SENSITIVE\_CODE |
| --- | --- | --- |
| 1452. | H/O: Anaemia vit.B12 deficient | 0 |
| 2126H | Vitamin B12 deficiency resolved | 0 |
| 42T2. | Serum vitamin B12 low | 0 |
| 42U2. | Serum folate low | 0 |
| 42U8. | RBC folate low | 0 |
| 42UB. | Whole blood folate low | 0 |
| 42UD. | RBC folate abnormal | 0 |
| 66E.. | B12 deficiency monitoring | 0 |
| 66E3. | B12 injections started | 0 |
| 66E4. | B12 injections - at home | 0 |
| 66E5. | B12 injections - at surgery | 0 |
| 66E8. | B12 injections stopped | 0 |
| 66EZ. | B12 defic. monitoring NOS | 0 |
| 7L185 | Intramuscular injection of vitamin B12 | 0 |
| C2620 | Folic acid deficiency | 0 |
| C2621 | Vitamin B12 deficiency | 0 |
| D010. | Pernicious anaemia | 0 |
| D011. | Other vitamin B12 deficiency anaemias | 0 |
| D0110 | Vitamin B12 deficiency anaemia due to dietary causes | 0 |
| D0111 | Vitamin B12 deficiency anaemia due to malabsorption with proteinuria | 0 |
| D0112 | Transcobalamin II deficiency | 0 |
| D011X | Vitamin B12 deficiency anaemia, unspecified | 0 |
| D011z | Other vitamin B12 deficiency anaemia NOS | 0 |
| D012. | Folate-deficiency anaemia | 0 |
| D0120 | Congenital folate malabsorption anaemia | 0 |
| D0121 | Folate-deficiency anaemia due to dietary causes | 0 |
| D0122 | Folate-deficiency anaemia, drug induced | 0 |
| D0123 | Folate-deficiency anaemia due to malabsorption | 0 |
| D0124 | Folate-deficiency anaemia due to liver disorders | 0 |
| D012z | Folate-deficiency anaemia NOS | 0 |
| D0130 | Combined B12 and folate deficiency anaemia | 0 |
| Dyu01 | [X]Other dietary vitamin B12 deficiency anaemia | 0 |
| Dyu02 | [X]Other vitamin B12 deficiency anaemias | 0 |
| Dyu03 | [X]Other folate deficiency anaemias | 0 |
| Dyu06 | [X]Vitamin B12 deficiency anaemia, unspecified | 0 |
| F11x6 | Cerebral degeneration due to vitamin B12 deficiency | 0 |
| F3815 | Myasthenic syndrome due to pernicious anaemia | 0 |

---

#### CONCEPT: Aneurysm¶

| READ\_CODE | DESCRIPTION\_SAIL | SENSITIVE\_CODE |
| --- | --- | --- |
| 14AE. | H/O: aortic aneurysm | 0 |
| 585I0 | Abdominal aortic aneurysm screen ultrasound scan abnormal | 0 |
| 68B51 | Aortic aneurysm screening abnormal | 0 |
| 7A11. | Replacement of aneurysmal bifurcation of aorta | 0 |
| 7A110 | Emergency replacement of aneurysmal bifurcation of aorta by anastomosis of aorta to femoral artery | 0 |
| 7A111 | Replacement of aneurysmal bifurcation of aorta by anastomosis of aorta to femoral artery | 0 |
| 7A112 | Emergency replacement of aneurysmal bifurcation of aorta by anastomosis of aorta to iliac artery | 0 |
| 7A113 | Replacement of aneurysmal bifurcation of aorta by anastomosis of aorta to iliac artery | 0 |
| 7A11y | Other specified replacement of aneurysmal bifurcation of aorta | 0 |
| 7A11z | Replacement of aneurysmal bifurcation of aorta NOS | 0 |
| 7A13. | Emergency replacement of aneurysmal segment of aorta | 0 |
| 7A130 | Emergency replacement of aneurysmal segment of ascending aorta by anastomosis of aorta to aorta | 0 |
| 7A131 | Emergency replacement of aneurysmal segment of thoracic aorta by anastomosis of aorta to aorta | 0 |
| 7A132 | Emergency replacement of aneurysmal segment of suprarenal abdominal aorta by anastomosis of aorta to aorta | 0 |
| 7A133 | Emergency replacement of aneurysmal segment of infrarenal abdominal aorta by anastomosis of aorta to aorta | 0 |
| 7A134 | Emergency replacement of aneurysmal segment of abdominal aorta by anastomosis of aorta to aorta NEC | 0 |
| 7A13y | Other specified emergency replacement of aneurysmal segment of aorta | 0 |
| 7A13z | Emergency replacement of aneurysmal segment of aorta NOS | 0 |
| 7A14. | Other replacement of aneurysmal segment of aorta | 0 |
| 7A140 | Replacement of aneurysmal segment of ascending aorta by anastomosis of aorta to aorta NEC | 0 |
| 7A141 | Replacement of aneurysmal segment of thoracic aorta by anastomosis of aorta to aorta NEC | 0 |
| 7A142 | Replacement of aneurysmal segment of suprarenal abdominal aorta by anastomosis of aorta to aorta NEC | 0 |
| 7A143 | Replacement of aneurysmal segment of infrarenal abdominal aorta by anastomosis of aorta to aorta NEC | 0 |
| 7A144 | Replacement of aneurysmal segment of abdominal aorta by anastomosis of aorta to aorta NEC | 0 |
| 7A14y | Other specified other replacement of aneurysmal segment of aorta | 0 |
| 7A14z | Other replacement of aneurysmal segment of aorta NOS | 0 |
| 7A15. | Other emergency bypass of segment of aorta | 0 |
| 7A150 | Emergency bypass of segment of ascending aorta by anastomosis of aorta to aorta NEC | 0 |
| 7A151 | Emergency bypass of segment of thoracic aorta by anastomosis of aorta to aorta NEC | 0 |
| 7A152 | Emergency bypass of segment of suprarenal abdominal aorta by anastomosis of aorta to aorta NEC | 0 |
| 7A153 | Emergency bypass of segment of infrarenal abdominal aorta by anastomosis of aorta to aorta NEC | 0 |
| 7A154 | Emergency bypass of segment of abdominal aorta by anastomosis of aorta to aorta NEC | 0 |
| 7A15y | Other specified other emergency bypass of segment of aorta | 0 |
| 7A15z | Other emergency bypass of segment of aorta NOS | 0 |
| 7A180 | Plastic repair of aorta and end to end anastomosis of aorta | 0 |
| 7A194 | Operation on aneurysm of aorta NEC | 0 |
| 7A1B. | Transluminal operations on aneurysmal segment of aorta | 0 |
| 7A1B0 | Endovascular stenting of infrarenal abdominal aortic aneurysm | 0 |
| 7A1B1 | Endovascular stenting of suprarenal aortic aneurysm | 0 |
| 7A1B2 | Endovascular stenting of thoracic aortic aneurysm | 0 |
| 7A1B5 | Endovascular stenting of aorto-uniiliac aneurysm | 0 |
| 7A1B6 | Endovascular stenting for aortic aneurysm of bifurcation NEC | 0 |
| 7A1B7 | Endovascular stenting for aorto-uniiliac aneurysm | 0 |
| 7A1B8 | Endovascular insertion of stent for infrarenal abdominal aortic aneurysm | 0 |
| 7A1B9 | Endovascular insertion of stent for suprarenal aortic aneurysm | 0 |
| 7A1BA | Endovascular insertion of stent for thoracic aortic aneurysm | 0 |
| 7A1BC | Endovascular insertion of stent for aortic aneurysm of bifurcation NEC | 0 |
| 7A1BD | Endovascular insertion of stent for aorto-uniiliac aneurysm | 0 |
| 7A1By | Other specified transluminal operations on aneurysmal segment of aorta | 0 |
| 7A1Bz | Transluminal operations on aneurysmal segment of aorta NOS | 0 |
| 7A1C. | Transluminal insertion of stent graft for aneurysmal segment of aorta | 0 |
| 7A1C0 | Endovascular insertion of stent graft for infrarenal abdominal aortic aneurysm | 0 |
| 7A1C1 | Endovascular insertion of stent graft for suprarenal aortic aneurysm | 0 |
| 7A1C2 | Endovascular insertion of stent graft for thoracic aortic aneurysm | 0 |
| 7A1C5 | Endovascular insertion of stent graft for aorto-uniiliac aneurysm | 0 |
| 7A1Cy | Other specified transluminal insertion of stent graft for aneurysmal segment of aorta | 0 |
| 7A1Cz | Transluminal insertion of stent graft for aneurysmal segment of aorta NOS | 0 |
| A930. | Syphilitic aortic aneurysm | 1 |
| G71.. | Aortic aneurysm | 0 |
| G710. | Dissecting aortic aneurysm | 0 |
| G711. | Thoracic aortic aneurysm which has ruptured | 0 |
| G712. | Thoracic aortic aneurysm without mention of rupture | 0 |
| G713. | Abdominal aortic aneurysm which has ruptured | 0 |
| G7130 | Ruptured suprarenal aortic aneurysm | 0 |
| G714. | Abdominal aortic aneurysm without mention of rupture | 0 |
| G7140 | Juxtarenal aortic aneurysm | 0 |
| G7141 | Inflammatory abdominal aortic aneurysm | 0 |
| G7142 | Infrarenal abdominal aortic aneurysm | 0 |
| G7143 | Aneurysm of suprarenal aorta | 0 |
| G715. | Ruptured aortic aneurysm NOS | 0 |
| G7150 | Thoracoabdominal aortic aneurysm, ruptured | 0 |
| G716. | Aortic aneurysm without mention of rupture NOS | 0 |
| G7160 | Thoracoabdominal aortic aneurysm, without mention of rupture | 0 |
| G717. | Aortic aneurysm - syphilitic | 1 |
| G718. | Leaking abdominal aortic aneurysm | 0 |
| G71z. | Aortic aneurysm NOS | 0 |
| Gyu71 | [X]Aortic aneurysm of unspecified site, ruptured | 0 |
| Gyu72 | [X]Aortic aneurysm of unspecified site, nonruptured | 0 |

---

#### CONCEPT: Ankylosing Spondylitis¶

| READ\_CODE | DESCRIPTION\_SAIL | SENSITIVE\_CODE |
| --- | --- | --- |
| 2377. | O/E - ankyl.spondyl.chest def. | 0 |
| 388p. | BASDAI - Bath ankylosing spondylitis disease activity index | 0 |
| 388p0 | Bath Ankylosing Spondylitis Functional Index | 0 |
| 38QL. | Bath Ankylosing Spondylitis Metrology Index | 0 |
| N0401 | Other rheumatoid arthritis of spine | 0 |
| N0450 | Juvenile ankylosing spondylitis | 0 |
| N100. | Ankylosing spondylitis | 0 |

---

#### CONCEPT: Anxiety¶

| READ\_CODE | DESCRIPTION\_SAIL | SENSITIVE\_CODE |
| --- | --- | --- |
| 1466. | H/O: anxiety state | 0 |
| 146G. | H/O: agoraphobia | 0 |
| 1B13. | Anxiousness | 0 |
| 2126J | Anxiety resolved | 0 |
| 2258. | O/E - anxious | 0 |
| 225J. | O/E - panic attack | 0 |
| 285.. | Neurotic condition, insight present | 0 |
| 286.. | Poor insight into neurotic condition | 0 |
| 8G94. | Anxiety management training | 0 |
| 8HHp. | Referral for guided self-help for anxiety | 0 |
| 8IH53 | Referral for guided self-help for anxiety declined | 0 |
| 8IH57 | Referral for psychological management of anxiety declined | 0 |
| 8T23. | Referral for psychological management of anxiety | 0 |
| E2... | Neurotic, personality and other nonpsychotic disorders | 0 |
| E20.. | Neurotic disorders | 0 |
| E200. | Anxiety states | 0 |
| E2000 | Anxiety state unspecified | 0 |
| E2001 | Panic disorder | 0 |
| E2002 | Generalised anxiety disorder | 0 |
| E2003 | Anxiety with depression | 0 |
| E2004 | Chronic anxiety | 0 |
| E2005 | Recurrent anxiety | 0 |
| E200z | Anxiety state NOS | 0 |
| E201. | Hysteria | 0 |
| E2010 | Hysteria unspecified | 0 |
| E2011 | Hysterical blindness | 0 |
| E2012 | Hysterical deafness | 0 |
| E2013 | Hysterical tremor | 0 |
| E2014 | Hysterical paralysis | 0 |
| E2015 | Hysterical seizures | 0 |
| E2016 | Other conversion disorder | 0 |
| E2017 | Hysterical amnesia | 0 |
| E2018 | Hysterical fugue | 0 |
| E2019 | Multiple personality | 0 |
| E201A | Dissociative reaction unspecified | 0 |
| E201B | Compensation neurosis | 0 |
| E201C | Phantom pregnancy | 0 |
| E201z | Hysteria NOS | 0 |
| E202. | Phobic disorders | 0 |
| E2020 | Phobia unspecified | 0 |
| E2021 | Agoraphobia with panic attacks | 0 |
| E2022 | Agoraphobia without mention of panic attacks | 0 |
| E2023 | Social phobia, fear of eating in public | 0 |
| E2024 | Social phobia, fear of public speaking | 0 |
| E2025 | Social phobia, fear of public washing | 0 |
| E2026 | Acrophobia | 0 |
| E2027 | Animal phobia | 0 |
| E2028 | Claustrophobia | 0 |
| E2029 | Fear of crowds | 0 |
| E202A | Fear of flying | 0 |
| E202B | Cancer phobia | 0 |
| E202C | Dental phobia | 0 |
| E202D | Fear of death | 0 |
| E202E | Fear of pregnancy | 0 |
| E202z | Phobic disorder NOS | 0 |
| E203. | Obsessive-compulsive disorders | 0 |
| E2030 | Compulsive neurosis | 0 |
| E2031 | Obsessional neurosis | 0 |
| E203z | Obsessive-compulsive disorder NOS | 0 |
| E204. | Neurotic depression reactive type | 0 |
| E205. | Neurasthenia - nervous debility | 0 |
| E206. | Depersonalisation syndrome | 0 |
| E207. | Hypochondriasis | 0 |
| E20y. | Other neurotic disorders | 0 |
| E20y0 | Somatization disorder | 0 |
| E20y1 | Writer's cramp neurosis | 0 |
| E20y2 | Other occupational neurosis | 0 |
| E20y3 | Psychasthenic neurosis | 0 |
| E20yz | Other neurotic disorder NOS | 0 |
| E20z. | Neurotic disorder NOS | 0 |
| E21.. | Personality disorders | 0 |
| E210. | Paranoid personality disorder | 0 |
| E211. | Affective personality disorder | 0 |
| E2110 | Unspecified affective personality disorder | 0 |
| E2111 | Hypomanic personality disorder | 0 |
| E2112 | Depressive personality disorder | 0 |
| E2113 | Cyclothymic personality disorder | 0 |
| E211z | Affective personality disorder NOS | 0 |
| E212. | Schizoid personality disorder | 0 |
| E2120 | Unspecified schizoid personality disorder | 0 |
| E2121 | Introverted personality | 0 |
| E2122 | Schizotypal personality | 0 |
| E212z | Schizoid personality disorder NOS | 0 |
| E213. | Explosive personality disorder | 0 |
| E214. | Compulsive personality disorders | 0 |
| E2140 | Anankastic personality | 0 |
| E2141 | Obsessional personality | 0 |
| E214z | Compulsive personality disorder NOS | 0 |
| E215. | Histrionic personality disorders | 0 |
| E2150 | Unspecified histrionic personality disorder | 0 |
| E2151 | Munchausen's syndrome | 0 |
| E2152 | Emotionally unstable personality | 0 |
| E2153 | Psychoinfantile personality | 0 |
| E215z | Histrionic personality disorder NOS | 0 |
| E216. | Inadequate personality disorder | 0 |
| E217. | Antisocial or sociopathic personality disorder | 0 |
| E21y. | Other personality disorders | 0 |
| E21y0 | Narcissistic personality disorder | 0 |
| E21y1 | Avoidant personality disorder | 0 |
| E21y2 | Borderline personality disorder | 0 |
| E21y3 | Passive-aggressive personality disorder | 0 |
| E21y4 | Eccentric personality disorder | 0 |
| E21y5 | Immature personality disorder | 0 |
| E21y6 | Masochistic personality disorder | 0 |
| E21y7 | Psychoneurotic personality disorder | 0 |
| E21yz | Other personality disorder NOS | 0 |
| E21z. | Personality disorder NOS | 0 |
| E26.. | Physiological malfunction arising from mental factors | 0 |
| E260. | Psychogenic musculoskeletal symptoms | 0 |
| E2600 | Psychogenic paralysis | 0 |
| E2601 | Psychogenic torticollis | 0 |
| E260z | Psychogenic musculoskeletal symptoms NOS | 0 |
| E261. | Psychogenic respiratory symptoms | 0 |
| E2610 | Psychogenic air hunger | 0 |
| E2611 | Psychogenic cough | 0 |
| E2612 | Psychogenic hiccough | 0 |
| E2613 | Psychogenic hyperventilation | 0 |
| E2614 | Psychogenic yawning | 0 |
| E2615 | Psychogenic aphonia | 0 |
| E261z | Psychogenic respiratory symptom NOS | 0 |
| E262. | Psychogenic cardiovascular symptoms | 0 |
| E2620 | Cardiac neurosis | 0 |
| E2621 | Cardiovascular neurosis | 0 |
| E2622 | Neurocirculatory asthenia | 0 |
| E2623 | Psychogenic cardiovascular disorder | 0 |
| E262z | Psychogenic cardiovascular symptom NOS | 0 |
| E263. | Psychogenic skin symptoms | 0 |
| E2630 | Psychogenic pruritus | 0 |
| E263z | Psychogenic skin symptoms NOS | 0 |
| E264. | Psychogenic gastrointestinal tract symptoms | 0 |
| E2640 | Psychogenic aerophagy | 0 |
| E2642 | Cyclical vomiting - psychogenic | 0 |
| E2643 | Psychogenic diarrhoea | 0 |
| E2644 | Psychogenic dyspepsia | 0 |
| E2645 | Psychogenic constipation | 0 |
| E264z | Psychogenic gastrointestinal tract symptom NOS | 0 |
| E265. | Psychogenic genitourinary tract symptoms | 0 |
| E2650 | Psychogenic genitourinary tract malfunction unspecified | 0 |
| E2651 | Psychogenic vaginismus | 0 |
| E2652 | Psychogenic dysmenorrhea | 0 |
| E2653 | Psychogenic dysuria | 0 |
| E265z | Psychogenic genitourinary tract symptom NOS | 0 |
| E266. | Psychogenic endocrine malfunction | 0 |
| E267. | Psychogenic symptom of special sense organ | 0 |
| E26y. | Other psychogenic malfunction | 0 |
| E26y0 | Bruxism (teeth grinding) | 0 |
| E26yz | Other psychogenic malfunction NOS | 0 |
| E26z. | Psychosomatic disorder NOS | 0 |
| E278. | Psychalgia | 0 |
| E2780 | Psychogenic pain unspecified | 0 |
| E2781 | Tension headache | 0 |
| E2782 | Psychogenic backache | 0 |
| E278z | Psychalgia NOS | 0 |
| E28.. | Acute reaction to stress | 0 |
| E280. | Acute panic state due to acute stress reaction | 0 |
| E281. | Acute fugue state due to acute stress reaction | 0 |
| E282. | Acute stupor state due to acute stress reaction | 0 |
| E283. | Other acute stress reactions | 0 |
| E2830 | Acute situational disturbance | 0 |
| E2831 | Acute posttrauma stress state | 0 |
| E283z | Other acute stress reaction NOS | 0 |
| E284. | Stress reaction causing mixed disturbance of emotion and conduct | 0 |
| E28z. | Acute stress reaction NOS | 0 |
| E29.. | Adjustment reaction | 0 |
| E2900 | Grief reaction | 0 |
| E292. | Adjustment reaction with predominant disturbance of other emotions | 0 |
| E2920 | Separation anxiety disorder | 0 |
| E2921 | Adolescent emancipation disorder | 0 |
| E2922 | Early adult emancipation disorder | 0 |
| E2923 | Specific academic or work inhibition | 0 |
| E2924 | Adjustment reaction with anxious mood | 0 |
| E2925 | Culture shock | 0 |
| E292y | Adjustment reaction with mixed disturbance of emotion | 0 |
| E292z | Adjustment reaction with disturbance of other emotion NOS | 0 |
| E293. | Adjustment reaction with predominant disturbance of conduct | 0 |
| E2930 | Adjustment reaction with aggression | 0 |
| E2931 | Adjustment reaction with antisocial behaviour | 0 |
| E2932 | Adjustment reaction with destructiveness | 0 |
| E293z | Adjustment reaction with predominant disturbance of conduct NOS | 0 |
| E294. | Adjustment reaction with mixed disturbance of emotion and conduct | 0 |
| E29y. | Other adjustment reactions | 0 |
| E29y0 | Concentration camp syndrome | 0 |
| E29y1 | Other post-traumatic stress disorder | 0 |
| E29y2 | Adjustment reaction with physical symptoms | 0 |
| E29y3 | Elective mutism due to an adjustment reaction | 0 |
| E29y4 | Adjustment reaction due to hospitalisation | 0 |
| E29y5 | Other adjustment reaction with withdrawal | 0 |
| E29yz | Other adjustment reactions NOS | 0 |
| E29z. | Adjustment reaction NOS | 0 |
| E2D0. | Disturbance of anxiety and fearfulness in childhood and adolescence | 0 |
| E2D0z | Disturbance of anxiety and fearfulness in childhood and adolescence NOS | 0 |
| Eu054 | [X]Organic anxiety disorder | 0 |
| Eu341 | [X]Dysthymia | 0 |
| Eu4.. | [X]Neurotic, stress - related and somoform disorders | 0 |
| Eu40. | [X]Phobic anxiety disorders | 0 |
| Eu400 | [X]Agoraphobia | 0 |
| Eu401 | [X]Social phobias | 0 |
| Eu402 | [X]Specific (isolated) phobias | 0 |
| Eu403 | [X]Needle phobia | 0 |
| Eu40y | [X]Other phobic anxiety disorders | 0 |
| Eu40z | [X]Phobic anxiety disorder, unspecified | 0 |
| Eu41. | [X]Other anxiety disorders | 0 |
| Eu410 | [X]Panic disorder [episodic paroxysmal anxiety] | 0 |
| Eu411 | [X]Generalized anxiety disorder | 0 |
| Eu412 | [X]Mixed anxiety and depressive disorder | 0 |
| Eu413 | [X]Other mixed anxiety disorders | 0 |
| Eu41y | [X]Other specified anxiety disorders | 0 |
| Eu41z | [X]Anxiety disorder, unspecified | 0 |
| Eu42. | [X]Obsessive - compulsive disorder | 0 |
| Eu420 | [X]Predominantly obsessional thoughts or ruminations | 0 |
| Eu421 | [X]Predominantly compulsive acts [obsessional rituals] | 0 |
| Eu422 | [X]Mixed obsessional thoughts and acts | 0 |
| Eu42y | [X]Other obsessive-compulsive disorders | 0 |
| Eu42z | [X]Obsessive-compulsive disorder, unspecified | 0 |
| Eu43. | [X]Reaction to severe stress, and adjustment disorders | 0 |
| Eu430 | [X]Acute stress reaction | 0 |
| Eu431 | [X]Post - traumatic stress disorder | 0 |
| Eu432 | [X]Adjustment disorders | 0 |
| Eu43y | [X]Other reactions to severe stress | 0 |
| Eu43z | [X]Reaction to severe stress, unspecified | 0 |
| Eu44. | [X]Dissociative [conversion] disorders | 0 |
| Eu440 | [X]Dissociative amnesia | 0 |
| Eu441 | [X]Dissociative fugue | 0 |
| Eu442 | [X]Dissociative stupor | 0 |
| Eu443 | [X]Trance and possession disorders | 0 |
| Eu444 | [X]Dissociative motor disorders | 0 |
| Eu445 | [X]Dissociative convulsions | 0 |
| Eu446 | [X]Dissociative anaesthesia and sensory loss | 0 |
| Eu447 | [X]Mixed dissociative [conversion] disorders | 0 |
| Eu44y | [X]Other dissociative [conversion] disorders | 0 |
| Eu44z | [X]Dissociative [conversion] disorder, unspecified | 0 |
| Eu45. | [X]Somatoform disorders | 0 |
| Eu450 | [X]Somatization disorder | 0 |
| Eu451 | [X]Undifferentiated somatoform disorder | 0 |
| Eu452 | [X]Hypochondriacal disorder | 0 |
| Eu453 | [X]Somatoform autonomic dysfunction | 0 |
| Eu454 | [X]Persistent somatoform pain disorder | 0 |
| Eu455 | [X]Globus pharyngeus | 0 |
| Eu45y | [X]Other somatoform disorders | 0 |
| Eu45z | [X]Somatoform disorder, unspecified | 0 |
| Eu46. | [X]Other neurotic disorders | 0 |
| Eu460 | [X]Neurasthenia | 0 |
| Eu461 | [X]Depersonalization - derealization syndrome | 0 |
| Eu46y | [X]Other specified neurotic disorders | 0 |
| Eu46z | [X]Neurotic disorder, unspecified | 0 |
| Eu515 | [X]Nightmares | 0 |
| Eu606 | [X]Anxious [avoidant] personality disorder | 0 |
| Eu930 | [X]Separation anxiety disorder of childhood | 0 |
| Eu931 | [X]Phobic anxiety disorder of childhood | 0 |
| Eu932 | [X]Social anxiety disorder of childhood | 0 |
| Eu93y | [X]Other childhood emotional disorders | 0 |
| M240E | Alopecia neurotica | 0 |

---

#### CONCEPT: Arrhythmia¶

| READ\_CODE | DESCRIPTION\_SAIL | SENSITIVE\_CODE |
| --- | --- | --- |
| 14AD. | H/O ventricular fibrillation | 0 |
| 14AN. | H/O: atrial fibrillation | 0 |
| 14AP. | History of ventricular tachycardia | 0 |
| 14AQ. | History of supraventricular tachycardia | 0 |
| 14AR. | History of atrial flutter | 0 |
| 14O71 | At risk of atrial fibrillation | 0 |
| 14V1. | H/O: cardiac pacemaker in situ | 0 |
| 1R00. | For attempted cardiopulmonary resuscitation | 0 |
| 1R10. | Not for attempted CPR (cardiopulmonary resuscitation) | 0 |
| 212R. | Atrial fibrillation resolved | 0 |
| 2241. | O/E - collapse -cardiac arrest | 0 |
| 2JS.. | Patient with internal cardiac defibrillator pacemaker | 0 |
| 32130 | Exercise ECG normal | 0 |
| 32131 | Exercise ECG abnormal | 0 |
| 32141 | Ambulatory ECG abnormal | 0 |
| 3217. | ECG abnormal | 0 |
| 3218. | ECG - improved | 0 |
| 3219. | ECG equivocal | 0 |
| 321C. | ECG sinus rhythm | 0 |
| 321C0 | ECG: sinus bradycardia | 0 |
| 321C1 | ECG: sinus tachycardia | 0 |
| 322.. | ECG: myocardial ischaemia | 0 |
| 3222. | ECG:shows myocardial ischaemia | 0 |
| 322Z. | ECG: myocardial ischaemia NOS | 0 |
| 323.. | ECG: myocardial infarction | 0 |
| 3232. | ECG: old myocardial infarction | 0 |
| 3233. | ECG: antero-septal infarct. | 0 |
| 3234. | ECG:posterior/inferior infarct | 0 |
| 3235. | ECG: subendocardial infarct | 0 |
| 3236. | ECG: lateral infarction | 0 |
| 323Z. | ECG: myocardial infarct NOS | 0 |
| 324.. | ECG:left ventricle hypertrophy | 0 |
| 3242. | ECG: shows LVH | 0 |
| 324Z. | ECG: LVH NOS | 0 |
| 325.. | ECG:right ventricle hypertrop. | 0 |
| 3252. | ECG: shows RVH | 0 |
| 325Z. | ECG: RVH NOS | 0 |
| 326.. | ECG: ectopic beats | 0 |
| 3262. | ECG: extrasystole | 0 |
| 3263. | ECG: ventricular ectopics | 0 |
| 3264. | ECG: atrial ectopics | 0 |
| 326Z. | ECG: ectopic beats NOS | 0 |
| 327.. | ECG: supraventricular arrhythmia | 0 |
| 3272. | ECG: atrial fibrillation | 0 |
| 3273. | ECG: atrial flutter | 0 |
| 3274. | ECG: paroxysmal atrial tachy. | 0 |
| 327Z. | ECG: supraventric. arryth. NOS | 0 |
| 328.. | ECG: ventricular arrhythmia | 0 |
| 3282. | ECG: ventricular tachycardia | 0 |
| 3283. | ECG: ventricular fibrillation | 0 |
| 328Z. | ECG: ventricular arrhythmia NOS | 0 |
| 329.. | ECG: heart block | 0 |
| 3292. | ECG: partial sinu-atrial block | 0 |
| 3293. | ECG:complete sinu-atrial block | 0 |
| 3294. | ECG:partial A-V block-long P-R | 0 |
| 3295. | ECG: partial A-V block - 2:1 | 0 |
| 3296. | ECG: partial A-V block - 3:1 | 0 |
| 3297. | ECG: Wenckebach phenomenon | 0 |
| 3298. | ECG: complete A-V block | 0 |
| 3299. | ECG: right bundle branch block | 0 |
| 329A. | ECG: left bundle branch block | 0 |
| 329B. | ECG: trifascicular block | 0 |
| 329C. | ECG: bifascicular block | 0 |
| 329D. | ECG: left anterior fascicular block | 0 |
| 329E. | ECG: left posterior fascicular block | 0 |
| 329F. | ECG: right bundle branch and left anterior fascicular block | 0 |
| 329G. | ECG: right bundle branch and left posterior fascicular block | 0 |
| 329H. | Electrocardiogram: Mobitz type 2 second degree atrioventricular block | 0 |
| 329Z. | ECG: heart block NOS | 0 |
| 32A2. | ECG: P wave abnormal | 0 |
| 32A3. | ECG: P mitrale | 0 |
| 32A4. | ECG: P pulmonale | 0 |
| 32B.. | ECG: Q wave | 0 |
| 32B2. | ECG: Q wave abnormal | 0 |
| 32B3. | ECG: Q wave pathological | 0 |
| 32BZ. | ECG: Q wave NOS | 0 |
| 32C.. | ECG: R wave | 0 |
| 32C2. | ECG: R wave abnormal | 0 |
| 32C3. | ECG: R wave tall | 0 |
| 32CZ. | ECG: R wave NOS | 0 |
| 32D2. | ECG: S wave abnormal | 0 |
| 32D3. | ECG: S wave deep | 0 |
| 32E2. | ECG: S-T interval abnormal | 0 |
| 32E3. | ECG: S-T elevation | 0 |
| 32E4. | ECG: S-T depression | 0 |
| 32F2. | ECG: T wave abnormal | 0 |
| 32F3. | ECG: T wave flattened | 0 |
| 32F4. | ECG: T wave inverted | 0 |
| 32FZ. | ECG: T wave NOS | 0 |
| 32G2. | ECG: U wave abnormal | 0 |
| 32G3. | ECG: U wave exaggerated | 0 |
| 32GZ. | ECG: U wave NOS | 0 |
| 32H.. | ECG: F wave | 0 |
| 32H1. | ECG: F wave absent | 0 |
| 32H2. | ECG: F wave present | 0 |
| 32HZ. | ECG: F wave NOS | 0 |
| 32I2. | ECG: P-R interval abnormal | 0 |
| 32I3. | ECG: P-R interval prolonged | 0 |
| 32I4. | ECG: P-R interval shortened | 0 |
| 32J2. | ECG: QRS complex abnormal | 0 |
| 32J3. | ECG: QRS complex prolonged | 0 |
| 32J4. | ECG: QRS complex shortened | 0 |
| 32J5. | Left axis deviation | 0 |
| 32J6. | Right axis deviation | 0 |
| 32K2. | ECG: Q-T interval abnormal | 0 |
| 32K3. | ECG: Q-T interval prolonged | 0 |
| 32K4. | ECG: Q-T interval shortened | 0 |
| 32L.. | ECG: left ventricular strain | 0 |
| 662S. | Atrial fibrillation monitoring | 0 |
| 6A9.. | Atrial fibrillation annual review | 0 |
| 79321 | Open chest massage of heart | 0 |
| 7936. | Introduction of cardiac pacemaker system via vein | 0 |
| 79360 | Implantation of intravenous cardiac pacemaker system | 0 |
| 79361 | Resiting of lead of intravenous pacemaker system | 0 |
| 79362 | Maintenance of battery of intravenous cardiac pacemaker system | 0 |
| 79363 | Maintenance of intravenous cardiac pacemaker system NEC | 0 |
| 79364 | Removal of intravenous cardiac pacemaker system | 0 |
| 79365 | Implantation of emergency intravenous cardiac pacemaker | 0 |
| 79366 | Implantation of permanent intravenous cardiac pacemaker | 0 |
| 79367 | Implantation of intravenous fixed-rate cardiac pacemaker | 0 |
| 79368 | Implantation of intravenous triggered cardiac pacemaker | 0 |
| 79369 | Implantation of intravenous atrial overdrive pacemaker | 0 |
| 7936A | Implantation of intravenous pacemaker for atrial fibrillation | 0 |
| 7936B | Implantation of simple one wire intravenous cardiac pacemaker | 0 |
| 7936C | Implantation of complex one wire intravenous cardiac pacemaker | 0 |
| 7936D | Implantation of complex two wire intravenous cardiac pacemaker | 0 |
| 7936E | Implantation of intravenous dual chamber permanent pacemaker | 0 |
| 7936F | Renewal of intravenous cardiac pacemaker system | 0 |
| 7936G | Implantation of intravenous single chamber cardiac pacemaker system | 0 |
| 7936H | Implantation of intravenous dual chamber cardiac pacemaker system | 0 |
| 7936J | Implantation of intravenous biventricular cardiac pacemaker system | 0 |
| 7936K | Implantation of intravenous cardiac pacemaker system NEC | 0 |
| 7936y | Other specified cardiac pacemaker system introduced via vein | 0 |
| 7936z | Cardiac pacemaker system introduced via vein NOS | 0 |
| 7937. | Other cardiac pacemaker system | 0 |
| 79370 | Implantation of cardiac pacemaker system NEC | 0 |
| 79371 | Resiting of lead of cardiac pacemaker system NEC | 0 |
| 79372 | Maintenance of battery of cardiac pacemaker system NEC | 0 |
| 79373 | Maintenance of cardiac pacemaker system NEC | 0 |
| 79374 | Removal of cardiac pacemaker system NEC | 0 |
| 79375 | Implantation of internal cardiac defibrillator | 0 |
| 79376 | Removal of internal cardiac defibrillator | 0 |
| 79377 | Implantation of single chamber cardiac pacemaker system | 0 |
| 79378 | Implantation of dual chamber cardiac pacemaker system | 0 |
| 79379 | Implantation of biventricular cardiac pacemaker system | 0 |
| 7937y | Other specified other cardiac pacemaker system | 0 |
| 7937z | Other cardiac pacemaker system NOS | 0 |
| 793F. | Cardioverter defibrillator introduced through the vein | 0 |
| 793F0 | Implantation of cardioverter defibrillator using one electrode lead | 0 |
| 793F1 | Implantation of cardioverter defibrillator using two electrode leads | 0 |
| 793F2 | Resiting of lead of cardioverter defibrillator | 0 |
| 793F3 | Renewal of cardioverter defibrillator | 0 |
| 793F4 | Removal of cardioverter defibrillator | 0 |
| 793F5 | Implantation of cardioverter defibrillator using three electrode leads | 0 |
| 793Fy | Other specified cardioverter defibrillator introduced through the vein | 0 |
| 793Fz | Cardioverter defibrillator introduced through the vein NOS | 0 |
| 793M1 | Percutaneous transluminal ablation of atrial wall for atrial flutter | 0 |
| 793M2 | Percutaneous transluminal internal cardioversion NEC | 0 |
| 793M3 | Percutaneous transluminal ablation of conducting system of heart for atrial flutter NEC | 0 |
| 793P. | Other cardioverter defibrillator | 0 |
| 793P0 | Implantation of subcutaneous cardioverter defibrillator | 0 |
| 793P1 | Resiting of lead of subcutaneous cardioverter defibrillator | 0 |
| 793P2 | Renewal of subcutaneous cardioverter defibrillator | 0 |
| 793P3 | Removal of subcutaneous cardioverter defibrillator | 0 |
| 793Py | Other specified other cardioverter defibrillator | 0 |
| 793Pz | Unspecified other cardioverter defibrillator | 0 |
| 7L1H. | External resuscitation | 0 |
| 7L1H0 | Direct current cardioversion | 0 |
| 7L1H1 | External cardioversion NEC | 0 |
| 7L1H2 | Internal electrode cardioversion | 0 |
| 7L1H6 | Advanced cardiopulmonary resuscitation | 0 |
| 7L1H7 | External ventricular defibrillation | 0 |
| 7L1H8 | Chemical cardioversion | 0 |
| 7P19. | Pacemaker testing | 0 |
| 7P190 | Distant pacemaker test | 0 |
| 7P19y | Other specified pacemaker testing | 0 |
| 7P19z | Pacemaker testing NOS | 0 |
| 7P1D. | Evaluation of cardioverter defibrillator | 0 |
| 840.. | Direct current cardioversion planned | 0 |
| 853.. | Cardiac massage - external | 0 |
| 8531. | Closed cardiac massage alone | 0 |
| 8532. | Closed cardiac massage+ventil. | 0 |
| 853Z. | External cardiac massage NOS | 0 |
| 88A9. | Deactivation of automatic implantable cardioverter defibrillator | 0 |
| 88AA. | Automatic implantable cardioverter defibrillator check | 0 |
| 8HRF. | Referral for cardiac pacemaker check | 0 |
| 8HTy. | Referral to atrial fibrillation clinic | 0 |
| 8OAD. | Provision of written information about atrial fibrillation | 0 |
| 9N2b. | Seen by cardiac pacemaker technician | 0 |
| F2560 | Hypsarrhythmia | 0 |
| G559. | Arrhythmogenic right ventricular cardiomyopathy | 0 |
| G56.. | Conduction disorders | 0 |
| G560. | Complete atrioventricular block | 0 |
| G561. | Partial atrioventricular block | 0 |
| G5610 | Atrioventricular block unspecified | 0 |
| G5611 | First degree atrioventricular block | 0 |
| G5612 | Mobitz type II atrioventricular block | 0 |
| G5613 | Mobitz type I (Wenckebach) atrioventricular block | 0 |
| G5614 | Second degree atrioventricular block | 0 |
| G561z | Atrioventricular block NOS | 0 |
| G562. | Left bundle branch hemiblock | 0 |
| G5620 | Left anterior fascicular block | 0 |
| G5621 | Left posterior fascicular block | 0 |
| G562z | Left bundle branch hemiblock NOS | 0 |
| G563. | Left main stem bundle branch block | 0 |
| G564. | Right bundle branch block | 0 |
| G565. | Other bundle branch block | 0 |
| G5650 | Bundle branch block unspecified | 0 |
| G5651 | Right bundle branch block with left posterior fascicular block | 0 |
| G5652 | Right bundle branch block with left anterior fascicular block | 0 |
| G5653 | Other bilateral bundle branch block | 0 |
| G5654 | Trifascicular block | 0 |
| G5655 | Bifascicular block | 0 |
| G565z | Other bundle branch block NOS | 0 |
| G566. | Other heart block | 0 |
| G5660 | Sinoatrial block | 0 |
| G5661 | Interventricular block NOS | 0 |
| G5662 | Right fascicular block | 0 |
| G566z | Other heart block NOS | 0 |
| G567. | Anomalous atrioventricular excitation | 0 |
| G5670 | Accelerated atrioventricular conduction | 0 |
| G5671 | Accessory atrioventricular conduction | 0 |
| G5672 | Pre-excitation atrioventricular conduction | 0 |
| G5673 | Ventricular pre-excitation | 0 |
| G5674 | Wolff-Parkinson-White syndrome | 0 |
| G567z | Anomalous atrioventricular excitation NOS | 0 |
| G56y. | Other conduction disorders | 0 |
| G56y0 | Lown-Ganong-Levine syndrome | 0 |
| G56y1 | Atrioventricular dissociation | 0 |
| G56y2 | Romano - Ward syndrome | 0 |
| G56y3 | Jervell and Lange-Nielsen syndrome | 0 |
| G56y4 | Right fascicular block | 0 |
| G56y5 | Long Q-T syndrome | 0 |
| G56y6 | Pacemaker twiddler's syndrome | 0 |
| G56yz | Other conduction disorders NOS | 0 |
| G56z. | Conduction disorders unspecified | 0 |
| G56z0 | Stokes-Adams syndrome | 0 |
| G56zz | Conduction disorders NOS | 0 |
| G57.. | Cardiac dysrhythmias | 0 |
| G570. | Paroxysmal supraventricular tachycardia | 0 |
| G5700 | Paroxysmal atrial tachycardia | 0 |
| G5701 | Paroxysmal atrioventricular tachycardia | 0 |
| G5702 | Paroxysmal junctional tachycardia | 0 |
| G5703 | Paroxysmal nodal tachycardia | 0 |
| G570z | Paroxysmal supraventricular tachycardia NOS | 0 |
| G571. | Paroxysmal ventricular tachycardia | 0 |
| G572. | Paroxysmal tachycardia unspecified | 0 |
| G5720 | Essential paroxysmal tachycardia | 0 |
| G5721 | Bouveret-Hoffmann syndrome | 0 |
| G572z | Paroxysmal tachycardia NOS | 0 |
| G573. | Atrial fibrillation and flutter | 0 |
| G5730 | Atrial fibrillation | 0 |
| G5731 | Atrial flutter | 0 |
| G5732 | Paroxysmal atrial fibrillation | 0 |
| G5733 | Non-rheumatic atrial fibrillation | 0 |
| G5734 | Permanent atrial fibrillation | 0 |
| G5735 | Persistent atrial fibrillation | 0 |
| G5736 | Paroxysmal atrial flutter | 0 |
| G5737 | Chronic atrial fibrillation | 0 |
| G5738 | Typical atrial flutter | 0 |
| G5739 | Atypical atrial flutter | 0 |
| G573z | Atrial fibrillation and flutter NOS | 0 |
| G574. | Ventricular fibrillation and flutter | 0 |
| G5740 | Ventricular fibrillation | 0 |
| G5741 | Ventricular flutter | 0 |
| G574z | Ventricular fibrillation and flutter NOS | 0 |
| G575. | Cardiac arrest | 0 |
| G5750 | Cardiac arrest with successful resuscitation | 0 |
| G5751 | Sudden cardiac death, so described | 0 |
| G5752 | Electromechanical dissociation with successful resuscitation | 0 |
| G5753 | Electromechanical dissociation | 0 |
| G575z | Cardiac arrest, unspecified | 0 |
| G576. | Ectopic beats | 0 |
| G5760 | Ectopic beats unspecified | 0 |
| G5761 | Supraventricular ectopic beats | 0 |
| G5762 | Ventricular ectopic beats | 0 |
| G5763 | Atrial premature depolarization | 0 |
| G5764 | Junctional premature depolarization | 0 |
| G5765 | Ventricular premature depolarization | 0 |
| G576z | Ectopic beats NOS | 0 |
| G577. | Sinus arrhythmia | 0 |
| G578. | Atrial standstill | 0 |
| G57y. | Other cardiac dysrhythmias | 0 |
| G57y0 | Persistent sinus bradycardia | 0 |
| G57y1 | Severe sinus bradycardia | 0 |
| G57y2 | Brugada syndrome | 0 |
| G57y3 | Sick sinus syndrome | 0 |
| G57y4 | Sinoatrial node dysfunction NOS | 0 |
| G57y5 | Wandering atrial pacemaker | 0 |
| G57y6 | Nodal rhythm disorder | 0 |
| G57y7 | Sinus tachycardia | 0 |
| G57y8 | Bigeminal pulse | 0 |
| G57y9 | Supraventricular tachycardia NOS | 0 |
| G57yA | Re-entry ventricular arrhythmia | 0 |
| G57yz | Other cardiac dysrhythmia NOS | 0 |
| G57z. | Cardiac dysrhythmia NOS | 0 |
| Gyu5U | [X]Other and unspecified atrioventricular block | 0 |
| Gyu5W | [X]Other and unspecified right bundle-branch block | 0 |
| Gyu5X | [X]Other specified heart block | 0 |
| Gyu5Z | [X]Other and unspecified premature depolarization | 0 |
| Gyu5a | [X]Other specified cardiac arrhythmias | 0 |
| L0911 | Defibrination syndrome following abortive pregnancy | 1 |
| L09y1 | Cardiac arrest following abortive pregnancy | 1 |
| P6y5z | Congenital heart block NOS | 0 |
| PG57. | Infantile cortical hyperostosis | 0 |
| Q491. | Neonatal cardiac dysrhythmia | 0 |
| R0026 | [D]Asystolic vasovagal syncope | 0 |
| R1431 | [D]Electrocardiogram (ECG) abnormal | 0 |
| SP001 | Mechanical complication of cardiac pacemaker | 0 |
| SP110 | Cardiac arrest as a complication of care | 0 |
| TB010 | Implant of cardiac pacemaker as the cause of abnormal reaction of patient, or of later complication, without mention of misadventure at the time of operation | 0 |
| ZV450 | [V]Cardiac pacemaker in situ | 0 |
| ZV45M | [V]Biventricular pacemaker in situ | 0 |
| ZV530 | [V]Fitting or adjustment of device related to nervous system or special senses | 0 |
| ZV533 | [V]Fitting or adjustment of cardiac pacemaker | 0 |

---

#### CONCEPT: Asthma¶

| READ\_CODE | DESCRIPTION\_SAIL | SENSITIVE\_CODE |
| --- | --- | --- |
| 13Y4. | Asthma society member | 0 |
| 14B4. | H/O: asthma | 0 |
| 14Ok0 | At risk of severe asthma exacerbation | 0 |
| 173A. | Exercise induced asthma | 0 |
| 173c. | Occupational asthma | 0 |
| 173d. | Work aggravated asthma | 0 |
| 178.. | Asthma trigger | 0 |
| 1780. | Aspirin induced asthma | 0 |
| 1781. | Asthma trigger - pollen | 0 |
| 1782. | Asthma trigger - tobacco smoke | 0 |
| 1783. | Asthma trigger - warm air | 0 |
| 1784. | Asthma trigger - emotion | 0 |
| 1785. | Asthma trigger - damp | 0 |
| 1786. | Asthma trigger - animals | 0 |
| 1787. | Asthma trigger - seasonal | 0 |
| 1788. | Asthma trigger - cold air | 0 |
| 1789. | Asthma trigger - respiratory infection | 0 |
| 178A. | Asthma trigger - airborne dust | 0 |
| 178B. | Asthma trigger - exercise | 0 |
| 1J70. | Suspected asthma | 0 |
| 1O2.. | Asthma confirmed | 0 |
| 21262 | Asthma resolved | 0 |
| 212G. | Asthma resolved | 0 |
| 388t0 | Royal College of Physicians asthma assessment three questions score | 0 |
| 38B8. | Severe asthma exacerbation risk assessment | 0 |
| 38DL. | Asthma control test | 0 |
| 38DT. | Asthma control questionnaire | 0 |
| 38DV. | Mini asthma quality of life questionnaire | 0 |
| 38QM. | Childhood Asthma Control Test | 0 |
| 661M1 | Asthma self-management plan agreed | 0 |
| 661N1 | Asthma self-management plan review | 0 |
| 663.. | Respiratory disease monitoring | 0 |
| 6636. | Inhaler technique shown | 0 |
| 663I. | Inhaler technique - poor | 0 |
| 663N. | Asthma disturbing sleep | 0 |
| 663N0 | Asthma causing night waking | 0 |
| 663N1 | Asthma disturbs sleep weekly | 0 |
| 663N2 | Asthma disturbs sleep frequently | 0 |
| 663O. | Asthma not disturbing sleep | 0 |
| 663O0 | Asthma never disturbs sleep | 0 |
| 663P. | Asthma limiting activities | 0 |
| 663P0 | Asthma limits activities 1 to 2 times per month | 0 |
| 663P1 | Asthma limits activities 1 to 2 times per week | 0 |
| 663P2 | Asthma limits activities most days | 0 |
| 663Q. | Asthma not limiting activities | 0 |
| 663U. | Asthma management plan given | 0 |
| 663V. | Asthma severity | 0 |
| 663V0 | Occasional asthma | 0 |
| 663V1 | Mild asthma | 0 |
| 663V2 | Moderate asthma | 0 |
| 663V3 | Severe asthma | 0 |
| 663W. | Asthma prophylactic medication used | 0 |
| 663d. | Emergency asthma admission since last appointment | 0 |
| 663e. | Asthma restricts exercise | 0 |
| 663e0 | Asthma sometimes restricts exercise | 0 |
| 663e1 | Asthma severely restricts exercise | 0 |
| 663f. | Asthma never restricts exercise | 0 |
| 663h. | Asthma - currently dormant | 0 |
| 663j. | Asthma - currently active | 0 |
| 663m. | Asthma accident and emergency attendance since last visit | 0 |
| 663n. | Asthma treatment compliance satisfactory | 0 |
| 663p. | Asthma treatment compliance unsatisfactory | 0 |
| 663q. | Asthma daytime symptoms | 0 |
| 663r. | Asthma causes night symptoms 1 to 2 times per month | 0 |
| 663s. | Asthma never causes daytime symptoms | 0 |
| 663t. | Asthma causes daytime symptoms 1 to 2 times per month | 0 |
| 663u. | Asthma causes daytime symptoms 1 to 2 times per week | 0 |
| 663v. | Asthma causes daytime symptoms most days | 0 |
| 663w. | Asthma limits walking up hills or stairs | 0 |
| 663x. | Asthma limits walking on the flat | 0 |
| 663y. | Number of asthma exacerbations in past year | 0 |
| 66Y5. | Change in asthma management plan | 0 |
| 66Y9. | Step up change in asthma management plan | 0 |
| 66YA. | Step down change in asthma management plan | 0 |
| 66YC. | Absent from work or school due to asthma | 0 |
| 66YE. | Asthma monitoring due | 0 |
| 66YJ. | Asthma annual review | 0 |
| 66YK. | Asthma follow-up | 0 |
| 66YP. | Asthma night-time symptoms | 0 |
| 66YQ. | Asthma monitoring by nurse | 0 |
| 66YR. | Asthma monitoring by doctor | 0 |
| 66YZ. | Does not have asthma management plan | 0 |
| 66Yp. | Asthma review using Royal College of Physicians three questions | 0 |
| 66Yq. | Asthma causes night time symptoms 1 to 2 times per week | 0 |
| 66Yr. | Asthma causes symptoms most nights | 0 |
| 66Ys. | Asthma never causes night symptoms | 0 |
| 66Yu. | Number of days absent from school due to asthma in past 6 months | 0 |
| 66Yz0 | Asthma management plan declined | 0 |
| 66Yz5 | Telehealth asthma monitoring | 0 |
| 679J. | Health education - asthma | 0 |
| 679J0 | Health education - asthma self management | 0 |
| 679J1 | Health education - structured asthma discussion | 0 |
| 679J2 | Health education - structured patient focused asthma discussion | 0 |
| 68C3. | Asthma screening | 0 |
| 6AP.. | Review of patient at risk of asthma | 0 |
| 8791. | Further asthma - drug prevent. | 0 |
| 8793. | Asthma control step 0 | 0 |
| 8794. | Asthma control step 1 | 0 |
| 8795. | Asthma control step 2 | 0 |
| 8796. | Asthma control step 3 | 0 |
| 8797. | Asthma control step 4 | 0 |
| 8798. | Asthma control step 5 | 0 |
| 8B3j. | Asthma medication review | 0 |
| 8CMA0 | Patient has a written asthma personal action plan | 0 |
| 8CR0. | Asthma clinical management plan | 0 |
| 8H2P. | Emergency admission, asthma | 0 |
| 9N1d. | Seen in asthma clinic | 0 |
| 9N1d0 | Seen in school asthma clinic | 0 |
| 9N4Q. | DNA - Did not attend asthma clinic | 0 |
| 9NI8. | Asthma outreach clinic | 0 |
| 9NNX. | Under care of asthma specialist nurse | 0 |
| 9OJ.. | Asthma monitoring admin. | 0 |
| 9OJ1. | Attends asthma monitoring | 0 |
| 9OJ2. | Refuses asthma monitoring | 0 |
| 9OJ3. | Asthma monitor offer default | 0 |
| 9OJ4. | Asthma monitor 1st letter | 0 |
| 9OJ5. | Asthma monitor 2nd letter | 0 |
| 9OJ6. | Asthma monitor 3rd letter | 0 |
| 9OJ7. | Asthma monitor verbal invite | 0 |
| 9OJ8. | Asthma monitor phone invite | 0 |
| 9OJ9. | Asthma monitoring deleted | 0 |
| 9OJA. | Asthma monitoring check done | 0 |
| 9OJB. | Asthma monitoring invitation SMS (short message service) text message | 0 |
| 9OJB0 | Asthma monitoring SMS (short message service) text message first invitation | 0 |
| 9OJB1 | Asthma monitoring SMS (short message service) text message second invitation | 0 |
| 9OJB2 | Asthma monitoring SMS (short message service) text message third invitation | 0 |
| 9OJC. | Asthma monitoring invitation email | 0 |
| 9OJZ. | Asthma monitoring admin.NOS | 0 |
| 9Oj0. | Hypothyroidism monitoring first letter | 0 |
| 9Q21. | Patient in asthma study | 0 |
| 9hA.. | Exception reporting: asthma quality indicators | 0 |
| 9hA1. | Excepted from asthma quality indicators: Patient unsuitable | 0 |
| 9hA2. | Excepted from asthma quality indicators: Informed dissent | 0 |
| G581. | Left ventricular failure | 0 |
| H3120 | Chronic asthmatic bronchitis | 0 |
| H33.. | Asthma | 0 |
| H330. | Extrinsic (atopic) asthma | 0 |
| H3300 | Extrinsic asthma without status asthmaticus | 0 |
| H3301 | Extrinsic asthma with status asthmaticus | 0 |
| H330z | Extrinsic asthma NOS | 0 |
| H331. | Intrinsic asthma | 0 |
| H3310 | Intrinsic asthma without status asthmaticus | 0 |
| H3311 | Intrinsic asthma with status asthmaticus | 0 |
| H331z | Intrinsic asthma NOS | 0 |
| H332. | Mixed asthma | 0 |
| H333. | Acute exacerbation of asthma | 0 |
| H334. | Brittle asthma | 0 |
| H335. | Chronic asthma with fixed airflow obstruction | 0 |
| H33z. | Asthma unspecified | 0 |
| H33z0 | Status asthmaticus NOS | 0 |
| H33z1 | Asthma attack | 0 |
| H33z2 | Late-onset asthma | 0 |
| H33zz | Asthma NOS | 0 |
| H35y6 | Sequoiosis (red-cedar asthma) | 0 |
| H35y7 | Wood asthma | 0 |
| H3B.. | Asthma-chronic obstructive pulmonary disease overlap syndrome | 0 |
| H47y0 | Detergent asthma | 0 |
| SLF7. | Antiasthmatic poisoning | 0 |
| SLF7z | Antiasthmatic poisoning NOS | 0 |
| TJF7. | Adverse reaction to antiasthmatics | 0 |
| TJF73 | Adverse reaction to theophylline (asthma) | 0 |
| TJF7z | Adverse reaction to antiasthmatic NOS | 0 |
| U60F6 | [X]Antiasthmatics causing adverse effects in therapeutic use, not elsewhere classified | 0 |

---

#### CONCEPT: Atopic Eczema¶

| READ\_CODE | DESCRIPTION\_SAIL | SENSITIVE\_CODE |
| --- | --- | --- |
| 14F1. | H/O: eczema | 0 |
| M07y. | Other specified local infection of skin or subcutaneous tissue | 0 |
| M102. | Infectious eczematoid dermatitis | 0 |
| M11.. | Atopic dermatitis and related conditions | 0 |
| M111. | Atopic dermatitis/eczema | 0 |
| M113. | Flexural eczema | 0 |
| M114. | Allergic (intrinsic) eczema | 0 |
| M115. | Besnier's prurigo | 0 |
| M117. | Neurodermatitis - atopic | 0 |
| M11z. | Atopic dermatitis NOS | 0 |
| M12z1 | Eczema NOS | 0 |
| M12z2 | Infected eczema | 0 |
| M12z4 | Erythrodermic eczema | 0 |
| Myu2. | [X]Dermatitis and eczema | 0 |
| Myu22 | [X]Exacerbation of eczema | 0 |

---

#### CONCEPT: Autism and ADHD¶

| READ\_CODE | DESCRIPTION\_SAIL | SENSITIVE\_CODE |
| --- | --- | --- |
| 6A61. | Attention deficit hyperactivity disorder annual review | 0 |
| 8BPT. | Drug therapy for ADHD (attention deficit hyperactivity disorder) | 0 |
| 8BPT0 | Stimulant drug therapy for ADHD (attention deficit hyperactivity disorder) | 0 |
| 8BPT1 | Non-stimulant drug therapy for ADHD (attention deficit hyperactivity disorder) | 0 |
| 9Ngp. | On drug therapy for ADHD (attention deficit hyperactivity disorder) | 0 |
| 9Ngp0 | On stimulant drug therapy for ADHD (attention deficit hyperactivity disorder) | 0 |
| 9Ngp1 | On non-stimulant drug therapy for ADHD (attention deficit hyperactivity disorder) | 0 |
| 9Ol8. | Attention deficit hyperactivity disorder monitoring invitation first letter | 0 |
| 9Ol9. | Attention deficit hyperactivity disorder monitoring invitation second letter | 0 |
| 9OlA. | Attention deficit hyperactivity disorder monitoring invitation third letter | 0 |
| E140. | Infantile autism | 0 |
| E1400 | Active infantile autism | 0 |
| E1401 | Residual infantile autism | 0 |
| E140z | Infantile autism NOS | 0 |
| E2E.. | Childhood hyperkinetic syndrome | 0 |
| E2E0. | Child attention deficit disorder | 0 |
| E2E01 | Attention deficit with hyperactivity | 0 |
| E2E0z | Child attention deficit disorder NOS | 0 |
| E2E1. | Hyperkinesis with developmental delay | 0 |
| Eu840 | [X]Childhood autism | 0 |
| Eu841 | [X]Atypical autism | 0 |
| Eu845 | [X]Asperger's syndrome | 0 |
| Eu84z | [X]Pervasive developmental disorder, unspecified | 0 |
| Eu900 | [X]Disturbance of activity and attention | 0 |
| Eu902 | [X]Deficits in attention, motor control and perception | 0 |
| Eu9y7 | [X]Attention deficit disorder | 0 |

---

#### CONCEPT: Bipolar Disorder¶

| READ\_CODE | DESCRIPTION\_SAIL | SENSITIVE\_CODE |
| --- | --- | --- |
| 146D. | H/O: manic depressive disorder | 0 |
| 212T. | Psychosis, schizophrenia and bipolar affective disorder resolved | 0 |
| 212V. | Bipolar affective disorder resolved | 0 |
| E11.. | Affective psychoses | 0 |
| E110. | Manic disorder, single episode | 0 |
| E1100 | Single manic episode, unspecified | 0 |
| E1101 | Single manic episode, mild | 0 |
| E1102 | Single manic episode, moderate | 0 |
| E1103 | Single manic episode, severe without mention of psychosis | 0 |
| E1104 | Single manic episode, severe, with psychosis | 0 |
| E1105 | Single manic episode in partial or unspecified remission | 0 |
| E1106 | Single manic episode in full remission | 0 |
| E110z | Manic disorder, single episode NOS | 0 |
| E111. | Recurrent manic episodes | 0 |
| E1110 | Recurrent manic episodes, unspecified | 0 |
| E1111 | Recurrent manic episodes, mild | 0 |
| E1112 | Recurrent manic episodes, moderate | 0 |
| E1113 | Recurrent manic episodes, severe without mention of psychosis | 0 |
| E1114 | Recurrent manic episodes, severe, with psychosis | 0 |
| E1115 | Recurrent manic episodes, in partial or unspecified remission | 0 |
| E1116 | Recurrent manic episodes, in full remission | 0 |
| E111z | Recurrent manic episode NOS | 0 |
| E114. | Bipolar affective disorder, currently manic | 0 |
| E1140 | Bipolar affective disorder, currently manic, unspecified | 0 |
| E1141 | Bipolar affective disorder, currently manic, mild | 0 |
| E1142 | Bipolar affective disorder, currently manic, moderate | 0 |
| E1143 | Bipolar affective disorder, currently manic, severe, without mention of psychosis | 0 |
| E1144 | Bipolar affective disorder, currently manic, severe, with psychosis | 0 |
| E1145 | Bipolar affective disorder, currently manic, in partial or unspecified remission | 0 |
| E1146 | Bipolar affective disorder, currently manic, in full remission | 0 |
| E114z | Bipolar affective disorder, currently manic, NOS | 0 |
| E115. | Bipolar affective disorder, currently depressed | 0 |
| E1150 | Bipolar affective disorder, currently depressed, unspecified | 0 |
| E1151 | Bipolar affective disorder, currently depressed, mild | 0 |
| E1152 | Bipolar affective disorder, currently depressed, moderate | 0 |
| E1153 | Bipolar affective disorder, currently depressed, severe, without mention of psychosis | 0 |
| E1154 | Bipolar affective disorder, currently depressed, severe, with psychosis | 0 |
| E1155 | Bipolar affective disorder, currently depressed, in partial or unspecified remission | 0 |
| E1156 | Bipolar affective disorder, currently depressed, in full remission | 0 |
| E115z | Bipolar affective disorder, currently depressed, NOS | 0 |
| E116. | Mixed bipolar affective disorder | 0 |
| E1160 | Mixed bipolar affective disorder, unspecified | 0 |
| E1161 | Mixed bipolar affective disorder, mild | 0 |
| E1162 | Mixed bipolar affective disorder, moderate | 0 |
| E1163 | Mixed bipolar affective disorder, severe, without mention of psychosis | 0 |
| E1164 | Mixed bipolar affective disorder, severe, with psychosis | 0 |
| E1165 | Mixed bipolar affective disorder, in partial or unspecified remission | 0 |
| E1166 | Mixed bipolar affective disorder, in full remission | 0 |
| E116z | Mixed bipolar affective disorder, NOS | 0 |
| E117. | Unspecified bipolar affective disorder | 0 |
| E1170 | Unspecified bipolar affective disorder, unspecified | 0 |
| E1171 | Unspecified bipolar affective disorder, mild | 0 |
| E1172 | Unspecified bipolar affective disorder, moderate | 0 |
| E1173 | Unspecified bipolar affective disorder, severe, without mention of psychosis | 0 |
| E1174 | Unspecified bipolar affective disorder, severe, with psychosis | 0 |
| E1175 | Unspecified bipolar affective disorder, in partial or unspecified remission | 0 |
| E1176 | Unspecified bipolar affective disorder, in full remission | 0 |
| E117z | Unspecified bipolar affective disorder, NOS | 0 |
| E11y. | Other and unspecified manic-depressive psychoses | 0 |
| E11y0 | Unspecified manic-depressive psychoses | 0 |
| E11y3 | Other mixed manic-depressive psychoses | 0 |
| E11yz | Other and unspecified manic-depressive psychoses NOS | 0 |
| Eu30. | [X]Manic episode | 0 |
| Eu301 | [X]Mania without psychotic symptoms | 0 |
| Eu302 | [X]Mania with psychotic symptoms | 0 |
| Eu30y | [X]Other manic episodes | 0 |
| Eu30z | [X]Manic episode, unspecified | 0 |
| Eu31. | [X]Bipolar affective disorder | 0 |
| Eu310 | [X]Bipolar affective disorder, current episode hypomanic | 0 |
| Eu311 | [X]Bipolar affective disorder, current episode manic without psychotic symptoms | 0 |
| Eu312 | [X]Bipolar affective disorder, current episode manic with psychotic symptoms | 0 |
| Eu313 | [X]Bipolar affective disorder, current episode mild or moderate depression | 0 |
| Eu314 | [X]Bipolar affective disorder, current episode severe depression without psychotic symptoms | 0 |
| Eu315 | [X]Bipolar affective disorder, current episode severe depression with psychotic symptoms | 0 |
| Eu316 | [X]Bipolar affective disorder, current episode mixed | 0 |
| Eu317 | [X]Bipolar affective disorder, currently in remission | 0 |
| Eu318 | [X]Bipolar affective disorder type I | 0 |
| Eu319 | [X]Bipolar affective disorder type II | 0 |
| Eu31y | [X]Other bipolar affective disorders | 0 |
| Eu31z | [X]Bipolar affective disorder, unspecified | 0 |
| Eu332 | [X]Recurrent depressive disorder, current episode severe without psychotic symptoms | 0 |
| Eu333 | [X]Recurrent depressive disorder, current episode severe with psychotic symptoms | 0 |
| Eu340 | [X]Cyclothymia | 0 |
| ZV111 | [V]Personal history of affective disorder | 0 |

---

#### CONCEPT: Blindness and low vision¶

| READ\_CODE | DESCRIPTION\_SAIL | SENSITIVE\_CODE |
| --- | --- | --- |
| 13o8J | Uses deafblind intervener | 0 |
| 13o8N | Uses Deafblind Manual Alphabet | 0 |
| 1B75. | Loss of vision | 0 |
| 1B77. | Deteriorating vision | 0 |
| 1a000 | Uses guide dog for the blind | 0 |
| 2B6B. | O/E - R-eye completely blind | 0 |
| 2B6S. | O/E - pinhole R-eye completely blind | 0 |
| 2B7B. | O/E - L-eye completely blind | 0 |
| 2B7S. | O/E - pinhole L-eye completely blind | 0 |
| 2BBr. | Impaired vision due to diabetic retinopathy | 0 |
| 6688. | Registered partially sighted | 0 |
| 6689. | Registered blind | 0 |
| 668B. | Poor visual acuity | 0 |
| 668C. | Certificate of vision impairment | 0 |
| 668D. | Registered sight impaired | 0 |
| 8F6.. | Specific disability rehab. | 0 |
| 8F61. | Blind rehabilitation | 0 |
| 8F62. | Blind lead dog rehabilitation | 0 |
| 9NfB. | Requires deafblind communicator guide | 0 |
| 9Nfa. | Requires deafblind manual alphabet interpreter | 0 |
| 9Nfb. | Requires deafblind block alphabet interpreter | 0 |
| 9Nfc. | Requires deafblind haptic communication interpreter | 0 |
| 9m08. | Excluded from diabetic retinopathy screening as blind | 0 |
| C245. | Vitamin A deficiency with night blindness | 0 |
| E2011 | Hysterical blindness | 0 |
| F4041 | Blind hypotensive eye | 0 |
| F4042 | Blind hypertensive eye | 0 |
| F4653 | After-cataract with vision obscured | 0 |
| F4811 | Sudden visual loss | 0 |
| F4814 | Other transient visual loss | 0 |
| F486. | Night blindness | 0 |
| F4860 | Unspecified night blindness | 0 |
| F4862 | Hereditary night blindness NOS | 0 |
| F4863 | Congenital night blindness NOS | 0 |
| F4864 | Acquired night blindness | 0 |
| F486z | Night blindness NOS | 0 |
| F49.. | Blindness and low vision | 0 |
| F490. | Blindness, both eyes | 0 |
| F4900 | Unspecified blindness both eyes | 0 |
| F4901 | Both eyes total visual impairment | 0 |
| F4902 | Better eye: near total visual impairment, Lesser eye: unspecified | 0 |
| F4903 | Better eye: near total visual impairment, Lesser eye: total visual impairment | 0 |
| F4904 | Better eye: near total visual impairment, Lesser eye: near total visual impairment | 0 |
| F4905 | Better eye: profound visual impairment, Lesser eye: unspecified | 0 |
| F4906 | Better eye: profound visual impairment, Lesser eye: total visual impairment | 0 |
| F4907 | Better eye: profound visual impairment, Lesser eye: near total visual impairment | 0 |
| F4908 | Better eye: profound visual impairment, Lesser eye: profound visual impairment | 0 |
| F4909 | Acquired blindness, both eyes | 0 |
| F490z | Blindness both eyes NOS | 0 |
| F491. | Better eye: low vision, Lesser eye: profound visual impairment | 0 |
| F4910 | One eye blind, one eye low vision | 0 |
| F4911 | Better eye: severe visual impairment, Lesser eye: blind, unspecified | 0 |
| F4912 | Better eye: severe visual impairment, Lesser eye: total visual impairment | 0 |
| F4913 | Better eye: severe visual impairment, Lesser eye: near total visual impairment | 0 |
| F4914 | Better eye: severe visual impairment, Lesser eye: profound visual impairment | 0 |
| F4915 | Better eye: moderate visual impairment, Lesser eye: blind, unspecified | 0 |
| F4916 | Better eye: moderate visual impairment, Lesser eye: total visual impairment | 0 |
| F4917 | Better eye: moderate visual impairment, Lesser eye: near total visual impairment | 0 |
| F4918 | Better eye: moderate visual impairment, Lesser eye: profound visual impairment | 0 |
| F491z | One eye blind, one eye low vision NOS | 0 |
| F492. | Low vision, both eyes | 0 |
| F4920 | Low vision, both eyes unspecified | 0 |
| F4921 | Better eye: severe visual impairment, Lesser eye: low vision unspecified | 0 |
| F4922 | Better eye: severe visual impairment, Lesser eye: severe visual impairment | 0 |
| F4923 | Better eye: moderate visual impairment, Lesser eye: low vision unspecified | 0 |
| F4924 | Better eye: moderate visual impairment, Lesser eye: severe visual impairment | 0 |
| F4925 | Better eye: moderate visual impairment, Lesser eye: moderate visual impairment | 0 |
| F492z | Low vision, both eyes NOS | 0 |
| F493. | Visual loss, both eyes unqualified | 0 |
| F494. | Legal blindness USA | 0 |
| F495. | Profound impairment, one eye | 0 |
| F4950 | Blindness, one eye, unspecified | 0 |
| F4951 | Lesser eye: total visual impairment, Better eye: unspecified | 0 |
| F4952 | Lesser eye: total visual impairment, Better eye: near normal vision | 0 |
| F4953 | Lesser eye: total visual impairment, Better eye: normal vision | 0 |
| F4954 | Lesser eye: near total visual impairment, Better eye: unspecified | 0 |
| F4955 | Lesser eye: near total visual impairment, Better eye: near normal vision | 0 |
| F4956 | Lesser eye: near total visual impairment, Better eye: normal vision | 0 |
| F4957 | Lesser eye: profound visual impairment, Better eye: unspecified | 0 |
| F4958 | Lesser eye: profound visual impairment, Better eye: near normal vision | 0 |
| F4959 | Lesser eye: profound visual impairment, Better eye: normal vision | 0 |
| F495A | Acquired blindness, one eye | 0 |
| F495z | Profound impairment one eye NOS | 0 |
| F496. | Low vision, one eye | 0 |
| F4960 | Low vision, one eye, unspecified | 0 |
| F4961 | Lesser eye: severe visual impairment, Better eye: unspecified | 0 |
| F4962 | Lesser eye: severe visual impairment, Better eye: near normal vision | 0 |
| F4963 | Lesser eye: severe visual impairment, Better eye: normal vision | 0 |
| F4964 | Lesser eye: moderate visual impairment, Better eye: unspecified | 0 |
| F4965 | Lesser eye: moderate visual impairment, Better eye: near normal vision | 0 |
| F4966 | Lesser eye: moderate visual impairment, Better eye: normal vision | 0 |
| F496z | Low vision, one eye NOS | 0 |
| F497. | Severe visual impairment, binocular | 0 |
| F49D. | Visual impairment | 0 |
| F49y. | Visual loss, one eye, unqualified | 0 |
| F49z. | Visual loss NOS | 0 |
| F4H73 | Cortical blindness | 0 |
| Fy1.. | Combined visual and hearing impairment | 0 |
| FyuL. | [X]Visual disturbances and blindness | 0 |
| P3410 | Congenital corneal opacity with visual deficit | 0 |
| P3411 | Congenital corneal opacity without visual deficit | 0 |
| SJ0z. | Optic nerve or pathway injury NOS | 0 |

---

#### CONCEPT: Breast Cancer¶

| READ\_CODE | DESCRIPTION\_SAIL | SENSITIVE\_CODE |
| --- | --- | --- |
| B34.. | Malignant neoplasm of female breast | 0 |
| B340. | Malignant neoplasm of nipple and areola of female breast | 0 |
| B3400 | Malignant neoplasm of nipple of female breast | 0 |
| B3401 | Malignant neoplasm of areola of female breast | 0 |
| B340z | Malignant neoplasm of nipple or areola of female breast NOS | 0 |
| B341. | Malignant neoplasm of central part of female breast | 0 |
| B342. | Malignant neoplasm of upper-inner quadrant of female breast | 0 |
| B343. | Malignant neoplasm of lower-inner quadrant of female breast | 0 |
| B344. | Malignant neoplasm of upper-outer quadrant of female breast | 0 |
| B345. | Malignant neoplasm of lower-outer quadrant of female breast | 0 |
| B346. | Malignant neoplasm of axillary tail of female breast | 0 |
| B347. | Malignant neoplasm, overlapping lesion of breast | 0 |
| B34y. | Malignant neoplasm of other site of female breast | 0 |
| B34yz | Malignant neoplasm of other site of female breast NOS | 0 |
| B34z. | Malignant neoplasm of female breast NOS | 0 |
| B35.. | Malignant neoplasm of male breast | 0 |
| B350. | Malignant neoplasm of nipple and areola of male breast | 0 |
| B3500 | Malignant neoplasm of nipple of male breast | 0 |
| B3501 | Malignant neoplasm of areola of male breast | 0 |
| B350z | Malignant neoplasm of nipple or areola of male breast NOS | 0 |
| B35z. | Malignant neoplasm of other site of male breast | 0 |
| B35zz | Malignant neoplasm of male breast NOS | 0 |
| B36.. | Local recurrence of malignant tumour of breast | 0 |
| B77.. | Benign neoplasm of breast | 0 |
| B770. | Benign neoplasm of female breast | 0 |
| B771. | Benign neoplasm of male breast | 0 |
| B77z. | Benign neoplasm of breast NOS | 0 |
| B83.. | Carcinoma in situ of breast and genitourinary system | 0 |
| B830. | Carcinoma in situ of breast | 0 |
| B8300 | Lobular carcinoma in situ of breast | 0 |
| B8301 | Intraductal carcinoma in situ of breast | 0 |
| BB94. | [M]Juvenile breast carcinoma | 0 |
| BB9J. | [M]Paget's disease, mammary | 0 |
| BB9K. | [M]Paget's disease and infiltrating breast duct carcinoma | 0 |
| BB9K0 | [M]Paget's disease and intraductal carcinoma of breast | 0 |
| Byu6. | [X]Malignant neoplasm of breast | 0 |
| ByuFG | [X]Other carcinoma in situ of breast | 0 |
| ZV103 | [V]Personal history of malignant neoplasm of breast | 0 |
| ZV13A | [V]Personal history of non-neoplastic breast disease | 0 |

---

#### CONCEPT: Bronchiectasis¶

| READ\_CODE | DESCRIPTION\_SAIL | SENSITIVE\_CODE |
| --- | --- | --- |
| 14BA. | H/O: bronchiectasis | 0 |
| A115. | Tuberculous bronchiectasis | 0 |
| H34.. | Bronchiectasis | 0 |
| H340. | Recurrent bronchiectasis | 0 |
| H341. | Post-infective bronchiectasis | 0 |
| H34z. | Bronchiectasis NOS | 0 |
| P861. | Congenital bronchiectasis | 0 |

---

#### CONCEPT: Chronic Back Pain¶

| READ\_CODE | DESCRIPTION\_SAIL | SENSITIVE\_CODE |
| --- | --- | --- |
| 16C9. | Chronic low back pain | 0 |
| 8HTH. | Referral to back pain clinic | 0 |

---

#### CONCEPT: Chronic Fatigue Syndrome¶

| READ\_CODE | DESCRIPTION\_SAIL | SENSITIVE\_CODE |
| --- | --- | --- |
| F03y. | Other causes of encephalitis | 0 |
| F286. | Chronic fatigue syndrome | 0 |
| F2860 | Mild chronic fatigue syndrome | 0 |
| F2861 | Moderate chronic fatigue syndrome | 0 |
| F2862 | Severe chronic fatigue syndrome | 0 |
| G8522 | Oesophageal varices in cirrhosis of the liver | 0 |
| J601. | Subacute necrosis of liver | 0 |
| J6010 | Subacute hepatic failure | 0 |
| J6011 | Subacute hepatitis - noninfective | 0 |
| J6012 | Subacute yellow atrophy | 0 |
| J601z | Subacute necrosis of liver NOS | 0 |
| J60z. | Acute and subacute liver necrosis NOS | 0 |
| J61.. | Cirrhosis and chronic liver disease | 0 |
| J614. | Chronic hepatitis | 0 |
| J6140 | Chronic persistent hepatitis | 0 |
| J6141 | Chronic active hepatitis | 0 |
| J6142 | Chronic aggressive hepatitis | 0 |
| J6143 | Recurrent hepatitis | 0 |
| J6144 | Chronic lobular hepatitis | 0 |
| J614y | Chronic hepatitis unspecified | 0 |
| J614z | Chronic hepatitis NOS | 0 |
| J615. | Cirrhosis - non alcoholic | 0 |
| J6150 | Unilobular portal cirrhosis | 0 |
| J6151 | Multilobular portal cirrhosis | 0 |
| J6152 | Mixed portal cirrhosis | 0 |
| J6153 | Diffuse nodular cirrhosis | 0 |
| J6154 | Fatty portal cirrhosis | 0 |
| J6155 | Hypertrophic portal cirrhosis | 0 |
| J6156 | Capsular portal cirrhosis | 0 |
| J6157 | Cardiac portal cirrhosis | 0 |
| J6158 | Juvenile portal cirrhosis | 0 |
| J6159 | Pigmentary portal cirrhosis | 0 |
| J615A | Pipe-stem portal cirrhosis | 0 |
| J615B | Toxic portal cirrhosis | 0 |
| J615C | Xanthomatous portal cirrhosis | 0 |
| J615D | Bacterial portal cirrhosis | 0 |
| J615E | Cardituberculous cirrhosis | 0 |
| J615F | Syphilitic portal cirrhosis | 1 |
| J615G | Zooparasitic portal cirrhosis | 0 |
| J615H | Infectious cirrhosis NOS | 0 |
| J615y | Portal cirrhosis unspecified | 0 |
| J615z | Non-alcoholic cirrhosis NOS | 0 |
| J616. | Biliary cirrhosis | 0 |
| J6160 | Primary biliary cirrhosis | 0 |
| J6161 | Secondary biliary cirrhosis | 0 |
| J6162 | Biliary cirrhosis of children | 0 |
| J616z | Biliary cirrhosis NOS | 0 |
| J617. | Alcoholic hepatitis | 0 |
| J6170 | Chronic alcoholic hepatitis | 0 |
| J61y. | Other non-alcoholic chronic liver disease | 0 |
| J61y0 | Chronic yellow liver atrophy | 0 |
| J61y1 | Non-alcoholic fatty liver | 0 |
| J61y2 | Hepatosplenomegaly | 0 |
| J61y3 | Portal fibrosis without cirrhosis | 0 |
| J61y4 | Hepatic fibrosis | 0 |
| J61y5 | Hepatic sclerosis | 0 |
| J61y6 | Hepatic fibrosis with hepatic sclerosis | 0 |
| J61y7 | Steatosis of liver | 0 |
| J61y8 | Nonalcoholic steatohepatitis | 0 |
| J61yz | Other non-alcoholic chronic liver disease NOS | 0 |
| J61z. | Chronic liver disease NOS | 0 |
| J623. | Portal hypertension | 0 |
| J624. | Hepatorenal syndrome | 0 |
| J62y. | Other sequelae of chronic liver disease | 0 |
| J62z. | Liver abscess and chronic liver disease causing sequelae NOS | 0 |
| J630. | Chronic passive liver congestion | 0 |
| Jyu71 | [X]Other and unspecified cirrhosis of liver | 0 |
| PB62. | Congenital cystic liver disease | 0 |
| PB620 | Congenital polycystic liver disease | 0 |

---

#### CONCEPT: Chronic Liver Disease¶

| READ\_CODE | DESCRIPTION\_SAIL | SENSITIVE\_CODE |
| --- | --- | --- |
| 141E. | History of hepatitis B | 1 |
| 14i.. | H/O hepatitis C antiviral drug therapy | 1 |
| 21267 | Hepatitis C resolved | 1 |
| 2J12. | Hepatitis C non immune | 0 |
| 43B4. | Hepatitis B surface antig +ve | 1 |
| 43B6. | Hepatitis B non immune | 0 |
| 43B7. | Hepatitis C non-immune | 0 |
| 43X3. | Hepatitis C antibody test positive | 1 |
| 4JQ3. | Hepatitis C virus genotype | 1 |
| 4JQD. | Hepatitis C viral ribonucleic acid polymerase chain reaction positive | 1 |
| 4JQF. | Hepatitis C antigen positive | 1 |
| 7609. | Open operations on oesophageal varices | 0 |
| 76093 | Local ligation of oesophageal varices | 0 |
| 76094 | Open injection sclerotherapy to oesophageal varices | 0 |
| 7609y | Other specified open operation on oesophageal varices | 0 |
| 7609z | Open operation on oesophageal varices NOS | 0 |
| 760C5 | Fibreoptic endoscopic banding of oesophageal varices | 0 |
| 760F3 | Endoscopic injection sclerotherapy to varices of oesophagus using rigid oesophagoscope | 0 |
| 760F4 | Rigid oesophagoscopic banding of oesophageal varices | 0 |
| 7L1f. | Compensation for liver failure | 0 |
| 7L1fz | Compensation for liver failure NOS | 0 |
| 7Q052 | Hepatitis B treatment drugs Band 1 | 1 |
| 7Q053 | Respiratory syncytial virus treatment and Hepatitis C treatment drugs Band 1 | 0 |
| 8BB5. | 12 week virologic response to hepatitis C treatment | 1 |
| 9NgR. | On hepatitis C treatment plan | 1 |
| 9kR.. | Chronic hepatitis annual review - enhanced services administration | 1 |
| 9kV.. | Hepatitis C screening positive - enhanced services administration | 0 |
| 9kZ.. | Hepatitis B screening positive - enhanced services administration | 1 |
| A702. | Viral hepatitis B with coma | 0 |
| A703. | Viral (serum) hepatitis B | 1 |
| A7040 | Viral hepatitis C with coma | 0 |
| A7050 | Viral hepatitis C without mention of hepatic coma | 1 |
| A7051 | Acute delta-(super)infection of hepatitis B carrier | 1 |
| A707. | Chronic viral hepatitis | 1 |
| A7070 | Chronic viral hepatitis B with delta-agent | 1 |
| A7071 | Chronic viral hepatitis B without delta-agent | 1 |
| A7072 | Chronic viral hepatitis C | 1 |
| A7073 | Chronic viral hepatitis B | 1 |
| A707X | Chronic viral hepatitis, unspecified | 1 |
| A70A. | Hepatitis C genotype 1 | 1 |
| A70B. | Hepatitis C genotype 2 | 1 |
| A70C. | Hepatitis C genotype 3 | 1 |
| A70D. | Hepatitis C genotype 4 | 1 |
| A70E. | Hepatitis C genotype 5 | 0 |
| A70F. | Hepatitis C genotype 6 | 0 |
| A70G. | Acute hepatitis C | 0 |
| A70z0 | Hepatitis C | 1 |
| AyuB1 | [X]Other chronic viral hepatitis | 1 |
| AyuB2 | [X]Chronic viral hepatitis, unspecified | 1 |
| C3104 | Glycogenosis with hepatic cirrhosis | 0 |
| C3500 | Haemochromatosis | 0 |
| C3707 | Liver disease due to cystic fibrosis | 0 |
| G85.. | Other varicose veins | 0 |
| G850. | Oesophageal varices with bleeding | 0 |
| G851. | Oesophageal varices without bleeding | 0 |
| G852. | Oesophageal varices in diseases EC | 0 |
| G8520 | Oesophageal varices with bleeding in diseases EC | 0 |
| G8521 | Oesophageal varices without bleeding in diseases EC | 0 |
| G8522 | Oesophageal varices in cirrhosis of the liver | 0 |
| G8523 | Oesophageal varices in alcoholic cirrhosis of the liver | 0 |
| G852z | Oesophageal varices in diseases EC NOS | 0 |
| G858. | Oesophageal varices NOS | 0 |
| Gyu94 | [X]Oesophageal varices in diseases classified elsewhere | 0 |
| J61.. | Cirrhosis and chronic liver disease | 0 |
| J610. | Alcoholic fatty liver | 0 |
| J611. | Acute alcoholic hepatitis | 0 |
| J612. | Alcoholic cirrhosis of liver | 0 |
| J6120 | Alcoholic fibrosis and sclerosis of liver | 0 |
| J613. | Alcoholic liver damage unspecified | 0 |
| J6130 | Alcoholic hepatic failure | 0 |
| J614. | Chronic hepatitis | 0 |
| J6140 | Chronic persistent hepatitis | 0 |
| J6141 | Chronic active hepatitis | 0 |
| J6142 | Chronic aggressive hepatitis | 0 |
| J6143 | Recurrent hepatitis | 0 |
| J6144 | Chronic lobular hepatitis | 0 |
| J614y | Chronic hepatitis unspecified | 0 |
| J614z | Chronic hepatitis NOS | 0 |
| J615. | Cirrhosis - non alcoholic | 0 |
| J6150 | Unilobular portal cirrhosis | 0 |
| J6151 | Multilobular portal cirrhosis | 0 |
| J6152 | Mixed portal cirrhosis | 0 |
| J6153 | Diffuse nodular cirrhosis | 0 |
| J6154 | Fatty portal cirrhosis | 0 |
| J6155 | Hypertrophic portal cirrhosis | 0 |
| J6156 | Capsular portal cirrhosis | 0 |
| J6157 | Cardiac portal cirrhosis | 0 |
| J6158 | Juvenile portal cirrhosis | 0 |
| J6159 | Pigmentary portal cirrhosis | 0 |
| J615A | Pipe-stem portal cirrhosis | 0 |
| J615B | Toxic portal cirrhosis | 0 |
| J615C | Xanthomatous portal cirrhosis | 0 |
| J615D | Bacterial portal cirrhosis | 0 |
| J615E | Cardituberculous cirrhosis | 0 |
| J615F | Syphilitic portal cirrhosis | 1 |
| J615G | Zooparasitic portal cirrhosis | 0 |
| J615H | Infectious cirrhosis NOS | 0 |
| J615y | Portal cirrhosis unspecified | 0 |
| J615z | Non-alcoholic cirrhosis NOS | 0 |
| J616. | Biliary cirrhosis | 0 |
| J6160 | Primary biliary cirrhosis | 0 |
| J6161 | Secondary biliary cirrhosis | 0 |
| J6162 | Biliary cirrhosis of children | 0 |
| J616z | Biliary cirrhosis NOS | 0 |
| J617. | Alcoholic hepatitis | 0 |
| J6170 | Chronic alcoholic hepatitis | 0 |
| J61y. | Other non-alcoholic chronic liver disease | 0 |
| J61y0 | Chronic yellow liver atrophy | 0 |
| J61y1 | Non-alcoholic fatty liver | 0 |
| J61y3 | Portal fibrosis without cirrhosis | 0 |
| J61y4 | Hepatic fibrosis | 0 |
| J61y5 | Hepatic sclerosis | 0 |
| J61y6 | Hepatic fibrosis with hepatic sclerosis | 0 |
| J61y8 | Nonalcoholic steatohepatitis | 0 |
| J61yz | Other non-alcoholic chronic liver disease NOS | 0 |
| J61z. | Chronic liver disease NOS | 0 |
| J62.. | Liver abscess and sequelae of chronic liver disease | 0 |
| J623. | Portal hypertension | 0 |
| J624. | Hepatorenal syndrome | 0 |
| J625. | [X] Hepatic failure | 0 |
| J62y. | Other sequelae of chronic liver disease | 0 |
| J62z. | Liver abscess and chronic liver disease causing sequelae NOS | 0 |
| J6353 | Toxic liver disease with chronic persistent hepatitis | 0 |
| J6354 | Toxic liver disease with chronic lobular hepatitis | 0 |
| J6355 | Toxic liver disease with chronic active hepatitis | 0 |
| J6356 | Toxic liver disease with fibrosis and cirrhosis of liver | 0 |
| J63B. | Autoimmune hepatitis | 0 |
| J6617 | Primary sclerosing cholangitis | 0 |
| Jyu71 | [X]Other and unspecified cirrhosis of liver | 0 |
| PB621 | Fibrocystic liver disease | 0 |
| Q4091 | Congenital hepatitis B infection | 1 |
| Q48yz | Other perinatal condition NOS | 0 |
| SP142 | Hepatic failure as a complication of care | 0 |
| SP143 | Hepatorenal syndrome as a complication of care | 0 |
| ZV02B | [V]Hepatitis B carrier | 1 |
| ZV02C | [V]Hepatitis C carrier | 1 |

---

#### CONCEPT: Chronic Pain¶

| READ\_CODE | DESCRIPTION\_SAIL | SENSITIVE\_CODE |
| --- | --- | --- |
| 16C9. | Chronic low back pain | 0 |
| 1M52. | Chronic pain | 0 |
| 8HTH. | Referral to back pain clinic | 0 |
| F347. | Complex regional pain syndrome type II | 0 |
| F369. | Complex regional pain syndrome | 0 |
| N337. | Algoneurodystrophy | 0 |
| N3370 | Disuse atrophy of bone | 0 |
| N3371 | Sudek's atrophy | 0 |
| N3372 | Algodystrophy of hand | 0 |
| N3373 | Algodystrophy of knee | 0 |
| N3374 | Algodystrophy of foot | 0 |
| N337z | Algoneurodystrophy NOS | 0 |
| N33C. | Complex regional pain syndrome type I | 0 |
| R00zC | [D]Chronic intractable pain | 0 |
| Ryu70 | [X]Other chronic pain | 0 |

---

#### CONCEPT: Chronic Sinusitis¶

| READ\_CODE | DESCRIPTION\_SAIL | SENSITIVE\_CODE |
| --- | --- | --- |
| H13.. | Chronic sinusitis | 0 |
| H130. | Chronic maxillary sinusitis | 0 |
| H131. | Chronic frontal sinusitis | 0 |
| H132. | Chronic ethmoidal sinusitis | 0 |
| H133. | Chronic sphenoidal sinusitis | 0 |
| H134. | Fistula of nasal sinus | 0 |
| H135. | Recurrent sinusitis | 0 |
| H13y. | Other chronic sinusitis | 0 |
| H13y0 | Chronic pansinusitis | 0 |
| H13y1 | Pansinusitis | 0 |
| H13yz | Other chronic sinusitis NOS | 0 |
| H13z. | Chronic sinusitis NOS | 0 |
| Hyu22 | [X]Other chronic sinusitis | 0 |
| J0835 | Oroantral fistula | 0 |

---

#### CONCEPT: Chronic Kidney Disease (CKD) Stage 3\_5¶

| READ\_CODE | DESCRIPTION\_SAIL | SENSITIVE\_CODE |
| --- | --- | --- |
| 1Z12. | Chronic kidney disease stage 3 | 0 |
| 1Z13. | Chronic kidney disease stage 4 | 0 |
| 1Z14. | Chronic kidney disease stage 5 | 0 |
| 1Z15. | Chronic kidney disease stage 3A | 0 |
| 1Z16. | Chronic kidney disease stage 3B | 0 |
| 1Z1B. | Chronic kidney disease stage 3 with proteinuria | 0 |
| 1Z1C. | Chronic kidney disease stage 3 without proteinuria | 0 |
| 1Z1D. | Chronic kidney disease stage 3A with proteinuria | 0 |
| 1Z1E. | Chronic kidney disease stage 3A without proteinuria | 0 |
| 1Z1F. | Chronic kidney disease stage 3B with proteinuria | 0 |
| 1Z1G. | Chronic kidney disease stage 3B without proteinuria | 0 |
| 1Z1H. | Chronic kidney disease stage 4 with proteinuria | 0 |
| 1Z1J. | Chronic kidney disease stage 4 without proteinuria | 0 |
| 1Z1K. | Chronic kidney disease stage 5 with proteinuria | 0 |
| 1Z1L. | Chronic kidney disease stage 5 without proteinuria | 0 |
| 1Z1T. | CKD G3aA1 - chronic kidney disease with glomerular filtration rate category G3a and albuminuria category A1 | 0 |
| 1Z1V. | CKD G3aA2 - chronic kidney disease with glomerular filtration rate category G3a and albuminuria category A2 | 0 |
| 1Z1W. | CKD G3aA3 - chronic kidney disease with glomerular filtration rate category G3a and albuminuria category A3 | 0 |
| 1Z1X. | CKD G3bA1 - chronic kidney disease with glomerular filtration rate category G3b and albuminuria category A1 | 0 |
| 1Z1Y. | CKD G3bA2 - chronic kidney disease with glomerular filtration rate category G3b and albuminuria category A2 | 0 |
| 1Z1Z. | CKD G3bA3 - chronic kidney disease with glomerular filtration rate category G3b and albuminuria category A3 | 0 |
| 1Z1a. | CKD G4A1 - chronic kidney disease with glomerular filtration rate category G4 and albuminuria category A1 | 0 |
| 1Z1b. | CKD G4A2 - chronic kidney disease with glomerular filtration rate category G4 and albuminuria category A2 | 0 |
| 1Z1c. | CKD G4A3 - chronic kidney disease with glomerular filtration rate category G4 and albuminuria category A3 | 0 |
| 1Z1d. | CKD G5A1 - chronic kidney disease with glomerular filtration rate category G5 and albuminuria category A1 | 0 |
| 1Z1e. | CKD G5A2 - chronic kidney disease with glomerular filtration rate category G5 and albuminuria category A2 | 0 |
| 1Z1f. | CKD G5A3 - chronic kidney disease with glomerular filtration rate category G5 and albuminuria category A3 | 0 |
| K05.. | Chronic renal failure | 0 |
| K050. | End stage renal failure | 0 |
| K053. | Chronic kidney disease stage 3 | 0 |
| K054. | Chronic kidney disease stage 4 | 0 |
| K055. | Chronic kidney disease stage 5 | 0 |

---

#### CONCEPT: Coeliac Disease¶

| READ\_CODE | DESCRIPTION\_SAIL | SENSITIVE\_CODE |
| --- | --- | --- |
| 6648. | Coeliac disease monitoring | 0 |
| 66480 | Coeliac disease annual review | 0 |
| 68W4. | Coeliac disease autoantibody profile positive | 0 |
| 8IAp. | Coeliac disease annual review declined | 0 |
| 9mB.. | Coeliac disease monitoring invitation | 0 |
| 9mB0. | Coeliac disease monitoring telephone invitation | 0 |
| 9mB1. | Coeliac disease monitoring invitation first letter | 0 |
| 9mB2. | Coeliac disease monitoring invitation second letter | 0 |
| 9mB3. | Coeliac disease monitoring invitation third letter | 0 |
| J690. | Coeliac disease | 0 |
| J6900 | Congenital coeliac disease | 0 |
| J6901 | Acquired coeliac disease | 0 |
| J690z | Coeliac disease NOS | 0 |

---

#### CONCEPT: Colon Cancer¶

| READ\_CODE | DESCRIPTION\_SAIL | SENSITIVE\_CODE |
| --- | --- | --- |
| 68W25 | Bowel scope (flexible sigmoidoscopy) screen: suspected cancer detected | 0 |
| 9Ow1. | Bowel cancer detected by national screening programme | 0 |
| B0720 | Malignant neoplasm of pharyngeal recess | 0 |
| B13.. | Malignant neoplasm of colon | 0 |
| B130. | Malignant neoplasm of hepatic flexure of colon | 0 |
| B131. | Malignant neoplasm of transverse colon | 0 |
| B132. | Malignant neoplasm of descending colon | 0 |
| B133. | Malignant neoplasm of sigmoid colon | 0 |
| B134. | Malignant neoplasm of caecum | 0 |
| B135. | Malignant neoplasm of appendix | 0 |
| B136. | Malignant neoplasm of ascending colon | 0 |
| B137. | Malignant neoplasm of splenic flexure of colon | 0 |
| B138. | Malignant neoplasm, overlapping lesion of colon | 0 |
| B139. | Hereditary nonpolyposis colon cancer | 0 |
| B13y. | Malignant neoplasm of other specified sites of colon | 0 |
| B13z. | Malignant neoplasm of colon NOS | 0 |
| B14.. | Malignant neoplasm of rectum, rectosigmoid junction and anus | 0 |
| B140. | Malignant neoplasm of rectosigmoid junction | 0 |
| B141. | Malignant neoplasm of rectum | 0 |
| B142. | Malignant neoplasm of anal canal | 0 |
| B1420 | Malignant neoplasm of cloacogenic zone | 0 |
| B143. | Malignant neoplasm of anus unspecified | 0 |
| B14y. | Malignant neoplasm of other sites of rectum, rectosigmoid junction and anus | 0 |
| B14z. | Malignant neoplasm of rectum, rectosigmoid junction and anus NOS | 0 |
| B18y2 | Malignant neoplasm of mesorectum | 0 |
| B1z0. | Malignant neoplasm of intestinal tract, part unspecified | 0 |
| B575. | Secondary malignant neoplasm of large intestine and rectum | 0 |
| B5750 | Secondary malignant neoplasm of colon | 0 |
| B5751 | Secondary malignant neoplasm of rectum | 0 |
| B575z | Secondary malignant neoplasm of large intestine or rectum NOS | 0 |
| B803. | Carcinoma in situ of colon | 0 |
| B8030 | Carcinoma in situ of hepatic flexure of colon | 0 |
| B8031 | Carcinoma in situ of transverse colon | 0 |
| B8032 | Carcinoma in situ of descending colon | 0 |
| B8033 | Carcinoma in situ of sigmoid colon | 0 |
| B8036 | Carcinoma in situ of ascending colon | 0 |
| B8037 | Carcinoma in situ of splenic flexure of colon | 0 |
| B8038 | High grade dysplasia of colon | 0 |
| B803z | Carcinoma in situ of colon NOS | 0 |
| B804. | Carcinoma in situ of rectum and rectosigmoid junction | 0 |
| B8040 | Carcinoma in situ of rectosigmoid junction | 0 |
| B8041 | Carcinoma in situ of rectum | 0 |
| B804z | Carcinoma in situ of rectum or rectosigmoid junction NOS | 0 |
| B902. | Neoplasm of uncertain behaviour of stomach, intestines and rectum | 0 |
| B9024 | Neoplasm of uncertain behaviour of colon | 0 |
| B9025 | Neoplasm of uncertain behaviour of rectum | 0 |
| B902z | Neoplasm of uncertain behaviour of stomach, intestine or rectum NOS | 0 |
| BB57. | [M]Adenocarcinoma, intestinal type | 0 |
| BB5N. | [M]Adenomatous and adenocarcinomatous polyps of colon | 0 |
| BB5Nz | [M]Adenomatous or adenocarcinomatous polyps of the colon NOS | 0 |
| PB5B. | Dysplasia of colon | 0 |
| ZV100 | [V]Personal history of malignant neoplasm of gastrointestinal tract | 0 |

---

#### CONCEPT: Congenital disease and chromosomal abnormalities¶

| READ\_CODE | DESCRIPTION\_SAIL | SENSITIVE\_CODE |
| --- | --- | --- |
| 12J3. | FH: Congenital CVS anomaly | 0 |
| 14AV. | History of ventricular septal defect | 0 |
| 21268 | Ostium secundum atrial septal defect resolved | 0 |
| 66g.. | Congenital heart condition monitoring | 0 |
| 7903y | Other specified atrial inversion operation for transposition of great vessels | 0 |
| 79063 | Closure of persistent ostium primum | 0 |
| 790A1 | Open atrial septostomy | 0 |
| 790N4 | Relief of left ventricular outflow tract obstruction | 0 |
| 9RD0. | Transfer of care from paediatric congenital heart service | 0 |
| G5y9. | Cardiac septal defect, acquired | 0 |
| L185. | Congenital cardiovascular disorders during pregnancy, childbirth and the puerperium | 0 |
| L1850 | Congenital cardiovascular disorder - unspecified whether during pregnancy or the puerperium | 0 |
| L1851 | Congenital cardiovascular disorder during pregnancy - baby delivered | 0 |
| L1852 | Congenital cardiovascular disorder in the puerperium - baby delivered during current episode of care | 0 |
| L1853 | Congenital cardiovascular disorder during pregnancy - baby not yet delivered | 0 |
| L1854 | Congenital cardiovascular disorder in the puerperium - baby delivered during previous episode of care | 0 |
| L185z | Congenital cardiovascular disorder during pregnancy, childbirth or the puerperium NOS | 0 |
| P5... | Bulbus cordis and cardiac septal closure anomalies | 0 |
| P50.. | Common aorto-pulmonary trunk | 0 |
| P500. | Absent septum between aorta and pulmonary artery | 0 |
| P501. | Aortic septal defect | 0 |
| P502. | Persistent truncus arteriosus | 0 |
| P50z. | Common aorto-pulmonary trunk NOS | 0 |
| P51.. | Transposition of great vessels | 0 |
| P510. | Total great vessel transposition | 0 |
| P511. | Double outlet right ventricle | 0 |
| P5110 | Double outlet right ventricle, unspecified | 0 |
| P5111 | Dextratransposition of aorta | 0 |
| P5112 | Incomplete great vessel transposition | 0 |
| P5113 | Taussig-Bing syndrome | 0 |
| P511z | Double outlet right ventricle NOS | 0 |
| P512. | Corrected great vessel transposition | 0 |
| P51y. | Other specified transposition of great vessels | 0 |
| P51z. | Great vessel transposition NOS | 0 |
| P52.. | Tetralogy of Fallot | 0 |
| P520. | Tetralogy of Fallot, unspecified | 0 |
| P521. | Pentalogy of Fallot | 0 |
| P52z. | Tetralogy of Fallot NOS | 0 |
| P53.. | Common ventricle | 0 |
| P54.. | Ventricular septal defect | 0 |
| P540. | Ventricular septal defect, unspecified | 0 |
| P541. | Interventricular septal defect | 0 |
| P542. | Left ventricle to right atrial communication | 0 |
| P543. | Eisenmenger's complex | 0 |
| P544. | Gerbode's defect | 0 |
| P545. | Roger's disease | 0 |
| P54y. | Other specified ventricular septal defect | 0 |
| P54z. | Ventricular septal defect NOS | 0 |
| P55.. | Ostium secundum atrial septal defect | 0 |
| P550. | Atrial septal defect NOS | 0 |
| P551. | Patent foramen ovale | 0 |
| P552. | Persistent ostium secundum | 0 |
| P553. | Lutembacher's syndrome | 0 |
| P55y. | Other specified ostium secundum atrial septal defect | 0 |
| P55z. | Ostium secundum atrial septal defect NOS | 0 |
| P56.. | Endocardial cushion defects | 0 |
| P560. | Endocardial cushion defects, unspecified | 0 |
| P561. | Ostium primum defect | 0 |
| P56y. | Other specified endocardial cushion defects | 0 |
| P56z. | Endocardial cushion defects NOS | 0 |
| P56z0 | Common atrium | 0 |
| P56z1 | Common atrioventricular canal | 0 |
| P56z2 | Common atrioventricular-type ventricular septal defect | 0 |
| P56zz | Endocardial cushion defects NOS | 0 |
| P57.. | Cor biloculare | 0 |
| P58.. | Double outlet left ventricle | 0 |
| P59.. | Isomerism of atrial appendages | 0 |
| P5X.. | Congenital malformations of cardiac chambers and connections | 0 |
| P5y.. | Other heart bulb and septal closure defect | 0 |
| P5z.. | Heart bulb or septal closure defects NOS | 0 |
| P6... | Other congenital heart anomalies | 0 |
| P60.. | Pulmonary valve anomalies | 0 |
| P600. | Pulmonary valve anomaly, unspecified | 0 |
| P601. | Congenital atresia of the pulmonary valve | 0 |
| P6010 | Hypoplasia of pulmonary valve | 0 |
| P601z | Congenital atresia of pulmonary valve NOS | 0 |
| P602. | Congenital pulmonary stenosis | 0 |
| P6020 | Congenital fusion of pulmonic cusps | 0 |
| P6021 | Congenital fusion of pulmonary valve segment | 0 |
| P602z | Congenital pulmonary stenosis NOS | 0 |
| P603. | Right hypoplastic heart syndrome | 0 |
| P60z. | Other pulmonary valve anomalies | 0 |
| P60z0 | Congenital insufficiency of the pulmonary valve | 0 |
| P60z1 | Fallot's trilogy | 0 |
| P60z2 | Supernumerary pulmonary valve cusps | 0 |
| P60zz | Other pulmonary valve anomaly NOS | 0 |
| P61.. | Congenital tricuspid atresia and stenosis | 0 |
| P610. | Congenital tricuspid atresia | 0 |
| P611. | Congenital tricuspid stenosis | 0 |
| P61z. | Congenital tricuspid atresia or stenosis NOS | 0 |
| P62.. | Ebstein's anomaly | 0 |
| P63.. | Congenital aortic valve stenosis | 0 |
| P64.. | Congenital aortic valve insufficiency | 0 |
| P640. | Congenital aortic valve insufficiency, unspecified | 0 |
| P641. | Bicuspid aortic valve | 0 |
| P64z. | Congenital aortic valve insufficiency NOS | 0 |
| P65.. | Congenital mitral stenosis | 0 |
| P650. | Congenital mitral stenosis, unspecified | 0 |
| P651. | Fused commissure of the mitral valve | 0 |
| P652. | Parachute deformity of the mitral valve | 0 |
| P653. | Supernumerary cusps of the mitral valve | 0 |
| P65z. | Congenital mitral stenosis NOS | 0 |
| P66.. | Congenital mitral insufficiency | 0 |
| P67.. | Hypoplastic left heart syndrome | 0 |
| P68.. | Congenital heart disease | 0 |
| P69.. | Left ventricular outflow tract obstruction | 0 |
| P6W.. | Congenital malformation of aortic and mitral valves unspecified | 0 |
| P6X.. | Congenital malformation of tricuspid valve, unspecified | 0 |
| P6y.. | Other specified heart anomalies | 0 |
| P6y0. | Subaortic stenosis | 0 |
| P6y1. | Cor triatriatum | 0 |
| P6y2. | Pulmonary infundibular stenosis | 0 |
| P6y3. | Obstructive heart anomaly NEC | 0 |
| P6y30 | Uhl's disease | 0 |
| P6y31 | Right ventricular outflow tract obstruction | 0 |
| P6y3z | Obstructive heart anomaly NEC NOS | 0 |
| P6y4. | Coronary artery anomaly | 0 |
| P6y40 | Congenital absence of coronary artery | 0 |
| P6y41 | Single coronary artery | 0 |
| P6y42 | Coronary artery from aorta | 0 |
| P6y43 | Coronary artery from pulmonary trunk | 0 |
| P6y44 | Anomalous coronary artery communication | 0 |
| P6y45 | Congenital coronary aneurysm | 0 |
| P6y46 | Congenital stricture of coronary artery | 0 |
| P6y4z | Coronary artery anomaly NOS | 0 |
| P6y5. | Congenital heart block | 0 |
| P6y50 | Congenital heart block, unspecified | 0 |
| P6y51 | Congenital complete atrio-ventricular heart block | 0 |
| P6y52 | Congenital incomplete atrio-ventricular heart block | 0 |
| P6y5z | Congenital heart block NOS | 0 |
| P6y6. | Heart and cardiac apex malposition | 0 |
| P6y60 | Dextrocardia | 0 |
| P6y61 | Levocardia | 0 |
| P6y62 | Mesocardia | 0 |
| P6y63 | Ectopia cordis | 0 |
| P6y64 | Abdominal heart | 0 |
| P6y6z | Heart or cardiac apex malposition NOS | 0 |
| P6y7. | Myocardial bridge of coronary artery | 0 |
| P6y8. | Congenital dextroposition of heart | 0 |
| P6yy. | Other specified heart anomalies | 0 |
| P6yy0 | Atresia of cardiac vein | 0 |
| P6yy1 | Hypoplasia of cardiac vein | 0 |
| P6yy2 | Congenital cardiomegaly | 0 |
| P6yy3 | Congenital left ventricular diverticulum | 0 |
| P6yy4 | Congenital pericardial defect | 0 |
| P6yy5 | Congenital anomaly of myocardium | 0 |
| P6yy6 | Congenital aneurysm of heart | 0 |
| P6yy7 | Atresia of heart valve NEC | 0 |
| P6yy8 | Cor triloculare | 0 |
| P6yy9 | Congenital epicardial cyst | 0 |
| P6yyA | Hemicardia | 0 |
| P6yyB | Supernumerary heart valve cusps NEC | 0 |
| P6yyC | Fusion of mitral valve cusps | 0 |
| P6yyD | Fusion of heart valve cusps NEC | 0 |
| P6yyz | Other specified heart anomalies NOS | 0 |
| P6z.. | Congenital heart anomaly NOS | 0 |
| P6z0. | Unspecified anomaly of heart valve | 0 |
| P6z1. | Anomalous bands of heart | 0 |
| P6z10 | Anomalous atrial bands | 0 |
| P6z11 | Anomalous ventricular bands | 0 |
| P6z1z | Anomalous bands of heart NOS | 0 |
| P6z2. | Acyanotic congenital heart disease NOS | 0 |
| P6z3. | Cyanotic congenital heart disease NOS | 0 |
| P6zz. | Congenital heart anomaly NOS | 0 |
| P7... | Other congenital circulatory system anomalies | 0 |
| P70.. | Patent ductus arteriosus | 0 |
| P71.. | Coarctation of aorta | 0 |
| P710. | Hypoplasia of aortic arch, unspecified | 0 |
| P711. | Preductal coarctation of aorta | 0 |
| P712. | Postductal coarctation of aorta | 0 |
| P713. | Interruption of aortic arch | 0 |
| P71z. | Coarctation of aorta NOS | 0 |
| P72.. | Other anomalies of aorta | 0 |
| P720. | Anomaly of aorta, unspecified | 0 |
| P721. | Aortic arch anomalies | 0 |
| P7210 | Anomalous origin of the aortic arch | 0 |
| P7211 | Dextraposition of aorta | 0 |
| P7212 | Double aortic arch | 0 |
| P7213 | Kommerell's diverticulum | 0 |
| P7214 | Persistent aortic arch convolutions | 0 |
| P7215 | Persistent right aortic arch | 0 |
| P7216 | Vascular ring, aorta | 0 |
| P7217 | Overriding aorta | 0 |
| P721z | Aortic arch anomalies NOS | 0 |
| P722. | Atresia and stenosis of aorta | 0 |
| P7220 | Congenital absence of aorta | 0 |
| P7221 | Aplasia of aorta | 0 |
| P7222 | Hypoplasia of aorta | 0 |
| P7223 | Stricture of aorta | 0 |
| P7224 | Supra-valvular aortic stenosis | 0 |
| P7225 | Atresia of aorta | 0 |
| P722z | Atresia or stenosis of aorta NOS | 0 |
| P72z. | Other anomalies of aorta NOS | 0 |
| P72z0 | Aneurysm of sinus of Valsalva | 0 |
| P72z1 | Congenital aneurysm of aorta | 0 |
| P72zz | Other anomaly of aorta NOS | 0 |
| P73.. | Pulmonary artery anomalies | 0 |
| P730. | Pulmonary artery anomaly, unspecified | 0 |
| P731. | Pulmonary artery agenesis | 0 |
| P732. | Pulmonary artery atresia | 0 |
| P733. | Coarctation of the pulmonary artery | 0 |
| P734. | Hypoplasia of the pulmonary artery | 0 |
| P735. | Stenosis of pulmonary artery | 0 |
| P736. | Pulmonary arterio-venous aneurysm | 0 |
| P737. | Pulmonary artery aneurysm | 0 |
| P738. | Atresia of pulmonary artery with septal defect | 0 |
| P73y. | Other specified anomaly of pulmonary artery | 0 |
| P73z. | Pulmonary artery anomaly NOS | 0 |
| P74.. | Anomalies of great veins | 0 |
| P740. | Anomaly of great veins, unspecified | 0 |
| P7400 | Anomaly of the pulmonary veins, unspecified | 0 |
| P7401 | Anomaly of the vena cava, unspecified | 0 |
| P740z | Unspecified anomaly of the great veins, NOS | 0 |
| P741. | Total anomalous pulmonary venous return - TAPVR | 0 |
| P7410 | Subdiaphragmatic total anomalous pulmonary venous return | 0 |
| P7411 | Supradiaphragmatic total anomalous pulmonary venous return | 0 |
| P741z | Total anomalous pulmonary venous return NOS | 0 |
| P742. | Partial anomalous pulmonary venous return | 0 |
| P743. | Anomalous portal vein termination | 0 |
| P744. | Portal vein - hepatic artery fistula | 0 |
| P74z. | Other great vein anomalies | 0 |
| P74z0 | Absence of inferior vena cava | 0 |
| P74z1 | Absence of superior vena cava | 0 |
| P74z2 | Stenosis of inferior vena cava | 0 |
| P74z3 | Stenosis of superior vena cava | 0 |
| P74z4 | Persistent left posterior cardinal vein | 0 |
| P74z5 | Persistent left superior vena cava | 0 |
| P74z6 | Scimitar syndrome | 0 |
| P74z7 | Transposition of pulmonary veins | 0 |
| P74z8 | Atresia of pulmonary vein | 0 |
| P74zz | Other great vein anomaly NOS | 0 |
| P76.. | Other peripheral vascular system anomalies | 0 |
| P760. | Absence of artery NEC | 0 |
| P761. | Anomaly of artery NEC | 0 |
| P762. | Atresia of artery NEC | 0 |
| P763. | Absence of vein NEC | 0 |
| P7X.. | Congenital malformation of great arteries, unspecified | 0 |
| PK33. | Complete situs inversus with dextrocardia | 0 |
| PK34. | Situs inversus with levocardia | 0 |
| Pyu20 | [X]Other congenital malformations of cardiac chambers and connections | 0 |
| Pyu21 | [X]Other congenital malformations of cardiac septa | 0 |
| Pyu22 | [X]Other congenital malformations of pulmonary valve | 0 |
| Pyu23 | [X]Other congenital malformations of tricuspid valve | 0 |
| Pyu24 | [X]Other congenital malformations of aortic and mitral valves | 0 |
| Pyu25 | [X]Other specified congenital malformations of the heart | 0 |
| Pyu26 | [X]Other congenital malformations of aorta | 0 |
| Pyu27 | [X]Other congenital malformations of pulmonary artery | 0 |
| Pyu28 | [X]Other congenital malformations of great arteries | 0 |
| Pyu29 | [X]Other congenital malformations of great veins | 0 |
| Pyu2F | [X]Congenital malformations of cardiac chambers and connections | 0 |
| Pyu2G | [X]Congenital malformation of tricuspid valve, unspecified | 0 |
| Pyu2H | [X]Congenital malformation of aortic and mitral valves unspecified | 0 |
| Pyu2J | [X]Congenital malformation of great arteries, unspecified | 0 |
| Q48y1 | Congenital cardiac failure | 0 |

---

#### CONCEPT: COPD (Chronic obstructive pulmonary disease)¶

| READ\_CODE | DESCRIPTION\_SAIL | SENSITIVE\_CODE |
| --- | --- | --- |
| 14B3. | H/O: chr.obstr. airway disease | 0 |
| 14OJ. | At risk of chronic obstructive pulmonary disease | 0 |
| 14OX. | At risk of chronic obstructive pulmonary disease exacerbation | 0 |
| 1J71. | Suspected chronic obstructive pulmonary disease | 0 |
| 661M3 | Chronic obstructive pulmonary disease self-management plan agreed | 0 |
| 661N3 | Chronic obstructive pulmonary disease self-management plan review | 0 |
| 66YB. | Chronic obstructive pulmonary disease monitoring | 0 |
| 66YB0 | Chronic obstructive pulmonary disease 3 monthly review | 0 |
| 66YB1 | Chronic obstructive pulmonary disease 6 monthly review | 0 |
| 66YB2 | Telehealth chronic obstructive pulmonary disease monitoring | 0 |
| 66YD. | Chronic obstructive pulmonary disease monitoring due | 0 |
| 66YI. | Chronic obstructive pulmonary disease self-management plan given | 0 |
| 66YL. | Chronic obstructive pulmonary disease follow-up | 0 |
| 66YM. | Chronic obstructive pulmonary disease annual review | 0 |
| 66YS. | Chronic obstructive pulmonary disease monitoring by nurse | 0 |
| 66YT. | Chronic obstructive pulmonary disease monitoring by doctor | 0 |
| 66Yd. | Chronic obstructive pulmonary disease accident and emergency attendance since last visit | 0 |
| 66Ye. | Emergency chronic obstructive pulmonary disease admission since last appointment | 0 |
| 66Yf. | Number of chronic obstructive pulmonary disease exacerbations in past year | 0 |
| 66Yg. | Chronic obstructive pulmonary disease disturbs sleep | 0 |
| 66Yh. | Chronic obstructive pulmonary disease does not disturb sleep | 0 |
| 66Yi. | Multiple chronic obstructive pulmonary disease emergency hospital admissions | 0 |
| 66Yz2 | Shared care chronic obstructive pulmonary disease monitoring | 0 |
| 679V. | Health education - chronic obstructive pulmonary disease | 0 |
| 8BMW. | Issue of chronic obstructive pulmonary disease rescue pack | 0 |
| 8CE6. | Chronic obstructive pulmonary disease leaflet given | 0 |
| 8CMV. | Has chronic obstructive pulmonary disease care plan | 0 |
| 8CMW5 | Chronic obstructive pulmonary disease care pathway | 0 |
| 8CR1. | Chronic obstructive pulmonary disease clinical management plan | 0 |
| 8CeD. | Preferred place of care for next exacerbation of chronic obstructive pulmonary disease | 0 |
| 8H2R. | Admit COPD emergency | 0 |
| 8Hkw. | Referral to chronic obstructive pulmonary disease community nursing team | 0 |
| 8IEZ. | Chronic obstructive pulmonary disease rescue pack declined | 0 |
| 9N4W. | DNA - Did not attend chronic obstructive pulmonary disease clinic | 0 |
| 9NgP. | On chronic obstructive pulmonary disease supportive care pathway | 0 |
| 9Nk70 | Seen in chronic obstructive pulmonary disease clinic | 0 |
| 9Oi.. | Chronic obstructive pulmonary disease monitoring administration | 0 |
| 9Oi0. | Chronic obstructive pulmonary disease monitoring 1st letter | 0 |
| 9Oi1. | Chronic obstructive pulmonary disease monitoring 2nd letter | 0 |
| 9Oi2. | Chronic obstructive pulmonary disease monitoring 3rd letter | 0 |
| 9Oi3. | Chronic obstructive pulmonary disease monitoring verbal invite | 0 |
| 9Oi4. | Chronic obstructive pulmonary disease monitoring phone invite | 0 |
| 9e03. | General practitioner out of hours service notified of chronic obstructive pulmonary disease care plan | 0 |
| 9kf.. | Chronic obstructive pulmonary disease - enhanced services administration | 0 |
| 9kf0. | Chronic obstructive pulmonary disease patient unsuitable for pulmonary rehabilitation - enhanced services administration | 0 |
| 9kf1. | Referred for chronic obstructive pulmonary disease structured smoking assessment - enhanced services administration | 0 |
| 9kf2. | Chronic obstructive pulmonary disease structured smoking assessment declined - enhanced services administration | 0 |
| H3... | Chronic obstructive pulmonary disease | 0 |
| H30.. | Bronchitis unspecified | 0 |
| H300. | Tracheobronchitis NOS | 0 |
| H301. | Laryngotracheobronchitis | 0 |
| H302. | Wheezy bronchitis | 0 |
| H30z. | Bronchitis NOS | 0 |
| H31.. | Chronic bronchitis | 0 |
| H310. | Simple chronic bronchitis | 0 |
| H3100 | Chronic catarrhal bronchitis | 0 |
| H3101 | Smokers' cough | 0 |
| H310z | Simple chronic bronchitis NOS | 0 |
| H311. | Mucopurulent chronic bronchitis | 0 |
| H3110 | Purulent chronic bronchitis | 0 |
| H3111 | Fetid chronic bronchitis | 0 |
| H311z | Mucopurulent chronic bronchitis NOS | 0 |
| H312. | Obstructive chronic bronchitis | 0 |
| H3120 | Chronic asthmatic bronchitis | 0 |
| H3121 | Emphysematous bronchitis | 0 |
| H3122 | Acute exacerbation of chronic obstructive airways disease | 0 |
| H312z | Obstructive chronic bronchitis NOS | 0 |
| H313. | Mixed simple and mucopurulent chronic bronchitis | 0 |
| H31y. | Other chronic bronchitis | 0 |
| H31y0 | Chronic tracheitis | 0 |
| H31y1 | Chronic tracheobronchitis | 0 |
| H31yz | Other chronic bronchitis NOS | 0 |
| H31z. | Chronic bronchitis NOS | 0 |
| H32.. | Emphysema | 0 |
| H320. | Chronic bullous emphysema | 0 |
| H3200 | Segmental bullous emphysema | 0 |
| H3201 | Zonal bullous emphysema | 0 |
| H3202 | Giant bullous emphysema | 0 |
| H3203 | Bullous emphysema with collapse | 0 |
| H320z | Chronic bullous emphysema NOS | 0 |
| H321. | Panlobular emphysema | 0 |
| H322. | Centrilobular emphysema | 0 |
| H32y. | Other emphysema | 0 |
| H32y0 | Acute vesicular emphysema | 0 |
| H32y1 | Atrophic (senile) emphysema | 0 |
| H32y2 | MacLeod's unilateral emphysema | 0 |
| H32yz | Other emphysema NOS | 0 |
| H32z. | Emphysema NOS | 0 |
| H36.. | Mild chronic obstructive pulmonary disease | 0 |
| H37.. | Moderate chronic obstructive pulmonary disease | 0 |
| H38.. | Severe chronic obstructive pulmonary disease | 0 |
| H39.. | Very severe chronic obstructive pulmonary disease | 0 |
| H3A.. | End stage chronic obstructive airways disease | 0 |
| H3B.. | Asthma-chronic obstructive pulmonary disease overlap syndrome | 0 |
| H3y.. | Other specified chronic obstructive airways disease | 0 |
| H3y0. | Chronic obstructive pulmonary disease with acute lower respiratory infection | 0 |
| H3y1. | Chronic obstructive pulmonary disease with acute exacerbation, unspecified | 0 |
| H3z.. | Chronic obstructive airways disease NOS | 0 |
| H4640 | Chronic emphysema due to chemical fumes | 0 |
| H4641 | Obliterative bronchiolitis due to chemical fumes | 0 |
| H581. | Interstitial emphysema | 0 |
| H5832 | Eosinophilic bronchitis | 0 |
| Hyu3. | [X]Chronic lower respiratory diseases | 0 |
| Hyu30 | [X]Other emphysema | 0 |
| Hyu31 | [X]Other specified chronic obstructive pulmonary disease | 0 |

---

#### CONCEPT: Coronary heart disease¶

| READ\_CODE | DESCRIPTION\_SAIL | SENSITIVE\_CODE |
| --- | --- | --- |
| 14A.. | H/O: cardiovascular disease | 0 |
| 14A5. | H/O: angina pectoris | 0 |
| 14AA. | H/O: heart disease NOS | 0 |
| 14AJ. | H/O: Angina in last year | 0 |
| 14AL. | H/O: Treatment for ischaemic heart disease | 0 |
| 14AW. | H/O acute coronary syndrome | 0 |
| 14Oh0 | High risk of cardiovascular disease | 0 |
| 187.. | Frequency of angina | 0 |
| 1J61. | Suspected ischaemic heart disease | 0 |
| 33B95 | Exercise tolerance test abnormal | 0 |
| 3889. | Euroscore for angina | 0 |
| 388E. | Canadian Cardiovascular Society classification of angina | 0 |
| 44H3. | Cardiac enzymes abnormal | 0 |
| 44H30 | Cardiac enzymes abnormal - first set | 0 |
| 5533. | Angiocardiography abnormal | 0 |
| 5543. | Coronary arteriograph.abnormal | 0 |
| 5C11. | Radionuclide heart study abnormal | 0 |
| 661M0 | Angina self-management plan agreed | 0 |
| 661N0 | Angina self-management plan review | 0 |
| 662K. | Angina control | 0 |
| 662K0 | Angina control - good | 0 |
| 662K1 | Angina control - poor | 0 |
| 662K2 | Angina control - improving | 0 |
| 662K3 | Angina control - worsening | 0 |
| 662K4 | Angina self management plan commenced | 0 |
| 662K5 | Angina self management plan completed | 0 |
| 662Kz | Angina control NOS | 0 |
| 68B2. | Ischaemic heart disease screen | 0 |
| 6A2.. | Coronary heart disease annual review | 0 |
| 6A4.. | Coronary heart disease review | 0 |
| 8B27. | Antianginal therapy | 0 |
| 8B3k. | Coronary heart disease medication review | 0 |
| 8B63. | Salicylate prophylaxis | 0 |
| 8F9.. | Cardiac rehabilitation | 0 |
| 8F90. | Cardiac rehabilitation - phase 1 | 0 |
| 8F91. | Cardiac rehabilitation - phase 2 | 0 |
| 8F92. | Cardiac rehabilitation - phase 3 | 0 |
| 8F93. | Cardiac rehabilitation - phase 4 | 0 |
| 8F97. | Cardiac rehabilitation programme completed | 0 |
| 8F98. | Cardiac rehabilitation programme offered | 0 |
| 8F99. | Cardiac rehabilitation initial assessment offered | 0 |
| 8H11. | Admit to cardiac ITU | 0 |
| 8H2Q. | Admit cardiology emergency | 0 |
| 8H2V. | Admit ischaemic heart disease emergency | 0 |
| 8H440 | Referral to cardiology multidisciplinary team | 0 |
| 8H7v. | Referral to cardiac rehabilitation nurse | 0 |
| 8Hkk. | Referral to cardiac rehabilitation programme | 0 |
| 8Hkl. | Referral to cardiac rehabilitation service by secondary care | 0 |
| 8Hkt. | Referral to community cardiology service | 0 |
| 8I3a. | Cardiac rehabilitation declined | 0 |
| 8IB9. | Referral to cardiac rehabilitation programme not indicated | 0 |
| 8IE3. | Referral to cardiac rehabilitation programme declined | 0 |
| 8IEY. | Referral to Angina Plan self-management programme declined | 0 |
| 8T04. | Referral to Angina Plan self-management programme | 0 |
| G.... | Circulatory system diseases | 0 |
| G3... | Ischaemic heart disease | 0 |
| G30.. | Acute myocardial infarction | 0 |
| G30B. | Acute posterolateral myocardial infarction | 0 |
| G31.. | Other acute and subacute ischaemic heart disease | 0 |
| G310. | Postmyocardial infarction syndrome | 0 |
| G311. | Preinfarction syndrome | 0 |
| G3110 | Myocardial infarction aborted | 0 |
| G3111 | Unstable angina | 0 |
| G3112 | Angina at rest | 0 |
| G3113 | Refractory angina | 0 |
| G3114 | Worsening angina | 0 |
| G3115 | Acute coronary syndrome | 0 |
| G311z | Preinfarction syndrome NOS | 0 |
| G312. | Coronary thrombosis not resulting in myocardial infarction | 0 |
| G31y. | Other acute and subacute ischaemic heart disease | 0 |
| G31y0 | Acute coronary insufficiency | 0 |
| G31y1 | Microinfarction of heart | 0 |
| G31y2 | Subendocardial ischaemia | 0 |
| G31y3 | Transient myocardial ischaemia | 0 |
| G31yz | Other acute and subacute ischaemic heart disease NOS | 0 |
| G32.. | Old myocardial infarction | 0 |
| G33.. | Angina pectoris | 0 |
| G330. | Angina decubitus | 0 |
| G3300 | Nocturnal angina | 0 |
| G330z | Angina decubitus NOS | 0 |
| G331. | Prinzmetal's angina | 0 |
| G332. | Coronary artery spasm | 0 |
| G33z. | Angina pectoris NOS | 0 |
| G33z0 | Status anginosus | 0 |
| G33z1 | Stenocardia | 0 |
| G33z2 | Syncope anginosa | 0 |
| G33z3 | Angina on effort | 0 |
| G33z4 | Ischaemic chest pain | 0 |
| G33z5 | Post infarct angina | 0 |
| G33z6 | New onset angina | 0 |
| G33z7 | Stable angina | 0 |
| G33zz | Angina pectoris NOS | 0 |
| G34.. | Other chronic ischaemic heart disease | 0 |
| G340. | Coronary atherosclerosis | 0 |
| G3400 | Single coronary vessel disease | 0 |
| G3401 | Double coronary vessel disease | 0 |
| G341. | Aneurysm of heart | 0 |
| G3410 | Ventricular cardiac aneurysm | 0 |
| G3411 | Other cardiac wall aneurysm | 0 |
| G3412 | Aneurysm of coronary vessels | 0 |
| G341z | Aneurysm of heart NOS | 0 |
| G342. | Atherosclerotic cardiovascular disease | 0 |
| G343. | Ischaemic cardiomyopathy | 0 |
| G344. | Silent myocardial ischaemia | 0 |
| G34y. | Other specified chronic ischaemic heart disease | 0 |
| G34y0 | Chronic coronary insufficiency | 0 |
| G34y1 | Chronic myocardial ischaemia | 0 |
| G34yz | Other specified chronic ischaemic heart disease NOS | 0 |
| G34z. | Other chronic ischaemic heart disease NOS | 0 |
| G34z0 | Asymptomatic coronary heart disease | 0 |
| G35.. | Subsequent myocardial infarction | 0 |
| G350. | Subsequent myocardial infarction of anterior wall | 0 |
| G351. | Subsequent myocardial infarction of inferior wall | 0 |
| G353. | Subsequent myocardial infarction of other sites | 0 |
| G35X. | Subsequent myocardial infarction of unspecified site | 0 |
| G36.. | Certain current complications following acute myocardial infarction | 0 |
| G360. | Haemopericardium as current complication following acute myocardial infarction | 0 |
| G361. | Atrial septal defect as current complication following acute myocardial infarction | 0 |
| G362. | Ventricular septal defect as current complication following acute myocardial infarction | 0 |
| G363. | Rupture of cardiac wall without haemopericardium as current complication following acute myocardial infarction | 0 |
| G364. | Rupture of chordae tendinae as current complication following acute myocardial infarction | 0 |
| G365. | Rupture of papillary muscle as current complication following acute myocardial infarction | 0 |
| G366. | Thrombosis of atrium, auricular appendage, and ventricle as current complications following acute myocardial infarction | 0 |
| G37.. | Cardiac syndrome X | 0 |
| G38.. | Postoperative myocardial infarction | 0 |
| G380. | Postoperative transmural myocardial infarction of anterior wall | 0 |
| G381. | Postoperative transmural myocardial infarction of inferior wall | 0 |
| G382. | Postoperative transmural myocardial infarction of other sites | 0 |
| G383. | Postoperative transmural myocardial infarction of unspecified site | 0 |
| G384. | Postoperative subendocardial myocardial infarction | 0 |
| G38z. | Postoperative myocardial infarction, unspecified | 0 |
| G39.. | Coronary microvascular disease | 0 |
| G3y.. | Other specified ischaemic heart disease | 0 |
| G3z.. | Ischaemic heart disease NOS | 0 |
| G501. | Post infarction pericarditis | 0 |
| G5y2. | Cardiovascular arteriosclerosis unspecified | 0 |
| G72B1 | Coronary artery dissection | 0 |
| Gyu3. | [X]Ischaemic heart diseases | 0 |
| Gyu30 | [X]Other forms of angina pectoris | 0 |
| Gyu31 | [X]Other current complications following acute myocardial infarction | 0 |
| Gyu32 | [X]Other forms of acute ischaemic heart disease | 0 |
| Gyu33 | [X]Other forms of chronic ischaemic heart disease | 0 |
| Gyu35 | [X]Subsequent myocardial infarction of other sites | 0 |
| Gyu36 | [X]Subsequent myocardial infarction of unspecified site | 0 |
| J421. | Chronic intestinal vascular insufficiency | 0 |
| ZV579 | [V]Cardiac rehabilitation | 0 |

---

#### CONCEPT: Cystic fibrosis¶

| READ\_CODE | DESCRIPTION\_SAIL | SENSITIVE\_CODE |
| --- | --- | --- |
| 66k.. | Cystic fibrosis monitoring | 0 |
| 66k0. | Cystic fibrosis annual review | 0 |
| C10N1 | Cystic fibrosis related diabetes mellitus | 0 |
| C370. | Cystic fibrosis | 0 |
| C3700 | Cystic fibrosis with no meconium ileus | 0 |
| C3701 | Cystic fibrosis with meconium ileus | 0 |
| C3702 | Cystic fibrosis with pulmonary manifestations | 0 |
| C3703 | Cystic fibrosis with intestinal manifestations | 0 |
| C3704 | Arthropathy in cystic fibrosis | 0 |
| C3705 | Cystic fibrosis with distal intestinal obstruction syndrome | 0 |
| C3707 | Liver disease due to cystic fibrosis | 0 |
| C3708 | Cystic fibrosis related cirrhosis | 0 |
| C3709 | Exacerbation of cystic fibrosis | 0 |
| C370y | Cystic fibrosis with other manifestations | 0 |
| C370z | Cystic fibrosis NOS | 0 |
| Q4352 | Perinatal jaundice due to mucoviscidosis | 0 |

---

#### CONCEPT: Deafness¶

| READ\_CODE | DESCRIPTION\_SAIL | SENSITIVE\_CODE |
| --- | --- | --- |
| 13ZL. | Using lip-reading | 0 |
| 13ZM. | Using British sign language | 0 |
| 13ZP. | Using Makaton sign language | 0 |
| 13o5. | Preferred method of communication: Makaton | 0 |
| 13o6. | Preferred method of communication: British Sign Language | 0 |
| 13o85 | Uses British Sign Language interpreter | 0 |
| 13o8M | Uses sign language | 0 |
| 1493. | H/O: hearing problem | 0 |
| 1C12. | Hearing difficulty | 0 |
| 1C13. | Deafness | 0 |
| 1C131 | Unilateral deafness | 0 |
| 1C132 | Partial deafness | 0 |
| 1C133 | Bilateral deafness | 0 |
| 1C16. | Deteriorating hearing | 0 |
| 1C17. | Hearing aid problem | 0 |
| 1C18. | Difficulty hearing with background noise | 0 |
| 1C19. | Difficulty hearing whispers | 0 |
| 1C1Z. | Hearing symptom NOS | 0 |
| 2BL2. | O/E - slightly deaf | 0 |
| 2BL3. | O/E - significantly deaf | 0 |
| 2BL4. | O/E - very deaf | 0 |
| 2BL5. | O/E - completely deaf | 0 |
| 2BM2. | O/E -tune fork=conductive deaf | 0 |
| 2BM3. | O/E tune fork=perceptive deaf | 0 |
| 2BM4. | O/E - High tone deafness | 0 |
| 2DG.. | Hearing aid worn | 0 |
| 2DH0. | Uses hearing loop | 0 |
| 2DH1. | Does use hearing aid | 0 |
| 31343 | Hearing test abnormal | 0 |
| 31344 | Hearing test bilateral abnormality | 0 |
| 31345 | Hearing test left abnormality | 0 |
| 31346 | Hearing test right abnormality | 0 |
| 73084 | Placement of hearing implant in external ear | 0 |
| 73085 | Attention to hearing implant in external ear | 0 |
| 73086 | Removal of hearing implant from external ear | 0 |
| 7311A | Insertion of bone anchors for subcutaneous bone anchored hearing aid | 0 |
| 7317C | Placement of hearing implant in middle ear | 0 |
| 7317D | Attention to hearing implant in middle ear | 0 |
| 7317E | Removal of hearing implant from middle ear | 0 |
| 7319. | Attachment of bone anchored hearing prosthesis | 0 |
| 73190 | Insertion of fixtures for bone anchored hearing prosthesis Stage 1 | 0 |
| 73191 | Insertion of fixtures for bone anchored hearing prosthesis Stage 2 | 0 |
| 73192 | Reduction of soft tissue for bone anchored hearing prosthesis | 0 |
| 73193 | Attention to fixtures for bone anchored hearing prosthesis | 0 |
| 73194 | One stage insertion of fixtures for bone anchored hearing prosthesis | 0 |
| 73195 | Fitting of external hearing prosthesis to bone anchored fixtures | 0 |
| 73196 | First stage insertion of fixtures for bone anchored hearing prosthesis | 0 |
| 7319y | Other specified attachment of bone anchored hearing prosthesis | 0 |
| 7319z | Attachment of bone anchored hearing prosthesis NOS | 0 |
| 73210 | Implantation of intracochlear prosthesis | 0 |
| 73211 | Implantation of extracochlear prosthesis | 0 |
| 73212 | Attention to cochlear prosthesis | 0 |
| 73218 | Removal of cochlear prosthesis | 0 |
| 8D2.. | Auditory aid | 0 |
| 8D21. | Provide head worn hearing aid | 0 |
| 8D22. | Provide body worn hearing aid | 0 |
| 8D23. | Ear fitting hearing aid | 0 |
| 8D24. | Replace hearing aid battery | 0 |
| 8D26. | Provision of replacement hearing aid | 0 |
| 8E31. | Lip reading training | 0 |
| 8M41. | Hearing aid requested | 0 |
| 8O0E. | Provision of support as Makaton user | 0 |
| 8O0F. | Provision of support as signing user | 0 |
| 9NUw. | Interpreter needed - British Sign Language | 0 |
| 9NUx. | Interpreter needed - Makaton Sign Language | 0 |
| 9NfL. | Requires information in Makaton | 0 |
| 9NnR. | Visual frame sign language interpreter needed | 0 |
| 9NnS. | Hands-on signing interpreter needed | 0 |
| F5801 | Presbyacusis | 0 |
| F5812 | Noise-induced hearing loss | 0 |
| F582. | Unspecified sudden hearing loss | 0 |
| F59.. | Hearing loss | 0 |
| F590. | Conductive hearing loss | 0 |
| F5900 | Unspecified conductive hearing loss | 0 |
| F5901 | Conductive hearing loss due to disorder of external ear | 0 |
| F5902 | Conductive hearing loss due to disorder of tympanic membrane | 0 |
| F5903 | Conductive hearing loss due to disorder of middle ear | 0 |
| F5904 | Conductive hearing loss due to disorder of inner ear | 0 |
| F5905 | Conductive hearing loss, bilateral | 0 |
| F5906 | Conductive hearing loss, unilateral with unrestricted hearing on the contralateral side | 0 |
| F590y | Combined conductive hearing loss | 0 |
| F590z | Conductive hearing loss NOS | 0 |
| F591. | Sensorineural hearing loss | 0 |
| F5910 | Unspecified perceptive hearing loss | 0 |
| F5911 | Sensory hearing loss | 0 |
| F5912 | Neural hearing loss | 0 |
| F5913 | Central hearing loss | 0 |
| F5914 | Congenital sensorineural deafness | 0 |
| F5915 | Ototoxicity - deafness | 0 |
| F5916 | Sensorineural hearing loss, bilateral | 0 |
| F5917 | Sensorineural hearing loss, unilateral with unrestricted hearing on the contralateral side | 0 |
| F5918 | Congenital prelingual deafness | 0 |
| F5919 | Bilateral profound sensorineural hearing loss | 0 |
| F591B | Profound sensorineural hearing loss | 0 |
| F591E | Severe sensorineural hearing loss | 0 |
| F591y | Combined perceptive hearing loss | 0 |
| F591z | Perceptive hearing loss NOS | 0 |
| F592. | Mixed conductive and sensorineural deafness | 0 |
| F5920 | Mixed conductive and sensorineural hearing loss, unilateral with unrestricted hearing on the contralateral side | 0 |
| F5921 | Mixed conductive and sensorineural hearing loss, bilateral | 0 |
| F593. | Deaf mutism, NEC | 0 |
| F594. | High frequency deafness | 0 |
| F595. | Low frequency deafness | 0 |
| F596. | Maternally inherited deafness | 0 |
| F597. | Mild acquired hearing loss | 0 |
| F598. | Moderate acquired hearing loss | 0 |
| F599. | Severe acquired hearing loss | 0 |
| F59A. | Profound acquired hearing loss | 0 |
| F59y. | Other specified forms of hearing loss | 0 |
| F59z. | Deafness NOS | 0 |
| F5A.. | Hearing impairment | 0 |
| FyuU0 | [X]Deaf mutism, not elsewhere classified | 0 |
| FyuU1 | [X]Other specified hearing loss | 0 |
| P40.. | Ear anomalies with hearing impairment | 0 |
| P400. | Ear anomalies with hearing impaired, unspecified | 0 |
| P402. | Other external ear anomaly with hearing impairment | 0 |
| P402z | Other external ear anomaly with hearing impairment NOS | 0 |
| P40z. | Other and unspecified ear anomaly with hearing impaired | 0 |
| P40zz | Ear anomaly with hearing impaired NOS | 0 |
| ZV412 | [V]Problems with hearing | 0 |
| ZV45G | [V]Presence of external hearing-aid | 0 |
| ZV45N | [V]Bone anchored hearing aid in situ | 0 |
| ZV532 | [V]Fitting or adjustment of hearing aid | 0 |
| ZV53D | [V]Adjustment and management of implanted hearing device | 0 |

---

#### CONCEPT: Dementia alzheimer¶

| READ\_CODE | DESCRIPTION\_SAIL | SENSITIVE\_CODE |
| --- | --- | --- |
| 1461. | H/O: dementia | 0 |
| 66h.. | Dementia monitoring | 0 |
| 6AB.. | Dementia annual review | 0 |
| 8BM02 | Dementia medication review | 0 |
| 8BPa. | Antipsychotic drug therapy for dementia | 0 |
| 8CMG2 | Review of dementia advance care plan | 0 |
| 8CMZ. | Dementia care plan | 0 |
| 8CMZ0 | Dementia care plan agreed | 0 |
| 8CMZ1 | Dementia care plan reviewed | 0 |
| 8CMZ2 | Dementia care plan declined | 0 |
| 8CMZ3 | Dementia care plan review declined | 0 |
| 8CMe0 | Dementia advance care plan | 0 |
| 8CSA. | Dementia advance care plan agreed | 0 |
| 8IAe0 | Dementia advance care plan declined | 0 |
| 8IAe2 | Dementia advance care plan review declined | 0 |
| 9Ou.. | Dementia monitoring administration | 0 |
| 9Ou1. | Dementia monitoring first letter | 0 |
| 9Ou2. | Dementia monitoring second letter | 0 |
| 9Ou3. | Dementia monitoring third letter | 0 |
| 9Ou4. | Dementia monitoring verbal invite | 0 |
| 9Ou5. | Dementia monitoring telephone invite | 0 |
| 9hD.. | Exception reporting: dementia quality indicators | 0 |
| 9hD0. | Excepted from dementia quality indicators: Patient unsuitable | 0 |
| 9hD1. | Excepted from dementia quality indicators: Informed dissent | 0 |
| A411. | Jakob-Creutzfeldt disease | 0 |
| A4110 | Sporadic Creutzfeldt-Jakob disease | 0 |
| E00.. | Senile and presenile organic psychotic conditions | 0 |
| E000. | Uncomplicated senile dementia | 0 |
| E001. | Presenile dementia | 0 |
| E0010 | Uncomplicated presenile dementia | 0 |
| E0011 | Presenile dementia with delirium | 0 |
| E0012 | Presenile dementia with paranoia | 0 |
| E0013 | Presenile dementia with depression | 0 |
| E001z | Presenile dementia NOS | 0 |
| E002. | Senile dementia with depressive or paranoid features | 0 |
| E0020 | Senile dementia with paranoia | 0 |
| E0021 | Senile dementia with depression | 0 |
| E002z | Senile dementia with depressive or paranoid features NOS | 0 |
| E003. | Senile dementia with delirium | 0 |
| E004. | Arteriosclerotic dementia | 0 |
| E0040 | Uncomplicated arteriosclerotic dementia | 0 |
| E0041 | Arteriosclerotic dementia with delirium | 0 |
| E0042 | Arteriosclerotic dementia with paranoia | 0 |
| E0043 | Arteriosclerotic dementia with depression | 0 |
| E004z | Arteriosclerotic dementia NOS | 0 |
| E00z. | Senile or presenile psychoses NOS | 0 |
| E012. | Other alcoholic dementia | 0 |
| E02y1 | Drug-induced dementia | 0 |
| E040. | Non-alcoholic amnestic syndrome | 0 |
| E041. | Dementia in conditions EC | 0 |
| Eu00. | [X]Dementia in Alzheimer's disease | 0 |
| Eu000 | [X]Dementia in Alzheimer's disease with early onset | 0 |
| Eu001 | [X]Dementia in Alzheimer's disease with late onset | 0 |
| Eu002 | [X]Dementia in Alzheimer's dis, atypical or mixed type | 0 |
| Eu00z | [X]Dementia in Alzheimer's disease, unspecified | 0 |
| Eu01. | [X]Vascular dementia | 0 |
| Eu010 | [X]Vascular dementia of acute onset | 0 |
| Eu011 | [X]Multi-infarct dementia | 0 |
| Eu012 | [X]Subcortical vascular dementia | 0 |
| Eu013 | [X]Mixed cortical and subcortical vascular dementia | 0 |
| Eu01y | [X]Other vascular dementia | 0 |
| Eu01z | [X]Vascular dementia, unspecified | 0 |
| Eu02. | [X]Dementia in other diseases classified elsewhere | 0 |
| Eu020 | [X]Dementia in Pick's disease | 0 |
| Eu021 | [X]Dementia in Creutzfeldt-Jakob disease | 0 |
| Eu022 | [X]Dementia in Huntington's disease | 0 |
| Eu023 | [X]Dementia in Parkinson's disease | 0 |
| Eu024 | [X]Dementia in human immunodef virus [HIV] disease | 1 |
| Eu025 | [X]Lewy body dementia | 0 |
| Eu02y | [X]Dementia in other specified diseases classif elsewhere | 0 |
| Eu02z | [X] Unspecified dementia | 0 |
| Eu041 | [X]Delirium superimposed on dementia | 0 |
| Eu107 | [X]Mental and behavioural disorders due to use of alcohol: residual and late-onset psychotic disorder | 0 |
| F110. | Alzheimer's disease | 0 |
| F1100 | Alzheimer's disease with early onset | 0 |
| F1101 | Alzheimer's disease with late onset | 0 |
| F111. | Pick's disease | 0 |
| F112. | Senile degeneration of brain | 0 |
| F1130 | Normal pressure hydrocephalus | 0 |
| F116. | Lewy body disease | 0 |
| F118. | Frontotemporal degeneration | 0 |
| F11x7 | Cerebral degeneration due to Jakob - Creutzfeldt disease | 0 |
| Fyu30 | [X]Other Alzheimer's disease | 0 |

---

#### CONCEPT: Depression¶

| READ\_CODE | DESCRIPTION\_SAIL | SENSITIVE\_CODE |
| --- | --- | --- |
| 12K8. | Maternal postnatal depression | 0 |
| 1465. | H/O: depression | 0 |
| 1B17. | Depressed | 0 |
| 1B1U. | Symptoms of depression | 0 |
| 1BT.. | Depressed mood | 0 |
| 212S. | Depression resolved | 0 |
| 2257. | O/E - depressed | 0 |
| 62T1. | Puerperal depression | 0 |
| 66590 | Antidepressant drug treatment started | 0 |
| 6G00. | Postnatal depression counselling | 0 |
| 8BK0. | Depression management programme | 0 |
| 8CAa. | Patient given advice about management of depression | 0 |
| 8HHq. | Referral for guided self-help for depression | 0 |
| 8HHq0 | Referral for depression self-help video | 0 |
| 8IH52 | Referral for guided self-help for depression declined | 0 |
| 9H90. | Depression annual review | 0 |
| 9H91. | Depression medication review | 0 |
| 9H92. | Depression interim review | 0 |
| 9HA0. | On depression register | 0 |
| 9Ov.. | Depression monitoring administration | 0 |
| 9Ov0. | Depression monitoring first letter | 0 |
| 9Ov1. | Depression monitoring second letter | 0 |
| 9Ov2. | Depression monitoring third letter | 0 |
| 9Ov3. | Depression monitoring verbal invite | 0 |
| 9Ov4. | Depression monitoring telephone invite | 0 |
| 9k4.. | Depression - enhanced services administration | 0 |
| 9k40. | Depression - enhanced service completed | 0 |
| 9kQ.. | On full dose long term treatment for depression - enhanced services administration | 0 |
| E0013 | Presenile dementia with depression | 0 |
| E002. | Senile dementia with depressive or paranoid features | 0 |
| E0021 | Senile dementia with depression | 0 |
| E002z | Senile dementia with depressive or paranoid features NOS | 0 |
| E0043 | Arteriosclerotic dementia with depression | 0 |
| E02y3 | Drug-induced depressive state | 0 |
| E11.. | Affective psychoses | 0 |
| E112. | Single major depressive episode | 0 |
| E1120 | Single major depressive episode, unspecified | 0 |
| E1121 | Single major depressive episode, mild | 0 |
| E1122 | Single major depressive episode, moderate | 0 |
| E1123 | Single major depressive episode, severe, without mention of psychosis | 0 |
| E1124 | Single major depressive episode, severe, with psychosis | 0 |
| E1125 | Single major depressive episode, in partial or unspecified remission | 0 |
| E1126 | Single major depressive episode, in full remission | 0 |
| E112z | Single major depressive episode NOS | 0 |
| E113. | Recurrent major depressive episode | 0 |
| E1130 | Recurrent major depressive episodes, unspecified | 0 |
| E1131 | Recurrent major depressive episodes, mild | 0 |
| E1132 | Recurrent major depressive episodes, moderate | 0 |
| E1133 | Recurrent major depressive episodes, severe, without mention of psychosis | 0 |
| E1134 | Recurrent major depressive episodes, severe, with psychosis | 0 |
| E1135 | Recurrent major depressive episodes, in partial or unspecified remission | 0 |
| E1136 | Recurrent major depressive episodes, in full remission | 0 |
| E1137 | Recurrent depression | 0 |
| E113z | Recurrent major depressive episode NOS | 0 |
| E118. | Seasonal affective disorder | 0 |
| E11y2 | Atypical depressive disorder | 0 |
| E11z2 | Masked depression | 0 |
| E130. | Reactive depressive psychosis | 0 |
| E135. | Agitated depression | 0 |
| E2003 | Anxiety with depression | 0 |
| E204. | Neurotic depression reactive type | 0 |
| E290. | Brief depressive reaction | 0 |
| E290z | Brief depressive reaction NOS | 0 |
| E291. | Prolonged depressive reaction | 0 |
| E2B.. | Depressive disorder NEC | 0 |
| E2B0. | Postviral depression | 0 |
| E2B1. | Chronic depression | 0 |
| Eu204 | [X]Post-schizophrenic depression | 0 |
| Eu251 | [X]Schizoaffective disorder, depressive type | 0 |
| Eu32. | [X]Depressive episode | 0 |
| Eu320 | [X]Mild depressive episode | 0 |
| Eu321 | [X]Moderate depressive episode | 0 |
| Eu322 | [X]Severe depressive episode without psychotic symptoms | 0 |
| Eu323 | [X]Severe depressive episode with psychotic symptoms | 0 |
| Eu324 | [X]Mild depression | 0 |
| Eu325 | [X]Major depression, mild | 0 |
| Eu326 | [X]Major depression, moderately severe | 0 |
| Eu327 | [X]Major depression, severe without psychotic symptoms | 0 |
| Eu328 | [X]Major depression, severe with psychotic symptoms | 0 |
| Eu329 | [X]Single major depressive episode, severe, with psychosis, psychosis in remission | 0 |
| Eu32A | [X]Recurrent major depressive episodes, severe, with psychosis, psychosis in remission | 0 |
| Eu32B | [X]Antenatal depression | 0 |
| Eu32y | [X]Other depressive episodes | 0 |
| Eu32z | [X]Depressive episode, unspecified | 0 |
| Eu33. | [X]Recurrent depressive disorder | 0 |
| Eu330 | [X]Recurrent depressive disorder, current episode mild | 0 |
| Eu331 | [X]Recurrent depressive disorder, current episode moderate | 0 |
| Eu332 | [X]Recurrent depressive disorder, current episode severe without psychotic symptoms | 0 |
| Eu333 | [X]Recurrent depressive disorder, current episode severe with psychotic symptoms | 0 |
| Eu334 | [X]Recurrent depressive disorder, currently in remission | 0 |
| Eu33y | [X]Other recurrent depressive disorders | 0 |
| Eu33z | [X]Recurrent depressive disorder, unspecified | 0 |
| Eu341 | [X]Dysthymia | 0 |
| Eu3y1 | [X]Other recurrent mood affective disorders | 0 |
| Eu412 | [X]Mixed anxiety and depressive disorder | 0 |
| Eu530 | [X]Mild mental and behavioural disorders associated with the puerperium, not elsewhere classified | 0 |
| Eu920 | [X]Depressive conduct disorder | 0 |
| R007z | [D]Malaise and fatigue NOS | 0 |

---

#### CONCEPT: Diabetes Type 1¶

| READ\_CODE | DESCRIPTION\_SAIL | SENSITIVE\_CODE |
| --- | --- | --- |
| 66At0 | Type I diabetic dietary review | 0 |
| C1000 | Diabetes mellitus, juvenile type, with no mention of complication | 0 |
| C108. | Insulin dependent diabetes mellitus | 0 |
| C1080 | Insulin-dependent diabetes mellitus with renal complications | 0 |
| C1081 | Insulin-dependent diabetes mellitus with ophthalmic complications | 0 |
| C1082 | Insulin-dependent diabetes mellitus with neurological complications | 0 |
| C1083 | Insulin dependent diabetes mellitus with multiple complications | 0 |
| C1084 | Unstable insulin dependent diabetes mellitus | 0 |
| C1085 | Insulin dependent diabetes mellitus with ulcer | 0 |
| C1086 | Insulin dependent diabetes mellitus with gangrene | 0 |
| C1087 | Insulin dependent diabetes mellitus with retinopathy | 0 |
| C1088 | Insulin dependent diabetes mellitus - poor control | 0 |
| C1089 | Insulin dependent diabetes maturity onset | 0 |
| C108A | Insulin-dependent diabetes without complication | 0 |
| C108B | Insulin dependent diabetes mellitus with mononeuropathy | 0 |
| C108C | Insulin dependent diabetes mellitus with polyneuropathy | 0 |
| C108D | Insulin dependent diabetes mellitus with nephropathy | 0 |
| C108E | Insulin dependent diabetes mellitus with hypoglycaemic coma | 0 |
| C108F | Insulin dependent diabetes mellitus with diabetic cataract | 0 |
| C108G | Insulin dependent diabetes mellitus with peripheral angiopathy | 0 |
| C108H | Insulin dependent diabetes mellitus with arthropathy | 0 |
| C108J | Insulin dependent diabetes mellitus with neuropathic arthropathy | 0 |
| C10E. | Type 1 diabetes mellitus | 0 |
| C10E0 | Type 1 diabetes mellitus with renal complications | 0 |
| C10E1 | Type 1 diabetes mellitus with ophthalmic complications | 0 |
| C10E2 | Type 1 diabetes mellitus with neurological complications | 0 |
| C10E3 | Type 1 diabetes mellitus with multiple complications | 0 |
| C10E4 | Unstable type 1 diabetes mellitus | 0 |
| C10E5 | Type 1 diabetes mellitus with ulcer | 0 |
| C10E6 | Type 1 diabetes mellitus with gangrene | 0 |
| C10E7 | Type 1 diabetes mellitus with retinopathy | 0 |
| C10E8 | Type 1 diabetes mellitus - poor control | 0 |
| C10E9 | Type 1 diabetes mellitus maturity onset | 0 |
| C10EA | Type 1 diabetes mellitus without complication | 0 |
| C10EB | Type 1 diabetes mellitus with mononeuropathy | 0 |
| C10EC | Type 1 diabetes mellitus with polyneuropathy | 0 |
| C10ED | Type 1 diabetes mellitus with nephropathy | 0 |
| C10EE | Type 1 diabetes mellitus with hypoglycaemic coma | 0 |
| C10EF | Type 1 diabetes mellitus with diabetic cataract | 0 |
| C10EG | Type 1 diabetes mellitus with peripheral angiopathy | 0 |
| C10EH | Type 1 diabetes mellitus with arthropathy | 0 |
| C10EJ | Type 1 diabetes mellitus with neuropathic arthropathy | 0 |
| C10EK | Type 1 diabetes mellitus with persistent proteinuria | 0 |
| C10EL | Type 1 diabetes mellitus with persistent microalbuminuria | 0 |
| C10EM | Type 1 diabetes mellitus with ketoacidosis | 0 |
| C10EN | Type 1 diabetes mellitus with ketoacidotic coma | 0 |
| C10EP | Type 1 diabetes mellitus with exudative maculopathy | 0 |
| C10EQ | Type 1 diabetes mellitus with gastroparesis | 0 |
| C10P0 | Type I diabetes mellitus in remission | 0 |

---

#### CONCEPT: Diabetes Type 2¶

| READ\_CODE | DESCRIPTION\_SAIL | SENSITIVE\_CODE |
| --- | --- | --- |
| 66A3. | Diabetic on diet only | 0 |
| 66A4. | Diabetic on oral treatment | 0 |
| 66AH3 | Conversion to non-insulin injectable medication | 0 |
| 66o2. | Diabetic on non-insulin injectable medication | 0 |
| 66o5. | Diabetic on oral treatment and glucagon-like peptide 1 receptor agonist | 0 |
| C1001 | Diabetes mellitus, adult onset, with no mention of complication | 0 |
| C1074 | NIDDM with peripheral circulatory disorder | 0 |
| C109. | Non-insulin dependent diabetes mellitus | 0 |
| C1090 | Non-insulin-dependent diabetes mellitus with renal complications | 0 |
| C1091 | Non-insulin-dependent diabetes mellitus with ophthalmic complications | 0 |
| C1092 | Non-insulin-dependent diabetes mellitus with neurological complications | 0 |
| C1093 | Non-insulin-dependent diabetes mellitus with multiple complications | 0 |
| C1094 | Non-insulin dependent diabetes mellitus with ulcer | 0 |
| C1095 | Non-insulin dependent diabetes mellitus with gangrene | 0 |
| C1096 | Non-insulin-dependent diabetes mellitus with retinopathy | 0 |
| C1097 | Non-insulin dependent diabetes mellitus - poor control | 0 |
| C1099 | Non-insulin-dependent diabetes mellitus without complication | 0 |
| C109A | Non-insulin dependent diabetes mellitus with mononeuropathy | 0 |
| C109B | Non-insulin dependent diabetes mellitus with polyneuropathy | 0 |
| C109C | Non-insulin dependent diabetes mellitus with nephropathy | 0 |
| C109D | Non-insulin dependent diabetes mellitus with hypoglycaemic coma | 0 |
| C109E | Non-insulin dependent diabetes mellitus with diabetic cataract | 0 |
| C109F | Non-insulin-dependent diabetes mellitus with peripheral angiopathy | 0 |
| C109G | Non-insulin dependent diabetes mellitus with arthropathy | 0 |
| C109H | Non-insulin dependent diabetes mellitus with neuropathic arthropathy | 0 |
| C109J | Insulin treated Type 2 diabetes mellitus | 0 |
| C109K | Hyperosmolar non-ketotic state in type 2 diabetes mellitus | 0 |
| C10F. | Type 2 diabetes mellitus | 0 |
| C10F0 | Type 2 diabetes mellitus with renal complications | 0 |
| C10F1 | Type 2 diabetes mellitus with ophthalmic complications | 0 |
| C10F2 | Type 2 diabetes mellitus with neurological complications | 0 |
| C10F3 | Type 2 diabetes mellitus with multiple complications | 0 |
| C10F4 | Type 2 diabetes mellitus with ulcer | 0 |
| C10F5 | Type 2 diabetes mellitus with gangrene | 0 |
| C10F6 | Type 2 diabetes mellitus with retinopathy | 0 |
| C10F7 | Type 2 diabetes mellitus - poor control | 0 |
| C10F9 | Type 2 diabetes mellitus without complication | 0 |
| C10FA | Type 2 diabetes mellitus with mononeuropathy | 0 |
| C10FB | Type 2 diabetes mellitus with polyneuropathy | 0 |
| C10FC | Type 2 diabetes mellitus with nephropathy | 0 |
| C10FD | Type 2 diabetes mellitus with hypoglycaemic coma | 0 |
| C10FE | Type 2 diabetes mellitus with diabetic cataract | 0 |
| C10FF | Type 2 diabetes mellitus with peripheral angiopathy | 0 |
| C10FG | Type 2 diabetes mellitus with arthropathy | 0 |
| C10FH | Type 2 diabetes mellitus with neuropathic arthropathy | 0 |
| C10FJ | Insulin treated Type 2 diabetes mellitus | 0 |
| C10FK | Hyperosmolar non-ketotic state in type 2 diabetes mellitus | 0 |
| C10FL | Type 2 diabetes mellitus with persistent proteinuria | 0 |
| C10FM | Type 2 diabetes mellitus with persistent microalbuminuria | 0 |
| C10FN | Type 2 diabetes mellitus with ketoacidosis | 0 |
| C10FP | Type 2 diabetes mellitus with ketoacidotic coma | 0 |
| C10FQ | Type 2 diabetes mellitus with exudative maculopathy | 0 |
| C10FR | Type 2 diabetes mellitus with gastroparesis | 0 |
| C10P1 | Type II diabetes mellitus in remission | 0 |
| L1806 | Pre-existing diabetes mellitus, non-insulin-dependent | 0 |

---

#### CONCEPT: Diabetic retinopathy¶

| READ\_CODE | DESCRIPTION\_SAIL | SENSITIVE\_CODE |
| --- | --- | --- |
| 2BBF. | Retinal abnormality - diabetes related | 0 |
| 2BBL. | O/E - diabetic maculopathy present both eyes | 0 |
| 2BBP. | O/E - right eye background diabetic retinopathy | 0 |
| 2BBQ. | O/E - left eye background diabetic retinopathy | 0 |
| 2BBR. | O/E - right eye preproliferative diabetic retinopathy | 0 |
| 2BBS. | O/E - left eye preproliferative diabetic retinopathy | 0 |
| 2BBT. | O/E - right eye proliferative diabetic retinopathy | 0 |
| 2BBV. | O/E - left eye proliferative diabetic retinopathy | 0 |
| 2BBW. | O/E - right eye diabetic maculopathy | 0 |
| 2BBX. | O/E - left eye diabetic maculopathy | 0 |
| 2BBk. | O/E - right eye stable treated proliferative diabetic retinopathy | 0 |
| 2BBl. | O/E - left eye stable treated proliferative diabetic retinopathy | 0 |
| 2BBo. | O/E - sight threatening diabetic retinopathy | 0 |
| 2BBr. | Impaired vision due to diabetic retinopathy | 0 |
| 7276. | Pan retinal photocoagulation for diabetes | 0 |
| 8HBG. | Diabetic retinopathy 12 month review | 0 |
| 8HBH. | Diabetic retinopathy 6 month review | 0 |
| C1087 | Insulin dependent diabetes mellitus with retinopathy | 0 |
| C1096 | Non-insulin-dependent diabetes mellitus with retinopathy | 0 |
| C10E7 | Type 1 diabetes mellitus with retinopathy | 0 |
| C10EP | Type 1 diabetes mellitus with exudative maculopathy | 0 |
| C10F6 | Type 2 diabetes mellitus with retinopathy | 0 |
| C10FQ | Type 2 diabetes mellitus with exudative maculopathy | 0 |
| F420. | Diabetic retinopathy | 0 |
| F4200 | Background diabetic retinopathy | 0 |
| F4201 | Proliferative diabetic retinopathy | 0 |
| F4202 | Preproliferative diabetic retinopathy | 0 |
| F4203 | Advanced diabetic maculopathy | 0 |
| F4204 | Diabetic maculopathy | 0 |
| F4205 | Advanced diabetic retinal disease | 0 |
| F4206 | Non proliferative diabetic retinopathy | 0 |
| F4207 | High risk proliferative diabetic retinopathy | 0 |
| F4208 | High risk non proliferative diabetic retinopathy | 0 |
| F420z | Diabetic retinopathy NOS | 0 |

---

#### CONCEPT: Dialysis¶

| READ\_CODE | DESCRIPTION\_SAIL | SENSITIVE\_CODE |
| --- | --- | --- |
| 14V2. | H/O: renal dialysis | 0 |
| 4I29. | Peritoneal dialysis sample | 0 |
| 4N... | Dialysis fluid examination | 0 |
| 4N0.. | Dialysis fluid urea level | 0 |
| 4N1.. | Dialysis fluid creatinine level | 0 |
| 4N2.. | Dialysis fluid glucose level | 0 |
| 4N3.. | Peritoneal dialysis fluid cell count | 0 |
| 4N4.. | Dialysis fluid potassium level | 0 |
| 4N5.. | Dialysis fluid sodium level | 0 |
| 7A606 | Creation of graft fistula for dialysis | 0 |
| 7A619 | Ligation of arteriovenous dialysis fistula | 0 |
| 7A61A | Ligation of arteriovenous dialysis graft | 0 |
| 7L1A. | Compensation for renal failure | 0 |
| 7L1A0 | Renal dialysis | 0 |
| 7L1A1 | Peritoneal dialysis | 0 |
| 7L1A2 | Haemodialysis NEC | 0 |
| 7L1A3 | Haemofiltration | 0 |
| 7L1A4 | Automated peritoneal dialysis | 0 |
| 7L1A5 | Continuous ambulatory peritoneal dialysis | 0 |
| 7L1A6 | Peritoneal dialysis NEC | 0 |
| 7L1B. | Placement of ambulatory apparatus for compensation for renal failure | 0 |
| 7L1B0 | Insertion of ambulatory peritoneal dialysis catheter | 0 |
| 7L1B1 | Removal of ambulatory peritoneal dialysis catheter | 0 |
| 7L1B2 | Flushing of peritoneal dialysis catheter | 0 |
| 7L1C0 | Insertion of temporary peritoneal dialysis catheter | 0 |
| 7L1f0 | Extracorporeal albumin haemodialysis | 0 |
| 8882. | Intestinal dialysis | 0 |
| 8DD.. | Dependence on renal dialysis | 0 |
| G72C. | Ruptured aneurysm of dialysis vascular access | 0 |
| G72D. | Aneurysm of dialysis arteriovenous fistula | 0 |
| G72D0 | Aneurysm of superficialised artery of dialysis arteriovenous fistula | 0 |
| G72D1 | Aneurysm of needle site of dialysis arteriovenous fistula | 0 |
| G72D2 | Aneurysm of anastomotic site of dialysis arteriovenous fistula | 0 |
| G72E. | Aneurysm of dialysis vascular access | 0 |
| Gy1.. | Stenosis of dialysis vascular access | 0 |
| Gy10. | Stenosis of dialysis arteriovenous graft | 0 |
| Gy11. | Stenosis of dialysis arteriovenous shunt | 0 |
| Gy110 | Stenosis of arterial side of dialysis arteriovenous shunt | 0 |
| Gy111 | Stenosis of venous side of dialysis arteriovenous shunt | 0 |
| Gy2.. | Thrombosis of dialysis vascular access | 0 |
| Gy20. | Thrombosis of dialysis arteriovenous graft | 0 |
| Gy21. | Thrombosis of dialysis arteriovenous fistula | 0 |
| Gy22. | Thrombosis of dialysis arteriovenous shunt | 0 |
| Gy3.. | Occlusion of dialysis vascular access | 0 |
| Gy30. | Occlusion of dialysis arteriovenous graft | 0 |
| Gy31. | Occlusion of dialysis arteriovenous fistula | 0 |
| Gy32. | Occlusion of dialysis arteriovenous shunt | 0 |
| Gy4.. | Infection of dialysis vascular access | 0 |
| Gy40. | Infection of dialysis arteriovenous graft | 0 |
| Gy41. | Infection of dialysis arteriovenous fistula | 0 |
| Gy42. | Infection of dialysis arteriovenous shunt | 0 |
| Gy5.. | Haemorrhage of dialysis vascular access | 0 |
| Gy50. | Haemorrhage of dialysis arteriovenous graft | 0 |
| Gy51. | Haemorrhage of dialysis arteriovenous fistula | 0 |
| Gy52. | Haemorrhage of dialysis arteriovenous shunt | 0 |
| Gy6.. | Rupture of dialysis vascular access | 0 |
| Gy60. | Rupture of dialysis arteriovenous graft | 0 |
| Gy61. | Rupture of dialysis arteriovenous fistula | 0 |
| Gy62. | Rupture of dialysis arteriovenous shunt | 0 |
| SP015 | Mechanical complication of dialysis catheter | 0 |
| SP056 | [X]Infection and inflammatory reaction due to other internal prosthetic devices, implants and grafts | 0 |
| SP06B | Continuous ambulatory peritoneal dialysis associated peritonitis | 0 |
| SP07G | Stenosis of arteriovenous dialysis fistula | 0 |
| SP0E. | Disorders associated with peritoneal dialysis | 0 |
| SP0E0 | Bloodstained peritoneal dialysis effluent | 0 |
| SP0E1 | Thrombus in peritoneal dialysis catheter | 0 |
| SP0F. | Haemodialysis first use syndrome | 0 |
| SP0G. | Anaphylactoid reaction due to haemodialysis | 0 |
| SP0H. | Disorder associated with dialysis | 0 |
| SP0H0 | Dialysis disequilibrium | 0 |
| SP3y9 | Acute hypercalcaemia of dialysis | 0 |
| TA02. | Accidental cut, puncture, perforation or haemorrhage during kidney dialysis or other perfusion | 0 |
| TA020 | Accidental cut, puncture, perforation or haemorrhage during kidney dialysis | 0 |
| TA120 | Foreign object left in body during kidney dialysis | 0 |
| TA220 | Failure of sterile precautions during kidney dialysis | 0 |
| TA420 | Mechanical failure of instrument or apparatus during kidney dialysis | 0 |
| TB11. | Kidney dialysis as the cause of abnormal reaction of patient, or of later complication, without mention of misadventure at the time of procedure | 0 |
| U641. | [X]Kidney dialysis as the cause of abnormal reaction of the patient, or of later complication, without mention of misadventure at the time of the procedure | 0 |
| ZV451 | [V]Renal dialysis status | 0 |
| ZV56. | [V]Aftercare involving intermittent dialysis | 0 |
| ZV560 | [V]Aftercare involving extracorporeal dialysis | 0 |
| ZV561 | [V]Preparatory care for dialysis | 0 |
| ZV56y | [V]Other specified aftercare involving intermittent dialysis | 0 |
| ZV56z | [V]Unspecified aftercare involving intermittent dialysis | 0 |
| ZVu3G | [X]Other dialysis | 0 |

---

#### CONCEPT: Diverticular Disease¶

| READ\_CODE | DESCRIPTION\_SAIL | SENSITIVE\_CODE |
| --- | --- | --- |
| 76410 | Excision of Meckel diverticulum | 0 |
| 77180 | Excision of diverticulum of colon | 0 |
| J171. | Gastric diverticulum | 0 |
| J23z3 | Appendicular diverticulum | 0 |
| J51.. | Diverticula of intestine | 0 |
| J510. | Diverticulosis | 0 |
| J5100 | Diverticulosis of the duodenum | 0 |
| J5101 | Diverticulosis of the jejunum | 0 |
| J5102 | Diverticulosis of the ileum | 0 |
| J5103 | Diverticulosis of the small intestine unspecified | 0 |
| J5104 | Diverticulosis of the small intestine NOS | 0 |
| J5105 | Diverticulosis of the colon | 0 |
| J5106 | Diverticulosis of the large intestine unspecified | 0 |
| J5107 | Diverticulosis of the large intestine NOS | 0 |
| J5108 | Diverticular disease of both small and large intestine without perforation or abscess | 0 |
| J5109 | Bleeding diverticulosis | 0 |
| J510y | Diverticulosis unspecified | 0 |
| J510z | Diverticulosis NOS | 0 |
| J511. | Diverticulitis | 0 |
| J5110 | Diverticulitis of the duodenum | 0 |
| J5111 | Diverticulitis of the jejunum | 0 |
| J5112 | Diverticulitis of the ileum | 0 |
| J5113 | Diverticulitis of the small intestine unspecified | 0 |
| J5114 | Diverticulitis of the small intestine NOS | 0 |
| J5115 | Diverticulitis of the colon | 0 |
| J5116 | Diverticulitis of the large intestine unspecified | 0 |
| J5117 | Diverticulitis of the large intestine NOS | 0 |
| J511y | Diverticulitis unspecified | 0 |
| J511z | Diverticulitis NOS | 0 |
| J512. | Perforated diverticulum | 0 |
| J5120 | Perforated diverticulum of duodenum | 0 |
| J5121 | Perforated diverticulum of jejunum | 0 |
| J5122 | Perforated diverticulum of ileum | 0 |
| J5123 | Perforated diverticulum of small intestine unspecified | 0 |
| J5124 | Perforated diverticulum of small intestine NOS | 0 |
| J5125 | Perforated diverticulum of colon | 0 |
| J5126 | Perforated diverticulum of large intestine unspecified | 0 |
| J5127 | Perforated diverticulum of large intestine NOS | 0 |
| J5128 | Diverticular disease of both small and large intestine with perforation and abscess | 0 |
| J512y | Perforated diverticulum unspecified | 0 |
| J512z | Perforated diverticulum of intestine NOS | 0 |
| J513. | Diverticular abscess | 0 |
| J51z. | Diverticula of the intestine NOS | 0 |

---

#### CONCEPT: Drug or alcohol misuse¶

| READ\_CODE | DESCRIPTION\_SAIL | SENSITIVE\_CODE |
| --- | --- | --- |
| 136S. | Hazardous alcohol use | 0 |
| 136T. | Harmful alcohol use | 0 |
| 13Y8. | Alcoholics anonymous | 0 |
| 13ZY. | Disqualified from driving due to excess alcohol | 0 |
| 13c.. | Drug user | 0 |
| 13c0. | Injecting drug user | 0 |
| 13c1. | Intravenous drug user | 0 |
| 13c2. | Never injecting drug user | 0 |
| 13c3. | Intramuscular drug user | 0 |
| 13c4. | Intranasal drug user | 0 |
| 13c5. | Substance misuse increased | 0 |
| 13c6. | Substance misuse decreased | 0 |
| 13c7. | Current drug user | 0 |
| 13c8. | Reduced drugs misuse | 0 |
| 13c9. | Subcutaneous drug user | 0 |
| 13cA. | Smokes drugs | 0 |
| 13cB. | Misuses drugs orally | 0 |
| 13cC. | Continuous use of drugs | 0 |
| 13cD. | Episodic use of drugs | 0 |
| 13cE. | Prolonged high dose use of cannabis | 0 |
| 13cF. | Preoccupied with substance misuse | 0 |
| 13cH. | Persistent substance misuse | 0 |
| 13cJ. | Previously injecting drug user | 0 |
| 13cK. | Current non recreational drug user | 0 |
| 13cM. | Substance misuse | 0 |
| 13cM0 | Novel psychoactive substance misuse | 0 |
| 13cM1 | Opioid analgesic dependence | 0 |
| 13cN. | Has never shared drug injection equipment | 0 |
| 13cQ. | Behavioural tolerance to drug | 0 |
| 13cR. | Physical tolerance to drug | 0 |
| 13cS. | Psychological tolerance to drug | 0 |
| 13cT. | Reverse tolerance to drug | 0 |
| 13r0. | Abstinent from drug misuse | 0 |
| 13r1. | Abstinent from drug misuse in normal environment | 0 |
| 13r2. | Abstinent from drug misuse in protected environment | 0 |
| 13r3. | Abstinent from drug misuse on maintenance replacement | 0 |
| 13r4. | Abstinent from drug misuse when receiving blocking therapy | 0 |
| 13r5. | Abstinent from substance misuse | 0 |
| 1462. | H/O: alcoholism | 0 |
| 1463. | H/O: drug dependency | 0 |
| 146E. | H/O: recreational drug use | 0 |
| 146F. | H/O: drug abuse | 0 |
| 1B1c. | Alcohol induced hallucinations | 0 |
| 1P30. | Compulsive uncontrollable drug taking | 0 |
| 1P31. | Compulsive drug taking | 0 |
| 1P6.. | Craving for drugs | 0 |
| 1P60. | Craves for drugs | 0 |
| 1P62. | Abnormal craving for drugs | 0 |
| 1P63. | Excessive craving for drugs | 0 |
| 1P64. | Irresistible craving for drugs | 0 |
| 1T... | History of substance misuse | 0 |
| 1T0.. | H/O heroin misuse | 0 |
| 1T00. | H/O daily heroin misuse | 0 |
| 1T01. | H/O weekly heroin misuse | 0 |
| 1T02. | Previous history of heroin misuse | 0 |
| 1T03. | H/O infrequent heroin misuse | 0 |
| 1T1.. | H/O methadone misuse | 0 |
| 1T10. | H/O daily methadone misuse | 0 |
| 1T11. | H/O weekly methadone misuse | 0 |
| 1T12. | H/O infrequent methadone misuse | 0 |
| 1T13. | Previous history of methadone misuse | 0 |
| 1T2.. | H/O ecstasy misuse | 0 |
| 1T20. | H/O daily ecstasy misuse | 0 |
| 1T21. | H/O weekly ecstasy misuse | 0 |
| 1T22. | H/O infrequent ecstasy misuse | 0 |
| 1T23. | Previous history of ecstasy misuse | 0 |
| 1T3.. | H/O benzodiazepine misuse | 0 |
| 1T30. | H/O daily benzodiazepine misuse | 0 |
| 1T31. | H/O weekly benzodiazepine misuse | 0 |
| 1T32. | H/O infrequent benzodiazepine misuse | 0 |
| 1T33. | Previous history of benzodiazepine misuse | 0 |
| 1T4.. | H/O amphetamine misuse | 0 |
| 1T40. | H/O daily amphetamine misuse | 0 |
| 1T41. | H/O weekly amphetamine misuse | 0 |
| 1T42. | H/O infrequent amphetamine misuse | 0 |
| 1T43. | Previous history of amphetamine misuse | 0 |
| 1T5.. | H/O cocaine misuse | 0 |
| 1T50. | H/O daily cocaine misuse | 0 |
| 1T51. | H/O weekly cocaine misuse | 0 |
| 1T52. | H/O infrequent cocaine misuse | 0 |
| 1T53. | Previous history of cocaine misuse | 0 |
| 1T6.. | H/O crack cocaine misuse | 0 |
| 1T60. | H/O daily crack cocaine misuse | 0 |
| 1T61. | H/O weekly crack cocaine misuse | 0 |
| 1T62. | H/O infrequent crack cocaine misuse | 0 |
| 1T63. | Previous history of crack cocaine misuse | 0 |
| 1T7.. | H/O hallucinogen misuse | 0 |
| 1T70. | H/O daily hallucinogen misuse | 0 |
| 1T71. | H/O weekly hallucinogen misuse | 0 |
| 1T72. | H/O infrequent hallucinogen misuse | 0 |
| 1T73. | Previous history of hallucinogen misuse | 0 |
| 1T8.. | H/O cannabis misuse | 0 |
| 1T80. | H/O daily cannabis misuse | 0 |
| 1T81. | H/O weekly cannabis misuse | 0 |
| 1T82. | H/O infrequent cannabis misuse | 0 |
| 1T83. | Previous history of cannabis misuse | 0 |
| 1T9.. | H/O solvent misuse | 0 |
| 1T90. | H/O daily solvent misuse | 0 |
| 1T91. | H/O weekly solvent misuse | 0 |
| 1T92. | H/O infrequent solvent misuse | 0 |
| 1T93. | Previous history of solvent misuse | 0 |
| 1TA.. | H/O barbiturate misuse | 0 |
| 1TA0. | H/O daily barbiturate misuse | 0 |
| 1TA1. | H/O weekly barbiturate misuse | 0 |
| 1TA2. | H/O infrequent barbiturate misuse | 0 |
| 1TA3. | Previous history of barbiturate misuse | 0 |
| 1TB.. | H/O major tranquilliser misuse | 0 |
| 1TB0. | H/O daily major tranquilliser misuse | 0 |
| 1TB1. | H/O weekly major tranquilliser misuse | 0 |
| 1TB2. | H/O infrequent major tranquilliser misuse | 0 |
| 1TB3. | Previous history of major tranquilliser misuse | 0 |
| 1TC.. | H/O anti-depressant misuse | 0 |
| 1TC0. | H/O daily anti-depressant misuse | 0 |
| 1TC1. | H/O weekly anti-depressant misuse | 0 |
| 1TC2. | H/O infrequent anti-depressant misuse | 0 |
| 1TC3. | Previous history of anti-depressant misuse | 0 |
| 1TD.. | H/O opiate misuse | 0 |
| 1TD0. | H/O daily opiate misuse | 0 |
| 1TD1. | H/O weekly opiate misuse | 0 |
| 1TD2. | H/O infrequent opiate misuse | 0 |
| 1TD3. | Previous history of opiate misuse | 0 |
| 1TE.. | Uses heroin on top of substitution therapy | 0 |
| 1TF.. | Does not use heroin on top of substitution therapy | 0 |
| 1TG.. | H/O novel psychoactive substance misuse | 0 |
| 1V... | Drug misuse behaviour | 0 |
| 1V0.. | Misuses drugs | 0 |
| 1V00. | Occasional drug user | 0 |
| 1V01. | Long-term drug misuser | 0 |
| 1V02. | Poly-drug misuser | 0 |
| 1V03. | Misuses drugs sublingually | 0 |
| 1V04. | Misuses drugs rectally | 0 |
| 1V05. | Misuses drugs vaginally | 0 |
| 1V0E. | Health problem secondary to drug misuse | 0 |
| 1V2.. | Frequency of drug misuse | 0 |
| 1V22. | Age at starting drug misuse | 0 |
| 1V23. | Time since stopped drug misuse | 0 |
| 1V24. | Total time drugs misused | 0 |
| 1V26. | Misused drugs in past | 0 |
| 1V64. | Illicit drug use | 0 |
| 1V65. | Heroin misuse | 0 |
| 1V66. | Ecstasy misuse | 0 |
| 2126C | Alcohol dependence resolved | 0 |
| 66e.. | Alcohol disorder monitoring | 0 |
| 66e0. | Alcohol abuse monitoring | 0 |
| 67H3. | Lifestyle advice regarding drug misuse | 0 |
| 68U.. | Drugs of abuse screening | 0 |
| 68U0. | Drugs of abuse urine screening | 0 |
| 7P221 | Delivery of rehabilitation for alcohol addiction | 0 |
| 8AA.. | Drug abuse monitoring | 0 |
| 8B23. | Drug addiction therapy | 0 |
| 8B2N. | Drug addiction detoxification therapy - methadone | 0 |
| 8B2P. | Drug addiction maintenance therapy - methadone | 0 |
| 8B2Q. | Drug addiction maintenance therapy - buprenorphine | 0 |
| 8B2R. | Drug addiction detoxification therapy - buprenorphine | 0 |
| 8B2S. | Opioid agonist substitution therapy | 0 |
| 8B2T. | Opioid antagonist therapy | 0 |
| 8BA8. | Alcohol detoxification | 0 |
| 8BA9. | Detoxification dependence drug | 0 |
| 8BAW. | Drug dependence self detoxification | 0 |
| 8BAX. | Drug dependence home detoxification | 0 |
| 8BAc. | Substance misuse management stopped - self withdrawal | 0 |
| 8BAd. | Opiate dependence detoxification | 0 |
| 8BAo. | Benzodiazepine dependence detoxification | 0 |
| 8BE0. | Reinduction to methadone maintenance therapy | 0 |
| 8BE1. | Reinduction to buprenorphine maintenance therapy | 0 |
| 8FB.. | Drug rehabilitation | 0 |
| 8FB0. | Drug detoxification programme completed | 0 |
| 8G22. | Family intervention for substance misuse | 0 |
| 8G32. | Aversion therapy - alcoholism | 0 |
| 8H35. | Admitted to alcohol detoxification centre | 0 |
| 8H7p. | Referral to community alcohol team | 0 |
| 8HkG. | Referral to specialist alcohol treatment service | 0 |
| 8HkJ. | Referral to alcohol brief intervention service | 0 |
| 8Hq.. | Admission to substance misuse detoxification centre | 0 |
| 8I2N. | Drug dependence home detoxification contraindicated | 0 |
| 8IAF. | Brief intervention for excessive alcohol consumption declined | 0 |
| 8IAt. | Extended intervention for excessive alcohol consumption declined | 0 |
| 8T24. | Referral for brief intervention for substance misuse | 0 |
| 8T25. | Referral for specialist substance misuse intervention | 0 |
| 9HC.. | Substance misuse monitoring | 0 |
| 9HC0. | Initial substance misuse assessment | 0 |
| 9HC1. | Follow up substance misuse assessment | 0 |
| 9HC2. | Substance misuse clinical management plan agreed | 0 |
| 9HC3. | Substance misuse clinical management plan reviewed | 0 |
| 9HC4. | Substance misuse treatment withdrawn | 0 |
| 9HC5. | Substance misuse treatment programme completed | 0 |
| 9HC6. | Substance misuse treatment declined | 0 |
| 9HC7. | Substance misuse treatment not available | 0 |
| 9HC8. | Declined to give substance misuse history | 0 |
| 9HCC. | On substance misuse programme | 0 |
| 9N1yJ | Seen in drug misuse clinic | 0 |
| 9NN2. | Under care of community alcohol team | 0 |
| 9NX2. | In-house substance misuse treatment | 0 |
| 9NdN. | Declined consent for notification of drug misuse | 0 |
| 9No5. | Seen in substance misuse clinic | 0 |
| 9k1A. | Brief intervention for excessive alcohol consumption completed | 0 |
| 9k1B. | Extended intervention for excessive alcohol consumption completed | 0 |
| 9k50. | Drug misuse - enhanced service completed | 0 |
| 9k51. | Shared care drug misuse treatment - enhanced services administration | 0 |
| 9k52. | Drug misuse treatment in primary care - enhanced services administration | 0 |
| 9k53. | Pharmacy attended for drug misuse - enhanced services administration | 0 |
| C1505 | Alcohol-induced pseudo-Cushing's syndrome | 0 |
| E01.. | Alcoholic psychoses | 0 |
| E010. | Alcohol withdrawal delirium | 0 |
| E011. | Alcohol amnestic syndrome | 0 |
| E0110 | Korsakov's alcoholic psychosis | 0 |
| E0111 | Korsakov's alcoholic psychosis with peripheral neuritis | 0 |
| E0112 | Wernicke-Korsakov syndrome | 0 |
| E011z | Alcohol amnestic syndrome NOS | 0 |
| E012. | Other alcoholic dementia | 0 |
| E0120 | Chronic alcoholic brain syndrome | 0 |
| E013. | Alcohol withdrawal hallucinosis | 0 |
| E014. | Pathological alcohol intoxication | 0 |
| E015. | Alcoholic paranoia | 0 |
| E01y. | Other alcoholic psychosis | 0 |
| E01y0 | Alcohol withdrawal syndrome | 0 |
| E01yz | Other alcoholic psychosis NOS | 0 |
| E01z. | Alcoholic psychosis NOS | 0 |
| E02.. | Drug psychoses | 0 |
| E020. | Drug withdrawal syndrome | 0 |
| E021. | Drug-induced paranoia or hallucinatory states | 0 |
| E0210 | Drug-induced paranoid state | 0 |
| E0211 | Drug-induced hallucinosis | 0 |
| E021z | Drug-induced paranoia or hallucinatory state NOS | 0 |
| E022. | Pathological drug intoxication | 0 |
| E02y. | Other drug psychoses | 0 |
| E02y0 | Drug-induced delirium | 0 |
| E02y1 | Drug-induced dementia | 0 |
| E02y2 | Drug-induced amnestic syndrome | 0 |
| E02y3 | Drug-induced depressive state | 0 |
| E02y4 | Drug-induced personality disorder | 0 |
| E02yz | Other drug psychoses NOS | 0 |
| E02z. | Drug psychosis NOS | 0 |
| E23.. | Alcohol dependence syndrome | 0 |
| E230. | Acute alcoholic intoxication in alcoholism | 0 |
| E2300 | Acute alcoholic intoxication, unspecified, in alcoholism | 0 |
| E2301 | Continuous acute alcoholic intoxication in alcoholism | 0 |
| E2302 | Episodic acute alcoholic intoxication in alcoholism | 0 |
| E2303 | Acute alcoholic intoxication in remission, in alcoholism | 0 |
| E230z | Acute alcoholic intoxication in alcoholism NOS | 0 |
| E231. | Chronic alcoholism | 0 |
| E2310 | Unspecified chronic alcoholism | 0 |
| E2311 | Continuous chronic alcoholism | 0 |
| E2312 | Episodic chronic alcoholism | 0 |
| E2313 | Chronic alcoholism in remission | 0 |
| E231z | Chronic alcoholism NOS | 0 |
| E23z. | Alcohol dependence syndrome NOS | 0 |
| E24.. | Drug dependence | 0 |
| E240. | Opioid type drug dependence | 0 |
| E2400 | Unspecified opioid dependence | 0 |
| E2401 | Continuous opioid dependence | 0 |
| E2402 | Episodic opioid dependence | 0 |
| E2403 | Opioid dependence in remission | 0 |
| E240z | Opioid drug dependence NOS | 0 |
| E241. | Hypnotic or anxiolytic dependence | 0 |
| E2410 | Hypnotic or anxiolytic dependence, unspecified | 0 |
| E2411 | Hypnotic or anxiolytic dependence, continuous | 0 |
| E2412 | Hypnotic or anxiolytic dependence, episodic | 0 |
| E2413 | Hypnotic or anxiolytic dependence in remission | 0 |
| E241z | Hypnotic or anxiolytic dependence NOS | 0 |
| E242. | Cocaine type drug dependence | 0 |
| E2420 | Cocaine dependence, unspecified | 0 |
| E2421 | Cocaine dependence, continuous | 0 |
| E2422 | Cocaine dependence, episodic | 0 |
| E2423 | Cocaine dependence in remission | 0 |
| E242z | Cocaine drug dependence NOS | 0 |
| E243. | Cannabis type drug dependence | 0 |
| E2430 | Cannabis dependence, unspecified | 0 |
| E2431 | Cannabis dependence, continuous | 0 |
| E2432 | Cannabis dependence, episodic | 0 |
| E2433 | Cannabis dependence in remission | 0 |
| E243z | Cannabis drug dependence NOS | 0 |
| E244. | Amphetamine or other psychostimulant dependence | 0 |
| E2440 | Amphetamine or psychostimulant dependence, unspecified | 0 |
| E2441 | Amphetamine or psychostimulant dependence, continuous | 0 |
| E2442 | Amphetamine or psychostimulant dependence, episodic | 0 |
| E2443 | Amphetamine or psychostimulant dependence in remission | 0 |
| E244z | Amphetamine or psychostimulant dependence NOS | 0 |
| E245. | Hallucinogen dependence | 0 |
| E2450 | Hallucinogen dependence, unspecified | 0 |
| E2451 | Hallucinogen dependence, continuous | 0 |
| E2452 | Hallucinogen dependence, episodic | 0 |
| E2453 | Hallucinogen dependence in remission | 0 |
| E245z | Hallucinogen dependence NOS | 0 |
| E246. | Glue sniffing dependence | 0 |
| E2460 | Glue sniffing dependence, unspecified | 0 |
| E2461 | Glue sniffing dependence, continuous | 0 |
| E2462 | Glue sniffing dependence, episodic | 0 |
| E2463 | Glue sniffing dependence in remission | 0 |
| E246z | Glue sniffing dependence NOS | 0 |
| E247. | Other specified drug dependence | 0 |
| E2470 | Other specified drug dependence, unspecified | 0 |
| E2471 | Other specified drug dependence, continuous | 0 |
| E2472 | Other specified drug dependence, episodic | 0 |
| E2473 | Other specified drug dependence in remission | 0 |
| E247z | Other specified drug dependence NOS | 0 |
| E248. | Combined opioid with other drug dependence | 0 |
| E2480 | Combined opioid with other drug dependence, unspecified | 0 |
| E2481 | Combined opioid with other drug dependence, continuous | 0 |
| E2482 | Combined opioid with other drug dependence, episodic | 0 |
| E2483 | Combined opioid with other drug dependence in remission | 0 |
| E248z | Combined opioid with other drug dependence NOS | 0 |
| E249. | Combined drug dependence, excluding opioids | 0 |
| E2490 | Combined drug dependence, excluding opioid, unspecified | 0 |
| E2491 | Combined drug dependence, excluding opioid, continuous | 0 |
| E2492 | Combined drug dependence, excluding opioid, episodic | 0 |
| E2493 | Combined drug dependence, excluding opioid, in remission | 0 |
| E249z | Combined drug dependence, excluding opioid, NOS | 0 |
| E24A. | Ecstasy type drug dependence | 0 |
| E24z. | Drug dependence NOS | 0 |
| E25.. | Nondependent abuse of drugs | 0 |
| E250. | Nondependent alcohol abuse | 0 |
| E2500 | Nondependent alcohol abuse, unspecified | 0 |
| E2501 | Nondependent alcohol abuse, continuous | 0 |
| E2502 | Nondependent alcohol abuse, episodic | 0 |
| E2503 | Nondependent alcohol abuse in remission | 0 |
| E250z | Nondependent alcohol abuse NOS | 0 |
| E252. | Nondependent cannabis abuse | 0 |
| E2520 | Nondependent cannabis abuse, unspecified | 0 |
| E2521 | Nondependent cannabis abuse, continuous | 0 |
| E2522 | Nondependent cannabis abuse, episodic | 0 |
| E2523 | Nondependent cannabis abuse in remission | 0 |
| E252z | Nondependent cannabis abuse NOS | 0 |
| E253. | Nondependent hallucinogen abuse | 0 |
| E2530 | Nondependent hallucinogen abuse, unspecified | 0 |
| E2531 | Nondependent hallucinogen abuse, continuous | 0 |
| E2532 | Nondependent hallucinogen abuse, episodic | 0 |
| E2533 | Nondependent hallucinogen abuse in remission | 0 |
| E253z | Nondependent hallucinogen abuse NOS | 0 |
| E254. | Nondependent hypnotic or anxiolytic abuse | 0 |
| E2540 | Nondependent hypnotic or anxiolytic abuse, unspecified | 0 |
| E2541 | Nondependent hypnotic or anxiolytic abuse, continuous | 0 |
| E2542 | Nondependent hypnotic or anxiolytic abuse, episodic | 0 |
| E2543 | Nondependent hypnotic or anxiolytic abuse in remission | 0 |
| E254z | Nondependent hypnotic or anxiolytic abuse NOS | 0 |
| E255. | Nondependent opioid abuse | 0 |
| E2550 | Nondependent opioid abuse, unspecified | 0 |
| E2551 | Nondependent opioid abuse, continuous | 0 |
| E2552 | Nondependent opioid abuse, episodic | 0 |
| E2553 | Nondependent opioid abuse in remission | 0 |
| E255z | Nondependent opioid abuse NOS | 0 |
| E256. | Nondependent cocaine abuse | 0 |
| E2560 | Nondependent cocaine abuse, unspecified | 0 |
| E2561 | Nondependent cocaine abuse, continuous | 0 |
| E2562 | Nondependent cocaine abuse, episodic | 0 |
| E2563 | Nondependent cocaine abuse in remission | 0 |
| E256z | Nondependent cocaine abuse NOS | 0 |
| E257. | Nondependent amphetamine or other psychostimulant abuse | 0 |
| E2570 | Nondependent amphetamine or psychostimulant abuse, unspecified | 0 |
| E2571 | Nondependent amphetamine or psychostimulant abuse, continuous | 0 |
| E2572 | Nondependent amphetamine or psychostimulant abuse, episodic | 0 |
| E2573 | Nondependent amphetamine or psychostimulant abuse in remission | 0 |
| E257z | Nondependent amphetamine or psychostimulant abuse NOS | 0 |
| E258. | Nondependent antidepressant type drug abuse | 0 |
| E2580 | Nondependent antidepressant type drug abuse, unspecified | 0 |
| E2581 | Nondependent antidepressant type drug abuse, continuous | 0 |
| E2582 | Nondependent antidepressant type drug abuse, episodic | 0 |
| E2583 | Nondependent antidepressant type drug abuse in remission | 0 |
| E258z | Nondependent antidepressant type drug abuse NOS | 0 |
| E259. | Nondependent mixed drug abuse | 0 |
| E2590 | Nondependent mixed drug abuse, unspecified | 0 |
| E2591 | Nondependent mixed drug abuse, continuous | 0 |
| E2592 | Nondependent mixed drug abuse, episodic | 0 |
| E2593 | Nondependent mixed drug abuse in remission | 0 |
| E2594 | Misuse of prescription only drugs | 0 |
| E259z | Nondependent mixed drug abuse NOS | 0 |
| E25y. | Nondependent other drug abuse | 0 |
| E25y0 | Nondependent other drug abuse, unspecified | 0 |
| E25y1 | Nondependent other drug abuse, continuous | 0 |
| E25y2 | Nondependent other drug abuse, episodic | 0 |
| E25y3 | Nondependent other drug abuse in remission | 0 |
| E25yz | Nondependent other drug abuse NOS | 0 |
| E25z. | Misuse of drugs NOS | 0 |
| Eu04. | [X]Delirium, not induced by alcohol and other psychoactive subs | 0 |
| Eu1.. | [X]Mental and behavioural disorders due to psychoactive substance use | 0 |
| Eu10. | [X]Mental and behavioural disorders due to use of alcohol | 0 |
| Eu100 | [X]Mental and behavioural disorders due to use of alcohol: acute intoxication | 0 |
| Eu101 | [X]Mental and behavioural disorders due to use of alcohol: harmful use | 0 |
| Eu102 | [X]Mental and behavioural disorders due to use of alcohol: dependence syndrome | 0 |
| Eu103 | [X]Mental and behavioural disorders due to use of alcohol: withdrawal state | 0 |
| Eu104 | [X]Mental and behavioural disorders due to use of alcohol: withdrawal state with delirium | 0 |
| Eu105 | [X]Mental and behavioural disorders due to use of alcohol: psychotic disorder | 0 |
| Eu106 | [X]Mental and behavioural disorders due to use of alcohol: amnesic syndrome | 0 |
| Eu107 | [X]Mental and behavioural disorders due to use of alcohol: residual and late-onset psychotic disorder | 0 |
| Eu108 | [X]Alcohol withdrawal-induced seizure | 0 |
| Eu10y | [X]Mental and behavioural disorders due to use of alcohol: other mental and behavioural disorders | 0 |
| Eu10z | [X]Mental and behavioural disorders due to use of alcohol: unspecified mental and behavioural disorder | 0 |
| Eu11. | [X]Mental and behavioural disorders due to use of opioids | 0 |
| Eu110 | [X]Mental and behavioural disorders due to use of opioids: acute intoxication | 0 |
| Eu111 | [X]Mental and behavioural disorders due to use of opioids: harmful use | 0 |
| Eu112 | [X]Mental and behavioural disorders due to use of opioids: dependence syndrome | 0 |
| Eu113 | [X]Mental and behavioural disorders due to use of opioids: withdrawal state | 0 |
| Eu114 | [X]Mental and behavioural disorders due to use of opioids: withdrawal state with delirium | 0 |
| Eu115 | [X]Mental and behavioural disorders due to use of opioids: psychotic disorder | 0 |
| Eu116 | [X]Mental and behavioural disorders due to use of opioids: amnesic syndrome | 0 |
| Eu117 | [X]Mental and behavioural disorders due to use of opioids: residual and late-onset psychotic disorder | 0 |
| Eu11y | [X]Mental and behavioural disorders due to use of opioids: other mental and behavioural disorders | 0 |
| Eu11z | [X]Mental and behavioural disorders due to use of opioids: unspecified mental and behavioural disorder | 0 |
| Eu12. | [X]Mental and behavioural disorders due to use of cannabinoids | 0 |
| Eu120 | [X]Mental and behavioural disorders due to use of cannabinoids: acute intoxication | 0 |
| Eu121 | [X]Mental and behavioural disorders due to use of cannabinoids: harmful use | 0 |
| Eu122 | [X]Mental and behavioural disorders due to use of cannabinoids: dependence syndrome | 0 |
| Eu123 | [X]Mental and behavioural disorders due to use of cannabinoids: withdrawal state | 0 |
| Eu124 | [X]Mental and behavioural disorders due to use of cannabinoids: withdrawal state with delirium | 0 |
| Eu125 | [X]Mental and behavioural disorders due to use of cannabinoids: psychotic disorder | 0 |
| Eu126 | [X]Mental and behavioural disorders due to use of cannabinoids: amnesic syndrome | 0 |
| Eu127 | [X]Mental and behavioural disorders due to use of cannabinoids: residual and late-onset psychotic disorder | 0 |
| Eu12y | [X]Mental and behavioural disorders due to use of cannabinoids: other mental and behavioural disorders | 0 |
| Eu12z | [X]Mental and behavioural disorders due to use of cannabinoids: unspecified mental and behavioural disorder | 0 |
| Eu13. | [X]Mental and behavioural disorders due to use of sedatives or hypnotics | 0 |
| Eu130 | [X]Mental and behavioural disorders due to use of sedatives or hypnotics: acute intoxication | 0 |
| Eu131 | [X]Mental and behavioural disorders due to use of sedatives or hypnotics: harmful use | 0 |
| Eu132 | [X]Mental and behavioural disorders due to use of sedatives or hypnotics: dependence syndrome | 0 |
| Eu133 | [X]Mental and behavioural disorders due to use of sedatives or hypnotics: withdrawal state | 0 |
| Eu134 | [X]Mental and behavioural disorders due to use of sedatives or hypnotics: withdrawal state with delirium | 0 |
| Eu135 | [X]Mental and behavioural disorders due to use of sedatives or hypnotics: psychotic disorder | 0 |
| Eu136 | [X]Mental and behavioural disorders due to use of sedatives or hypnotics: amnesic syndrome | 0 |
| Eu137 | [X]Mental and behavioural disorders due to use of sedatives or hypnotics: residual and late-onset psychotic disorder | 0 |
| Eu13y | [X]Mental and behavioural disorders due to use of sedatives or hypnotics: other mental and behavioural disorders | 0 |
| Eu13z | [X]Mental and behavioural disorders due to use of sedatives or hypnotics: unspecified mental and behavioural disorder | 0 |
| Eu14. | [X]Mental and behavioural disorders due to use of cocaine | 0 |
| Eu140 | [X]Mental and behavioural disorders due to use of cocaine: acute intoxication | 0 |
| Eu141 | [X]Mental and behavioural disorders due to use of cocaine: harmful use | 0 |
| Eu142 | [X]Mental and behavioural disorders due to use of cocaine: dependence syndrome | 0 |
| Eu143 | [X]Mental and behavioural disorders due to use of cocaine: withdrawal state | 0 |
| Eu144 | [X]Mental and behavioural disorders due to use of cocaine: withdrawal state with delirium | 0 |
| Eu145 | [X]Mental and behavioural disorders due to use of cocaine: psychotic disorder | 0 |
| Eu146 | [X]Mental and behavioural disorders due to use of cocaine: amnesic syndrome | 0 |
| Eu147 | [X]Mental and behavioural disorders due to use of cocaine: residual and late-onset psychotic disorder | 0 |
| Eu14y | [X]Mental and behavioural disorders due to use of cocaine: other mental and behavioural disorders | 0 |
| Eu14z | [X]Mental and behavioural disorders due to use of cocaine: unspecified mental and behavioural disorder | 0 |
| Eu16. | [X]Mental and behavioural disorders due to use of hallucinogens | 0 |
| Eu160 | [X]Mental and behavioural disorders due to use of hallucinogens: acute intoxication | 0 |
| Eu161 | [X]Mental and behavioural disorders due to use of hallucinogens: harmful use | 0 |
| Eu162 | [X]Mental and behavioural disorders due to use of hallucinogens: dependence syndrome | 0 |
| Eu163 | [X]Mental and behavioural disorders due to use of hallucinogens: withdrawal state | 0 |
| Eu164 | [X]Mental and behavioural disorders due to use of hallucinogens: withdrawal state with delirium | 0 |
| Eu165 | [X]Mental and behavioural disorders due to use of hallucinogens: psychotic disorder | 0 |
| Eu166 | [X]Mental and behavioural disorders due to use of hallucinogens: amnesic syndrome | 0 |
| Eu167 | [X]Mental and behavioural disorders due to use of hallucinogens: residual and late-onset psychotic disorder | 0 |
| Eu16y | [X]Mental and behavioural disorders due to use of hallucinogens: other mental and behavioural disorders | 0 |
| Eu16z | [X]Mental and behavioural disorders due to use of hallucinogens: unspecified mental and behavioural disorder | 0 |
| Eu18. | [X]Mental and behavioural disorders due to use of volatile solvents | 0 |
| Eu180 | [X]Mental and behavioural disorders due to use of volatile solvents: acute intoxication | 0 |
| Eu181 | [X]Mental and behavioural disorders due to use of volatile solvents: harmful use | 0 |
| Eu182 | [X]Mental and behavioural disorders due to use of volatile solvents: dependence syndrome | 0 |
| Eu183 | [X]Mental and behavioural disorders due to use of volatile solvents: withdrawal state | 0 |
| Eu184 | [X]Mental and behavioural disorders due to use of volatile solvents: withdrawal state with delirium | 0 |
| Eu185 | [X]Mental and behavioural disorders due to use of volatile solvents: psychotic disorder | 0 |
| Eu186 | [X]Mental and behavioural disorders due to use of volatile solvents: amnesic syndrome | 0 |
| Eu187 | [X]Mental and behavioural disorders due to use of volatile solvents: residual and late-onset psychotic disorder | 0 |
| Eu18y | [X]Mental and behavioural disorders due to use of volatile solvents: other mental and behavioural disorders | 0 |
| Eu18z | [X]Mental and behavioural disorders due to use of volatile solvents: unspecified mental and behavioural disorder | 0 |
| Eu19. | [X]Mental and behavioural disorders due to multiple drug use and use of other psychoactive substances | 0 |
| Eu190 | [X]Mental and behavioural disorders due to multiple drug use and use of other psychoactive substances: acute intoxication | 0 |
| Eu191 | [X]Mental and behavioural disorders due to multiple drug use and use of other psychoactive substances: harmful use | 0 |
| Eu192 | [X]Mental and behavioural disorders due to multiple drug use and use of other psychoactive substances: dependence syndrome | 0 |
| Eu193 | [X]Mental and behavioural disorders due to multiple drug use and use of other psychoactive substances: withdrawal state | 0 |
| Eu194 | [X]Mental and behavioural disorders due to multiple drug use and use of other psychoactive substances: withdrawal state with delirium | 0 |
| Eu195 | [X]Mental and behavioural disorders due to multiple drug use and use of other psychoactive substances: psychotic disorder | 0 |
| Eu196 | [X]Mental and behavioural disorders due to multiple drug use and use of other psychoactive substances: amnesic syndrome | 0 |
| Eu197 | [X]Mental and behavioural disorders due to multiple drug use and use of other psychoactive substances: residual and late-onset psychotic | 0 |
| Eu19y | [X]Mental and behavioural disorders due to multiple drug use and use of other psychoactive substances: other mental and behavioural disorders | 0 |
| Eu19z | [X]Mental and behavioural disorders due to multiple drug use and use of other psychoactive substances: unspecified mental and behavioural disorder | 0 |
| Eu1A. | [X]Mental and behavioural disorders due to use of crack cocaine | 0 |
| Eu1A0 | [X]Mental and behavioural disorders due to use of crack cocaine: acute intoxication | 0 |
| Eu1A1 | [X]Mental and behavioural disorders due to use of crack cocaine: harmful use | 0 |
| Eu1A2 | [X]Mental and behavioural disorders due to use of crack cocaine: dependence syndrome | 0 |
| Eu1A3 | [X]Mental and behavioural disorders due to use of crack cocaine: withdrawal state | 0 |
| Eu1A4 | [X]Mental and behavioural disorders due to use of crack cocaine: withdrawal state with delirium | 0 |
| Eu1A5 | [X]Mental and behavioural disorders due to use of crack cocaine: psychotic disorder | 0 |
| Eu1A6 | [X]Mental and behavioural disorders due to use of crack cocaine: amnesic syndrome | 0 |
| Eu1A7 | [X]Mental and behavioural disorders due to use of crack cocaine: residual and late-onset psychotic disorder | 0 |
| Eu1Ay | [X]Mental and behavioural disorders due to use of crack cocaine: other mental and behavioural disorders | 0 |
| Eu1Az | [X]Mental and behavioural disorders due to use of crack cocaine: unspecified mental and behavioural disorder | 0 |
| Eu55. | [X]Abuse of non-dependence-producing substances | 0 |
| F11x0 | Cerebral degeneration due to alcoholism | 0 |
| F1440 | Cerebellar ataxia due to alcoholism | 0 |
| F25B. | Alcohol-induced epilepsy | 0 |
| F375. | Alcoholic polyneuropathy | 0 |
| F3941 | Alcoholic myopathy | 0 |
| G555. | Alcoholic cardiomyopathy | 0 |
| G801E | Deep vein thrombosis of leg related to intravenous drug use | 0 |
| G8523 | Oesophageal varices in alcoholic cirrhosis of the liver | 0 |
| J153. | Alcoholic gastritis | 0 |
| J610. | Alcoholic fatty liver | 0 |
| J611. | Acute alcoholic hepatitis | 0 |
| J612. | Alcoholic cirrhosis of liver | 0 |
| J6120 | Alcoholic fibrosis and sclerosis of liver | 0 |
| J613. | Alcoholic liver damage unspecified | 0 |
| J6130 | Alcoholic hepatic failure | 0 |
| J615. | Cirrhosis - non alcoholic | 0 |
| J617. | Alcoholic hepatitis | 0 |
| J6170 | Chronic alcoholic hepatitis | 0 |
| J6708 | Alcohol-induced acute pancreatitis | 0 |
| J6710 | Alcohol-induced chronic pancreatitis | 0 |
| L183. | Drug dependence during pregnancy, childbirth and the puerperium | 0 |
| L1830 | Drug dependence - unspecified whether during pregnancy or the puerperium | 0 |
| L1831 | Drug dependence during pregnancy - baby delivered | 0 |
| L1832 | Drug dependence in the puerperium - baby delivered | 0 |
| L1833 | Drug dependence during pregnancy - baby not yet delivered | 0 |
| L1834 | Drug dependence in the puerperium - baby delivered during previous episode of care | 0 |
| Q4850 | Neonatal withdrawal symptoms from maternal use of drugs of addiction | 0 |
| U6097 | [X]Psychostimulants with abuse potential causing adverse effects in therapeutic use | 0 |
| ZV113 | [V]Personal history of alcoholism | 0 |
| ZV114 | [V]Personal history of psychoactive substance abuse | 0 |
| ZV115 | [V]Personal history of drug abuse by injection | 0 |
| ZV4K1 | [V]Drug use | 0 |
| ZVu54 | [X]Other problems related to care provider dependency | 0 |

---

#### CONCEPT: Eating disorders¶

| READ\_CODE | DESCRIPTION\_SAIL | SENSITIVE\_CODE |
| --- | --- | --- |
| 1467. | H/O: anorexia nervosa | 0 |
| 1JZ.. | Suspected binge eating disorder | 0 |
| 8HTN. | Referral to eating disorders clinic | 0 |
| 9Nk9. | Seen in eating disorder clinic | 0 |
| E2642 | Cyclical vomiting - psychogenic | 0 |
| E271. | Anorexia nervosa | 0 |
| E275. | Other and unspecified non-organic eating disorders | 0 |
| E2750 | Unspecified non-organic eating disorder | 0 |
| E2751 | Bulimia (non-organic overeating) | 0 |
| E2752 | Pica | 0 |
| E2754 | Psychogenic vomiting NOS | 0 |
| E275y | Other specified non-organic eating disorder | 0 |
| E275z | Non-organic eating disorder NOS | 0 |
| Eu50. | [X]Eating disorders | 0 |
| Eu500 | [X]Anorexia nervosa | 0 |
| Eu501 | [X]Atypical anorexia nervosa | 0 |
| Eu502 | [X]Bulimia nervosa | 0 |
| Eu503 | [X]Atypical bulimia nervosa | 0 |
| Eu504 | [X]Overeating associated with other psychological disturbances | 0 |
| Eu505 | [X]Vomiting associated with other psychological disturbances | 0 |
| Eu50y | [X]Other eating disorders | 0 |
| Eu50z | [X]Eating disorder, unspecified | 0 |
| Fy05. | Nocturnal sleep-related eating disorder | 0 |
| R0360 | [D]Excessive eating | 0 |

---

#### CONCEPT: Ehlers Danlos Syndrome¶

| READ\_CODE | DESCRIPTION\_SAIL | SENSITIVE\_CODE |
| --- | --- | --- |
| N235. | Hypermobility syndrome | 0 |
| PGy22 | Ehlers-Danlos syndrome type III | 0 |

---

#### CONCEPT: Endometriosis¶

| READ\_CODE | DESCRIPTION\_SAIL | SENSITIVE\_CODE |
| --- | --- | --- |
| 7E0D8 | Laparoscopic laser destruction of endometriosis | 0 |
| BBL1. | [M]Endolymphatic stromal myosis | 0 |
| K50.. | Endometriosis | 0 |
| K500. | Endometriosis of uterus | 0 |
| K5000 | Internal endometriosis | 0 |
| K5001 | Endometriosis of myometrium | 0 |
| K5002 | Endometriosis of cervix | 0 |
| K500z | Endometriosis of uterus NOS | 0 |
| K501. | Endometriosis of ovary | 0 |
| K502. | Endometriosis of the fallopian tube | 0 |
| K503. | Endometriosis of the pelvic peritoneum | 0 |
| K5030 | Endometriosis of the broad ligament | 0 |
| K5031 | Endometriosis of the pouch of Douglas | 0 |
| K5032 | Endometriosis of the parametrium | 0 |
| K5033 | Endometriosis of the round ligament | 0 |
| K503z | Endometriosis of the pelvic peritoneum NOS | 0 |
| K504. | Endometriosis of the rectovaginal septum and vagina | 0 |
| K5040 | Endometriosis of the rectovaginal septum | 0 |
| K5041 | Endometriosis of the vagina | 0 |
| K504z | Endometriosis of the rectovaginal septum and vagina NOS | 0 |
| K505. | Endometriosis of the intestine | 0 |
| K5050 | Endometriosis of the appendix | 0 |
| K5051 | Endometriosis of the colon | 0 |
| K5052 | Endometriosis of the rectum | 0 |
| K505z | Endometriosis of the intestine NOS | 0 |
| K506. | Endometriosis in scar of skin | 0 |
| K50y. | Other endometriosis | 0 |
| K50y0 | Endometriosis of the bladder | 0 |
| K50y1 | Endometriosis of the lung | 0 |
| K50y2 | Endometriosis of the umbilicus | 0 |
| K50y3 | Endometriosis of the vulva | 0 |
| K50yz | Other endometriosis NOS | 0 |
| K50z. | Endometriosis NOS | 0 |
| Kyu90 | [X]Other endometriosis | 0 |

---

#### CONCEPT: Epilepsy¶

| READ\_CODE | DESCRIPTION\_SAIL | SENSITIVE\_CODE |
| --- | --- | --- |
| 1473. | H/O: epilepsy | 0 |
| 14On. | At risk of sudden unexpected death in epilepsy | 0 |
| 1B1W. | Transient epileptic amnesia | 0 |
| 1O30. | Epilepsy confirmed | 0 |
| 2828. | Absence seizure | 0 |
| 667.. | Epilepsy monitoring | 0 |
| 6671. | Initial epilepsy assessment | 0 |
| 6672. | Follow-up epilepsy assessment | 0 |
| 6673. | Driving licence | 0 |
| 6674. | Epilepsy associated problems | 0 |
| 6675. | Fit frequency | 0 |
| 6676. | Last fit | 0 |
| 6677. | Epilepsy drug side effects | 0 |
| 6678. | Epilepsy treatment changed | 0 |
| 6679. | Epilepsy treatment started | 0 |
| 667A. | Epilepsy treatment stopped | 0 |
| 667B. | Nocturnal epilepsy | 0 |
| 667C. | Epilepsy control good | 0 |
| 667D. | Epilepsy control poor | 0 |
| 667E. | Epilepsy care arrangement | 0 |
| 667G. | Epilepsy restricts employment | 0 |
| 667H. | Epilepsy prevents employment | 0 |
| 667J. | Epilepsy impairs education | 0 |
| 667K. | Epilepsy limits activities | 0 |
| 667L. | Epilepsy does not limit activities | 0 |
| 667M. | Epilepsy management plan given | 0 |
| 667N. | Epilepsy severity | 0 |
| 667Q. | 1 to 12 seizures a year | 0 |
| 667R. | 2 to 4 seizures a month | 0 |
| 667S. | 1 to 7 seizures a week | 0 |
| 667T. | Daily seizures | 0 |
| 667V. | Many seizures a day | 0 |
| 667W. | Emergency epilepsy treatment since last appointment | 0 |
| 667X. | No epilepsy drug side effects | 0 |
| 667Z. | Epilepsy monitoring NOS | 0 |
| 8B66. | Anticonvulsant therapy | 0 |
| 8BIF. | Epilepsy medication review | 0 |
| Eu052 | [X]Organic delusional [schizophrenia-like] disorder | 0 |
| Eu05y | [X]Other specified mental disorder due to brain damage and dysfunction and to physical disease | 0 |
| Eu060 | [X]Organic personality disorder | 0 |
| Eu803 | [X]Acquired aphasia with epilepsy [Landau - Kleffner] | 0 |
| F1321 | Progressive myoclonic epilepsy | 0 |
| F25.. | Epilepsy | 0 |
| F250. | Generalised nonconvulsive epilepsy | 0 |
| F2500 | Petit mal (minor) epilepsy | 0 |
| F2501 | Pykno-epilepsy | 0 |
| F2502 | Epileptic seizures - atonic | 0 |
| F2503 | Epileptic seizures - akinetic | 0 |
| F2504 | Juvenile absence epilepsy | 0 |
| F2505 | Lennox-Gastaut syndrome | 0 |
| F250y | Other specified generalised nonconvulsive epilepsy | 0 |
| F250z | Generalised nonconvulsive epilepsy NOS | 0 |
| F251. | Generalised convulsive epilepsy | 0 |
| F2510 | Grand mal (major) epilepsy | 0 |
| F2511 | Neonatal myoclonic epilepsy | 0 |
| F2512 | Epileptic seizures - clonic | 0 |
| F2513 | Epileptic seizures - myoclonic | 0 |
| F2514 | Epileptic seizures - tonic | 0 |
| F2515 | Tonic-clonic epilepsy | 0 |
| F2516 | Grand mal seizure | 0 |
| F251y | Other specified generalised convulsive epilepsy | 0 |
| F251z | Generalised convulsive epilepsy NOS | 0 |
| F252. | Petit mal status | 0 |
| F253. | Grand mal status | 0 |
| F254. | Partial epilepsy with impairment of consciousness | 0 |
| F2540 | Temporal lobe epilepsy | 0 |
| F2541 | Psychomotor epilepsy | 0 |
| F2542 | Psychosensory epilepsy | 0 |
| F2543 | Limbic system epilepsy | 0 |
| F2544 | Epileptic automatism | 0 |
| F2545 | Complex partial epileptic seizure | 0 |
| F254z | Partial epilepsy with impairment of consciousness NOS | 0 |
| F255. | Partial epilepsy without mention of impairment of consciousness | 0 |
| F2550 | Jacksonian, focal or motor epilepsy | 0 |
| F2551 | Sensory induced epilepsy | 0 |
| F2552 | Somatosensory epilepsy | 0 |
| F2553 | Visceral reflex epilepsy | 0 |
| F2554 | Visual reflex epilepsy | 0 |
| F2555 | Unilateral epilepsy | 0 |
| F2556 | Simple partial epileptic seizure | 0 |
| F255y | Other specified partial epilepsy without mention of impairment of consciousness | 0 |
| F255z | Partial epilepsy without mention of impairment of consciousness NOS | 0 |
| F256. | Infantile spasms | 0 |
| F2560 | Hypsarrhythmia | 0 |
| F2561 | Salaam attacks | 0 |
| F256z | Infantile spasms NOS | 0 |
| F257. | Kojevnikov's epilepsy | 0 |
| F258. | Post-ictal state | 0 |
| F259. | Early infantile epileptic encephalopathy with suppression bursts | 0 |
| F25A. | Juvenile myoclonic epilepsy | 0 |
| F25B. | Alcohol-induced epilepsy | 0 |
| F25C. | Drug-induced epilepsy | 0 |
| F25D. | Menstrual epilepsy | 0 |
| F25E. | Stress-induced epilepsy | 0 |
| F25F. | Photosensitive epilepsy | 0 |
| F25G. | Severe myoclonic epilepsy in infancy | 0 |
| F25X. | Status epilepticus, unspecified | 0 |
| F25y. | Other forms of epilepsy | 0 |
| F25y0 | Cursive (running) epilepsy | 0 |
| F25y1 | Gelastic epilepsy | 0 |
| F25y2 | Localization-related(focal)(partial)idiopathic epilepsy and epileptic syndromes with seizures of localised onset | 0 |
| F25y3 | Complex partial status epilepticus | 0 |
| F25y4 | Benign Rolandic epilepsy | 0 |
| F25y5 | Panayiotopoulos syndrome | 0 |
| F25yz | Other forms of epilepsy NOS | 0 |
| F25z. | Epilepsy NOS | 0 |
| Fyu50 | [X]Other generalized epilepsy and epileptic syndromes | 0 |
| Fyu51 | [X]Other epilepsy | 0 |
| Fyu52 | [X]Other status epilepticus | 0 |
| Fyu59 | [X]Status epilepticus, unspecified | 0 |
| SC200 | Traumatic epilepsy | 0 |

---

#### CONCEPT: Fibromyalgia¶

| READ\_CODE | DESCRIPTION\_SAIL | SENSITIVE\_CODE |
| --- | --- | --- |
| N239. | Fibromyalgia | 0 |
| N248. | Fibromyalgia | 0 |

---

#### CONCEPT: Glaucoma¶

| READ\_CODE | DESCRIPTION\_SAIL | SENSITIVE\_CODE |
| --- | --- | --- |
| 1482. | H/O: glaucoma | 0 |
| 66T1. | Glaucoma monitoring | 0 |
| 7259. | Operations following glaucoma surgery | 0 |
| 72590 | Needling of bleb following glaucoma surgery | 0 |
| 72591 | Injection of bleb following glaucoma surgery | 0 |
| 72592 | Revision of bleb NEC following glaucoma surgery | 0 |
| 72593 | Removal of releasable suture following glaucoma surgery | 0 |
| 72594 | Laser suture lysis following glaucoma surgery | 0 |
| 7259y | Other specified operations following glaucoma surgery | 0 |
| 7259z | Operations following glaucoma surgery NOS | 0 |
| 7275. | Pan retinal photocoagulation for glaucoma | 0 |
| F4042 | Blind hypertensive eye | 0 |
| F4421 | Glaucomatocyclitic crises | 0 |
| F45.. | Glaucoma | 0 |
| F450. | Borderline glaucoma | 0 |
| F4500 | Unspecified preglaucoma | 0 |
| F4501 | Open angle glaucoma with borderline intraocular pressure | 0 |
| F4502 | Borderline glaucoma with anatomical narrow angle | 0 |
| F4503 | Borderline glaucoma steroid responder | 0 |
| F450z | Borderline glaucoma NOS | 0 |
| F451. | Open-angle glaucoma | 0 |
| F4510 | Unspecified open-angle glaucoma | 0 |
| F4511 | Primary open-angle glaucoma | 0 |
| F4512 | Low tension glaucoma | 0 |
| F4513 | Pigmentary glaucoma | 0 |
| F4514 | Glaucoma of childhood | 0 |
| F4515 | Open-angle glaucoma residual stage | 0 |
| F451z | Open-angle glaucoma NOS | 0 |
| F452. | Primary angle-closure glaucoma | 0 |
| F4520 | Unspecified primary angle-closure glaucoma | 0 |
| F4521 | Intermittent primary angle-closure glaucoma | 0 |
| F4522 | Acute primary angle-closure glaucoma | 0 |
| F4523 | Chronic primary angle-closure glaucoma | 0 |
| F4524 | Primary angle-closure glaucoma residual stage | 0 |
| F452z | Primary angle-closure glaucoma NOS | 0 |
| F453. | Steroid-induced glaucoma | 0 |
| F4530 | Steroid-induced glaucoma glaucomatous stage | 0 |
| F4531 | Steroid-induced glaucoma residual stage | 0 |
| F453z | Steroid-induced glaucoma NOS | 0 |
| F454. | Glaucoma due to disease EC | 0 |
| F4540 | Glaucoma due to chamber angle anomaly | 0 |
| F4541 | Glaucoma due to iris anomaly | 0 |
| F4542 | Glaucoma due to other anterior segment anomaly | 0 |
| F4543 | Glaucoma due to systemic syndrome | 0 |
| F4544 | Glaucoma in endocrine, nutritional and metabolic diseases | 0 |
| F454z | Glaucoma due to disease NOS | 0 |
| F455. | Glaucoma associated with disorders of the lens | 0 |
| F4550 | Phacolytic glaucoma | 0 |
| F4551 | Pseudoexfoliation glaucoma | 0 |
| F455z | Glaucoma associated with disorders of the lens NOS | 0 |
| F456. | Glaucoma associated with other ocular disorders | 0 |
| F4560 | Glaucoma due to unspecified ocular disorder | 0 |
| F4561 | Glaucoma due to pupillary block | 0 |
| F4562 | Glaucoma due to ocular inflammation | 0 |
| F4563 | Glaucoma due to ocular vascular disorder | 0 |
| F4564 | Glaucoma due to ocular tumour or cyst | 0 |
| F4565 | Glaucoma due to ocular trauma | 0 |
| F4566 | Neovascular glaucoma | 0 |
| F456z | Glaucoma associated with other ocular disorders NOS | 0 |
| F45y. | Other specified forms of glaucoma | 0 |
| F45y0 | Hypersecretion glaucoma | 0 |
| F45y1 | Glaucoma due to episode of increased venous pressure | 0 |
| F45y2 | Low tension glaucoma | 0 |
| F45yz | Other specified glaucoma NOS | 0 |
| F45z. | Glaucoma NOS | 0 |
| F4631 | Glaucomatous subcapsular flecks | 0 |
| F4H14 | Optic disc glaucomatous atrophy | 0 |
| FyuG. | [X]Glaucoma | 0 |
| FyuG0 | [X]Other glaucoma | 0 |
| FyuG1 | [X]Glaucoma in endocrine, nutritional and metabolic diseases classified elsewhere | 0 |
| FyuG2 | [X]Glaucoma in other diseases classified elsewhere | 0 |
| P3200 | Congenital glaucoma | 0 |
| Q20y7 | Traumatic glaucoma due to birth trauma | 0 |

---

#### CONCEPT: Gout¶

| READ\_CODE | DESCRIPTION\_SAIL | SENSITIVE\_CODE |
| --- | --- | --- |
| 1443. | H/O: gout | 0 |
| 2D52. | O/E - auricle of ear - tophi | 0 |
| 669.. | Gout monitoring | 0 |
| 6691. | Initial gout assessment | 0 |
| 6692. | Follow-up gout assessment | 0 |
| 6693. | Joints gout affected | 0 |
| 6695. | Date gout treatment started | 0 |
| 6696. | Date of last gout attack | 0 |
| 6697. | Gout associated problems | 0 |
| 6698. | Gout drug side effects | 0 |
| 6699. | Gout treatment changed | 0 |
| 669A. | Date gout treatment stopped | 0 |
| 669Z. | Gout monitoring NOS | 0 |
| C34.. | Gout | 0 |
| C340. | Gouty arthropathy | 0 |
| C341. | Gouty nephropathy | 0 |
| C3410 | Gouty nephropathy unspecified | 0 |
| C341z | Gouty nephropathy NOS | 0 |
| C342. | Idiopathic gout | 0 |
| C343. | Lead-induced gout | 0 |
| C344. | Drug-induced gout | 0 |
| C345. | Gout due to impairment of renal function | 0 |
| C346. | Acute exacerbation of gout | 0 |
| C34y. | Other specified gouty manifestation | 0 |
| C34y0 | Gouty tophi of ear | 0 |
| C34y1 | Gouty tophi of heart | 0 |
| C34y2 | Gouty tophi of other sites | 0 |
| C34y3 | Gouty iritis | 0 |
| C34y4 | Gouty neuritis | 0 |
| C34y5 | Gouty tophi of hand | 0 |
| C34y6 | Gouty tophi of toe | 0 |
| C34yz | Other specified gouty manifestation NOS | 0 |
| C34z. | Gout NOS | 0 |
| G5573 | Gouty tophi of heart | 0 |
| N023. | Gouty arthritis | 0 |
| N0230 | Gouty arthritis of unspecified site | 0 |
| N0231 | Gouty arthritis of the shoulder region | 0 |
| N0232 | Gouty arthritis of the upper arm | 0 |
| N0233 | Gouty arthritis of the forearm | 0 |
| N0234 | Gouty arthritis of the hand | 0 |
| N0235 | Gouty arthritis of the pelvic region and thigh | 0 |
| N0236 | Gouty arthritis of the lower leg | 0 |
| N0237 | Gouty arthritis of the ankle and foot | 0 |
| N0238 | Gouty arthritis of toe | 0 |
| N023x | Gouty arthritis of multiple sites | 0 |
| N023y | Gouty arthritis of other specified site | 0 |
| N023z | Gouty arthritis NOS | 0 |
| Nyu17 | [X]Other secondary gout | 0 |
| Nyu1C | [X]Gouty arthropathy due to enzyme defects and other inherited disorders classified elsewhere | 0 |

---

#### CONCEPT: Heart Valve Disorders¶

| READ\_CODE | DESCRIPTION\_SAIL | SENSITIVE\_CODE |
| --- | --- | --- |
| 0Z1A. | Valveman | 0 |
| 14S4. | H/O: heart valve recipient | 0 |
| 14T3. | H/O: artificial heart valve | 0 |
| 24G.. | Aortic valve gradient | 0 |
| 70101 | Creation of ventriculovascular anastomosis | 0 |
| 70114 | Renewal of valve of cerebroventricular shunt | 0 |
| 70160 | Drainage of ventricle of brain NEC | 0 |
| 74493 | Insertion of endobronchial valve | 0 |
| 744D1 | Changing of tracheooesophageal valve | 0 |
| 74583 | Bag valve mask ventilation | 0 |
| 76112 | Partial gastrectomy and anastomosis of stomach to jejunum NEC | 0 |
| 79020 | Correction of tetralogy of Fallot using valved right ventricular outflow conduit | 0 |
| 79025 | Repair of tetralogy of Fallot with absent pulmonary valve | 0 |
| 790D. | Creation of valved cardiac conduit | 0 |
| 790D0 | Creation of valved conduit between atrium and ventricle of heart | 0 |
| 790D1 | Creation of valved conduit between right atrium and pulmonary artery | 0 |
| 790D2 | Creation of valved conduit between right ventricle of heart and pulmonary artery | 0 |
| 790D3 | Creation of valved conduit between left ventricle of heart and aorta | 0 |
| 790D4 | Revision of valved cardiac conduit | 0 |
| 790D6 | Creation of valved conduit between left ventricle of heart and pulmonary artery | 0 |
| 790D7 | Replacement of valved cardiac conduit | 0 |
| 790Dy | Other specified creation of valved cardiac conduit | 0 |
| 790Dz | Creation of valved cardiac conduit NOS | 0 |
| 790M3 | Radical aortopulmonary reconstruction with right ventricle to pulmonary arterial valveless conduit | 0 |
| 790M8 | Aortopulmonary reconstruction with right ventricle to pulmonary arterial valveless conduit | 0 |
| 791.. | Valves of heart and adjacent structures operations | 0 |
| 7910. | Plastic repair of mitral valve | 0 |
| 79100 | Allograft replacement of mitral valve | 0 |
| 79101 | Xenograft replacement of mitral valve | 0 |
| 79102 | Prosthetic replacement of mitral valve | 0 |
| 79103 | Replacement of mitral valve NEC | 0 |
| 79104 | Mitral valvuloplasty NEC | 0 |
| 7910y | Other specified plastic repair of mitral valve | 0 |
| 7910z | Plastic repair of mitral valve NOS | 0 |
| 7911. | Plastic repair of aortic valve | 0 |
| 79110 | Allograft replacement of aortic valve | 0 |
| 79111 | Xenograft replacement of aortic valve | 0 |
| 79112 | Prosthetic replacement of aortic valve | 0 |
| 79113 | Replacement of aortic valve NEC | 0 |
| 79114 | Aortic valvuloplasty NEC | 0 |
| 79115 | Transapical aortic valve implantation | 0 |
| 79116 | Transluminal aortic valve implantation | 0 |
| 7911y | Other specified plastic repair of aortic valve | 0 |
| 7911z | Plastic repair of aortic valve NOS | 0 |
| 7912. | Plastic repair of tricuspid valve | 0 |
| 79120 | Allograft replacement of tricuspid valve | 0 |
| 79121 | Xenograft replacement of tricuspid valve | 0 |
| 79122 | Prosthetic replacement of tricuspid valve | 0 |
| 79123 | Replacement of tricuspid valve NEC | 0 |
| 79124 | Repositioning of tricuspid valve | 0 |
| 79125 | Tricuspid valvuloplasty NEC | 0 |
| 7912y | Other specified plastic repair of tricuspid valve | 0 |
| 7912z | Plastic repair of tricuspid valve NOS | 0 |
| 7913. | Plastic repair of pulmonary valve | 0 |
| 79130 | Allograft replacement of pulmonary valve | 0 |
| 79131 | Xenograft replacement of pulmonary valve | 0 |
| 79132 | Prosthetic replacement of pulmonary valve | 0 |
| 79133 | Replacement of pulmonary valve NEC | 0 |
| 79134 | Pulmonary valvuloplasty NEC | 0 |
| 7913y | Other specified plastic repair of pulmonary valve | 0 |
| 7913z | Plastic repair of pulmonary valve NOS | 0 |
| 7914. | Plastic repair of unspecified valve of heart | 0 |
| 79140 | Allograft replacement of valve of heart NEC | 0 |
| 79141 | Xenograft replacement of valve of heart NEC | 0 |
| 79142 | Prosthetic replacement of valve of heart NEC | 0 |
| 79143 | Replacement of valve of heart NEC | 0 |
| 79144 | Valvuloplasty of heart NEC | 0 |
| 79145 | Truncal valve repair | 0 |
| 79146 | Replacement of truncal valve | 0 |
| 7914y | Other specified plastic repair of unspecified valve of heart | 0 |
| 7914z | Plastic repair of unspecified valve of heart NOS | 0 |
| 7915. | Revision of plastic repair of valve of heart | 0 |
| 79150 | Revision of plastic repair of mitral valve | 0 |
| 79151 | Revision of plastic repair of aortic valve | 0 |
| 79152 | Revision of plastic repair of tricuspid valve | 0 |
| 79153 | Revision of plastic repair of pulmonary valve | 0 |
| 79154 | Revision of plastic repair of truncal valve | 0 |
| 7915y | Other specified revision of plastic repair of valve of heart | 0 |
| 7915z | Revision of plastic repair of valve of heart NOS | 0 |
| 7916. | Open incision of heart valve | 0 |
| 79160 | Open mitral valvotomy | 0 |
| 79161 | Open aortic valvotomy | 0 |
| 79162 | Open tricuspid valvotomy | 0 |
| 79163 | Open pulmonary valvotomy | 0 |
| 79164 | Open truncal valvotomy | 0 |
| 7916y | Other specified open specified incision of valve of heart | 0 |
| 7916z | Open incision of valve of heart NOS | 0 |
| 7917. | Closed incision of heart valve | 0 |
| 79170 | Closed mitral valvotomy | 0 |
| 79171 | Closed aortic valvotomy | 0 |
| 79172 | Closed tricuspid valvotomy | 0 |
| 79173 | Closed pulmonary valvotomy | 0 |
| 7917y | Other specified closed incision of valve of heart | 0 |
| 7917z | Closed incision of valve of heart NOS | 0 |
| 7918. | Other open operations on valve of heart | 0 |
| 79180 | Annuloplasty of mitral valve | 0 |
| 79181 | Annuloplasty of tricuspid valve | 0 |
| 79182 | Annuloplasty of valve of heart NEC | 0 |
| 79183 | Excision of vegetations of valve of heart | 0 |
| 79184 | Closure of tricuspid valve | 0 |
| 79185 | Closure of pulmonary valve | 0 |
| 7918y | Other specified other open operation on valve of heart | 0 |
| 7918z | Other open operation on valve of heart NOS | 0 |
| 7919. | Therapeutic transluminal operations on valve of heart | 0 |
| 79190 | Percutaneous transluminal mitral valvotomy | 0 |
| 79191 | Percutaneous transluminal aortic valvotomy | 0 |
| 79192 | Percutaneous transluminal tricuspid valvotomy | 0 |
| 79193 | Percutaneous transluminal pulmonary valvotomy | 0 |
| 79194 | Percutaneous transluminal valvuloplasty | 0 |
| 79195 | Percutaneous transluminal pulmonary valve perforation and dilation | 0 |
| 79196 | Percutaneous transluminal pulmonary valve replacement | 0 |
| 7919y | Other specified therapeutic transluminal operation on valve of heart | 0 |
| 7919z | Therapeutic transluminal operation on heart valve NOS | 0 |
| 791A. | Removal of obstruction from structure adjacent to valve of heart | 0 |
| 791A0 | Infundibulectomy of heart using patch | 0 |
| 791A1 | Infundibulectomy of heart NEC | 0 |
| 791A2 | Repair of subaortic stenosis | 0 |
| 791A3 | Repair of supraaortic stenosis | 0 |
| 791A4 | Excision of supramitral ring | 0 |
| 791A5 | Aortoventriculoplasty | 0 |
| 791Ay | Other specified removal of obstruction from structure adjacent to valve of heart | 0 |
| 791Az | Removal of obstruction from structure adjacent to valve of heart NOS | 0 |
| 791B. | Other operations on structure adjacent to valve of heart | 0 |
| 791B0 | Operation on papillary muscle | 0 |
| 791B1 | Operation on chordae tendineae | 0 |
| 791B2 | Operations on mitral subvalvar apparatus | 0 |
| 791B3 | Closure of aorto-left ventricular tunnel | 0 |
| 791B5 | Repair of aortic sinus of Valsalva aneurysm | 0 |
| 791By | Other specified operation on structure adjacent to valve of heart | 0 |
| 791Bz | Operation on structure adjacent to valve of heart NOS | 0 |
| 791C0 | Aortic root replacement using pulmonary valve autograft with right ventricle to pulmonary artery valved conduit | 0 |
| 791C1 | Aortic root replacement using pulmonary valve autograft with right ventricle to pulmonary artery valved conduit and aortoventriculoplasty | 0 |
| 791C5 | Aortoventriculoplasty with pulmonary valve autograft | 0 |
| 791D. | Excision of valve of heart | 0 |
| 791D0 | Tricuspid valvectomy | 0 |
| 791D1 | Pulmonary valvectomy | 0 |
| 791Dy | Other specified excision of valve of heart | 0 |
| 791Dz | Excision of valve of heart NOS | 0 |
| 791y. | Other specified operations on heart valve or adjacent structures | 0 |
| 791z. | Heart valve and adjacent structures operations NOS | 0 |
| 79310 | Inspection of valve of heart | 0 |
| 7A65. | Repair of valve of vein | 0 |
| 7A650 | Transposition of valve of vein | 0 |
| 7A651 | Interposition of valve of vein | 0 |
| 7A65y | Other specified repair of valve of vein | 0 |
| 7A65z | Repair of valve of vein NOS | 0 |
| 7B402 | Open excision of urethral lesion | 0 |
| 7B435 | Endoscopic destruction of posterior urethral valve | 0 |
| 7B43A | Endoscopic destruction of urethral valves | 0 |
| 7B456 | Hook ablation of posterior urethral valve | 0 |
| 7N40. | [SO]Valve of heart | 0 |
| 7N400 | [SO]Mitral valve | 0 |
| 7N401 | [SO]Aortic valve | 0 |
| 7N402 | [SO]Tricuspid valve | 0 |
| 7N403 | [SO]Pulmonary valve | 0 |
| 7N404 | [SO]Truncal valve | 0 |
| 7N40y | [SO]Specified valve of heart NEC | 0 |
| 7N40z | [SO]Valve of heart NEC | 0 |
| A932. | Syphilitic endocarditis | 1 |
| A9320 | Syphilitic endocarditis of unspecified valve | 1 |
| A9321 | Syphilitic endocarditis of mitral valve | 1 |
| A9322 | Syphilitic endocarditis of aortic valve | 1 |
| A9323 | Syphilitic endocarditis of tricuspid valve | 1 |
| A9324 | Syphilitic endocarditis of pulmonary valve | 1 |
| A932z | Syphilitic endocarditis of heart valve NOS | 1 |
| B713. | Benign neoplasm of colon | 0 |
| B7139 | Benign neoplasm of ileocaecal valve | 0 |
| G11.. | Mitral valve diseases | 0 |
| G110. | Mitral stenosis | 0 |
| G111. | Rheumatic mitral insufficiency | 0 |
| G112. | Mitral stenosis with insufficiency | 0 |
| G113. | Nonrheumatic mitral valve stenosis | 0 |
| G114. | Ruptured mitral valve cusp | 0 |
| G11z. | Mitral valve disease NOS | 0 |
| G12.. | Rheumatic aortic valve disease | 0 |
| G120. | Rheumatic aortic stenosis | 0 |
| G121. | Rheumatic aortic insufficiency | 0 |
| G122. | Rheumatic aortic stenosis with insufficiency | 0 |
| G12z. | Rheumatic aortic valve disease NOS | 0 |
| G13.. | Diseases of mitral and aortic valves | 0 |
| G130. | Mitral and aortic stenosis | 0 |
| G131. | Mitral stenosis and aortic insufficiency | 0 |
| G132. | Mitral insufficiency and aortic stenosis | 0 |
| G133. | Mitral and aortic incompetence | 0 |
| G13y. | Multiple mitral and aortic valve involvement | 0 |
| G13z. | Mitral and aortic valve disease NOS | 0 |
| G14.. | Other chronic rheumatic endocardial disease | 0 |
| G140. | Tricuspid valve disease NEC | 0 |
| G1400 | Rheumatic tricuspid stenosis | 0 |
| G1401 | Rheumatic tricuspid insufficiency | 0 |
| G1402 | Rheumatic tricuspid stenosis and insufficiency | 0 |
| G1403 | Tricuspid stenosis, cause unspecified | 0 |
| G1404 | Tricuspid insufficiency, cause unspecified | 0 |
| G1405 | Tricuspid stenosis and insufficiency, cause unspecified | 0 |
| G140z | Rheumatic tricuspid valve disease NOS | 0 |
| G141. | Rheumatic pulmonary valve disease | 0 |
| G1410 | Rheumatic pulmonary stenosis | 0 |
| G1411 | Rheumatic pulmonary insufficiency | 0 |
| G1412 | Rheumatic pulmonary stenosis and insufficiency | 0 |
| G141z | Rheumatic pulmonary valve disease NOS | 0 |
| G14z. | Rheumatic endocarditis NOS | 0 |
| G1y.. | Other specified chronic rheumatic heart disease | 0 |
| G54.. | Other diseases of endocardium | 0 |
| G540. | Mitral valve incompetence | 0 |
| G5400 | Mitral incompetence, non-rheumatic | 0 |
| G5401 | Mitral incompetence, cause unspecified | 0 |
| G5402 | Mitral valve prolapse | 0 |
| G5403 | Mitral valve leaf prolapse | 0 |
| G540z | Mitral valve disorders NOS | 0 |
| G541. | Aortic valve disorders | 0 |
| G5410 | Aortic incompetence, non-rheumatic | 0 |
| G5411 | Aortic stenosis, non-rheumatic | 0 |
| G5412 | Aortic incompetence alone, cause unspecified | 0 |
| G5413 | Aortic stenosis alone, cause unspecified | 0 |
| G5414 | Aortic valve stenosis with insufficiency | 0 |
| G5415 | Aortic stenosis | 0 |
| G5416 | Aortic valve sclerosis | 0 |
| G5417 | Aortic valve calcification | 0 |
| G541z | Aortic valve disorders NOS | 0 |
| G542. | Tricuspid valve disorders, non-rheumatic | 0 |
| G5420 | Tricuspid incompetence, non-rheumatic | 0 |
| G5421 | Tricuspid stenosis, non-rheumatic | 0 |
| G5422 | Nonrheumatic tricuspid valve stenosis with insufficiency | 0 |
| G542X | Nonrheumatic tricuspid valve disorder, unspecified | 0 |
| G542z | Tricuspid valve disorders NOS | 0 |
| G543. | Pulmonary valve disorders | 0 |
| G5430 | Pulmonary incompetence, non-rheumatic | 0 |
| G5431 | Pulmonary stenosis, non-rheumatic | 0 |
| G5432 | Pulmonary incompetence, cause unspecified | 0 |
| G5433 | Pulmonary stenosis, cause unspecified | 0 |
| G5434 | Pulmonary valve stenosis with insufficiency | 0 |
| G543z | Pulmonary valve disorders NOS | 0 |
| G544. | Multiple valve diseases | 0 |
| G5440 | Disorders of both aortic and tricuspid valves | 0 |
| G5441 | Disorders of both mitral and tricuspid valves | 0 |
| G5442 | Combined disorders of mitral, aortic and tricuspid valves | 0 |
| G544X | Multiple valve disease, unspecified | 0 |
| G54z. | Endocarditis, valve unspecified | 0 |
| G54z0 | Incompetence of unspecified heart valve | 0 |
| G54z1 | Stenosis of unspecified heart valve | 0 |
| G54z2 | Chronic cardiac valvulitis NOS | 0 |
| G54z3 | Endocarditis, valve unspecified, OS | 0 |
| G54z5 | Valvular heart disease | 0 |
| G54zz | Endocarditis, valve unspecified, NOS | 0 |
| G5804 | Congestive heart failure due to valvular disease | 0 |
| Gyu10 | [X]Other mitral valve diseases | 0 |
| Gyu11 | [X]Other rheumatic aortic valve diseases | 0 |
| Gyu12 | [X]Other tricuspid valve diseases | 0 |
| Gyu13 | [X]Other multiple valve diseases | 0 |
| Gyu15 | [X]Multiple valve disease, unspecified | 0 |
| Gyu55 | [X]Other nonrheumatic mitral valve disorders | 0 |
| Gyu56 | [X]Other aortic valve disorders | 0 |
| Gyu57 | [X]Other nonrheumatic tricuspid valve disorders | 0 |
| Gyu58 | [X]Other pulmonary valve disorders | 0 |
| Gyu59 | [X]Mitral valve disorders in diseases classified elsewhere | 0 |
| Gyu5A | [X]Aortic valve disorders in diseases classified elsewhere | 0 |
| Gyu5B | [X]Tricuspid valve disorders in diseases classified elsewhere | 0 |
| Gyu5C | [X]Pulmonary valve disorders in diseases classified elsewhere | 0 |
| Gyu5D | [X]Multiple valve disorders in diseases classified elsewhere | 0 |
| Gyu5E | [X]Endocarditis, valve unspecified, in diseases classified elsewhere | 0 |
| Gyu5f | [X]Nonrheumatic tricuspid valve disorder, unspecified | 0 |
| P60.. | Pulmonary valve anomalies | 0 |
| P600. | Pulmonary valve anomaly, unspecified | 0 |
| P601. | Congenital atresia of the pulmonary valve | 0 |
| P6010 | Hypoplasia of pulmonary valve | 0 |
| P601z | Congenital atresia of pulmonary valve NOS | 0 |
| P602. | Congenital pulmonary stenosis | 0 |
| P6021 | Congenital fusion of pulmonary valve segment | 0 |
| P602z | Congenital pulmonary stenosis NOS | 0 |
| P60z. | Other pulmonary valve anomalies | 0 |
| P60z0 | Congenital insufficiency of the pulmonary valve | 0 |
| P60z2 | Supernumerary pulmonary valve cusps | 0 |
| P60zz | Other pulmonary valve anomaly NOS | 0 |
| P611. | Congenital tricuspid stenosis | 0 |
| P62.. | Ebstein's anomaly | 0 |
| P63.. | Congenital aortic valve stenosis | 0 |
| P64.. | Congenital aortic valve insufficiency | 0 |
| P640. | Congenital aortic valve insufficiency, unspecified | 0 |
| P641. | Bicuspid aortic valve | 0 |
| P64z. | Congenital aortic valve insufficiency NOS | 0 |
| P65.. | Congenital mitral stenosis | 0 |
| P650. | Congenital mitral stenosis, unspecified | 0 |
| P651. | Fused commissure of the mitral valve | 0 |
| P652. | Parachute deformity of the mitral valve | 0 |
| P653. | Supernumerary cusps of the mitral valve | 0 |
| P65z. | Congenital mitral stenosis NOS | 0 |
| P66.. | Congenital mitral insufficiency | 0 |
| P6W.. | Congenital malformation of aortic and mitral valves unspecified | 0 |
| P6X.. | Congenital malformation of tricuspid valve, unspecified | 0 |
| P6y0. | Subaortic stenosis | 0 |
| P6y2. | Pulmonary infundibular stenosis | 0 |
| P6yy. | Other specified heart anomalies | 0 |
| P6yy7 | Atresia of heart valve NEC | 0 |
| P6yyB | Supernumerary heart valve cusps NEC | 0 |
| P6yyC | Fusion of mitral valve cusps | 0 |
| P6yyD | Fusion of heart valve cusps NEC | 0 |
| P6z0. | Unspecified anomaly of heart valve | 0 |
| P711. | Preductal coarctation of aorta | 0 |
| P712. | Postductal coarctation of aorta | 0 |
| P7224 | Supra-valvular aortic stenosis | 0 |
| PD21. | Occlusion of ureter | 0 |
| PD62. | Congenital urethral valvular stricture | 0 |
| PD67. | Congenital posterior urethral valves | 0 |
| Pyu22 | [X]Other congenital malformations of pulmonary valve | 0 |
| Pyu23 | [X]Other congenital malformations of tricuspid valve | 0 |
| Pyu24 | [X]Other congenital malformations of aortic and mitral valves | 0 |
| Pyu2G | [X]Congenital malformation of tricuspid valve, unspecified | 0 |
| Pyu2H | [X]Congenital malformation of aortic and mitral valves unspecified | 0 |
| SP002 | Mechanical complication of heart valve prosthesis | 0 |
| SP004 | Infection and inflammatory reaction due to cardiac valve prosthesis | 0 |
| SyuK6 | [X]Other complications of cardiac and vascular prosthetic devices, implants and grafts | 0 |
| TB012 | Implant of heart valve prosthesis as the cause of abnormal reaction of patient, or of later complication, without mention of misadventure at the time of operation | 0 |
| ZV422 | [V]Heart valve transplanted | 0 |
| ZV433 | [V]Has artificial heart valve | 0 |
| ZV452 | [V]Has cerebrospinal fluid drainage device in situ | 0 |
| ZV45H | [V]Presence of prosthetic heart valve | 0 |
| ZVu6e | [X]Presence of other heart valve replacement | 0 |

---

#### CONCEPT: HF (Heart Failure)¶

| READ\_CODE | DESCRIPTION\_SAIL | SENSITIVE\_CODE |
| --- | --- | --- |
| 14A6. | H/O: heart failure | 0 |
| 14AM. | H/O: Heart failure in last year | 0 |
| 14S3. | H/O: heart recipient | 0 |
| 14T7. | H/O: artificial heart | 0 |
| 1O1.. | Heart failure confirmed | 0 |
| 23E1. | O/E - pulmonary oedema | 0 |
| 2JZ.. | On optimal heart failure therapy | 0 |
| 33BA. | Impaired left ventricular function | 0 |
| 388D. | New York Heart Association classification of heart failure symptoms | 0 |
| 585f. | Echocardiogram shows left ventricular systolic dysfunction | 0 |
| 585g. | Echocardiogram shows left ventricular diastolic dysfunction | 0 |
| 662T. | Congestive heart failure monitoring | 0 |
| 662W. | Heart failure annual review | 0 |
| 662f. | New York Heart Association classification - class I | 0 |
| 662g. | New York Heart Association classification - class II | 0 |
| 662h. | New York Heart Association classification - class III | 0 |
| 662i. | New York Heart Association classification - class IV | 0 |
| 662p. | Heart failure 6 month review | 0 |
| 7900. | Transplantation of heart and lung | 0 |
| 79000 | Allotransplantation of heart and lung | 0 |
| 79001 | Revision of transplantation of heart and lung | 0 |
| 7900y | Other specified transplantation of heart and lung | 0 |
| 7900z | Transplantation of heart and lung NOS | 0 |
| 7901. | Other transplantation of heart | 0 |
| 79010 | Allotransplantation of heart NEC | 0 |
| 79011 | Xenotransplantation of heart | 0 |
| 79012 | Implantation of prosthetic heart | 0 |
| 79013 | Piggyback transplantation of heart | 0 |
| 79015 | Revision of transplantation of heart NEC | 0 |
| 7901y | Other specified other transplantation of heart | 0 |
| 7901z | Other transplantation of heart NOS | 0 |
| 8B29. | Cardiac failure therapy | 0 |
| 8CeC. | Preferred place of care for next exacerbation of heart failure | 0 |
| 8H2S. | Admit heart failure emergency | 0 |
| 8HBE. | Heart failure follow-up | 0 |
| 8HHb. | Referral to heart failure nurse | 0 |
| 8HHz. | Referral to heart failure exercise programme | 0 |
| 8HTL. | Referral to heart failure clinic | 0 |
| 8HTL0 | Referral to rapid access heart failure clinic | 0 |
| 8HgD. | Discharge from heart failure nurse service | 0 |
| 8Hk0. | Referred to heart failure education group | 0 |
| 8IE1. | Referral to heart failure exercise programme declined | 0 |
| 9N0k. | Seen in heart failure clinic | 0 |
| 9N2p. | Seen by community heart failure nurse | 0 |
| 9N6T. | Referred by heart failure nurse specialist | 0 |
| 9Or0. | Heart failure review completed | 0 |
| G1yz1 | Rheumatic left ventricular failure | 0 |
| G21.. | Hypertensive heart disease | 0 |
| G210. | Malignant hypertensive heart disease | 0 |
| G2100 | Malignant hypertensive heart disease without congestive cardiac failure | 0 |
| G2101 | Malignant hypertensive heart disease with congestive cardiac failure | 0 |
| G210z | Malignant hypertensive heart disease NOS | 0 |
| G211. | Benign hypertensive heart disease | 0 |
| G2110 | Benign hypertensive heart disease without congestive cardiac failure | 0 |
| G2111 | Benign hypertensive heart disease with congestive cardiac failure | 0 |
| G211z | Benign hypertensive heart disease NOS | 0 |
| G21z. | Hypertensive heart disease NOS | 0 |
| G21z1 | Hypertensive heart disease NOS with congestive cardiac failure | 0 |
| G21zz | Hypertensive heart disease NOS | 0 |
| G23.. | Hypertensive heart and renal disease | 0 |
| G230. | Malignant hypertensive heart and renal disease | 0 |
| G231. | Benign hypertensive heart and renal disease | 0 |
| G232. | Hypertensive heart and renal disease with (congestive) heart failure | 0 |
| G233. | Hypertensive heart and renal disease with renal failure | 0 |
| G234. | Hypertensive heart and renal disease with both (congestive) heart failure and renal failure | 0 |
| G23z. | Hypertensive heart and renal disease NOS | 0 |
| G400. | Acute cor pulmonale | 0 |
| G41z. | Chronic pulmonary heart disease NOS | 0 |
| G5540 | Congestive cardiomyopathy | 0 |
| G5571 | Beriberi heart disease | 0 |
| G58.. | Heart failure | 0 |
| G580. | Congestive heart failure | 0 |
| G5800 | Acute congestive heart failure | 0 |
| G5801 | Chronic congestive heart failure | 0 |
| G5802 | Decompensated cardiac failure | 0 |
| G5803 | Compensated cardiac failure | 0 |
| G5804 | Congestive heart failure due to valvular disease | 0 |
| G581. | Left ventricular failure | 0 |
| G5810 | Acute left ventricular failure | 0 |
| G582. | Acute heart failure | 0 |
| G583. | Heart failure with normal ejection fraction | 0 |
| G584. | Right ventricular failure | 0 |
| G58z. | Heart failure NOS | 0 |
| G5y34 | Ventricular hypertrophy | 0 |
| G5y4z | Post cardiac operation heart failure NOS | 0 |
| G5yy9 | Left ventricular systolic dysfunction | 0 |
| G5yyA | Left ventricular diastolic dysfunction | 0 |
| G5yyD | Left ventricular cardiac dysfunction | 0 |
| Gyu5M | [X]Other hypertrophic cardiomyopathy | 0 |
| H54.. | Pulmonary congestion and hypostasis | 0 |
| H541. | Pulmonary congestion | 0 |
| H5410 | Chronic pulmonary oedema | 0 |
| H541z | Pulmonary oedema NOS | 0 |
| H54z. | Pulmonary congestion and hypostasis NOS | 0 |
| H584. | Acute pulmonary oedema unspecified | 0 |
| H584z | Acute pulmonary oedema NOS | 0 |
| L09y2 | Cardiac failure following abortive pregnancy | 1 |
| L1280 | Pre-existing hypertensive heart disease complicating pregnancy, childbirth and the puerperium | 0 |
| L1281 | Pre-existing hypertensive heart and renal disease complicating pregnancy, childbirth and the puerperium | 0 |
| P69.. | Left ventricular outflow tract obstruction | 0 |
| Q48y1 | Congenital cardiac failure | 0 |
| Q490. | Neonatal cardiac failure | 0 |
| R2y10 | [D]Cardiorespiratory failure | 0 |
| SP111 | Cardiac insufficiency as a complication of care | 0 |
| TB000 | Transplantation of heart as the cause of abnormal reaction of patient, or of later complication, without mention of misadventure at the time of operation | 0 |
| ZV421 | [V]Heart transplanted | 0 |
| ZV432 | [V]Has artificial heart | 0 |

---

#### CONCEPT: HIV/AIDS¶

| READ\_CODE | DESCRIPTION\_SAIL | SENSITIVE\_CODE |
| --- | --- | --- |
| 43C3. | HTLV-3 antibody positive | 1 |
| 43j7. | HIV 1 nucleic acid detection | 1 |
| 4J3P. | Human immunodeficiency virus type 1 subtype identification | 1 |
| 65QA. | AIDS carrier | 1 |
| A78.. | Other viral or chlamydial diseases | 1 |
| A788. | Acquired immune deficiency syndrome | 1 |
| A7880 | Acute human immunodeficiency virus infection | 1 |
| A7881 | Asymptomatic human immunodeficiency virus infection | 1 |
| A7882 | Human immunodeficiency virus infection with persistent generalised lymphadenopathy | 1 |
| A7883 | Human immunodeficiency virus with constitutional disease | 1 |
| A7884 | Human immunodeficiency virus with neurological disease | 1 |
| A7885 | Human immunodeficiency virus infection with secondary clinical infectious disease | 1 |
| A7886 | Human immunodeficiency virus with secondary cancers | 1 |
| A788U | HIV disease resulting in haematological and immunological abnormalities, not elsewhere classified | 1 |
| A788V | HIV disease resulting in multiple diseases classified elsewhere | 1 |
| A788W | HIV disease resulting in unspecified malignant neoplasm | 1 |
| A788X | HIV disease resulting in unspecified infectious and parasitic disease | 1 |
| A788y | Human immunodeficiency virus with other clinical findings | 1 |
| A788z | Acquired human immunodeficiency virus infection syndrome NOS | 1 |
| A789. | Human immunodef virus resulting in other disease | 1 |
| A7890 | HIV disease resulting in mycobacterial infection | 1 |
| A7891 | HIV disease resulting in cytomegaloviral disease | 1 |
| A7892 | HIV disease resulting in candidiasis | 1 |
| A7893 | HIV disease resulting in Pneumocystis carinii pneumonia | 1 |
| A7894 | HIV disease resulting in multiple infections | 1 |
| A7895 | HIV disease resulting in Kaposi's sarcoma | 1 |
| A7896 | HIV disease resulting in Burkitt's lymphoma | 1 |
| A7897 | HIV disease resulting in other types of non-Hodgkin's lymphoma | 1 |
| A7898 | HIV disease resulting in multiple malignant neoplasms | 1 |
| A7899 | HIV disease resulting in lymphoid interstitial pneumonitis | 1 |
| A789A | HIV disease resulting in wasting syndrome | 1 |
| A789X | HIV disease resulting in other malignant neoplasms of lymphoid, haematopoietic and related tissue | 1 |
| AyuC. | [X]Human immunodeficiency virus disease | 1 |
| AyuC0 | [X]HIV disease resulting in other bacterial infections | 1 |
| AyuC1 | [X]HIV disease resulting in other viral infections | 1 |
| AyuC2 | [X]HIV disease resulting in other mycoses | 1 |
| AyuC3 | [X]HIV disease resulting in multiple infections | 1 |
| AyuC4 | [X]Hiv disease resulting in other infectious and parasitic diseases | 1 |
| AyuC5 | [X]Hiv disease resulting in unspecified infectious and parasitic disease | 1 |
| AyuC6 | [X]HIV disease resulting in other non-Hodgkin's lymphoma | 1 |
| AyuC7 | [X]Hiv disease resulting in other malignant neoplasms of lymphoid, haematopoietic and related tissue | 1 |
| AyuC8 | [X]HIV disease resulting in other malignant neoplasms | 1 |
| AyuC9 | [X]HIV disease resulting in unspecified malignant neoplasm | 1 |
| AyuCA | [X]Hiv disease resulting in multiple diseases classified elsewhere | 1 |
| AyuCB | [X]Hiv disease resulting in haematological and immunological abnormalities, not elsewhere classified | 1 |
| AyuCC | [X]HIV disease resulting in other specified conditions | 1 |
| AyuCD | [X]Unspecified human immunodeficiency virus [HIV] disease | 1 |
| Eu024 | [X]Dementia in human immunodef virus [HIV] disease | 1 |
| L179. | Human immunodeficiency [HIV] disease complicating pregnancy, childbirth and the puerperium | 1 |
| R109. | [D]Laboratory evidence of human immunodeficiency virus [HIV] | 1 |
| ZV01A | [V]Asymptomatic human immunodeficiency virus infection status | 1 |

---

#### CONCEPT: Hypertension¶

| READ\_CODE | DESCRIPTION\_SAIL | SENSITIVE\_CODE |
| --- | --- | --- |
| 14A2. | H/O: hypertension | 0 |
| 21261 | Hypertension resolved | 0 |
| 212K. | Hypertension resolved | 0 |
| 61462 | Hypertension induced by oral contraceptive pill | 0 |
| 6624. | Borderline hyperten:yearly obs | 0 |
| 6627. | Good hypertension control | 0 |
| 6628. | Poor hypertension control | 0 |
| 6629. | Hypertension:follow-up default | 0 |
| 662F. | Hypertension treatm. started | 0 |
| 662G. | Hypertensive treatm.changed | 0 |
| 662H. | Hypertension treatm.stopped | 0 |
| 662O. | On treatment for hypertension | 0 |
| 662P. | Hypertension monitoring | 0 |
| 662P0 | Hypertension 9 month review | 0 |
| 662b. | Moderate hypertension control | 0 |
| 662c. | Hypertension six month review | 0 |
| 662d. | Hypertension annual review | 0 |
| 662q. | Trial reduction of antihypertensive therapy | 0 |
| 662r. | Trial withdrawal of antihypertensive therapy | 0 |
| 7Q01. | High cost hypertension drugs | 0 |
| 7Q01y | Other specified high cost hypertension drugs | 0 |
| 7Q01z | High cost hypertension drugs NOS | 0 |
| 8B26. | Antihypertensive therapy | 0 |
| 8BL0. | Patient on maximal tolerated antihypertensive therapy | 0 |
| 8CR4. | Hypertension clinical management plan | 0 |
| 8I3N. | Hypertension treatment refused | 0 |
| 8IA5. | Trial withdrawal of antihypertensive therapy declined | 0 |
| 8IA6. | Trial reduction of antihypertensive therapy declined | 0 |
| 9N03. | Seen in hypertension clinic | 0 |
| 9N1y2 | Seen in hypertension clinic | 0 |
| 9OI9. | Hypertens.monitor deleted | 0 |
| F4042 | Blind hypertensive eye | 0 |
| F4213 | Hypertensive retinopathy | 0 |
| G2... | Hypertensive disease | 0 |
| G20.. | Essential hypertension | 0 |
| G200. | Malignant essential hypertension | 0 |
| G201. | Benign essential hypertension | 0 |
| G202. | Systolic hypertension | 0 |
| G203. | Diastolic hypertension | 0 |
| G20z. | Essential hypertension NOS | 0 |
| G21.. | Hypertensive heart disease | 0 |
| G210. | Malignant hypertensive heart disease | 0 |
| G2100 | Malignant hypertensive heart disease without congestive cardiac failure | 0 |
| G2101 | Malignant hypertensive heart disease with congestive cardiac failure | 0 |
| G210z | Malignant hypertensive heart disease NOS | 0 |
| G211. | Benign hypertensive heart disease | 0 |
| G2110 | Benign hypertensive heart disease without congestive cardiac failure | 0 |
| G2111 | Benign hypertensive heart disease with congestive cardiac failure | 0 |
| G211z | Benign hypertensive heart disease NOS | 0 |
| G21z. | Hypertensive heart disease NOS | 0 |
| G21z0 | Hypertensive heart disease NOS without congestive cardic failure | 0 |
| G21z1 | Hypertensive heart disease NOS with congestive cardiac failure | 0 |
| G21zz | Hypertensive heart disease NOS | 0 |
| G22.. | Hypertensive renal disease | 0 |
| G220. | Malignant hypertensive renal disease | 0 |
| G221. | Benign hypertensive renal disease | 0 |
| G222. | Hypertensive renal disease with renal failure | 0 |
| G22z. | Hypertensive renal disease NOS | 0 |
| G23.. | Hypertensive heart and renal disease | 0 |
| G230. | Malignant hypertensive heart and renal disease | 0 |
| G231. | Benign hypertensive heart and renal disease | 0 |
| G232. | Hypertensive heart and renal disease with (congestive) heart failure | 0 |
| G233. | Hypertensive heart and renal disease with renal failure | 0 |
| G234. | Hypertensive heart and renal disease with both (congestive) heart failure and renal failure | 0 |
| G23z. | Hypertensive heart and renal disease NOS | 0 |
| G24.. | Secondary hypertension | 0 |
| G240. | Secondary malignant hypertension | 0 |
| G2400 | Secondary malignant renovascular hypertension | 0 |
| G240z | Secondary malignant hypertension NOS | 0 |
| G241. | Secondary benign hypertension | 0 |
| G2410 | Secondary benign renovascular hypertension | 0 |
| G241z | Secondary benign hypertension NOS | 0 |
| G244. | Hypertension secondary to endocrine disorders | 0 |
| G24z. | Secondary hypertension NOS | 0 |
| G24z0 | Secondary renovascular hypertension NOS | 0 |
| G24z1 | Hypertension secondary to drug | 0 |
| G24zz | Secondary hypertension NOS | 0 |
| G25.. | Stage 1 hypertension (NICE - National Institute for Health and Clinical Excellence 2011) | 0 |
| G250. | Stage 1 hypertension (NICE 2011) without evidence of end organ damage | 0 |
| G251. | Stage 1 hypertension (NICE 2011) with evidence of end organ damage | 0 |
| G26.. | Severe hypertension (NICE - National Institute for Health and Clinical Excellence 2011) | 0 |
| G27.. | Hypertension resistant to drug therapy | 0 |
| G28.. | Stage 2 hypertension (NICE - National Institute for Health and Clinical Excellence 2011) | 0 |
| G2y.. | Other specified hypertensive disease | 0 |
| G2z.. | Hypertensive disease NOS | 0 |
| G672. | Hypertensive encephalopathy | 0 |
| Gyu2. | [X]Hypertensive diseases | 0 |
| Gyu20 | [X]Other secondary hypertension | 0 |
| Gyu21 | [X]Hypertension secondary to other renal disorders | 0 |
| L120. | Benign essential hypertension complicating pregnancy, childbirth and the puerperium | 0 |
| L122. | Other pre-existing hypertension in preg/childbirth/puerp | 0 |
| L1220 | Other pre-existing hypertension complicating pregnancy, childbirth and the puerperium unspecified | 0 |
| L1221 | Other pre-existing hypertension complicating pregnancy, childbirth and the puerperium - delivered | 0 |
| L1223 | Other pre-existing hypertension complicating pregnancy, childbirth and the puerperium - not delivered | 0 |
| L1224 | Other pre-existing hypertension complicating pregnancy, childbirth and the puerperium with postnatal complication | 0 |
| L122z | Other pre-existing hypertension complicating pregnancy, childbirth and the puerperium NOS | 0 |
| L127z | Pre-eclampsia or eclampsia with pre-existing hypertension NOS | 0 |
| L128. | Pre-existing hypertension complicating pregnancy, childbirth and puerperium | 0 |
| L1280 | Pre-existing hypertensive heart disease complicating pregnancy, childbirth and the puerperium | 0 |
| Q000. | Fetus or neonate affected by maternal hypertensive disease | 0 |
| SLC6. | Other hypertensive agent poisoning | 0 |
| TJC7. | Adverse reaction to other antihypertensives | 0 |
| TJC7z | Adverse reaction to antihypertensives NOS | 0 |
| U60C5 | [X]Other antihypertensive drugs causing adverse effects in therapeutic use, not elsewhere classified | 0 |

---

#### CONCEPT: Hyperthyroidism¶

| READ\_CODE | DESCRIPTION\_SAIL | SENSITIVE\_CODE |
| --- | --- | --- |
| 1431. | H/O: hyperthyroidism | 0 |
| 212P. | Hyperthyroidism resolved | 0 |
| C02.. | Thyrotoxicosis | 0 |
| C020. | Toxic diffuse goitre | 0 |
| C0200 | Toxic diffuse goitre with no crisis | 0 |
| C0201 | Toxic diffuse goitre with crisis | 0 |
| C020z | Toxic diffuse goitre NOS | 0 |
| C021. | Toxic uninodular goitre | 0 |
| C0210 | Toxic uninodular goitre with no crisis | 0 |
| C0211 | Toxic uninodular goitre with crisis | 0 |
| C021z | Toxic uninodular goitre NOS | 0 |
| C022. | Toxic multinodular goitre | 0 |
| C0220 | Toxic multinodular goitre with no crisis | 0 |
| C0221 | Toxic multinodular goitre with crisis | 0 |
| C022z | Toxic multinodular goitre NOS | 0 |
| C023. | Toxic nodular goitre unspecified | 0 |
| C0230 | Toxic nodular goitre unspecified with no crisis | 0 |
| C0231 | Toxic nodular goitre unspecified with crisis | 0 |
| C023z | Toxic nodular goitre NOS | 0 |
| C024. | Thyrotoxicosis from ectopic thyroid nodule | 0 |
| C0240 | Thyrotoxicosis from ectopic thyroid nodule with no crisis | 0 |
| C0241 | Thyrotoxicosis from ectopic thyroid nodule with crisis | 0 |
| C024z | Thyrotoxicosis from ectopic thyroid nodule NOS | 0 |
| C02y. | Thyrotoxicosis of other specified origin | 0 |
| C02y0 | Thyrotoxicosis of other specified origin with no crisis | 0 |
| C02y1 | Thyrotoxicosis of other specified origin with crisis | 0 |
| C02yz | Thyrotoxicosis of other specified origin NOS | 0 |
| C02z. | Thyrotoxicosis without mention of goitre or other cause | 0 |
| C02z0 | Thyrotoxicosis without mention of goitre or other cause with no crisis | 0 |
| C02z1 | Thyrotoxicosis without mention of goitre or other cause with crisis | 0 |
| C02zz | Thyrotoxicosis NOS | 0 |
| F4G20 | Thyrotoxic exophthalmos | 0 |
| G5575 | Thyrotoxic heart disease | 0 |

---

#### CONCEPT: Hypothyroidism¶

| READ\_CODE | DESCRIPTION\_SAIL | SENSITIVE\_CODE |
| --- | --- | --- |
| 1432. | H/O: hypothyroidism | 0 |
| 66BB. | Hypothyroidism annual review | 0 |
| 71100 | Total thyroidectomy | 0 |
| 71110 | Excision of substernal thyroid tissue | 0 |
| 8CR5. | Hypothyroidism clinical management plan | 0 |
| 9Oj.. | Hypothyroidism monitoring administration | 0 |
| 9Oj0. | Hypothyroidism monitoring first letter | 0 |
| 9Oj1. | Hypothyroidism monitoring second letter | 0 |
| 9Oj2. | Hypothyroidism monitoring third letter | 0 |
| 9Oj3. | Hypothyroidism monitoring verbal invite | 0 |
| 9Oj4. | Hypothyroidism monitoring telephone invitation | 0 |
| C03.. | Congenital hypothyroidism | 0 |
| C031. | Goitrous cretin | 0 |
| C03y. | Other specified congenital hypothyroidism | 0 |
| C03y0 | Congenital hypothyroidism with diffuse goitre | 0 |
| C03y1 | Congenital hypothyroidism without goitre | 0 |
| C03z. | Congenital hypothyroidism NOS | 0 |
| C04.. | Acquired hypothyroidism | 0 |
| C040. | Postsurgical hypothyroidism | 0 |
| C041. | Other postablative hypothyroidism | 0 |
| C0410 | Irradiation hypothyroidism | 0 |
| C041z | Postablative hypothyroidism NOS | 0 |
| C042. | Iodine hypothyroidism | 0 |
| C043. | Other iatrogenic hypothyroidism | 0 |
| C0430 | Hypothyroidism resulting from para-aminosalicylic acid | 0 |
| C0431 | Hypothyroidism resulting from phenylbutazone | 0 |
| C0432 | Hypothyroidism resulting from resorcinol | 0 |
| C043z | Iatrogenic hypothyroidism NOS | 0 |
| C044. | Postinfectious hypothyroidism | 0 |
| C045. | Acquired atrophy of thyroid | 0 |
| C046. | Autoimmune myxoedema | 0 |
| C04y. | Other acquired hypothyroidism | 0 |
| C04z. | Hypothyroidism NOS | 0 |
| C04z0 | Premature puberty due to hypothyroidism | 0 |
| C04z1 | Myxoedema coma | 0 |
| C06y1 | Thyroid atrophy | 0 |
| C0A1. | Congenital iodine-deficiency syndrome, myxoedematous type | 0 |
| Cyu11 | [X]Other sp cified hypothyroidism | 0 |
| F11x5 | Cerebral degeneration due to myxoedema | 0 |
| F1441 | Cerebellar ataxia due to myxoedema | 0 |
| F3814 | Myasthenic syndrome due to hypothyroidism | 0 |
| F3953 | Myopathy due to myxoedema | 0 |
| Fyu15 | [X]Systemic atrophy affecting the central nervous system in myxoedema | 0 |
| Q4337 | Neonatal jaundice with congenital hypothyroidism | 0 |

---

#### CONCEPT: IBD (Inflammatory bowel disease)¶

| READ\_CODE | DESCRIPTION\_SAIL | SENSITIVE\_CODE |
| --- | --- | --- |
| 14C4. | H/O: colitis | 0 |
| J08z9 | Orofacial Crohn's disease | 0 |
| J40.. | Regional enteritis - Crohn's disease | 0 |
| J400. | Regional enteritis of the small bowel | 0 |
| J4000 | Regional enteritis of the duodenum | 0 |
| J4001 | Regional enteritis of the jejunum | 0 |
| J4002 | Crohn's disease of the terminal ileum | 0 |
| J4003 | Crohn's disease of the ileum unspecified | 0 |
| J4004 | Crohn's disease of the ileum NOS | 0 |
| J4005 | Exacerbation of Crohn's disease of small intestine | 0 |
| J400z | Crohn's disease of the small bowel NOS | 0 |
| J401. | Regional enteritis of the large bowel | 0 |
| J4010 | Regional enteritis of the colon | 0 |
| J4011 | Regional enteritis of the rectum | 0 |
| J4012 | Exacerbation of Crohn's disease of large intestine | 0 |
| J401z | Crohn's disease of the large bowel NOS | 0 |
| J402. | Regional ileocolitis | 0 |
| J40z. | Regional enteritis NOS | 0 |
| J41.. | Idiopathic proctocolitis | 0 |
| J410. | Ulcerative proctocolitis | 0 |
| J4100 | Ulcerative ileocolitis | 0 |
| J4101 | Ulcerative colitis | 0 |
| J4102 | Ulcerative rectosigmoiditis | 0 |
| J4103 | Ulcerative proctitis | 0 |
| J4104 | Exacerbation of ulcerative colitis | 0 |
| J410z | Ulcerative proctocolitis NOS | 0 |
| J411. | Ulcerative (chronic) enterocolitis | 0 |
| J412. | Ulcerative (chronic) ileocolitis | 0 |
| J413. | Ulcerative pancolitis | 0 |
| J41y. | Other idiopathic proctocolitis | 0 |
| J4212 | Chronic ischaemic enterocolitis | 0 |
| J4213 | Chronic ischaemic colitis | 0 |
| J42z0 | Ischaemic colitis NOS | 0 |
| J4302 | Radiation enterocolitis | 0 |
| J4303 | Radiation colitis | 0 |
| J4312 | Toxic enterocolitis | 0 |
| J4313 | Toxic colitis | 0 |
| J4322 | Allergic enterocolitis | 0 |
| J4323 | Allergic colitis | 0 |
| J4332 | Dietetic enterocolitis | 0 |
| J4333 | Dietetic colitis | 0 |
| J436. | Microscopic colitis | 0 |
| J4360 | Collagenous colitis | 0 |
| J4361 | Lymphocytic colitis | 0 |
| J437. | Colitis | 0 |
| J4z3. | Non-infective colitis NOS | 0 |
| J4z5. | Exacerbation of non-infective colitis | 0 |
| J4z6. | Indeterminate colitis | 0 |
| Jyu4. | [X]Noninfective enteritis and colitis | 0 |
| Jyu40 | [X]Other Crohn's disease | 0 |
| Jyu41 | [X]Other ulcerative colitis | 0 |
| N0310 | Arthropathy in ulcerative colitis | 0 |
| N0311 | Arthropathy in Crohn's disease | 0 |
| N0453 | Juvenile arthritis in Crohn's disease | 0 |
| N0454 | Juvenile arthritis in ulcerative colitis | 0 |

---

#### CONCEPT: IBS (Irritable Bowel Syndrome)¶

| READ\_CODE | DESCRIPTION\_SAIL | SENSITIVE\_CODE |
| --- | --- | --- |
| 14CF. | History of irritable bowel syndrome | 0 |
| J521. | Irritable colon - Irritable bowel syndrome | 0 |
| J5210 | Irritable bowel syndrome with diarrhoea | 0 |
| J529. | Generalised intestinal dysmotility | 0 |
| J52y. | Other functional disorders of the intestine | 0 |
| J52yz | Other functional disorder of the intestine NOS | 0 |
| J52z. | Functional gastrointestinal tract disorders NEC NOS | 0 |
| Jyu53 | [X]Other specified functional disorders of intestine | 0 |

---

#### CONCEPT: ILD (Inflammatory Lung Disease)¶

| READ\_CODE | DESCRIPTION\_SAIL | SENSITIVE\_CODE |
| --- | --- | --- |
| H35.. | Extrinsic allergic alveolitis | 0 |
| H350. | Farmers' lung | 0 |
| H351. | Bagassosis | 0 |
| H352. | Bird-fancier's lung | 0 |
| H3520 | Budgerigar-fanciers' lung | 0 |
| H3521 | Pigeon-fanciers' lung | 0 |
| H352z | Bird-fancier's lung NOS | 0 |
| H353. | Suberosis ( cork-handlers' lung ) | 0 |
| H354. | Malt workers' lung | 0 |
| H355. | Mushroom workers' lung | 0 |
| H356. | Maple bark strippers' lung | 0 |
| H35y. | Other allergic alveolitis | 0 |
| H35y0 | Cheese-washers' lung | 0 |
| H35y1 | Coffee-workers' lung | 0 |
| H35y2 | Fish-meal workers' lung | 0 |
| H35y5 | Pituitary snuff-takers' disease | 0 |
| H35y8 | Air-conditioner and humidifier lung | 0 |
| H35yz | Other allergic alveolitis NOS | 0 |
| H35z. | Allergic alveolitis and pneumonitis NOS | 0 |
| H35z0 | Allergic extrinsic alveolitis NOS | 0 |
| H35zz | Allergic alveolitis and pneumonitis NOS | 0 |
| H4... | Lung disease due to external agents | 0 |
| H40.. | Coal workers' pneumoconiosis | 0 |
| H42.. | Silica and silicate pneumoconiosis | 0 |
| H420. | Talc pneumoconiosis | 0 |
| H421. | Simple silicosis | 0 |
| H422. | Complicated silicosis | 0 |
| H423. | Massive silicotic fibrosis | 0 |
| H42z. | Silica pneumoconiosis NOS | 0 |
| H43.. | Pneumoconiosis due to other inorganic dust | 0 |
| H431. | Bauxite fibrosis of lung | 0 |
| H432. | Berylliosis | 0 |
| H433. | Graphite fibrosis of lung | 0 |
| H434. | Siderosis | 0 |
| H435. | Stannosis | 0 |
| H43z. | Pneumoconiosis due to inorganic dust NOS | 0 |
| H45.. | Pneumoconiosis NOS | 0 |
| H450. | Pneumoconiosis associated with tuberculosis | 0 |
| H4642 | Chronic pulmonary fibrosis due to chemical fumes | 0 |
| H4y10 | Chronic pulmonary fibrosis following radiation | 0 |
| H4y2. | Drug-induced interstitial lung disorders | 0 |
| H4y20 | Acute drug-induced interstitial lung disorders | 0 |
| H4y21 | Chronic drug-induced interstitial lung disorders | 0 |
| H55.. | Postinflammatory pulmonary fibrosis | 0 |
| H563. | Idiopathic fibrosing alveolitis | 0 |
| H5631 | Diffuse pulmonary fibrosis | 0 |
| H5632 | Pulmonary fibrosis | 0 |
| H5633 | Usual interstitial pneumonitis | 0 |
| H563z | Idiopathic fibrosing alveolitis NOS | 0 |
| H58y3 | Interstitial lung disease NEC | 0 |
| H58y5 | Respiratory bronchiolitis associated interstitial lung disease | 0 |
| H58y6 | Interstitial lung disease due to collagen vascular disease | 0 |
| H58y7 | Interstitial lung disease due to connective tissue disease | 0 |
| Hyu40 | [X]Pneumoconiosis due to other dust containing silica | 0 |
| Hyu48 | [X]Chronic and other pulmonary manifestations due to radiation | 0 |
| Hyu5. | [X]Other respiratory diseases principally affecting the interstitium | 0 |
| Hyu50 | [X]Other interstitial pulmonary diseases with fibrosis | 0 |
| Hyu51 | [X]Other specified interstitial pulmonary diseases | 0 |
| N04y0 | Rheumatoid lung | 0 |
| Q3171 | Prematurity with interstitial pulmonary fibrosis | 0 |

---

#### CONCEPT: Learning disability¶

| READ\_CODE | DESCRIPTION\_SAIL | SENSITIVE\_CODE |
| --- | --- | --- |
| 12K6. | Maternal learning disability | 0 |
| 12L5. | Paternal learning disability | 0 |
| 13Z4E | Learning difficulties | 0 |
| 1IN.. | Learning disability excluded | 0 |
| 69DB. | Learning disability health examination | 0 |
| 918W. | Carer of a person with learning disability | 0 |
| 918e. | On learning disability register | 0 |
| 949H. | Died in learning disability unit | 0 |
| 9HB.. | Learning disabilities administration status | 0 |
| 9HB1. | Learning disabilities health action plan offered | 0 |
| 9HB2. | Learning disabilities health action plan reviewed | 0 |
| 9HB3. | Learning disabilities health assessment | 0 |
| 9HB4. | Learning disabilities health action plan completed | 0 |
| 9HB5. | Learning disabilities annual health assessment | 0 |
| E2F2. | Other specific learning difficulty | 0 |
| E3... | Mental retardation | 0 |
| E30.. | Mild mental retardation, IQ in range 50-70 | 0 |
| E31.. | Other specified mental retardation | 0 |
| E310. | Moderate mental retardation, IQ in range 35-49 | 0 |
| E311. | Severe mental retardation, IQ in range 20-34 | 0 |
| E312. | Profound mental retardation with IQ less than 20 | 0 |
| E31z. | Other specified mental retardation NOS | 0 |
| E3y.. | Other specified mental retardation | 0 |
| E3z.. | Mental retardation NOS | 0 |
| Eu7.. | [X]Mental retardation | 0 |
| Eu70. | [X]Mild mental retardation | 0 |
| Eu700 | [X]Mild mental retardation with the statement of no, or minimal, impairment of behaviour | 0 |
| Eu701 | [X]Mild mental retardation, significant impairment of behaviour requiring attention or treatment | 0 |
| Eu70y | [X]Mild mental retardation, other impairments of behaviour | 0 |
| Eu70z | [X]Mild mental retardation without mention of impairment of behaviour | 0 |
| Eu71. | [X]Moderate mental retardation | 0 |
| Eu710 | [X]Moderate mental retardation with the statement of no, or minimal, impairment of behaviour | 0 |
| Eu711 | [X]Moderate mental retardation, significant impairment of behaviour requiring attention or treatment | 0 |
| Eu71y | [X]Mod retard oth behav impair | 0 |
| Eu71z | [X]Moderate mental retardation without mention of impairment of behaviour | 0 |
| Eu72. | [X]Severe mental retardation | 0 |
| Eu720 | [X]Severe mental retardation with the statement of no, or minimal, impairment of behaviour | 0 |
| Eu721 | [X]Severe mental retardation, significant impairment of behaviour requiring attention or treatment | 0 |
| Eu72y | [X]Severe mental retardation, other impairments of behaviour | 0 |
| Eu72z | [X]Severe mental retardation without mention of impairment of behaviour | 0 |
| Eu73. | [X]Profound mental retardation | 0 |
| Eu730 | [X]Profound mental retardation with the statement of no, or minimal, impairment of behaviour | 0 |
| Eu731 | [X]Profound mental retardation, significant impairment of behaviour requiring attention or treatment | 0 |
| Eu73y | [X]Profound mental retardation, other impairments of behaviour | 0 |
| Eu73z | [X]Profound mental retardation without mention of impairment of behaviour | 0 |
| Eu7y. | [X]Other mental retardation | 0 |
| Eu7y0 | [X]Other mental retardation with the statement of no, or minimal, impairment of behaviour | 0 |
| Eu7y1 | [X]Other mental retardation, significant impairment of behaviour requiring attention or treatment | 0 |
| Eu7yy | [X]Other mental retardation, other impairments of behaviour | 0 |
| Eu7yz | [X]Other mental retardation without mention of impairment of behaviour | 0 |
| Eu7z. | [X]Unspecified mental retardation | 0 |
| Eu7z0 | [X]Unspecified mental retardation with the statement of no, or minimal, impairment of behaviour | 0 |
| Eu7z1 | [X]Unspecified mental retardation, significant impairment of behaviour requiring attention or treatment | 0 |
| Eu7zy | [X]Unspecified mental retardation, other impairments of behaviour | 0 |
| Eu7zz | [X]Unspecified mental retardation without mention of impairment of behaviour | 0 |
| Eu814 | [X]Moderate learning disability | 0 |
| Eu815 | [X]Severe learning disability | 0 |
| Eu816 | [X]Mild learning disability | 0 |
| Eu817 | [X]Profound learning disability | 0 |
| Eu818 | [X]Specific learning disability | 0 |
| Eu81z | [X]Developmental disorder of scholastic skills, unspecified | 0 |

---

#### CONCEPT: Leukaemia¶

| READ\_CODE | DESCRIPTION\_SAIL | SENSITIVE\_CODE |
| --- | --- | --- |
| 1429. | H/O: \* leukaemia | 0 |
| B624. | Leukaemic reticuloendotheliosis | 0 |
| B64.. | Lymphoid leukaemia | 0 |
| B640. | Acute lymphoid leukaemia | 0 |
| B6400 | B-cell acute lymphoblastic leukaemia | 0 |
| B641. | Chronic lymphoid leukaemia | 0 |
| B6410 | B-cell chronic lymphocytic leukaemia | 0 |
| B6411 | Clinical stage A chronic lymphocytic leukaemia | 0 |
| B6412 | Clinical stage B chronic lymphocytic leukaemia | 0 |
| B6413 | Clinical stage C chronic lymphocytic leukaemia | 0 |
| B642. | Subacute lymphoid leukaemia | 0 |
| B64y. | Other lymphoid leukaemia | 0 |
| B64y0 | Aleukaemic lymphoid leukaemia | 0 |
| B64y1 | Prolymphocytic leukaemia | 0 |
| B64y2 | Adult T-cell leukaemia | 0 |
| B64y3 | B-cell prolymphocytic leukaemia | 0 |
| B64y4 | T-cell prolymphocytic leukaemia | 0 |
| B64y5 | Adult T-cell lymphoma/leukaemia (HTLV-1-associated) | 0 |
| B64yz | Other lymphoid leukaemia NOS | 0 |
| B64z. | Lymphoid leukaemia NOS | 0 |
| B65.. | Myeloid leukaemia | 0 |
| B650. | Acute myeloid leukaemia | 0 |
| B6500 | Acute myeloid leukaemia with 11q23 abnormality | 0 |
| B6501 | Acute myeloid leukaemia with multilineage dysplasia | 0 |
| B651. | Chronic myeloid leukaemia | 0 |
| B6510 | Chronic eosinophilic leukaemia | 0 |
| B6511 | Chronic myeloid leukaemia, BCR/ABL positive | 0 |
| B6512 | Chronic neutrophilic leukaemia | 0 |
| B6513 | Atypical chronic myeloid leukaemia, BCR/ABL negative | 0 |
| B651z | Chronic myeloid leukaemia NOS | 0 |
| B652. | Subacute myeloid leukaemia | 0 |
| B653. | Myeloid sarcoma | 0 |
| B6530 | Chloroma | 0 |
| B6531 | Granulocytic sarcoma | 0 |
| B653z | Myeloid sarcoma NOS | 0 |
| B654. | Acute myeloblastic leukaemia | 0 |
| B65y. | Other myeloid leukaemia | 0 |
| B65y0 | Aleukaemic myeloid leukaemia | 0 |
| B65y1 | Acute promyelocytic leukaemia | 0 |
| B65yz | Other myeloid leukaemia NOS | 0 |
| B65z. | Myeloid leukaemia NOS | 0 |
| B66.. | Monocytic leukaemia | 0 |
| B660. | Acute monocytic leukaemia | 0 |
| B661. | Chronic monocytic leukaemia | 0 |
| B662. | Subacute monocytic leukaemia | 0 |
| B663. | Acute monoblastic leukaemia | 0 |
| B66y. | Other monocytic leukaemia | 0 |
| B66y0 | Aleukaemic monocytic leukaemia | 0 |
| B66yz | Other monocytic leukaemia NOS | 0 |
| B66z. | Monocytic leukaemia NOS | 0 |
| B67.. | Other specified leukaemia | 0 |
| B670. | Acute erythraemia and erythroleukaemia | 0 |
| B671. | Chronic erythraemia | 0 |
| B672. | Megakaryocytic leukaemia | 0 |
| B673. | Mast cell leukaemia | 0 |
| B674. | Acute panmyelosis | 0 |
| B675. | Acute myelofibrosis | 0 |
| B676. | Acute erythroid leukaemia | 0 |
| B677. | Myelodysplastic and myeloproliferative disease | 0 |
| B67y. | Other and unspecified leukaemia | 0 |
| B67y0 | Lymphosarcoma cell leukaemia | 0 |
| B67yz | Other and unspecified leukaemia NOS | 0 |
| B67z. | Other specified leukaemia NOS | 0 |
| B68.. | Leukaemia of unspecified cell type | 0 |
| B680. | Acute leukaemia NOS | 0 |
| B681. | Chronic leukaemia NOS | 0 |
| B682. | Subacute leukaemia NOS | 0 |
| B68y. | Other leukaemia of unspecified cell type | 0 |
| B68z. | Leukaemia NOS | 0 |
| B69.. | Myelomonocytic leukaemia | 0 |
| B690. | Acute myelomonocytic leukaemia | 0 |
| B691. | Chronic myelomonocytic leukaemia | 0 |
| B692. | Subacute myelomonocytic leukaemia | 0 |
| B693. | Juvenile myelomonocytic leukaemia | 0 |
| B6y.. | Malignant neoplasm of lymphatic or haematopoietic tissue otherwise specified | 0 |
| B6y1. | Myelosclerosis with myeloid metaplasia | 0 |
| B6z.. | Malignant neoplasm lymphatic or haematopoietic tissue NOS | 0 |
| B937. | Neoplasm of uncertain behaviour of other lymphatic and haematopoietic tissue | 0 |
| BBr.. | [M]Leukaemias | 0 |
| BBr0. | [M]Leukaemias unspecified | 0 |
| BBr00 | [M]Leukaemia NOS | 0 |
| BBr01 | [M]Acute leukaemia NOS | 0 |
| BBr02 | [M]Subacute leukaemia NOS | 0 |
| BBr03 | [M]Chronic leukaemia NOS | 0 |
| BBr04 | [M]Aleukaemic leukaemia NOS | 0 |
| BBr0z | [M]Leukaemia unspecified, NOS | 0 |
| BBr1. | [M]Compound leukaemias | 0 |
| BBr10 | [M]Compound leukaemia | 0 |
| BBr1z | [M]Compound leukaemia NOS | 0 |
| BBr2. | [M]Lymphoid leukaemias | 0 |
| BBr20 | [M]Lymphoid leukaemia NOS | 0 |
| BBr21 | [M]Acute lymphoid leukaemia | 0 |
| BBr22 | [M]Subacute lymphoid leukaemia | 0 |
| BBr23 | [M]Chronic lymphoid leukaemia | 0 |
| BBr24 | [M]Aleukaemic lymphoid leukaemia | 0 |
| BBr25 | [M]Prolymphocytic leukaemia | 0 |
| BBr26 | [M]Burkitt's cell leukaemia | 0 |
| BBr2z | [M]Other lymphoid leukaemia NOS | 0 |
| BBr4. | [M]Erythroleukaemias | 0 |
| BBr40 | [M]Erythroleukaemia | 0 |
| BBr4z | [M]Erythroleukaemia NOS | 0 |
| BBr5. | [M]Lymphosarcoma cell leukaemias | 0 |
| BBr50 | [M]Lymphosarcoma cell leukaemia | 0 |
| BBr5z | [M]Lymphosarcoma cell leukaemia NOS | 0 |
| BBr6. | [M]Myeloid leukaemias | 0 |
| BBr60 | [M]Myeloid leukaemia NOS | 0 |
| BBr61 | [M]Acute myeloid leukaemia | 0 |
| BBr62 | [M]Subacute myeloid leukaemia | 0 |
| BBr63 | [M]Chronic myeloid leukaemia | 0 |
| BBr64 | [M]Aleukaemic myeloid leukaemia | 0 |
| BBr65 | [M]Neutrophilic leukaemia | 0 |
| BBr66 | [M]Acute promyelocytic leukaemia | 0 |
| BBr67 | [M]Acute myelomonocytic leukaemia | 0 |
| BBr68 | [M]Chronic myelomonocytic leukaemia | 0 |
| BBr69 | [M]Juvenile myelomonocytic leukaemia | 0 |
| BBr6z | [M]Other myeloid leukaemia NOS | 0 |
| BBr7. | [M]Basophilic leukaemias | 0 |
| BBr70 | [M]Basophilic leukaemia | 0 |
| BBr7z | [M]Basophilic leukaemia NOS | 0 |
| BBr8. | [M]Eosinophilic leukaemias | 0 |
| BBr80 | [M]Eosinophilic leukaemia | 0 |
| BBr8z | [M]Eosinophilic leukaemia NOS | 0 |
| BBr9. | [M]Monocytic leukaemias | 0 |
| BBr90 | [M]Monocytic leukaemia NOS | 0 |
| BBr91 | [M]Acute monocytic leukaemia | 0 |
| BBr92 | [M]Subacute monocytic leukaemia | 0 |
| BBr93 | [M]Chronic monocytic leukaemia | 0 |
| BBr94 | [M]Aleukaemic monocytic leukaemia | 0 |
| BBr9z | [M]Other monocytic leukaemia NOS | 0 |
| BBrA. | [M]Miscellaneous leukaemias | 0 |
| BBrA0 | [M]Mast cell leukaemia | 0 |
| BBrA1 | [M]Megakaryocytic leukaemia | 0 |
| BBrA3 | [M]Myeloid sarcoma | 0 |
| BBrA4 | [M]Hairy cell leukaemia | 0 |
| BBrA5 | [M]Acute megakaryoblastic leukaemia | 0 |
| BBrA6 | [M]Acute panmyelosis | 0 |
| BBrA7 | [M]Acute myelofibrosis | 0 |
| BBrAz | [M]Miscellaneous leukaemia NOS | 0 |
| BBrz. | [M]Leukaemia NOS | 0 |
| BBs1. | [M]Acute panmyelosis | 0 |
| BBs3. | [M]Myelosclerosis with myeloid metaplasia | 0 |
| ByuD. | [X]Malignant neoplasms of lymphoid, haematopoietic and related tissue | 0 |
| ByuD5 | [X]Other lymphoid leukaemia | 0 |
| ByuD6 | [X]Other myeloid leukaemia | 0 |
| ByuD7 | [X]Other monocytic leukaemia | 0 |
| ByuD8 | [X]Other specified leukaemias | 0 |
| ByuD9 | [X]Other leukaemia of unspecified cell type | 0 |
| D41y1 | Myelofibrosis | 0 |
| ZV106 | [V]Personal history of leukaemia | 0 |
| ZV678 | [V]Follow-up examination after chemotherapy for malignant neoplasm | 0 |

---

#### CONCEPT: Lymphoma¶

| READ\_CODE | DESCRIPTION\_SAIL | SENSITIVE\_CODE |
| --- | --- | --- |
| 4M20. | Lymphoma stage I | 0 |
| 4M21. | Lymphoma stage II | 0 |
| 4M22. | Lymphoma stage III | 0 |
| 4M23. | Lymphoma stage IV | 0 |
| A7896 | HIV disease resulting in Burkitt's lymphoma | 1 |
| A7897 | HIV disease resulting in other types of non-Hodgkin's lymphoma | 1 |
| AyuC6 | [X]HIV disease resulting in other non-Hodgkin's lymphoma | 1 |
| B5601 | Secondary and unspecified malignant neoplasm of mastoid lymph nodes | 0 |
| B6... | Malignant neoplasm of lymphatic and haemopoietic tissue | 0 |
| B60.. | Lymphosarcoma and reticulosarcoma | 0 |
| B600. | Reticulosarcoma | 0 |
| B6000 | Reticulosarcoma of unspecified site | 0 |
| B6001 | Reticulosarcoma of lymph nodes of head, face and neck | 0 |
| B6002 | Reticulosarcoma of intrathoracic lymph nodes | 0 |
| B6003 | Reticulosarcoma of intra-abdominal lymph nodes | 0 |
| B6004 | Reticulosarcoma of lymph nodes of axilla and upper limb | 0 |
| B6005 | Reticulosarcoma of lymph nodes of inguinal region and lower limb | 0 |
| B6006 | Reticulosarcoma of intrapelvic lymph nodes | 0 |
| B6007 | Reticulosarcoma of spleen | 0 |
| B6008 | Reticulosarcoma of lymph nodes of multiple sites | 0 |
| B600z | Reticulosarcoma NOS | 0 |
| B601. | Lymphosarcoma | 0 |
| B6010 | Lymphosarcoma of unspecified site | 0 |
| B6011 | Lymphosarcoma of lymph nodes of head, face and neck | 0 |
| B6012 | Lymphosarcoma of intrathoracic lymph nodes | 0 |
| B6013 | Lymphosarcoma of intra-abdominal lymph nodes | 0 |
| B6014 | Lymphosarcoma of lymph nodes of axilla and upper limb | 0 |
| B6015 | Lymphosarcoma of lymph nodes of inguinal region and lower limb | 0 |
| B6016 | Lymphosarcoma of intrapelvic lymph nodes | 0 |
| B6017 | Lymphosarcoma of spleen | 0 |
| B6018 | Lymphosarcoma of lymph nodes of multiple sites | 0 |
| B601z | Lymphosarcoma NOS | 0 |
| B602. | Burkitt's lymphoma | 0 |
| B6020 | Burkitt's lymphoma of unspecified site | 0 |
| B6021 | Burkitt's lymphoma of lymph nodes of head, face and neck | 0 |
| B6022 | Burkitt's lymphoma of intrathoracic lymph nodes | 0 |
| B6023 | Burkitt's lymphoma of intra-abdominal lymph nodes | 0 |
| B6024 | Burkitt's lymphoma of lymph nodes of axilla and upper limb | 0 |
| B6025 | Burkitt's lymphoma of lymph nodes of inguinal region and lower limb | 0 |
| B6026 | Burkitt's lymphoma of intrapelvic lymph nodes | 0 |
| B6027 | Burkitt's lymphoma of spleen | 0 |
| B6028 | Burkitt's lymphoma of lymph nodes of multiple sites | 0 |
| B602z | Burkitt's lymphoma NOS | 0 |
| B60y. | Other specified reticulosarcoma or lymphosarcoma | 0 |
| B60z. | Reticulosarcoma or lymphosarcoma NOS | 0 |
| B61.. | Hodgkin's disease | 0 |
| B610. | Hodgkin's paragranuloma | 0 |
| B6100 | Hodgkin's paragranuloma of unspecified site | 0 |
| B6101 | Hodgkin's paragranuloma of lymph nodes of head, face, and neck | 0 |
| B6102 | Hodgkin's paragranuloma of intrathoracic lymph nodes | 0 |
| B6103 | Hodgkin's paragranuloma of intra-abdominal lymph nodes | 0 |
| B6104 | Hodgkin's paragranuloma of lymph nodes of axilla and upper limb | 0 |
| B6105 | Hodgkin's paragranuloma of lymph nodes of inguinal region and lower limb | 0 |
| B6106 | Hodgkin's paragranuloma of intrapelvic lymph nodes | 0 |
| B6107 | Hodgkin's paragranuloma of spleen | 0 |
| B6108 | Hodgkin's paragranuloma of lymph nodes of multiple sites | 0 |
| B610z | Hodgkin's paragranuloma NOS | 0 |
| B611. | Hodgkin's granuloma | 0 |
| B6110 | Hodgkin's granuloma of unspecified site | 0 |
| B6111 | Hodgkin's granuloma of lymph nodes of head, face and neck | 0 |
| B6112 | Hodgkin's granuloma of intrathoracic lymph nodes | 0 |
| B6113 | Hodgkin's granuloma of intra-abdominal lymph nodes | 0 |
| B6114 | Hodgkin's granuloma of lymph nodes of axilla and upper limb | 0 |
| B6115 | Hodgkin's granuloma of lymph nodes of inguinal region and lower limb | 0 |
| B6116 | Hodgkin's granuloma of intrapelvic lymph nodes | 0 |
| B6117 | Hodgkin's granuloma of spleen | 0 |
| B6118 | Hodgkin's granuloma of lymph nodes of multiple sites | 0 |
| B611z | Hodgkin's granuloma NOS | 0 |
| B612. | Hodgkin's sarcoma | 0 |
| B6120 | Hodgkin's sarcoma of unspecified site | 0 |
| B6121 | Hodgkin's sarcoma of lymph nodes of head, face and neck | 0 |
| B6122 | Hodgkin's sarcoma of intrathoracic lymph nodes | 0 |
| B6123 | Hodgkin's sarcoma of intra-abdominal lymph nodes | 0 |
| B6124 | Hodgkin's sarcoma of lymph nodes of axilla and upper limb | 0 |
| B6125 | Hodgkin's sarcoma of lymph nodes of inguinal region and lower limb | 0 |
| B6126 | Hodgkin's sarcoma of intrapelvic lymph nodes | 0 |
| B6127 | Hodgkin's sarcoma of spleen | 0 |
| B6128 | Hodgkin's sarcoma of lymph nodes of multiple sites | 0 |
| B612z | Hodgkin's sarcoma NOS | 0 |
| B613. | Hodgkin's disease, lymphocytic-histiocytic predominance | 0 |
| B6130 | Hodgkin's disease, lymphocytic-histiocytic predominance of unspecified site | 0 |
| B6131 | Hodgkin's disease, lymphocytic-histiocytic predominance of lymph nodes of head, face and neck | 0 |
| B6132 | Hodgkin's disease, lymphocytic-histiocytic predominance of intrathoracic lymph nodes | 0 |
| B6133 | Hodgkin's disease, lymphocytic-histiocytic predominance of intra-abdominal lymph nodes | 0 |
| B6134 | Hodgkin's disease, lymphocytic-histiocytic predominance of lymph nodes of axilla and upper limb | 0 |
| B6135 | Hodgkin's disease, lymphocytic-histiocytic predominance of lymph nodes of inguinal region and lower limb | 0 |
| B6136 | Hodgkin's disease, lymphocytic-histiocytic predominance of intrapelvic lymph nodes | 0 |
| B6137 | Hodgkin's, lymphocytic-histiocytic predominance of spleen | 0 |
| B6138 | Hodgkin's disease, lymphocytic-histiocytic predominance of lymph nodes of multiple sites | 0 |
| B613z | Hodgkin's, lymphocytic-histiocytic predominance NOS | 0 |
| B614. | Hodgkin's disease, nodular sclerosis | 0 |
| B6140 | Hodgkin's disease, nodular sclerosis of unspecified site | 0 |
| B6141 | Hodgkin's disease, nodular sclerosis of lymph nodes of head, face and neck | 0 |
| B6142 | Hodgkin's disease, nodular sclerosis of intrathoracic lymph nodes | 0 |
| B6143 | Hodgkin's disease, nodular sclerosis of intra-abdominal lymph nodes | 0 |
| B6144 | Hodgkin's disease, nodular sclerosis of lymph nodes of axilla and upper limb | 0 |
| B6145 | Hodgkin's disease, nodular sclerosis of lymph nodes of inguinal region and lower limb | 0 |
| B6146 | Hodgkin's disease, nodular sclerosis of intrapelvic lymph nodes | 0 |
| B6147 | Hodgkin's disease, nodular sclerosis of spleen | 0 |
| B6148 | Hodgkin's disease, nodular sclerosis of lymph nodes of multiple sites | 0 |
| B614z | Hodgkin's disease, nodular sclerosis NOS | 0 |
| B615. | Hodgkin's disease, mixed cellularity | 0 |
| B6150 | Hodgkin's disease, mixed cellularity of unspecified site | 0 |
| B6151 | Hodgkin's disease, mixed cellularity of lymph nodes of head, face and neck | 0 |
| B6152 | Hodgkin's disease, mixed cellularity of intrathoracic lymph nodes | 0 |
| B6153 | Hodgkin's disease, mixed cellularity of intra-abdominal lymph nodes | 0 |
| B6154 | Hodgkin's disease, mixed cellularity of lymph nodes of axilla and upper limb | 0 |
| B6155 | Hodgkin's disease, mixed cellularity of lymph nodes of inguinal region and lower limb | 0 |
| B6156 | Hodgkin's disease, mixed cellularity of intrapelvic lymph nodes | 0 |
| B6157 | Hodgkin's disease, mixed cellularity of spleen | 0 |
| B6158 | Hodgkin's disease, mixed cellularity of lymph nodes of multiple sites | 0 |
| B615z | Hodgkin's disease, mixed cellularity NOS | 0 |
| B616. | Hodgkin's disease, lymphocytic depletion | 0 |
| B6160 | Hodgkin's lymphocytic depletion of unspecified site | 0 |
| B6161 | Hodgkin's disease, lymphocytic depletion of lymph nodes of head, face and neck | 0 |
| B6162 | Hodgkin's disease, lymphocytic depletion of intrathoracic lymph nodes | 0 |
| B6163 | Hodgkin's disease, lymphocytic depletion of intra-abdominal lymph nodes | 0 |
| B6164 | Hodgkin's disease, lymphocytic depletion of lymph nodes of axilla and upper limb | 0 |
| B6165 | Hodgkin's disease, lymphocytic depletion of lymph nodes of inguinal region and lower limb | 0 |
| B6166 | Hodgkin's disease, lymphocytic depletion of intrapelvic lymph nodes | 0 |
| B6167 | Hodgkin's disease, lymphocytic depletion of spleen | 0 |
| B6168 | Hodgkin's disease, lymphocytic depletion of lymph nodes of multiple sites | 0 |
| B616z | Hodgkin's disease, lymphocytic depletion NOS | 0 |
| B617. | Nodular lymphocyte predominant Hodgkin lymphoma | 0 |
| B618. | Nodular sclerosis classical Hodgkin lymphoma | 0 |
| B619. | Mixed cellularity classical Hodgkin lymphoma | 0 |
| B61A. | Lymphocyte depleted classical Hodgkin lymphoma | 0 |
| B61B. | Lymphocyte-rich classical Hodgkin lymphoma | 0 |
| B61C. | Other classical Hodgkin lymphoma | 0 |
| B61z. | Hodgkin's disease NOS | 0 |
| B61z0 | Hodgkin's disease NOS, unspecified site | 0 |
| B61z1 | Hodgkin's disease NOS of lymph nodes of head, face and neck | 0 |
| B61z2 | Hodgkin's disease NOS of intrathoracic lymph nodes | 0 |
| B61z3 | Hodgkin's disease NOS of intra-abdominal lymph nodes | 0 |
| B61z4 | Hodgkin's disease NOS of lymph nodes of axilla and upper limb | 0 |
| B61z5 | Hodgkin's disease NOS of lymph nodes of inguinal region and lower limb | 0 |
| B61z6 | Hodgkin's disease NOS of intrapelvic lymph nodes | 0 |
| B61z7 | Hodgkin's disease NOS of spleen | 0 |
| B61z8 | Hodgkin's disease NOS of lymph nodes of multiple sites | 0 |
| B61zz | Hodgkin's disease NOS | 0 |
| B62.. | Other malignant neoplasm of lymphoid and histiocytic tissue | 0 |
| B620. | Nodular lymphoma (Brill - Symmers disease) | 0 |
| B6200 | Nodular lymphoma of unspecified site | 0 |
| B6201 | Nodular lymphoma of lymph nodes of head, face and neck | 0 |
| B6202 | Nodular lymphoma of intrathoracic lymph nodes | 0 |
| B6203 | Nodular lymphoma of intra-abdominal lymph nodes | 0 |
| B6204 | Nodular lymphoma of lymph nodes of axilla and upper limb | 0 |
| B6205 | Nodular lymphoma of lymph nodes of inguinal region and lower limb | 0 |
| B6206 | Nodular lymphoma of intrapelvic lymph nodes | 0 |
| B6207 | Nodular lymphoma of spleen | 0 |
| B6208 | Nodular lymphoma of lymph nodes of multiple sites | 0 |
| B620z | Nodular lymphoma NOS | 0 |
| B621. | Mycosis fungoides | 0 |
| B6210 | Mycosis fungoides of unspecified site | 0 |
| B6211 | Mycosis fungoides of the lymph nodes of head, face and neck | 0 |
| B6212 | Mycosis fungoides of intrathoracic lymph nodes | 0 |
| B6213 | Mycosis fungoides of intra-abdominal lymph nodes | 0 |
| B6214 | Mycosis fungoides of lymph nodes of axilla and upper limb | 0 |
| B6215 | Mycosis fungoides of lymph nodes of inguinal region and lower limb | 0 |
| B6216 | Mycosis fungoides of intrapelvic lymph nodes | 0 |
| B6217 | Mycosis fungoides of spleen | 0 |
| B6218 | Mycosis fungoides of lymph nodes of multiple sites | 0 |
| B621z | Mycosis fungoides NOS | 0 |
| B622. | Sezary's disease | 0 |
| B6220 | Sezary's disease of unspecified site | 0 |
| B6221 | Sezary's disease of lymph nodes of head, face and neck | 0 |
| B6222 | Sezary's disease of intrathoracic lymph nodes | 0 |
| B6223 | Sezary's disease of intra-abdominal lymph nodes | 0 |
| B6224 | Sezary's disease of lymph nodes of axilla and upper limb | 0 |
| B6225 | Sezary's disease of lymph nodes of inguinal region and lower limb | 0 |
| B6226 | Sezary's disease of intrapelvic lymph nodes | 0 |
| B6227 | Sezary's disease of spleen | 0 |
| B6228 | Sezary's disease of lymph nodes of multiple sites | 0 |
| B622z | Sezary's disease NOS | 0 |
| B623. | Malignant histiocytosis | 0 |
| B6230 | Malignant histiocytosis of unspecified site | 0 |
| B6231 | Malignant histiocytosis of lymph nodes of head, face and neck | 0 |
| B6232 | Malignant histiocytosis of intrathoracic lymph nodes | 0 |
| B6233 | Malignant histiocytosis of intra-abdominal lymph nodes | 0 |
| B6234 | Malignant histiocytosis of lymph nodes of axilla and upper limb | 0 |
| B6235 | Malignant histiocytosis of lymph nodes of inguinal region and lower limb | 0 |
| B6236 | Malignant histiocytosis of intrapelvic lymph nodes | 0 |
| B6237 | Malignant histiocytosis of spleen | 0 |
| B6238 | Malignant histiocytosis of lymph nodes of multiple sites | 0 |
| B623z | Malignant histiocytosis NOS | 0 |
| B624. | Leukaemic reticuloendotheliosis | 0 |
| B6240 | Leukaemic reticuloendotheliosis of unspecified sites | 0 |
| B6241 | Leukaemic reticuloendotheliosis of lymph nodes of head, face and neck | 0 |
| B6242 | Leukaemic reticuloendotheliosis of intrathoracic lymph nodes | 0 |
| B6243 | Leukaemic reticuloendotheliosis of intra-abdominal lymph nodes | 0 |
| B6244 | Leukaemic reticuloendotheliosis of lymph nodes of axilla and upper limb | 0 |
| B6245 | Leukaemic reticuloendotheliosis of lymph nodes of inguinal region and lower limb | 0 |
| B6246 | Leukaemic reticuloendotheliosis of intrapelvic lymph nodes | 0 |
| B6247 | Leukaemic reticuloendotheliosis of spleen | 0 |
| B6248 | Leukaemic reticuloendotheliosis of lymph nodes of multiple sites | 0 |
| B624z | Leukaemic reticuloendotheliosis NOS | 0 |
| B625. | Letterer-Siwe disease | 0 |
| B6250 | Letterer-Siwe disease of unspecified sites | 0 |
| B6251 | Letterer-Siwe disease of lymph nodes of head, face and neck | 0 |
| B6252 | Letterer-Siwe disease of intrathoracic lymph nodes | 0 |
| B6253 | Letterer-Siwe disease of intra-abdominal lymph nodes | 0 |
| B6254 | Letterer-Siwe disease of lymph nodes of axilla and upper limb | 0 |
| B6255 | Letterer-Siwe disease of lymph nodes of inguinal region and lower limb | 0 |
| B6256 | Letterer-Siwe disease of intrapelvic lymph nodes | 0 |
| B6257 | Letterer-Siwe disease of spleen | 0 |
| B6258 | Letterer-Siwe disease of lymph nodes of multiple sites | 0 |
| B625z | Letterer-Siwe disease NOS | 0 |
| B626. | Malignant mast cell tumours | 0 |
| B6260 | Mast cell malignancy of unspecified site | 0 |
| B6261 | Mast cell malignancy of lymph nodes of head, face and neck | 0 |
| B6262 | Mast cell malignancy of intrathoracic lymph nodes | 0 |
| B6263 | Mast cell malignancy of intra-abdominal lymph nodes | 0 |
| B6264 | Mast cell malignancy of lymph nodes of axilla and upper limb | 0 |
| B6265 | Mast cell malignancy of lymph nodes of inguinal region and lower limb | 0 |
| B6266 | Mast cell malignancy of intrapelvic lymph nodes | 0 |
| B6267 | Mast cell malignancy of spleen | 0 |
| B6268 | Mast cell malignancy of lymph nodes of multiple sites | 0 |
| B626z | Malignant mast cell tumour NOS | 0 |
| B627. | Non - Hodgkin's lymphoma | 0 |
| B6270 | Follicular non-Hodgkin's small cleaved cell lymphoma | 0 |
| B6271 | Follicular non-Hodgkin's mixed small cleaved and large cell lymphoma | 0 |
| B6272 | Follicular non-Hodgkin's large cell lymphoma | 0 |
| B6273 | Diffuse non-Hodgkin's small cell (diffuse) lymphoma | 0 |
| B6274 | Diffuse non-Hodgkin's small cleaved cell (diffuse) lymphoma | 0 |
| B6275 | Diffuse non-Hodgkin's mixed small and large cell (diffuse) lymphoma | 0 |
| B6276 | Diffuse non-Hodgkin's immunoblastic (diffuse) lymphoma | 0 |
| B6277 | Diffuse non-Hodgkin's lymphoblastic (diffuse) lymphoma | 0 |
| B6278 | Diffuse non-Hodgkin's lymphoma undifferentiated (diffuse) | 0 |
| B6279 | Mucosa-associated lymphoma | 0 |
| B627A | Diffuse non-Hodgkin's large cell lymphoma | 0 |
| B627B | Other types of follicular non-Hodgkin's lymphoma | 0 |
| B627C | Follicular non-Hodgkin's lymphoma | 0 |
| B627D | Diffuse non-Hodgkin's centroblastic lymphoma | 0 |
| B627E | Diffuse large B-cell lymphoma | 0 |
| B627F | Extranodal marginal zone B-cell lymphoma of mucosa-associated lymphoid tissue | 0 |
| B627G | Mediastinal (thymic) large B-cell lymphoma | 0 |
| B627W | Unspecified B-cell non-Hodgkin's lymphoma | 0 |
| B627X | Diffuse non-Hodgkin's lymphoma, unspecified | 0 |
| B628. | Follicular lymphoma | 0 |
| B6280 | Follicular lymphoma grade 1 | 0 |
| B6281 | Follicular lymphoma grade 2 | 0 |
| B6282 | Follicular lymphoma grade 3 | 0 |
| B6283 | Follicular lymphoma grade 3a | 0 |
| B6284 | Follicular lymphoma grade 3b | 0 |
| B6285 | Diffuse follicle centre lymphoma | 0 |
| B6286 | Cutaneous follicle centre lymphoma | 0 |
| B6287 | Other types of follicular lymphoma | 0 |
| B629. | Multifocal and multisystemic (disseminated) Langerhans-cell histiocytosis | 0 |
| B62A. | Sarcoma of dendritic cells | 0 |
| B62B. | Multifocal and unisystemic Langerhans-cell histiocytosis | 0 |
| B62C. | Unifocal Langerhans-cell histiocytosis | 0 |
| B62D. | Histiocytic sarcoma | 0 |
| B62E. | T/NK-cell lymphoma | 0 |
| B62E0 | Mature T/NK-cell lymphoma | 0 |
| B62E1 | Anaplastic large cell lymphoma, ALK-positive | 0 |
| B62E2 | Anaplastic large cell lymphoma, ALK-negative | 0 |
| B62E3 | Cutaneous T-cell lymphoma | 0 |
| B62E4 | Extranodal NK/T-cell lymphoma, nasal type | 0 |
| B62E5 | Hepatosplenic T-cell lymphoma | 0 |
| B62E6 | Enteropathy-associated T-cell lymphoma | 0 |
| B62E7 | Subcutaneous panniculitic T-cell lymphoma | 0 |
| B62E8 | Blastic NK-cell lymphoma | 0 |
| B62E9 | Angioimmunoblastic T-cell lymphoma | 0 |
| B62EA | Primary cutaneous CD30-positive T-cell proliferations | 0 |
| B62Ew | Other mature T/NK-cell lymphoma | 0 |
| B62F. | Nonfollicular lymphoma | 0 |
| B62F0 | Small cell B-cell lymphoma | 0 |
| B62F1 | Mantle cell lymphoma | 0 |
| B62F2 | Lymphoblastic (diffuse) lymphoma | 0 |
| B62Fy | Other non-follicular lymphoma | 0 |
| B62x. | Malignant lymphoma otherwise specified | 0 |
| B62x0 | T-zone lymphoma | 0 |
| B62x1 | Lymphoepithelioid lymphoma | 0 |
| B62x2 | Peripheral T-cell lymphoma | 0 |
| B62x3 | Malignant reticuloendotheliosis | 0 |
| B62x4 | Malignant reticulosis | 0 |
| B62x5 | Malignant immunoproliferative small intestinal disease | 0 |
| B62x6 | True histiocytic lymphoma | 0 |
| B62xX | Other and unspecified peripheral and cutaneous T-cell lymphomas | 0 |
| B62y. | Malignant lymphoma NOS | 0 |
| B62y0 | Malignant lymphoma NOS of unspecified site | 0 |
| B62y1 | Malignant lymphoma NOS of lymph nodes of head, face and neck | 0 |
| B62y2 | Malignant lymphoma NOS of intrathoracic lymph nodes | 0 |
| B62y3 | Malignant lymphoma NOS of intra-abdominal lymph nodes | 0 |
| B62y4 | Malignant lymphoma NOS of lymph nodes of axilla and upper limb | 0 |
| B62y5 | Malignant lymphoma NOS of lymph nodes of inguinal region and lower limb | 0 |
| B62y6 | Malignant lymphoma NOS of intrapelvic lymph nodes | 0 |
| B62y7 | Malignant lymphoma NOS of spleen | 0 |
| B62y8 | Malignant lymphoma NOS of lymph nodes of multiple sites | 0 |
| B62yz | Malignant lymphoma NOS | 0 |
| B62z. | Malignant neoplasms of lymphoid and histiocytic tissue NOS | 0 |
| B62z0 | Unspecified malignant neoplasm of lymphoid and histiocytic tissue of unspecified site | 0 |
| B62z1 | Unspecified malignant neoplasm of lymphoid and histiocytic tissue of lymph nodes of head, face and neck | 0 |
| B62z2 | Unspecified malignant neoplasm of lymphoid and histiocytic tissue of intrathoracic lymph nodes | 0 |
| B62z3 | Unspecified malignant neoplasm of lymphoid and histiocytic tissue of intra-abdominal lymph nodes | 0 |
| B62z4 | Unspecified malignant neoplasm of lymphoid and histiocytic tissue of lymph nodes of axilla and upper limb | 0 |
| B62z5 | Unspecified malignant neoplasm of lymphoid and histiocytic tissue of lymph nodes of inguinal region and lower limb | 0 |
| B62z6 | Unspecified malignant neoplasm of lymphoid and histiocytic tissue of intrapelvic lymph nodes | 0 |
| B62z7 | Unspecified malignant neoplasm of lymphoid and histiocytic tissue of spleen | 0 |
| B62z8 | Unspecified malignant neoplasm of lymphoid and histiocytic tissue of lymph nodes of multiple sites | 0 |
| B62zz | Lymphoid and histiocytic malignancy NOS | 0 |
| B63y. | Other immunoproliferative neoplasms | 0 |
| B64y5 | Adult T-cell lymphoma/leukaemia (HTLV-1-associated) | 0 |
| B67y0 | Lymphosarcoma cell leukaemia | 0 |
| B6y.. | Malignant neoplasm of lymphatic or haematopoietic tissue otherwise specified | 0 |
| B6z.. | Malignant neoplasm lymphatic or haematopoietic tissue NOS | 0 |
| BBGF. | [M]Fibrous histiocytoma, malignant | 0 |
| BBg.. | [M]Lymphomas, NOS or diffuse | 0 |
| BBg1. | [M]Malignant lymphoma NOS | 0 |
| BBg10 | [M]Malignant lymphoma, diffuse NOS | 0 |
| BBg2. | [M]Malignant lymphoma, non Hodgkin's type | 0 |
| BBg3. | [M]Malignant lymphoma, undifferentiated cell type NOS | 0 |
| BBg4. | [M]Malignant lymphoma, stem cell type | 0 |
| BBg5. | [M]Malignant lymphoma, convoluted cell type NOS | 0 |
| BBg6. | [M]Lymphosarcoma NOS | 0 |
| BBg7. | [M]Malignant lymphoma, lymphoplasmacytoid type | 0 |
| BBg8. | [M]Malignant lymphoma, immunoblastic type | 0 |
| BBg9. | [M]Malignant lymphoma, mixed lymphocytic-histiocytic NOS | 0 |
| BBgA. | [M]Malignant lymphoma, centroblastic-centrocytic, diffuse | 0 |
| BBgB. | [M]Malignant lymphoma, follicular centre cell NOS | 0 |
| BBgC. | [M]Malignant lymphoma, lymphocytic, well differentiated NOS | 0 |
| BBgD. | [M]Malignant lymphoma, lymphocytic, intermediate differentiation NOS | 0 |
| BBgE. | [M]Malignant lymphoma, centrocytic | 0 |
| BBgF. | [M]Malignant lymphoma, follicular centre cell, cleaved NOS | 0 |
| BBgG. | [M]Malignant lymphoma, lymphocytic, poorly differentiated NOS | 0 |
| BBgH. | [M]Prolymphocytic lymphosarcoma | 0 |
| BBgJ. | [M]Malignant lymphoma, centroblastic type NOS | 0 |
| BBgK. | [M]Malignant lymphoma, follicular centre cell, non-cleaved NOS | 0 |
| BBgL. | [M]Malignant lymphoma, small lymphocytic NOS | 0 |
| BBgM. | [M]Malignant lymphoma, small cleaved cell, diffuse | 0 |
| BBgN. | [M]Malignant lymphoma, lymphocytic, intermediate differentiation, diffuse | 0 |
| BBgP. | [M]Malignant lymphoma, mixed small and large cell, diffuse | 0 |
| BBgQ. | [M]Malignant lymphomatous polyposis | 0 |
| BBgR. | [M]Malignant lymphoma, large cell, diffuse NOS | 0 |
| BBgS. | [M]Malignant lymphoma, large cell, cleaved, diffuse | 0 |
| BBgT. | [M]Malignant lymphoma, large cell, noncleaved, diffuse | 0 |
| BBgV. | [M]Malignant lymphoma, small cell, noncleaved, diffuse | 0 |
| BBgz. | [M]Lymphoma, diffuse or NOS | 0 |
| BBh.. | [M]Reticulosarcomas | 0 |
| BBh0. | [M]Reticulosarcoma NOS | 0 |
| BBh1. | [M]Reticulosarcoma, pleomorphic cell type | 0 |
| BBh2. | [M]Reticulosarcoma, nodular | 0 |
| BBhz. | [M]Reticulosarcoma NOS | 0 |
| BBj.. | [M]Hodgkin's disease | 0 |
| BBj0. | [M]Hodgkin's disease NOS | 0 |
| BBj1. | [M]Hodgkin's disease, lymphocytic predominance | 0 |
| BBj10 | [M]Hodgkin,s disease, lymphocytic predominance, diffuse | 0 |
| BBj11 | [M]Hodgkin,s disease, lymphocytic predominance, nodular | 0 |
| BBj2. | [M]Hodgkin's disease, mixed cellularity | 0 |
| BBj3. | [M]Hodgkin's disease, lymphocytic depletion NOS | 0 |
| BBj4. | [M]Hodgkin's disease, lymphocytic depletion, diffuse fibrosis | 0 |
| BBj5. | [M]Hodgkin's disease, lymphocytic depletion, reticular type | 0 |
| BBj6. | [M]Hodgkin's disease, nodular sclerosis NOS | 0 |
| BBj60 | [M]Hodgkin,s disease, nodular sclerosis, lymphocytic predominance | 0 |
| BBj61 | [M]Hodgkin,s disease, nodular sclerosis, mixed cellularity | 0 |
| BBj62 | [M]Hodgkin,s disease, nodular sclerosis, lymphocytic depletion | 0 |
| BBj7. | [M]Hodgkin's disease, nodular sclerosis, cellular phase | 0 |
| BBj8. | [M]Hodgkin's paragranuloma | 0 |
| BBj9. | [M]Hodgkin's granuloma | 0 |
| BBjA. | [M]Hodgkin's sarcoma | 0 |
| BBjz. | [M]Hodgkin's disease NOS | 0 |
| BBk.. | [M]Lymphomas, nodular or follicular | 0 |
| BBk0. | [M]Malignant lymphoma, nodular NOS | 0 |
| BBk1. | [M]Malignant lymphoma, mixed lymphocytic-histiocytic, nodular | 0 |
| BBk2. | [M]Malignant lymphoma, centroblastic-centrocytic, follicular | 0 |
| BBk3. | [M]Malignant lymphoma, lymphocytic, well differentiated, nodular | 0 |
| BBk4. | [M]Malignant lymphoma, lymphocytic, intermediate differentiation, nodular | 0 |
| BBk5. | [M]Malignant lymphoma, follicular centre cell, cleaved, follicular | 0 |
| BBk6. | [M]Malignant lymphoma, lymphocytic, poorly differentiated, nodular | 0 |
| BBk7. | [M]Malignant lymphoma, centroblastic type, follicular | 0 |
| BBkz. | [M]Lymphoma, nodular or follicular NOS | 0 |
| BBl.. | [M]Mycosis fungoides | 0 |
| BBl0. | [M]Mycosis fungoides | 0 |
| BBl1. | [M]Sezary's disease | 0 |
| BBlz. | [M]Mycosis fungoides NOS | 0 |
| BBm1. | [M]Malignant histiocytosis | 0 |
| BBm3. | [M]Letterer - Siwe disease | 0 |
| BBm4. | [M]True histiocytic lymphoma | 0 |
| BBm5. | [M] Peripheral T-cell lymphoma NOS | 0 |
| BBm9. | [M] Monocytoid B-cell lymphoma | 0 |
| BBmD. | [M] Cutaneous lymphoma | 0 |
| BBmG. | [M] Immunoproliferative small intestinal disease | 0 |
| BBmH. | [M] Large cell lymphoma | 0 |
| BBmK. | [M]Waldenstrom's macroglobulinaemia | 0 |
| BBp1. | [M]Mast cell sarcoma | 0 |
| BBp2. | [M]Malignant mastocytosis | 0 |
| BBq.. | [M]Burkitt's tumours | 0 |
| BBq0. | [M]Burkitt's tumour | 0 |
| BBqz. | [M]Burkitt's tumour NOS | 0 |
| BBr27 | [M]Adult T-cell leukaemia/lymphoma | 0 |
| BBr5. | [M]Lymphosarcoma cell leukaemias | 0 |
| BBr50 | [M]Lymphosarcoma cell leukaemia | 0 |
| BBr5z | [M]Lymphosarcoma cell leukaemia NOS | 0 |
| BBv0. | [M]Monocytoid B-cell lymphoma | 0 |
| BBv2. | [M]AngiocentricT-cell lymphoma | 0 |
| ByuD. | [X]Malignant neoplasms of lymphoid, haematopoietic and related tissue | 0 |
| ByuD0 | [X]Other Hodgkin's disease | 0 |
| ByuD1 | [X]Other types of follicular non-Hodgkin's lymphoma | 0 |
| ByuD2 | [X]Other types of diffuse non-Hodgkin's lymphoma | 0 |
| ByuD3 | [X]Other specified types of non-Hodgkin's lymphoma | 0 |
| ByuD4 | [X]Other malignant immunoproliferative diseases | 0 |
| ByuDA | [X]Other specified malignant neoplasms of lymphoid, haematopoietic and related tissue | 0 |
| ByuDB | [X]Malignant neoplasm of lymphoid, haematopoietic and related tissue, unspecified | 0 |
| ByuDC | [X]Diffuse non-Hodgkin's lymphoma, unspecified | 0 |
| ByuDD | [X]Other and unspecified peripheral and cutaneous T-cell lymphomas | 0 |
| ByuDE | [X]Unspecified B-cell non-Hodgkin's lymphoma | 0 |
| ByuDF | [X]Non-Hodgkin's lymphoma, unspecified type | 0 |
| C333. | Macroglobulinaemia | 0 |
| C3330 | Waldenstrom's macroglobulinaemia | 0 |
| C333z | Macroglobulinaemia NOS | 0 |
| C37y0 | Hand - Schuller - Christian disease | 0 |
| C37y5 | Histiocytosis X , chronic | 0 |
| C37y6 | Histiocytosis X , unspecified | 0 |
| C37yB | Langerhans' cell histiocytosis | 0 |
| C37yz | Other specified disorder of metabolism NOS | 0 |
| Dyu44 | [X]Other histiocytosis syndromes | 0 |
| G753. | Lethal midline granuloma | 0 |
| M1628 | Lymphomatoid papulosis | 0 |
| ZV107 | [V]Personal history of other lymphatic or haematopoietic neoplasm | 0 |
| ZVu6G | [X]Personal history of other malignant neoplasms of lymphoid, haematopoietic and related tissues | 0 |

---

#### CONCEPT: Marfan Syndrome¶

| READ\_CODE | DESCRIPTION\_SAIL | SENSITIVE\_CODE |
| --- | --- | --- |
| PKy2. | Marfan's syndrome | 0 |

---

#### CONCEPT: Meniere's disease¶

| READ\_CODE | DESCRIPTION\_SAIL | SENSITIVE\_CODE |
| --- | --- | --- |
| 1491. | H/O: vertigo/Meniere's disease | 0 |
| F560. | Meniere's disease | 0 |
| F5600 | Unspecified Meniere's disease | 0 |
| F5601 | Active cochleovestibular Meniere's disease | 0 |
| F5602 | Active cochlear Meniere's disease | 0 |
| F5603 | Active vestibular Meniere's disease | 0 |
| F560z | Meniere's disease NOS | 0 |

---

#### CONCEPT: Metastatic cancers¶

| READ\_CODE | DESCRIPTION\_SAIL | SENSITIVE\_CODE |
| --- | --- | --- |
| 1D18. | Pain from metastases | 0 |
| 5136. | X-ray metastasis control | 0 |
| 5A12. | Thyroid tumour/metast irradiat | 0 |
| 5A15. | Bone tumour/metast.irradiat. | 0 |
| 8Hgg. | Discharge from secondary care cancer service | 0 |
| A7886 | Human immunodeficiency virus with secondary cancers | 1 |
| B153. | Secondary malignant neoplasm of liver | 0 |
| B56.. | Secondary and unspecified malignant neoplasm of lymph nodes | 0 |
| B560. | Secondary and unspecified malignant neoplasm of lymph nodes of head, face and neck | 0 |
| B5600 | Secondary and unspecified malignant neoplasm of superficial parotid lymph nodes | 0 |
| B5601 | Secondary and unspecified malignant neoplasm of mastoid lymph nodes | 0 |
| B5602 | Secondary and unspecified malignant neoplasm of superficial cervical lymph nodes | 0 |
| B5603 | Secondary and unspecified malignant neoplasm of occipital lymph nodes | 0 |
| B5604 | Secondary and unspecified malignant neoplasm of deep parotid lymph nodes | 0 |
| B5605 | Secondary and unspecified malignant neoplasm of submandibular lymph nodes | 0 |
| B5606 | Secondary and unspecified malignant neoplasm of facial lymph nodes | 0 |
| B5607 | Secondary and unspecified malignant neoplasm of submental lymph nodes | 0 |
| B5608 | Secondary and unspecified malignant neoplasm of anterior cervical lymph nodes | 0 |
| B5609 | Secondary and unspecified malignant neoplasm of deep cervical lymph nodes | 0 |
| B560z | Secondary and unspecified malignant neoplasm of lymph nodes of head, face and neck NOS | 0 |
| B561. | Secondary and unspecified malignant neoplasm of intrathoracic lymph nodes | 0 |
| B5610 | Secondary and unspecified malignant neoplasm of internal mammary lymph nodes | 0 |
| B5611 | Secondary and unspecified malignant neoplasm of intercostal lymph nodes | 0 |
| B5612 | Secondary and unspecified malignant neoplasm of diaphragmatic lymph nodes | 0 |
| B5613 | Secondary and unspecified malignant neoplasm of anterior mediastinal lymph nodes | 0 |
| B5614 | Secondary and unspecified malignant neoplasm of posterior mediastinal lymph nodes | 0 |
| B5615 | Secondary and unspecified malignant neoplasm of paratracheal lymph nodes | 0 |
| B5616 | Secondary and unspecified malignant neoplasm of superficial tracheobronchial lymph nodes | 0 |
| B5617 | Secondary and unspecified malignant neoplasm of inferior tracheobronchial lymph nodes | 0 |
| B5618 | Secondary and unspecified malignant neoplasm of bronchopulmonary lymph nodes | 0 |
| B5619 | Secondary and unspec malig neop pulmonary lymph nodes | 0 |
| B561z | Secondary and unspecified malignant neoplasm of intrathoracic lymph nodes NOS | 0 |
| B562. | Secondary and unspecified malignant neoplasm of intra-abdominal lymph nodes | 0 |
| B5620 | Secondary and unspecified malignant neoplasm of coeliac lymph nodes | 0 |
| B5621 | Secondary and unspecified malignant neoplasm of superficial mesenteric lymph nodes | 0 |
| B5622 | Secondary and unspecified malignant neoplasm of inferior mesenteric lymph nodes | 0 |
| B5623 | Secondary and unspecified malignant neoplasm of common iliac lymph nodes | 0 |
| B5624 | Secondary and unspecified malignant neoplasm of external iliac lymph nodes | 0 |
| B562z | Secondary and unspecified malignant neoplasm of intra-abdominal lymph nodes NOS | 0 |
| B563. | Secondary and unspecified malignant neoplasm of axilla and upper limb lymph nodes | 0 |
| B5630 | Secondary and unspecified malignant neoplasm of axillary lymph nodes | 0 |
| B5631 | Secondary and unspecified malignant neoplasm of supratrochlear lymph nodes | 0 |
| B5632 | Secondary and unspecified malignant neoplasm of infraclavicular lymph nodes | 0 |
| B5633 | Secondary and unspecified malignant neoplasm of pectoral lymph nodes | 0 |
| B563z | Secondary and unspecified malignant neoplasm of axilla and upper limb lymph nodes NOS | 0 |
| B564. | Secondary and unspecified malignant neoplasm of inguinal and lower limb lymph nodes | 0 |
| B5640 | Secondary and unspecified malignant neoplasm of superficial inguinal lymph nodes | 0 |
| B5641 | Secondary and unspecified malignant neoplasm of deep inguinal lymph nodes | 0 |
| B5642 | Secondary and unspecified malignant neoplasm of popliteal lymph nodes | 0 |
| B564z | Secondary and unspecified malignant neoplasm of inguinal and lower limb lymph nodes NOS | 0 |
| B565. | Secondary and unspecified malignant neoplasm of intrapelvic lymph nodes | 0 |
| B5650 | Secondary and unspecified malignant neoplasm of internal iliac lymph nodes | 0 |
| B5651 | Secondary and unspecified malignant neoplasm of inferior epigastric lymph nodes | 0 |
| B5652 | Secondary and unspecified malignant neoplasm of circumflex iliac lymph nodes | 0 |
| B5653 | Secondary and unspecified malignant neoplasm of sacral lymph nodes | 0 |
| B5654 | Secondary and unspecified malignant neoplasm of obturator lymph nodes | 0 |
| B565z | Secondary and unspecified malignant neoplasm of intrapelvic lymph nodes NOS | 0 |
| B56y. | Secondary and unspecified malignant neoplasm of lymph nodes in multiple sites | 0 |
| B56z. | Secondary and unspecified malignant neoplasm of lymph nodes NOS | 0 |
| B57.. | Secondary malignant neoplasm of respiratory and digestive systems | 0 |
| B570. | Secondary malignant neoplasm of lung | 0 |
| B571. | Secondary malignant neoplasm of mediastinum | 0 |
| B572. | Secondary malignant neoplasm of pleura | 0 |
| B573. | Secondary malignant neoplasm of other respiratory organs | 0 |
| B574. | Secondary malignant neoplasm of small intestine and duodenum | 0 |
| B5740 | Secondary malignant neoplasm of duodenum | 0 |
| B5741 | Secondary malignant neoplasm of jejunum | 0 |
| B5742 | Secondary malignant neoplasm of ileum | 0 |
| B574z | Secondary malignant neoplasm of small intestine or duodenum NOS | 0 |
| B575. | Secondary malignant neoplasm of large intestine and rectum | 0 |
| B5750 | Secondary malignant neoplasm of colon | 0 |
| B5751 | Secondary malignant neoplasm of rectum | 0 |
| B575z | Secondary malignant neoplasm of large intestine or rectum NOS | 0 |
| B576. | Secondary malignant neoplasm of retroperitoneum and peritoneum | 0 |
| B5760 | Secondary malignant neoplasm of retroperitoneum | 0 |
| B5761 | Secondary malignant neoplasm of peritoneum | 0 |
| B576z | Secondary malignant neoplasm of retroperitoneum or peritoneum NOS | 0 |
| B577. | Secondary malignant neoplasm of liver | 0 |
| B5770 | Secondary malignant neoplasm of liver and intrahepatic bile duct | 0 |
| B57y. | Secondary malignant neoplasm of other digestive organ | 0 |
| B57z. | Secondary malignant neoplasm of respiratory or digestive system NOS | 0 |
| B58.. | Secondary malignant neoplasm of other specified sites | 0 |
| B580. | Secondary malignant neoplasm of kidney | 0 |
| B581. | Secondary malignant neoplasm of other urinary organs | 0 |
| B5810 | Secondary malignant neoplasm of ureter | 0 |
| B5811 | Secondary malignant neoplasm of bladder | 0 |
| B5812 | Secondary malignant neoplasm of urethra | 0 |
| B581z | Secondary malignant neoplasm of other urinary organ NOS | 0 |
| B582. | Secondary malignant neoplasm of skin | 0 |
| B5820 | Secondary malignant neoplasm of skin of head | 0 |
| B5821 | Secondary malignant neoplasm of skin of face | 0 |
| B5822 | Secondary malignant neoplasm of skin of neck | 0 |
| B5823 | Secondary malignant neoplasm of skin of trunk | 0 |
| B5824 | Secondary malignant neoplasm of skin of shoulder and arm | 0 |
| B5825 | Secondary malignant neoplasm of skin of hip and leg | 0 |
| B5826 | Secondary malignant neoplasm of skin of breast | 0 |
| B582z | Secondary malignant neoplasm of skin NOS | 0 |
| B583. | Secondary malignant neoplasm of brain and spinal cord | 0 |
| B5830 | Secondary malignant neoplasm of brain | 0 |
| B5831 | Secondary malignant neoplasm of spinal cord | 0 |
| B5832 | Cerebral metastasis | 0 |
| B583z | Secondary malignant neoplasm of brain or spinal cord NOS | 0 |
| B584. | Secondary malignant neoplasm of other part of nervous system | 0 |
| B585. | Secondary malignant neoplasm of bone and bone marrow | 0 |
| B5850 | Pathological fracture due to metastatic bone disease | 0 |
| B586. | Secondary malignant neoplasm of ovary | 0 |
| B587. | Secondary malignant neoplasm of adrenal gland | 0 |
| B58y. | Secondary malignant neoplasm of other specified sites | 0 |
| B58y0 | Secondary malignant neoplasm of breast | 0 |
| B58y1 | Secondary malignant neoplasm of uterus | 0 |
| B58y2 | Secondary malignant neoplasm of cervix uteri | 0 |
| B58y3 | Secondary malignant neoplasm of vagina | 0 |
| B58y4 | Secondary malignant neoplasm of vulva | 0 |
| B58y5 | Secondary malignant neoplasm of prostate | 0 |
| B58y6 | Secondary malignant neoplasm of testis | 0 |
| B58y7 | Secondary malignant neoplasm of penis | 0 |
| B58y8 | Secondary malignant neoplasm of epididymis and vas deferens | 0 |
| B58y9 | Secondary malignant neoplasm of tongue | 0 |
| B58yz | Secondary malignant neoplasm of other specified site NOS | 0 |
| B58z. | Secondary malignant neoplasm of other specified site NOS | 0 |
| B590. | Disseminated malignancy NOS | 0 |
| B594. | Secondary malignant neoplasm of unknown site | 0 |
| BB03. | [M]Neoplasm, metastatic | 0 |
| BB04. | [M]Neoplasm, malignant, uncertain whether primary or metastatic | 0 |
| BB13. | [M]Carcinoma, metastatic, NOS | 0 |
| BB14. | [M]Carcinomatosis | 0 |
| BB2B. | [M]Squamous cell carcinoma, metastatic NOS | 0 |
| BB53. | [M]Adenocarcinoma, metastatic, NOS | 0 |
| BB851 | [M]Metastatic signet ring cell carcinoma | 0 |
| BBy2. | [M]No microscopic confirmation of tumour, clinically metastatic | 0 |
| ByuC. | [X]Malignant neoplasm of ill-defined, secondary and unspecified sites | 0 |
| ByuC2 | [X]Secondary and unspecified malignant neoplasm lymph nodes of multiple regions | 0 |
| ByuC3 | [X]Secondary malignant neoplasm of other and unspecified respiratory organs | 0 |
| ByuC4 | [X]Secondary malignant neoplasm of other and unspecified digestive organs | 0 |
| ByuC5 | [X]Secondary malignant neoplasm of bladder and other and unspecified urinary organs | 0 |
| ByuC6 | [X]Secondary malignant neoplasm of other and unspecified parts of nervous system | 0 |
| ByuC7 | [X]Secondary malignant neoplasm of other specified sites | 0 |
| C354A | Metastatic calcification | 0 |
| F4314 | Metastatic disseminated retinitis | 0 |

---

#### CONCEPT: Multiple sclerosis¶

| READ\_CODE | DESCRIPTION\_SAIL | SENSITIVE\_CODE |
| --- | --- | --- |
| 666A. | Multiple sclerosis review | 0 |
| 666B. | Multiple sclerosis multidisciplinary review | 0 |
| 8CS1. | Multiple sclerosis care plan agreed | 0 |
| 8Cc0. | Management of multiple sclerosis in onset phase | 0 |
| 8Cc1. | Management of multiple sclerosis in early disease phase | 0 |
| 8Cc2. | Management of multiple sclerosis in stable disability phase | 0 |
| 8Cc4. | Management of multiple sclerosis in palliative phase | 0 |
| 8IAb. | Multiple sclerosis review declined | 0 |
| 9kG.. | Specialised services for patients with multiple sclerosis - enhanced services administration | 0 |
| 9mD.. | Multiple sclerosis monitoring administration | 0 |
| 9mD0. | Multiple sclerosis monitoring first letter | 0 |
| 9mD1. | Multiple sclerosis monitoring second letter | 0 |
| 9mD2. | Multiple sclerosis monitoring telephone invitation | 0 |
| 9mD3. | Multiple sclerosis monitoring third letter | 0 |
| F20.. | Multiple sclerosis | 0 |
| F200. | Multiple sclerosis of the brain stem | 0 |
| F201. | Multiple sclerosis of the spinal cord | 0 |
| F202. | Generalised multiple sclerosis | 0 |
| F203. | Exacerbation of multiple sclerosis | 0 |
| F204. | Benign multiple sclerosis | 0 |
| F205. | Malignant multiple sclerosis | 0 |
| F206. | Primary progressive multiple sclerosis | 0 |
| F207. | Relapsing and remitting multiple sclerosis | 0 |
| F208. | Secondary progressive multiple sclerosis | 0 |
| F20z. | Multiple sclerosis NOS | 0 |

---

#### CONCEPT: Osteoarthritis¶

| READ\_CODE | DESCRIPTION\_SAIL | SENSITIVE\_CODE |
| --- | --- | --- |
| 14G2. | H/O: osteoarthritis | 0 |
| 7P204 | Delivery of rehabilitation for osteoarthritis | 0 |
| 9hQ.. | Exception reporting: osteoarthritis quality indicators | 0 |
| 9hQ0. | Excepted from osteoarthritis quality indicators: patient unsuitable | 0 |
| 9hQ1. | Excepted from osteoarthritis quality indicators: informed dissent | 0 |
| F1632 | Myelopathy due to spondylosis | 0 |
| F3372 | Nerve root and plexus compressions in spondylosis | 0 |
| N0... | Arthropathies and related disorders | 0 |
| N05.. | Osteoarthritis and allied disorders | 0 |
| N050. | Generalised osteoarthritis - OA | 0 |
| N0500 | Generalised osteoarthritis of unspecified site | 0 |
| N0501 | Generalised osteoarthritis of the hand | 0 |
| N0502 | Generalised osteoarthritis of multiple sites | 0 |
| N0504 | Primary generalized osteoarthrosis | 0 |
| N0505 | Secondary multiple arthrosis | 0 |
| N0506 | Erosive osteoarthrosis | 0 |
| N0507 | Heberden's nodes with arthropathy | 0 |
| N050z | Generalised osteoarthritis NOS | 0 |
| N051. | Localised, primary osteoarthritis | 0 |
| N0510 | Localised, primary osteoarthritis of unspecified site | 0 |
| N0511 | Localised, primary osteoarthritis of the shoulder region | 0 |
| N0512 | Localised, primary osteoarthritis of the upper arm | 0 |
| N0513 | Localised, primary osteoarthritis of the forearm | 0 |
| N0514 | Localised, primary osteoarthritis of the hand | 0 |
| N0515 | Localised, primary osteoarthritis of the pelvic region and thigh | 0 |
| N0516 | Localised, primary osteoarthritis of the lower leg | 0 |
| N0517 | Localised, primary osteoarthritis of the ankle and foot | 0 |
| N0518 | Localised, primary osteoarthritis of other specified site | 0 |
| N0519 | Primary coxarthrosis, bilateral | 0 |
| N051B | Primary gonarthrosis, bilateral | 0 |
| N051C | Primary arthrosis of first carpometacarpal joints, bilateral | 0 |
| N051D | Localised, primary osteoarthritis of the wrist | 0 |
| N051E | Localised, primary osteoarthritis of toe | 0 |
| N051F | Localised, primary osteoarthritis of elbow | 0 |
| N051G | Osteoarthritis of spinal facet joint | 0 |
| N051z | Localised, primary osteoarthritis NOS | 0 |
| N052. | Localised, secondary osteoarthritis | 0 |
| N0520 | Localised, secondary osteoarthritis of unspecified site | 0 |
| N0521 | Localised, secondary osteoarthritis of the shoulder region | 0 |
| N0522 | Localised, secondary osteoarthritis of the upper arm | 0 |
| N0523 | Localised, secondary osteoarthritis of the forearm | 0 |
| N0524 | Localised, secondary osteoarthritis of the hand | 0 |
| N0525 | Localised, secondary osteoarthritis of the pelvic region and thigh | 0 |
| N0526 | Localised, secondary osteoarthritis of the lower leg | 0 |
| N0527 | Localised, secondary osteoarthritis of the ankle and foot | 0 |
| N0528 | Localised, secondary osteoarthritis of other specified site | 0 |
| N0529 | Post-traumatic coxarthrosis, bilateral | 0 |
| N052A | Post-traumatic gonarthrosis, bilateral | 0 |
| N052B | Post-traumatic arthrosis of first carpometacarpal joints, bilateral | 0 |
| N052C | Post-traumatic gonarthrosis, unilateral | 0 |
| N052z | Localised, secondary osteoarthritis NOS | 0 |
| N053. | Localised osteoarthritis, unspecified | 0 |
| N0530 | Localised osteoarthritis, unspecified, of unspecified site | 0 |
| N0531 | Localised osteoarthritis, unspecified, of the shoulder region | 0 |
| N0532 | Localised osteoarthritis, unspecified, of the upper arm | 0 |
| N0533 | Localised osteoarthritis, unspecified, of the forearm | 0 |
| N0534 | Localised osteoarthritis, unspecified, of the hand | 0 |
| N0535 | Localised osteoarthritis, unspecified, of the pelvic region and thigh | 0 |
| N0536 | Localised osteoarthritis, unspecified, of the lower leg | 0 |
| N0537 | Localised osteoarthritis, unspecified, of the ankle and foot | 0 |
| N0538 | Localised osteoarthritis, unspecified, of other specified site | 0 |
| N0539 | Arthrosis of first carpometacarpal joint, unspecified | 0 |
| N053z | Localised osteoarthritis, unspecified, NOS | 0 |
| N054. | Oligoarticular osteoarthritis, unspecified | 0 |
| N0540 | Oligoarticular osteoarthritis, unspecified, of unspecified sites | 0 |
| N0541 | Oligoarticular osteoarthritis, unspecified, of the shoulder region | 0 |
| N0542 | Oligoarticular osteoarthritis, unspecified, of upper arm | 0 |
| N0543 | Oligoarticular osteoarthritis, unspecified, of forearm | 0 |
| N0544 | Oligoarticular osteoarthritis, unspecified, of hand | 0 |
| N0545 | Oligoarticular osteoarthritis, unspecified, of the pelvic region and thigh | 0 |
| N0546 | Oligoarticular osteoarthritis, unspecified, of lower leg | 0 |
| N0547 | Oligoarticular osteoarthritis, unspecified, of ankle and foot | 0 |
| N0548 | Oligoarticular osteoarthritis, unspecified, of other specified sites | 0 |
| N0549 | Oligoarticular osteoarthritis, unspecified, of multiple sites | 0 |
| N054z | Osteoarthritis of more than one site, unspecified, NOS | 0 |
| N05z. | Osteoarthritis NOS | 0 |
| N05z0 | Osteoarthritis NOS, of unspecified site | 0 |
| N05z1 | Osteoarthritis NOS, of shoulder region | 0 |
| N05z2 | Osteoarthritis NOS, of the upper arm | 0 |
| N05z3 | Osteoarthritis NOS, of the forearm | 0 |
| N05z4 | Osteoarthritis NOS, of the hand | 0 |
| N05z5 | Osteoarthritis NOS, pelvic region/thigh | 0 |
| N05z6 | Osteoarthritis NOS, of the lower leg | 0 |
| N05z7 | Osteoarthritis NOS, of ankle and foot | 0 |
| N05z8 | Osteoarthritis NOS, other specified site | 0 |
| N05z9 | Osteoarthritis NOS, of shoulder | 0 |
| N05zA | Osteoarthritis NOS, of sternoclavicular joint | 0 |
| N05zB | Osteoarthritis NOS, of acromioclavicular joint | 0 |
| N05zC | Osteoarthritis NOS, of elbow | 0 |
| N05zD | Osteoarthritis NOS, of distal radio-ulnar joint | 0 |
| N05zE | Osteoarthritis NOS, of wrist | 0 |
| N05zF | Osteoarthritis NOS, of metacarpophalangeal joint | 0 |
| N05zG | Osteoarthritis NOS, of proximal interphalangeal joint of finger | 0 |
| N05zH | Osteoarthritis NOS, of distal interphalangeal joint of finger | 0 |
| N05zJ | Osteoarthritis NOS, of hip | 0 |
| N05zK | Osteoarthritis NOS, of sacro-iliac joint | 0 |
| N05zL | Osteoarthritis NOS, of knee | 0 |
| N05zM | Osteoarthritis NOS, of tibio-fibular joint | 0 |
| N05zN | Osteoarthritis NOS, of ankle | 0 |
| N05zP | Osteoarthritis NOS, of subtalar joint | 0 |
| N05zQ | Osteoarthritis NOS, of talonavicular joint | 0 |
| N05zR | Osteoarthritis NOS, of other tarsal joint | 0 |
| N05zS | Osteoarthritis NOS, of 1st metatarsophalangeal joint | 0 |
| N05zT | Osteoarthritis NOS, of lesser metatarsophalangeal joint | 0 |
| N05zU | Osteoarthritis NOS, of interphalangeal joint of toe | 0 |
| N05zz | Osteoarthritis NOS | 0 |
| N11.. | Spondylosis and allied disorders | 0 |
| N110. | Cervical spondylosis without myelopathy | 0 |
| N1100 | Single-level cervical spondylosis without myelopathy | 0 |
| N1101 | Two-level cervical spondylosis without myelopathy | 0 |
| N1102 | Multiple-level cervical spondylosis without myelopathy | 0 |
| N111. | Cervical spondylosis with myelopathy | 0 |
| N1110 | Single-level cervical spondylosis with myelopathy | 0 |
| N1111 | Two-level cervical spondylosis with myelopathy | 0 |
| N1112 | Multiple-level cervical spondylosis with myelopathy | 0 |
| N112. | Thoracic spondylosis without myelopathy | 0 |
| N1120 | Single-level thoracic spondylosis without myelopathy | 0 |
| N1121 | Two-level thoracic spondylosis without myelopathy | 0 |
| N1122 | Multiple-level thoracic spondylosis without myelopathy | 0 |
| N1123 | Dorsal spondylosis without myelopathy | 0 |
| N113. | Thoracic spondylosis with myelopathy | 0 |
| N1130 | Single-level thoracic spondylosis with myelopathy | 0 |
| N1131 | Two-level thoracic spondylosis with myelopathy | 0 |
| N1132 | Multiple-level thoracic spondylosis with myelopathy | 0 |
| N114. | Lumbosacral spondylosis without myelopathy | 0 |
| N1140 | Single-level lumbosacral spondylosis without myelopathy | 0 |
| N1141 | Two-level lumbosacral spondylosis without myelopathy | 0 |
| N1142 | Multiple-level lumbosacral spondylosis without myelopathy | 0 |
| N115. | Lumbosacral spondylosis with myelopathy | 0 |
| N1150 | Single-level lumbosacral spondylosis with myelopathy | 0 |
| N1151 | Two-level lumbosacral spondylosis with myelopathy | 0 |
| N1152 | Multiple-level lumbosacral spondylosis with myelopathy | 0 |
| N119. | Cervical spondylosis with radiculopathy | 0 |
| N1190 | Single-level cervical spondylosis with radiculopathy | 0 |
| N1191 | Two-level cervical spondylosis with radiculopathy | 0 |
| N1192 | Multiple-level cervical spondylosis with radiculopathy | 0 |
| N11A. | Cervical spondylosis with vascular compression | 0 |
| N11B. | Thoracic spondylosis with radiculopathy | 0 |
| N11B0 | Single-level thoracic spondylosis with radiculopathy | 0 |
| N11B1 | Two-level thoracic spondylosis with radiculopathy | 0 |
| N11B2 | Multiple-level thoracic spondylosis with radiculopathy | 0 |
| N11C. | Lumbosacral spondylosis with radiculopathy | 0 |
| N11C0 | Single-level lumbosacral spondylosis with radiculopathy | 0 |
| N11C1 | Two-level lumbosacral spondylosis with radiculopathy | 0 |
| N11C2 | Multiple-level lumbosacral spondylosis with radiculopathy | 0 |
| N11D. | Osteoarthritis of spine | 0 |
| N11D0 | Osteoarthritis of cervical spine | 0 |
| N11D1 | Osteoarthritis of thoracic spine | 0 |
| N11D2 | Osteoarthritis of lumbar spine | 0 |
| N11D3 | Osteoarthritis of spine NOS | 0 |
| N11E. | Cervical spondylosis | 0 |
| N11y. | Other spondyloses and allied disorders | 0 |
| N11z. | Spondylosis NOS | 0 |
| N11z0 | Spondylosis without myelopathy, NOS | 0 |
| N11z1 | Spondylosis with myelopathy, NOS | 0 |
| N11zz | Spondylosis NOS | 0 |
| Nyu2. | [X]Arthrosis | 0 |
| Nyu20 | [X]Other polyarthrosis | 0 |
| Nyu21 | [X]Other primary coxarthrosis | 0 |
| Nyu23 | [X]Other post-traumatic coxarthrosis | 0 |
| Nyu24 | [X]Other secondary coxarthrosis, bilateral | 0 |
| Nyu25 | [X]Other primary gonarthrosis | 0 |
| Nyu26 | [X]Other post-traumatic gonarthrosis | 0 |
| Nyu27 | [X]Other secondary gonarthrosis, bilateral | 0 |
| Nyu28 | [X]Other secondary gonarthrosis | 0 |
| Nyu29 | [X]Other primary arthrosis of first carpometacarpal joint | 0 |
| Nyu2A | [X]Other post-traumatic arthrosis of first carpometacarpal joint | 0 |
| Nyu2B | [X]Other secondary arthrosis of first carpometacarpal joints, bilateral | 0 |
| Nyu2C | [X]Other secondary arthrosis of first carpometacarpal joint | 0 |
| Nyu2D | [X]Other specified arthrosis | 0 |
| Nyu2E | [X]Other secondary coxarthrosis | 0 |
| Nyu2F | [X]Post-traumatic arthrosis of other joints | 0 |
| Nyu62 | [X]Other spondylosis with myelopathy | 0 |
| Nyu63 | [X]Other spondylosis with radiculopathy | 0 |
| Nyu64 | [X]Other spondylosis | 0 |

---

#### CONCEPT: Osteoporosis¶

| READ\_CODE | DESCRIPTION\_SAIL | SENSITIVE\_CODE |
| --- | --- | --- |
| 14G6. | H/O: fragility fracture | 0 |
| 14GB. | History of osteoporosis | 0 |
| 585O. | Quantitative ultrasound scan of heel - result osteoporotic | 0 |
| 58E4. | Forearm DXA scan result osteoporotic | 0 |
| 58EA. | Heel DXA scan result osteoporotic | 0 |
| 58EG. | Hip DXA scan result osteoporotic | 0 |
| 58EM. | Lumbar DXA scan result osteoporotic | 0 |
| 58EV. | Femoral neck DEXA scan result osteoporotic | 0 |
| 66a2. | Osteoporosis treatment started | 0 |
| 66a3. | Osteoporosis treatment stopped | 0 |
| 66a4. | Osteoporosis treatment changed | 0 |
| 66a5. | Osteoporosis - no treatment | 0 |
| 66aA. | Osteoporosis - treatment response | 0 |
| 66aB. | Osteoporosis - no treatment response | 0 |
| 8B31E | Osteoporosis medication compliance review | 0 |
| 9kj.. | Osteoporosis - enhanced services administration | 0 |
| 9kj0. | Bone sparing drug treatment offered for osteoporosis - enhanced services administration | 0 |
| N330. | Osteoporosis | 0 |
| N3300 | Osteoporosis, unspecified | 0 |
| N3301 | Senile osteoporosis | 0 |
| N3302 | Postmenopausal osteoporosis | 0 |
| N3303 | Idiopathic osteoporosis | 0 |
| N3304 | Dissuse osteoporosis | 0 |
| N3305 | Drug-induced osteoporosis | 0 |
| N3306 | Postoophorectomy osteoporosis | 0 |
| N3307 | Postsurgical malabsorption osteoporosis | 0 |
| N3308 | Localized osteoporosis - Lequesne | 0 |
| N3309 | Osteoporosis in multiple myelomatosis | 0 |
| N330A | Osteoporosis in endocrine disorders | 0 |
| N330B | Vertebral osteoporosis | 0 |
| N330C | Osteoporosis localized to spine | 0 |
| N330D | Osteoporosis due to corticosteroids | 0 |
| N330z | Osteoporosis NOS | 0 |
| N331. | Pathological fracture | 0 |
| N3312 | Postoophorectomy osteoporosis with pathological fracture | 0 |
| N3313 | Osteoporosis of disuse with pathological fracture | 0 |
| N3314 | Postsurgical malabsorption osteoporosis with pathological fracture | 0 |
| N3315 | Drug-induced osteoporosis with pathological fracture | 0 |
| N3316 | Idiopathic osteoporosis with pathological fracture | 0 |
| N3318 | Osteoporosis + pathological fracture lumbar vertebrae | 0 |
| N3319 | Osteoporosis + pathological fracture thoracic vertebrae | 0 |
| N331A | Osteoporosis + pathological fracture cervical vertebrae | 0 |
| N331B | Postmenopausal osteoporosis with pathological fracture | 0 |
| N331H | Collapse of cervical vertebra due to osteoporosis | 0 |
| N331J | Collapse of lumbar vertebra due to osteoporosis | 0 |
| N331K | Collapse of thoracic vertebra due to osteoporosis | 0 |
| N331L | Collapse of vertebra due to osteoporosis NOS | 0 |
| N331M | Fragility fracture due to unspecified osteoporosis | 0 |
| N331N | Fragility fracture | 0 |
| N3370 | Disuse atrophy of bone | 0 |
| N3746 | Osteoporotic kyphosis | 0 |
| NyuB0 | [X]Other osteoporosis with pathological fracture | 0 |
| NyuB1 | [X]Other osteoporosis | 0 |
| NyuB2 | [X]Osteoporosis in other disorders classified elsewhere | 0 |
| NyuB8 | [X]Unspecified osteoporosis with pathological fracture | 0 |

---

#### CONCEPT: Pancreatic disease¶

| READ\_CODE | DESCRIPTION\_SAIL | SENSITIVE\_CODE |
| --- | --- | --- |
| 14CH. | History of chronic pancreatitis | 0 |
| J671. | Chronic pancreatitis | 0 |
| J6710 | Alcohol-induced chronic pancreatitis | 0 |
| J6711 | Gallstone chronic pancreatitis | 0 |
| J67y0 | Atrophy of pancreas | 0 |
| J67y2 | Fibrosis of pancreas | 0 |
| Jyu84 | [X]Other chronic pancreatitis | 0 |

---

#### CONCEPT: Paralysis¶

| READ\_CODE | DESCRIPTION\_SAIL | SENSITIVE\_CODE |
| --- | --- | --- |
| 2833. | O/E - hemiplegia | 0 |
| 2835. | O/E - paraplegia | 0 |
| 2836. | O/E - quadriplegia | 0 |
| F038. | Tropical spastic paraplegia | 0 |
| F141. | Hereditary spastic paraplegia | 0 |
| F22.. | Hemiplegia | 0 |
| F220. | Flaccid hemiplegia | 0 |
| F221. | Spastic hemiplegia | 0 |
| F222. | Left hemiplegia | 0 |
| F223. | Right hemiplegia | 0 |
| F22z. | Hemiplegia NOS | 0 |
| F230. | Congenital diplegia | 0 |
| F2300 | Congenital paraplegia | 0 |
| F231. | Congenital hemiplegia | 0 |
| F232. | Congenital quadriplegia | 0 |
| F234. | Infantile hemiplegia NOS | 0 |
| F240. | Quadriplegia | 0 |
| F2400 | Flaccid tetraplegia | 0 |
| F2401 | Spastic tetraplegia | 0 |
| F241. | Paraplegia | 0 |
| F2410 | Flaccid paraplegia | 0 |
| F2411 | Spastic paraplegia | 0 |
| F2B0. | Spastic quadriplegic cerebral palsy | 0 |
| F2B1. | Spastic hemiplegic cerebral palsy | 0 |
| N2310 | Muscular calcification and ossification, unspecified | 0 |

---

#### CONCEPT: Parkinson's disease¶

| READ\_CODE | DESCRIPTION\_SAIL | SENSITIVE\_CODE |
| --- | --- | --- |
| 147F. | History of Parkinson's disease | 0 |
| 297A. | O/E - Parkinsonian tremor | 0 |
| 2987. | O/E -Parkinson flexion posture | 0 |
| 2994. | O/E-festination-Parkinson gait | 0 |
| A94y1 | Syphilitic parkinsonism | 1 |
| Eu023 | [X]Dementia in Parkinson's disease | 0 |
| F11x9 | Cerebral degeneration in Parkinson's disease | 0 |
| F12.. | Parkinson's disease | 0 |
| F120. | Paralysis agitans | 0 |
| F121. | Parkinsonism secondary to drugs | 0 |
| F123. | Postencephalitic parkinsonism | 0 |
| F124. | Vascular parkinsonism | 0 |
| F12W. | Secondary parkinsonism due to other external agents | 0 |
| F12X. | Secondary parkinsonism, unspecified | 0 |
| F12z. | Parkinson's disease NOS | 0 |
| F1303 | Parkinsonism with orthostatic hypotension | 0 |
| Fyu20 | [X]Other drug-induced secondary parkinsonism | 0 |
| Fyu21 | [X]Other secondary parkinsonism | 0 |
| Fyu22 | [X]Parkinsonism in diseases classified elsewhere | 0 |
| Fyu29 | [X]Secondary parkinsonism, unspecified | 0 |
| Fyu2B | [X]Secondary parkinsonism due to other external agents | 0 |

---

#### CONCEPT: PCOS (Polycystic ovary syndrome)¶

| READ\_CODE | DESCRIPTION\_SAIL | SENSITIVE\_CODE |
| --- | --- | --- |
| 7E253 | Endoscopic drilling of ovary | 0 |
| C164. | Polycystic ovaries | 0 |
| C165. | Polycystic ovarian syndrome | 0 |

---

#### CONCEPT: Plasmacell¶

| READ\_CODE | DESCRIPTION\_SAIL | SENSITIVE\_CODE |
| --- | --- | --- |
| 4C53. | Bone marrow: myeloma cells | 0 |
| B63.. | Multiple myeloma and immunoproliferative neoplasms | 0 |
| B630. | Multiple myeloma | 0 |
| B6300 | Malignant plasma cell neoplasm, extramedullary plasmacytoma | 0 |
| B6301 | Solitary myeloma | 0 |
| B6302 | Plasmacytoma NOS | 0 |
| B6303 | Lambda light chain myeloma | 0 |
| B6304 | Solitary plasmacytoma | 0 |
| B631. | Plasma cell leukaemia | 0 |
| B63y. | Other immunoproliferative neoplasms | 0 |
| B63z. | Immunoproliferative neoplasm or myeloma NOS | 0 |
| B936. | Neoplasm of uncertain behaviour of plasma cells | 0 |
| BBm6. | [M] Alpha heavy chain disease | 0 |
| BBmE. | [M] Gamma heavy chain disease | 0 |
| BBn.. | [M]Plasma cell tumours | 0 |
| BBn0. | [M]Plasma cell myeloma | 0 |
| BBn2. | [M]Plasmacytoma NOS | 0 |
| BBn3. | [M]Plasma cell tumour, malignant | 0 |
| BBnz. | [M]Plasma cell tumour NOS | 0 |
| BBr3. | [M]Plasma cell leukaemias | 0 |
| BBr30 | [M]Plasma cell leukaemia | 0 |
| BBr3z | [M]Plasma cell leukaemia NOS | 0 |
| C3331 | Alpha heavy chain disease | 0 |
| C3332 | Gamma heavy chain disease | 0 |
| C3333 | Heavy chain disease | 0 |
| N3309 | Osteoporosis in multiple myelomatosis | 0 |

---

#### CONCEPT: PMR (Polymyalgia reheumatica)¶

| READ\_CODE | DESCRIPTION\_SAIL | SENSITIVE\_CODE |
| --- | --- | --- |
| N20.. | Polymyalgia rheumatica | 0 |
| N200. | Giant cell arteritis with polymyalgia rheumatica | 0 |

---

#### CONCEPT: Primary Lung Cancer¶

| READ\_CODE | DESCRIPTION\_SAIL | SENSITIVE\_CODE |
| --- | --- | --- |
| 1422. | H/O: \* bronchus | 0 |
| 1423. | H/O: \* trachea/lung | 0 |
| B22.. | Malignant neoplasm of trachea, bronchus and lung | 0 |
| B220. | Malignant neoplasm of trachea | 0 |
| B2200 | Malignant neoplasm of cartilage of trachea | 0 |
| B2201 | Malignant neoplasm of mucosa of trachea | 0 |
| B220z | Malignant neoplasm of trachea NOS | 0 |
| B221. | Malignant neoplasm of main bronchus | 0 |
| B2210 | Malignant neoplasm of carina of bronchus | 0 |
| B2211 | Malignant neoplasm of hilus of lung | 0 |
| B221z | Malignant neoplasm of main bronchus NOS | 0 |
| B222. | Malignant neoplasm of upper lobe, bronchus or lung | 0 |
| B2220 | Malignant neoplasm of upper lobe bronchus | 0 |
| B2221 | Malignant neoplasm of upper lobe of lung | 0 |
| B222z | Malignant neoplasm of upper lobe, bronchus or lung NOS | 0 |
| B223. | Malignant neoplasm of middle lobe, bronchus or lung | 0 |
| B2230 | Malignant neoplasm of middle lobe bronchus | 0 |
| B2231 | Malignant neoplasm of middle lobe of lung | 0 |
| B223z | Malignant neoplasm of middle lobe, bronchus or lung NOS | 0 |
| B224. | Malignant neoplasm of lower lobe, bronchus or lung | 0 |
| B2240 | Malignant neoplasm of lower lobe bronchus | 0 |
| B2241 | Malignant neoplasm of lower lobe of lung | 0 |
| B224z | Malignant neoplasm of lower lobe, bronchus or lung NOS | 0 |
| B225. | Malignant neoplasm of overlapping lesion of bronchus and lung | 0 |
| B22y. | Malignant neoplasm of other sites of bronchus or lung | 0 |
| B22z. | Malignant neoplasm of bronchus or lung NOS | 0 |
| BB5S2 | [M]Bronchiolo-alveolar adenocarcinoma | 0 |
| BB5S4 | [M]Alveolar adenocarcinoma | 0 |
| Byu2. | [X]Malignant neoplasm of respiratory and intrathoracic organs | 0 |
| Byu20 | [X]Malignant neoplasm of bronchus or lung, unspecified | 0 |
| ZV101 | [V]Personal history of malignant neoplasm of trachea, bronchus or lung | 0 |

---

#### CONCEPT: Prostate Cancer¶

| READ\_CODE | DESCRIPTION\_SAIL | SENSITIVE\_CODE |
| --- | --- | --- |
| 4M01. | Gleason prostate grade 5-7 (medium) | 0 |
| 4M02. | Gleason prostate grade 8-10 (high) | 0 |
| 5B51. | Microwave therapy to prostate | 0 |
| 7B3B0 | Endoscopic cryotherapy of prostate | 0 |
| 7B3B1 | Other endoscopic destruction of lesion of prostate | 0 |
| 7B3B6 | Ultrasound ablation of prostate | 0 |
| 7B3BC | Endoscopic radiofrequency ablation of lesion of prostate | 0 |
| 7B3C6 | Cryotherapy to prostate via perineal probe | 0 |
| 7B3E0 | High intensity focused ultrasound of prostate | 0 |
| 8AD0. | Active surveillance of prostate cancer | 0 |
| 8BAV0 | Prostate cancer care review | 0 |
| B46.. | Malignant neoplasm of prostate | 0 |
| B58y5 | Secondary malignant neoplasm of prostate | 0 |

---

#### CONCEPT: Prostate disorders¶

| READ\_CODE | DESCRIPTION\_SAIL | SENSITIVE\_CODE |
| --- | --- | --- |
| 14E1. | H/O: prostatism | 0 |
| 1AA.. | Prostatism | 0 |
| 7B395 | Endoscopic resection of prostate using an electrotome | 0 |
| 8L51. | Prostatectomy planned | 0 |
| A1650 | Tuberculosis of prostate | 0 |
| A9812 | Acute gonococcal prostatitis | 1 |
| A9832 | Chronic gonococcal prostatitis | 1 |
| AD103 | Trichomonal prostatitis | 1 |
| B46.. | Malignant neoplasm of prostate | 0 |
| B58y5 | Secondary malignant neoplasm of prostate | 0 |
| B7C2. | Benign neoplasm of prostate | 0 |
| B834. | Carcinoma in situ of prostate | 0 |
| B8340 | High grade prostatic intraepithelial neoplasia | 0 |
| B915. | Neoplasm of uncertain behaviour of prostate | 0 |
| K20.. | Benign prostatic hypertrophy | 0 |
| K200. | Prostatic hyperplasia unspecified | 0 |
| K201. | Prostatic hyperplasia of the lateral lobe | 0 |
| K202. | Prostatic hyperplasia of the medial lobe | 0 |
| K20z. | Prostatic hyperplasia NOS | 0 |
| K21.. | Prostate inflammatory diseases | 0 |
| K210. | Acute prostatitis | 0 |
| K211. | Chronic prostatitis | 0 |
| K212. | Abscess of prostate | 0 |
| K213. | Prostatocystitis | 0 |
| K214. | Prostatitis in diseases EC | 0 |
| K2140 | Prostatitis in actinomycosis | 0 |
| K2141 | Prostatitis in blastomycosis | 0 |
| K2142 | Prostatitis in syphilis | 1 |
| K2143 | Prostatitis in tuberculosis | 0 |
| K2144 | Prostatitis in gonorrhoea | 1 |
| K2145 | Prostatitis in moniliasis | 0 |
| K2146 | Prostatitis in trichomoniasis | 1 |
| K214z | Prostatitis in diseases EC NOS | 0 |
| K21y. | Other prostatic inflammatory diseases | 0 |
| K21z. | Prostatitis NOS | 0 |
| K22.. | Other disorders of prostate | 0 |
| K220. | Calculus of prostate | 0 |
| K221. | Prostatic congestion or haemorrhage | 0 |
| K2210 | Prostatic congestion | 0 |
| K2211 | Prostatic haemorrhage | 0 |
| K221z | Prostatic congestion or haemorrhage NOS | 0 |
| K222. | Atrophy of prostate | 0 |
| K22y. | Other disorders of prostate OS | 0 |
| K22y0 | Prostatic fistula | 0 |
| K22y1 | Infarction of prostate | 0 |
| K22y2 | Stricture of prostate | 0 |
| K22y3 | Periprostatic adhesions | 0 |
| K22yz | Other prostate disorders NOS | 0 |
| K22z. | Prostatic disorders NOS | 0 |
| Kyu60 | [X]Other inflammatory diseases of prostate | 0 |
| Kyu61 | [X]Other specified disorders of prostate | 0 |
| Kyu68 | [X]Disorders of prostate in diseases classified elsewhere | 0 |
| PCy01 | Congenital absence of prostate | 0 |
| PCyx. | Other congenital anomaly of vas deferens or prostate | 0 |
| Pyu69 | [X]Other congenital malformations of vas deferens, epididymis, seminal vesicles and prostate | 0 |

---

#### CONCEPT: Psoriasis¶

| READ\_CODE | DESCRIPTION\_SAIL | SENSITIVE\_CODE |
| --- | --- | --- |
| 14F2. | H/O: psoriasis | 0 |
| 38Gg. | Psoriasis area and severity index | 0 |
| M16.. | Psoriasis and similar disorders | 0 |
| M161. | Other psoriasis | 0 |
| M1610 | Psoriasis unspecified | 0 |
| M1611 | Psoriasis annularis | 0 |
| M1612 | Psoriasis circinata | 0 |
| M1613 | Psoriasis diffusa | 0 |
| M1614 | Psoriasis discoidea | 0 |
| M1615 | Psoriasis geographica | 0 |
| M1616 | Guttate psoriasis | 0 |
| M1617 | Psoriasis gyrata | 0 |
| M1618 | Psoriasis inveterata | 0 |
| M1619 | Psoriasis ostracea | 0 |
| M161A | Psoriasis palmaris | 0 |
| M161B | Psoriasis plantaris | 0 |
| M161C | Psoriasis punctata | 0 |
| M161D | Pustular psoriasis | 0 |
| M161E | Psoriasis universalis | 0 |
| M161F | Psoriasis vulgaris | 0 |
| M161H | Erythrodermic psoriasis | 0 |
| M161J | Flexural psoriasis | 0 |
| M161z | Psoriasis NOS | 0 |
| M166. | Palmoplantar pustular psoriasis | 0 |
| M16y. | Other psoriasis and similar disorders | 0 |
| M16y0 | Scalp psoriasis | 0 |
| M16z. | Psoriasis and similar disorders NOS | 0 |
| Myu30 | [X]Other psoriasis | 0 |

---

#### CONCEPT: Psoriasis Arthritis¶

| READ\_CODE | DESCRIPTION\_SAIL | SENSITIVE\_CODE |
| --- | --- | --- |
| M160. | Psoriatic arthropathy | 0 |
| M1600 | Psoriasis spondylitica | 0 |
| M1601 | Distal interphalangeal psoriatic arthropathy | 0 |
| M1602 | Arthritis mutilans | 0 |
| M160z | Psoriatic arthropathy NOS | 0 |
| N0452 | Juvenile arthritis in psoriasis | 0 |
| Nyu13 | [X]Other psoriatic arthropathies | 0 |

---

#### CONCEPT: PTSD (Post-traumatic stress disorders)¶

| READ\_CODE | DESCRIPTION\_SAIL | SENSITIVE\_CODE |
| --- | --- | --- |
| 1S1.. | Flashbacks | 0 |
| 1S10. | Short duration flashbacks | 0 |
| 1S11. | Long duration flashbacks | 0 |
| 1S12. | Episodic flashbacks | 0 |
| 1S13. | Duplicative flashbacks | 0 |
| E2831 | Acute posttrauma stress state | 0 |
| E29y1 | Other post-traumatic stress disorder | 0 |
| Eu431 | [X]Post - traumatic stress disorder | 0 |

---

#### CONCEPT: Pheripheral vascular disease¶

| READ\_CODE | DESCRIPTION\_SAIL | SENSITIVE\_CODE |
| --- | --- | --- |
| 14NB. | H/O: Peripheral vascular disease procedure | 0 |
| 1M110 | Ischaemic foot pain at rest | 0 |
| 1M111 | Ischaemic foot pain when walking | 0 |
| 2G63. | Ischaemic toe | 0 |
| 5593. | Femoral arteriogram abnormal | 0 |
| 55A2. | Lower limb arteriogram abnorm. | 0 |
| 727A4 | Scanning laser ophthalmoscopy evaluation of retina | 0 |
| 7A10. | Extraanatomic bypass of aorta | 0 |
| 7A100 | Emergency bypass of aorta by anastomosis of axillary artery to femoral artery | 0 |
| 7A101 | Bypass of aorta by anastomosis of axillary artery to femoral artery NEC | 0 |
| 7A102 | Axillo-bifemoral bypass graft | 0 |
| 7A103 | Axillo-unifemoral PTFE bypass graft | 0 |
| 7A104 | Bypass of aorta by anastomosis of axillary artery to bilateral femoral arteries | 0 |
| 7A10y | Other specified extraanatomic bypass of aorta | 0 |
| 7A10z | Extraanatomic bypass of aorta NOS | 0 |
| 7A192 | Open embolectomy of bifurcation of aorta | 0 |
| 7A4.. | Iliac and femoral artery operations | 0 |
| 7A40. | Replacement of aneurysmal iliac artery | 0 |
| 7A400 | Emergency replacement of aneurysmal iliac artery by anastomosis of iliac artery to femoral artery | 0 |
| 7A401 | Replacement of aneurysmal iliac artery by anastomosis of iliac artery to femoral artery NEC | 0 |
| 7A402 | Emergency replacement of aneurysmal iliac artery by anastomosis of femoral artery to femoral artery | 0 |
| 7A403 | Replacement of aneurysmal iliac artery by anastomosis of femoral artery to femoral artery NEC | 0 |
| 7A404 | Emergency replacement of aneurysmal common iliac artery by anastomosis of aorta to common iliac artery | 0 |
| 7A405 | Emergency replacement of aneurysmal iliac artery by anastomosis of aorta to external iliac artery | 0 |
| 7A406 | Emergency replacement of aneurysmal artery of leg by anastomosis of aorta to common femoral artery | 0 |
| 7A407 | Emergency replacement of aneurysmal artery of leg by anastomosis of aorta to superficial femoral artery | 0 |
| 7A408 | Emergency replacement of aneurysmal iliac artery by anastomosis of iliac artery to iliac artery | 0 |
| 7A409 | Replacement of aneurysmal common iliac artery by anastomosis of aorta to common iliac artery NEC | 0 |
| 7A40A | Replacement of aneurysmal iliac artery by anastomosis of aorta to external iliac artery NEC | 0 |
| 7A40B | Replacement of aneurysmal artery of leg by anastomosis of aorta to common femoral artery NEC | 0 |
| 7A40C | Replacement of aneurysmal artery of leg by anastomosis of aorta to superficial femoral artery NEC | 0 |
| 7A40D | Replacement of aneurysmal iliac artery by anastomosis of iliac artery to iliac artery NEC | 0 |
| 7A40y | Other specified replacement of aneurysmal iliac artery | 0 |
| 7A40z | Replacement of aneurysmal iliac artery NOS | 0 |
| 7A41. | Other bypass of iliac artery | 0 |
| 7A410 | Emergency bypass of iliac artery by anastomosis of iliac artery to femoral artery NEC | 0 |
| 7A411 | Bypass of iliac artery by anastomosis of iliac artery to femoral artery NEC | 0 |
| 7A412 | Emergency bypass of iliac artery by anastomosis of femoral artery to femoral artery NEC | 0 |
| 7A413 | Bypass of iliac artery by anastomosis of femoral artery to femoral artery NEC | 0 |
| 7A414 | Emergency bypass of common iliac artery by anastomosis of aorta to common iliac artery NEC | 0 |
| 7A415 | Emergency bypass of iliac artery by anastomosis of aorta to external iliac artery NEC | 0 |
| 7A416 | Emergency bypass of artery of leg by anastomosis of aorta to common femoral artery NEC | 0 |
| 7A417 | Emergency bypass of artery of leg by anastomosis of aorta to deep femoral artery NEC | 0 |
| 7A418 | Emergency bypass of iliac artery by anastomosis of iliac artery to iliac artery NEC | 0 |
| 7A419 | Bypass of common iliac artery by anastomosis of aorta to common iliac artery NEC | 0 |
| 7A41A | Bypass of iliac artery by anastomosis of aorta to external iliac NEC | 0 |
| 7A41B | Bypass of artery of leg by anastomosis of aorta to common femoral artery NEC | 0 |
| 7A41C | Bypass of artery of leg by anastomosis of aorta to deep femoral artery NEC | 0 |
| 7A41D | Bypass of iliac artery by anastomosis of iliac artery to iliac artery NEC | 0 |
| 7A41E | Emergency bypass of iliac artery by unspecified anastomosis | 0 |
| 7A41F | Ilio-femoral prosthetic cross over graft | 0 |
| 7A41y | Other specified other bypass of iliac artery | 0 |
| 7A41z | Other bypass of iliac artery NOS | 0 |
| 7A42. | Reconstruction of iliac artery | 0 |
| 7A420 | Endarterectomy of iliac artery and patch repair of iliac artery | 0 |
| 7A421 | Endarterectomy of iliac artery NEC | 0 |
| 7A42y | Other specified reconstruction of iliac artery | 0 |
| 7A42z | Reconstruction of iliac artery NOS | 0 |
| 7A43. | Other open operations on iliac artery | 0 |
| 7A430 | Repair of iliac artery NEC | 0 |
| 7A431 | Open embolectomy of iliac artery | 0 |
| 7A432 | Operation on aneurysm of iliac artery NEC | 0 |
| 7A433 | Open insertion of iliac artery stent | 0 |
| 7A43y | Other specified other open operation on iliac artery | 0 |
| 7A43z | Other open operation on iliac artery NOS | 0 |
| 7A44. | Transluminal operations on iliac artery | 0 |
| 7A440 | Percutaneous transluminal angioplasty of iliac artery | 0 |
| 7A441 | Percutaneous transluminal embolectomy of iliac artery | 0 |
| 7A442 | Arteriography of iliac artery | 0 |
| 7A443 | Insertion of iliac artery stent | 0 |
| 7A444 | Percutaneous transluminal insertion of iliac artery stent | 0 |
| 7A44y | Other specified transluminal operation on iliac artery | 0 |
| 7A44z | Transluminal operation on iliac artery NOS | 0 |
| 7A45. | Emergency replacement of aneurysmal femoral or popliteal artery | 0 |
| 7A450 | Emergency replacement of aneurysmal femoral artery by anastomosis of femoral artery to popliteal artery using prosthesis | 0 |
| 7A451 | Emergency replacement of aneurysmal popliteal artery by anastomosis of popliteal artery to popliteal artery using prosthesis | 0 |
| 7A452 | Emergency replacement of aneurysmal femoral artery by anastomosis of femoral artery to popliteal artery using vein graft | 0 |
| 7A453 | Emergency replacement of aneurysmal popliteal artery by anastomosis of popliteal artery to popliteal artery using vein graft | 0 |
| 7A454 | Emergency replacement of aneurysmal femoral artery by anastomosis of femoral artery to tibial artery using prosthesis | 0 |
| 7A455 | Emergency replacement of aneurysmal popliteal artery by anastomosis of popliteal artery to tibial artery using prosthesis | 0 |
| 7A456 | Emergency replacement of aneurysmal femoral artery by anastomosis of femoral artery to tibial artery using vein graft | 0 |
| 7A457 | Emergency replacement of aneurysmal popliteal artery by anastomosis of popliteal artery to tibial artery using vein graft | 0 |
| 7A458 | Emergency replacement of aneurysmal femoral artery by anastomosis of femoral artery to peroneal artery using prosthesis | 0 |
| 7A459 | Emergency replacement of aneurysmal popliteal artery by anastomosis of popliteal artery to peroneal artery using prosthesis | 0 |
| 7A45A | Emergency replacement of aneurysmal femoral artery by anastomosis of femoral artery to peroneal artery using vein graft | 0 |
| 7A45B | Emergency replacement of aneurysmal popliteal artery by anastomosis of popliteal artery to peroneal artery using vein graft | 0 |
| 7A45C | Emergency replacement of aneurysmal femoral artery by anastomosis of femoral artery to femoral artery | 0 |
| 7A45D | Emergency replacement of aneurysmal popliteal artery by anastomosis of popliteal artery to femoral artery | 0 |
| 7A45y | Other specified emergency replacement of aneurysmal femoral artery or popliteal artery | 0 |
| 7A45z | Emergency replacement of aneurysmal femoral artery or popliteal artery NOS | 0 |
| 7A46. | Other replacement of aneurysmal femoral artery | 0 |
| 7A460 | Replacement of aneurysmal femoral artery by anastomosis of femoral artery to popliteal artery using prosthesis NEC | 0 |
| 7A461 | Replacement of aneurysmal popliteal artery by anastomosis of popliteal artery to popliteal artery using prosthesis NEC | 0 |
| 7A462 | Replacement of aneurysmal femoral artery by anastomosis of femoral artery to popliteal artery using vein graft NEC | 0 |
| 7A463 | Replacement of aneurysmal popliteal artery by anastomosis of popliteal artery to popliteal artery using vein graft NEC | 0 |
| 7A464 | Replacement of aneurysmal femoral artery by anastomosis of femoral artery to tibial artery using prosthesis NEC | 0 |
| 7A465 | Replacement of aneurysmal popliteal artery by anastomosis of popliteal artery to tibial artery using prosthesis NEC | 0 |
| 7A466 | Replacement of aneurysmal femoral artery by anastomosis of femoral artery to tibial artery using vein graft NEC | 0 |
| 7A467 | Replacement of aneurysmal popliteal artery by anastomosis of popliteal artery to tibial artery using vein graft NEC | 0 |
| 7A468 | Replacement of aneurysmal femoral artery by anastomosis of femoral artery to peroneal artery using prosthesis NEC | 0 |
| 7A469 | Replacement of aneurysmal popliteal artery by anastomosis of popliteal artery to peroneal artery using prosthesis NEC | 0 |
| 7A46A | Replacement of aneurysmal femoral artery by anastomosis of femoral artery to peroneal artery using vein graft NEC | 0 |
| 7A46B | Replacement of aneurysmal popliteal artery by anastomosis of popliteal artery to peroneal artery using vein graft NEC | 0 |
| 7A46C | Replacement of aneurysmal femoral artery by anastomosis of femoral artery to femoral artery NEC | 0 |
| 7A46D | Replacement of aneurysmal popliteal artery by anastomosis of popliteal artery to femoral artery NEC | 0 |
| 7A46y | Other specified other replacement of aneurysmal femoral artery or popliteal artery | 0 |
| 7A46z | Other replacement of aneurysmal femoral artery or popliteal artery NOS | 0 |
| 7A47. | Other emergency bypass of femoral artery or popliteal artery | 0 |
| 7A470 | Emergency bypass of femoral artery by anastomosis of femoral artery to popliteal artery using prosthesis NEC | 0 |
| 7A471 | Emergency bypass of popliteal artery by anastomosis of popliteal artery to popliteal artery using prosthesis NEC | 0 |
| 7A472 | Emergency bypass of femoral artery by anastomosis of femoral artery to popliteal artery using vein graft NEC | 0 |
| 7A473 | Emergency bypass of popliteal artery by anastomosis of popliteal artery to popliteal artery using vein graft NEC | 0 |
| 7A474 | Emergency bypass of femoral artery by anastomosis of femoral artery to tibial artery using prosthesis NEC | 0 |
| 7A475 | Emergency bypass of popliteal artery by anastomosis of popliteal artery to tibial artery using prosthesis NEC | 0 |
| 7A476 | Emergency bypass of femoral artery by anastomosis of femoral artery to tibial artery using vein graft NEC | 0 |
| 7A477 | Emergency bypass of popliteal artery by anastomosis of popliteal artery to tibial artery using vein graft NEC | 0 |
| 7A478 | Emergency bypass of femoral artery by anastomosis of femoral artery to peroneal artery using prosthesis NEC | 0 |
| 7A479 | Emergency bypass of popliteal artery by anastomosis of popliteal artery to peroneal artery using prosthesis NEC | 0 |
| 7A47A | Emergency bypass of femoral artery by anastomosis of femoral artery to peroneal artery using vein graft NEC | 0 |
| 7A47B | Emergency bypass of popliteal artery by anastomosis of popliteal artery to peroneal artery using vein graft NEC | 0 |
| 7A47C | Emergency bypass of femoral artery by anastomosis of femoral artery to femoral artery NEC | 0 |
| 7A47D | Emergency bypass of popliteal artery by anastomosis of popliteal artery to femoral artery NEC | 0 |
| 7A47y | Other specified other emergency bypass of femoral artery or popliteal artery | 0 |
| 7A47z | Other emergency bypass of femoral artery or popliteal artery NOS | 0 |
| 7A48. | Other bypass of femoral artery or popliteal artery | 0 |
| 7A480 | Bypass of femoral artery by anastomosis of femoral artery to popliteal artery using prosthesis NEC | 0 |
| 7A481 | Bypass of popliteal artery by anastomosis of popliteal artery to popliteal artery using prosthesis NEC | 0 |
| 7A482 | Bypass of femoral artery by anastomosis of femoral artery to popliteal artery using vein graft NEC | 0 |
| 7A483 | Bypass of popliteal artery by anastomosis of popliteal artery to popliteal artery using vein graft NEC | 0 |
| 7A484 | Bypass of femoral artery by anastomosis of femoral artery to tibial artery using prosthesis NEC | 0 |
| 7A485 | Bypass of popliteal artery by anastomosis of popliteal artery to tibial artery using prosthesis NEC | 0 |
| 7A486 | Bypass of femoral artery by anastomosis of femoral artery to tibial artery using vein graft NEC | 0 |
| 7A487 | Bypass of popliteal artery by anastomosis of popliteal artery to tibial artery using vein graft NEC | 0 |
| 7A488 | Bypass of femoral artery by anastomosis of femoral artery to peroneal artery using prosthesis NEC | 0 |
| 7A489 | Bypass of popliteal artery by anastomosis of popliteal artery to peroneal artery using prosthesis NEC | 0 |
| 7A48A | Bypass of femoral artery by anastomosis of femoral artery to peroneal artery using vein graft NEC | 0 |
| 7A48B | Bypass of popliteal artery by anastomosis of popliteal artery to peroneal artery using vein graft NEC | 0 |
| 7A48C | Bypass of femoral artery by anastomosis of femoral artery to femoral artery NEC | 0 |
| 7A48D | Bypass of popliteal artery by anastomosis of popliteal artery to femoral artery NEC | 0 |
| 7A48E | Femoro-femoral prosthetic cross over graft | 0 |
| 7A48y | Other specified other bypass of femoral artery or popliteal artery | 0 |
| 7A48z | Other bypass of femoral artery or popliteal artery NOS | 0 |
| 7A49. | Reconstruction of femoral artery or popliteal artery | 0 |
| 7A490 | Endarterectomy of femoral artery and patch repair of femoral artery | 0 |
| 7A491 | Endarterectomy of popliteal artery and patch repair of popliteal artery | 0 |
| 7A492 | Endarterectomy of femoral artery NEC | 0 |
| 7A493 | Endarterectomy of popliteal artery NEC | 0 |
| 7A494 | Profundoplasty of femoral artery and patch repair of deep femoral artery | 0 |
| 7A495 | Profundoplasty of popliteal artery and patch repair of popliteal artery | 0 |
| 7A496 | Profundoplasty of femoral artery NEC | 0 |
| 7A497 | Profundoplasty of popliteal artery NEC | 0 |
| 7A498 | Reconstruction of femoral artery with vein graft | 0 |
| 7A499 | Reconstruction of popliteal artery with vein graft | 0 |
| 7A49y | Other specified reconstruction of femoral artery or popliteal artery | 0 |
| 7A49z | Reconstruction of femoral or popliteal artery NOS | 0 |
| 7A4A. | Other open operations on femoral artery or popliteal artery | 0 |
| 7A4A0 | Repair of femoral artery NEC | 0 |
| 7A4A1 | Repair of popliteal artery NEC | 0 |
| 7A4A2 | Open embolectomy of femoral artery | 0 |
| 7A4A3 | Open embolectomy popliteal artery | 0 |
| 7A4A4 | Ligation of aneurysm of popliteal artery | 0 |
| 7A4A5 | Operation on aneurysm of femoral artery NEC | 0 |
| 7A4A6 | Operation on popliteal artery NEC | 0 |
| 7A4A7 | Repair of femoral artery with temporary silastic shunt | 0 |
| 7A4A8 | Repair of popliteal artery with temporary silastic shunt | 0 |
| 7A4Ay | Other specified other open operation on femoral artery or popliteal artery | 0 |
| 7A4Az | Other open operation on femoral artery or popliteal artery NOS | 0 |
| 7A4B. | Transluminal operations on femoral artery or popliteal artery | 0 |
| 7A4B0 | Percutaneous transluminal angioplasty of femoral artery | 0 |
| 7A4B1 | Percutaneous transluminal angioplasty of popliteal artery | 0 |
| 7A4B2 | Percutaneous transluminal embolectomy of femoral artery | 0 |
| 7A4B3 | Percutaneous transluminal embolectomy of popliteal artery | 0 |
| 7A4B4 | Percutaneous transluminal embolisation of femoral artery | 0 |
| 7A4B5 | Percutaneous transluminal embolisation of popliteal artery | 0 |
| 7A4B6 | Arteriography of femoral artery | 0 |
| 7A4B7 | Arteriography of popliteal artery | 0 |
| 7A4B8 | Percutaneous transluminal thrombolysis of femoral graft using streptokinase | 0 |
| 7A4B9 | Percutaneous transluminal insertion of stent into femoral artery | 0 |
| 7A4By | Other specified transluminal operation on femoral artery or popliteal artery | 0 |
| 7A4Bz | Transluminal operation on femoral artery or popliteal artery NOS | 0 |
| 7A4y. | Other specified operations on iliac and femoral artery | 0 |
| 7A4z. | Iliac and femoral artery operations NOS | 0 |
| 7A50. | Revision of reconstruction of artery | 0 |
| 7A500 | Revision of reconstruction involving aorta | 0 |
| 7A501 | Revision of reconstruction involving iliac artery | 0 |
| 7A502 | Revision of reconstruction involving femoral artery | 0 |
| 7A503 | Revision of reconstruction of popliteal artery | 0 |
| 7A50y | Other specified revision of reconstruction of artery | 0 |
| 7A50z | Revision of reconstruction of artery NOS | 0 |
| C107. | Diabetes mellitus with peripheral circulatory disorder | 0 |
| C1070 | Diabetes mellitus, juvenile type, with peripheral circulatory disorder | 0 |
| C1071 | Diabetes mellitus, adult onset, with peripheral circulatory disorder | 0 |
| C1072 | Diabetes mellitus, adult with gangrene | 0 |
| C1073 | IDDM with peripheral circulatory disorder | 0 |
| C1074 | NIDDM with peripheral circulatory disorder | 0 |
| C107z | Diabetes mellitus NOS with peripheral circulatory disorder | 0 |
| C1086 | Insulin dependent diabetes mellitus with gangrene | 0 |
| C108G | Insulin dependent diabetes mellitus with peripheral angiopathy | 0 |
| C1095 | Non-insulin dependent diabetes mellitus with gangrene | 0 |
| C109F | Non-insulin-dependent diabetes mellitus with peripheral angiopathy | 0 |
| C10E6 | Type 1 diabetes mellitus with gangrene | 0 |
| C10EG | Type 1 diabetes mellitus with peripheral angiopathy | 0 |
| C10F5 | Type 2 diabetes mellitus with gangrene | 0 |
| C10FF | Type 2 diabetes mellitus with peripheral angiopathy | 0 |
| G702. | Extremity artery atheroma | 0 |
| G702z | Extremity artery atheroma NOS | 0 |
| G73.. | Other peripheral vascular disease | 0 |
| G730. | Raynaud's syndrome | 0 |
| G7300 | Raynaud's disease | 0 |
| G7301 | Raynaud's phenomenon | 0 |
| G730z | Raynaud's syndrome NOS | 0 |
| G731. | Thromboangiitis obliterans | 0 |
| G7310 | Buerger's disease | 0 |
| G7311 | Presenile gangrene | 0 |
| G731z | Thromboangiitis obliterans NOS | 0 |
| G732. | Peripheral gangrene | 0 |
| G7320 | Gangrene of toe | 0 |
| G7321 | Gangrene of foot | 0 |
| G7322 | Gangrene of finger | 0 |
| G7323 | Gangrene of thumb | 0 |
| G7324 | Gangrene of hand | 0 |
| G733. | Ischaemic foot | 0 |
| G734. | Peripheral arterial disease | 0 |
| G73y. | Other specified peripheral vascular disease | 0 |
| G73y0 | Diabetic peripheral angiopathy | 0 |
| G73y1 | Peripheral angiopathic disease EC NOS | 0 |
| G73y2 | Acrocyanosis | 0 |
| G73y4 | Acroparaesthesia - Schultze's type | 0 |
| G73y5 | Acroparaesthesia - Nothnagel's type | 0 |
| G73y6 | Acroparaesthesia - unspecified | 0 |
| G73y7 | Erythrocyanosis | 0 |
| G73y8 | Erythromelalgia | 0 |
| G73yz | Other specified peripheral vascular disease NOS | 0 |
| G73z. | Peripheral vascular disease NOS | 0 |
| G73z0 | Intermittent claudication | 0 |
| G73zz | Peripheral vascular disease NOS | 0 |
| G742z | Peripheral arterial embolism and thrombosis NOS | 0 |
| G830. | Varicose veins of the leg with ulcer | 0 |
| G831. | Varicose veins of the leg with eczema | 0 |
| G832. | Varicose veins of the leg with ulcer and eczema | 0 |
| G833. | Varicose veins of the leg with rupture | 0 |
| G835. | Infected varicose ulcer | 0 |
| G836. | Varicose vein of leg with phlebitis | 0 |
| G837. | Venous ulcer of leg | 0 |
| G8y1. | Postphlebitic syndrome | 0 |
| G8y3. | Chronic peripheral venous hypertension | 0 |
| G8yy0 | Chronic venous insufficiency NOS | 0 |
| Gyu74 | [X]Other specified peripheral vascular diseases | 0 |
| Gyu7A | [X]Peripheral angiopathy in diseases classified elsewhere | 0 |
| M271. | Non-pressure ulcer lower limb | 0 |
| M2710 | Ischaemic ulcer diabetic foot | 0 |
| P76.. | Other peripheral vascular system anomalies | 0 |
| R0542 | [D]Gangrene of toe in diabetic | 0 |
| R0543 | [D]Widespread diabetic foot gangrene | 0 |

---

#### CONCEPT: Renal transplant¶

| READ\_CODE | DESCRIPTION\_SAIL | SENSITIVE\_CODE |
| --- | --- | --- |
| 7B00. | Transplantation of kidney | 0 |
| 7B000 | Autotransplant of kidney | 0 |
| 7B001 | Transplantation of kidney from live donor | 0 |
| 7B002 | Transplantation of kidney from cadaver | 0 |
| 7B003 | Allotransplantation of kidney from cadaver, heart-beating | 0 |
| 7B004 | Allotransplantation of kidney from cadaver, heart non-beating | 0 |
| 7B005 | Allotransplantation of kidney from cadaver NEC | 0 |
| 7B006 | Xenograft renal transplant | 0 |
| 7B00y | Other specified transplantation of kidney | 0 |
| 7B00z | Transplantation of kidney NOS | 0 |
| 7B015 | Transplant nephrectomy | 0 |
| 7B019 | Excision of rejected transplanted kidney | 0 |
| 7B063 | Exploration of renal transplant | 0 |
| 7B0F. | Interventions associated with transplantation of kidney | 0 |
| 7B0F3 | Post-transplantation of kidney examination, recipient | 0 |
| 7B0F4 | Post-transplantation of kidney examination, live donor | 0 |
| 7B0Fy | Other specified interventions associated with transplantation of kidney | 0 |
| 7B0Fz | Interventions associated with transplantation of kidney NOS | 0 |
| 7L020 | Autotransplantation of adrenal medulla to caudate nucleus of brain | 0 |
| K0B5. | Renal tubulo-interstitial disorders in transplant rejection | 0 |
| Kyu1C | [X]Renal tubulo-interstitial disorders in transplant rejection | 0 |
| SP081 | Transplanted organ rejection | 0 |
| SP083 | Kidney transplant failure and rejection | 0 |
| SP08C | Accelerated rejection of renal transplant | 0 |
| SP08D | Acute-on-chronic rejection of renal transplant | 0 |
| SP08E | Acute rejection of renal transplant - grade I | 0 |
| SP08F | Acute rejection of renal transplant - grade II | 0 |
| SP08G | Acute rejection of renal transplant - grade III | 0 |
| SP08H | Acute rejection of renal transplant | 0 |
| SP08J | Chronic rejection of renal transplant | 0 |
| SP08K | Chronic rejection of renal transplant - grade 1 | 0 |
| SP08L | Chronic rejection of renal transplant - grade II | 0 |
| SP08M | Chronic rejection of renal transplant - grade III | 0 |
| SP08N | Unexplained episode of renal transplant dysfunction | 0 |
| SP08P | Stenosis of vein of transplanted kidney | 0 |
| SP08Q | Aneurysm of artery of transplanted kidney | 0 |
| SP08R | Renal transplant rejection | 0 |
| SP08S | Aneurysm of vein of transplanted kidney | 0 |
| SP08T | Urological complication of renal transplant | 0 |
| SP08V | Very mild acute rejection of renal transplant | 0 |
| SP08W | Vascular complication of renal transplant | 0 |
| SP08X | Rupture of artery of transplanted kidney | 0 |
| SP08Y | Rupture of vein of transplanted kidney | 0 |
| SP08Z | Thrombosis of artery of transplanted kidney | 0 |
| SP08a | Thrombosis of vein of transplanted kidney | 0 |
| TB001 | Transplantation of kidney as the cause of abnormal reaction of patient, or of later complication, without mention of misadventure at the time of operation | 0 |
| ZV420 | [V]Kidney transplanted | 0 |

---

#### CONCEPT: Rheumatoid Arthritis¶

| READ\_CODE | DESCRIPTION\_SAIL | SENSITIVE\_CODE |
| --- | --- | --- |
| 14G1. | H/O: rheumatoid arthritis | 0 |
| 38DZ. | Disease activity score in rheumatoid arthritis | 0 |
| 38DZ0 | Disease activity score 28 joint in rheumatoid arthritis | 0 |
| 66HB0 | Rheumatoid arthritis annual review | 0 |
| 7P203 | Delivery of rehabilitation for rheumatoid arthritis | 0 |
| 9hR.. | Exception reporting: rheumatoid arthritis quality indicators | 0 |
| 9hR0. | Excepted from rheumatoid arthritis quality indicators: patient unsuitable | 0 |
| 9hR1. | Excepted from rheumatoid arthritis quality indicators: informed dissent | 0 |
| 9mM.. | Rheumatoid arthritis monitoring invitation | 0 |
| 9mM0. | Rheumatoid arthritis monitoring invitation first letter | 0 |
| 9mM1. | Rheumatoid arthritis monitoring invitation second letter | 0 |
| 9mM2. | Rheumatoid arthritis monitoring invitation third letter | 0 |
| 9mM3. | Rheumatoid arthritis monitoring verbal invitation | 0 |
| 9mM4. | Rheumatoid arthritis monitoring telephone invitation | 0 |
| F3712 | Polyneuropathy in rheumatoid arthritis | 0 |
| F3964 | Myopathy due to rheumatoid arthritis | 0 |
| H570. | Rheumatoid lung | 0 |
| N04.. | Rheumatoid arthritis and other inflammatory polyarthropathies | 0 |
| N040. | Rheumatoid arthritis | 0 |
| N0400 | Rheumatoid arthritis of cervical spine | 0 |
| N0401 | Other rheumatoid arthritis of spine | 0 |
| N0402 | Rheumatoid arthritis of shoulder | 0 |
| N0403 | Rheumatoid arthritis of sternoclavicular joint | 0 |
| N0404 | Rheumatoid arthritis of acromioclavicular joint | 0 |
| N0405 | Rheumatoid arthritis of elbow | 0 |
| N0406 | Rheumatoid arthritis of distal radio-ulnar joint | 0 |
| N0407 | Rheumatoid arthritis of wrist | 0 |
| N0408 | Rheumatoid arthritis of metacarpophalangeal joint | 0 |
| N0409 | Rheumatoid arthritis of proximal interphalangeal joint of finger | 0 |
| N040A | Rheumatoid arthritis of distal interphalangeal joint of finger | 0 |
| N040B | Rheumatoid arthritis of hip | 0 |
| N040C | Rheumatoid arthritis of sacro-iliac joint | 0 |
| N040D | Rheumatoid arthritis of knee | 0 |
| N040E | Rheumatoid arthritis of tibio-fibular joint | 0 |
| N040F | Rheumatoid arthritis of ankle | 0 |
| N040G | Rheumatoid arthritis of subtalar joint | 0 |
| N040H | Rheumatoid arthritis of talonavicular joint | 0 |
| N040J | Rheumatoid arthritis of other tarsal joint | 0 |
| N040K | Rheumatoid arthritis of 1st metatarsophalangeal joint | 0 |
| N040L | Rheumatoid arthritis of lesser metatarsophalangeal joint | 0 |
| N040M | Rheumatoid arthritis of interphalangeal joint of toe | 0 |
| N040N | Rheumatoid vasculitis | 0 |
| N040P | Seronegative rheumatoid arthritis | 0 |
| N040Q | Rheumatoid bursitis | 0 |
| N040R | Rheumatoid nodule | 0 |
| N040S | Rheumatoid arthritis - multiple joint | 0 |
| N040T | Flare of rheumatoid arthritis | 0 |
| N041. | Felty's syndrome | 0 |
| N0421 | Rheumatoid lung disease | 0 |
| N0422 | Rheumatoid nodule | 0 |
| N047. | Seropositive errosive rheumatoid arthritis | 0 |
| N04X. | Seropositive rheumatoid arthritis, unspecified | 0 |
| N04y0 | Rheumatoid lung | 0 |
| Nyu10 | [X]Rheumatoid arthritis with involvement of other organs or systems | 0 |
| Nyu11 | [X]Other seropositive rheumatoid arthritis | 0 |
| Nyu12 | [X]Other specified rheumatoid arthritis | 0 |
| Nyu1G | [X]Seropositive rheumatoid arthritis, unspecified | 0 |

---

#### CONCEPT: Schizophrenia¶

| READ\_CODE | DESCRIPTION\_SAIL | SENSITIVE\_CODE |
| --- | --- | --- |
| 1284. | FH: Schizophrenia | 0 |
| 13L3. | Relative - psychotic illness | 0 |
| 13Y2. | Schizophrenia association member | 0 |
| 1464. | H/O: schizophrenia | 0 |
| 212T. | Psychosis, schizophrenia and bipolar affective disorder resolved | 0 |
| 212W. | Schizophrenia resolved | 0 |
| E10.. | Schizophrenic disorders | 0 |
| E100. | Simple schizophrenia | 0 |
| E1000 | Unspecified schizophrenia | 0 |
| E1001 | Subchronic schizophrenia | 0 |
| E1002 | Chronic schizophrenic | 0 |
| E1003 | Acute exacerbation of subchronic schizophrenia | 0 |
| E1004 | Acute exacerbation of chronic schizophrenia | 0 |
| E1005 | Schizophrenia in remission | 0 |
| E100z | Simple schizophrenia NOS | 0 |
| E101. | Hebephrenic schizophrenia | 0 |
| E1010 | Unspecified hebephrenic schizophrenia | 0 |
| E1011 | Subchronic hebephrenic schizophrenia | 0 |
| E1012 | Chronic hebephrenic schizophrenia | 0 |
| E1013 | Acute exacerbation of subchronic hebephrenic schizophrenia | 0 |
| E1014 | Acute exacerbation of chronic hebephrenic schizophrenia | 0 |
| E1015 | Hebephrenic schizophrenia in remission | 0 |
| E101z | Hebephrenic schizophrenia NOS | 0 |
| E102. | Catatonic schizophrenia | 0 |
| E1020 | Unspecified catatonic schizophrenia | 0 |
| E1021 | Subchronic catatonic schizophrenia | 0 |
| E1022 | Chronic catatonic schizophrenia | 0 |
| E1023 | Acute exacerbation of subchronic catatonic schizophrenia | 0 |
| E1024 | Acute exacerbation of chronic catatonic schizophrenia | 0 |
| E1025 | Catatonic schizophrenia in remission | 0 |
| E102z | Catatonic schizophrenia NOS | 0 |
| E103. | Paranoid schizophrenia | 0 |
| E1030 | Unspecified paranoid schizophrenia | 0 |
| E1031 | Subchronic paranoid schizophrenia | 0 |
| E1032 | Chronic paranoid schizophrenia | 0 |
| E1033 | Acute exacerbation of subchronic paranoid schizophrenia | 0 |
| E1034 | Acute exacerbation of chronic paranoid schizophrenia | 0 |
| E1035 | Paranoid schizophrenia in remission | 0 |
| E103z | Paranoid schizophrenia NOS | 0 |
| E104. | Acute schizophrenic episode | 0 |
| E105. | Latent schizophrenia | 0 |
| E1050 | Unspecified latent schizophrenia | 0 |
| E1051 | Subchronic latent schizophrenia | 0 |
| E1052 | Chronic latent schizophrenia | 0 |
| E1053 | Acute exacerbation of subchronic latent schizophrenia | 0 |
| E1054 | Acute exacerbation of chronic latent schizophrenia | 0 |
| E1055 | Latent schizophrenia in remission | 0 |
| E105z | Latent schizophrenia NOS | 0 |
| E106. | Residual schizophrenia | 0 |
| E107. | Schizo-affective schizophrenia | 0 |
| E1070 | Unspecified schizo-affective schizophrenia | 0 |
| E1071 | Subchronic schizo-affective schizophrenia | 0 |
| E1072 | Chronic schizo-affective schizophrenia | 0 |
| E1073 | Acute exacerbation of subchronic schizo-affective schizophrenia | 0 |
| E1074 | Acute exacerbation of chronic schizo-affective schizophrenia | 0 |
| E1075 | Schizo-affective schizophrenia in remission | 0 |
| E107z | Schizo-affective schizophrenia NOS | 0 |
| E10y. | Other schizophrenia | 0 |
| E10y0 | Atypical schizophrenia | 0 |
| E10y1 | Coenesthopathic schizophrenia | 0 |
| E10yz | Other schizophrenia NOS | 0 |
| E10z. | Schizophrenia NOS | 0 |
| E14z. | Child psychosis NOS | 0 |
| Eu052 | [X]Organic delusional [schizophrenia-like] disorder | 0 |
| Eu2.. | [X]Schizophrenia, schizotypal and delusional disorders | 0 |
| Eu20. | [X]Schizophrenia | 0 |
| Eu200 | [X]Paranoid schizophrenia | 0 |
| Eu201 | [X]Hebephrenic schizophrenia | 0 |
| Eu202 | [X]Catatonic schizophrenia | 0 |
| Eu203 | [X]Undifferentiated schizophrenia | 0 |
| Eu204 | [X]Post-schizophrenic depression | 0 |
| Eu205 | [X]Residual schizophrenia | 0 |
| Eu206 | [X]Simple schizophrenia | 0 |
| Eu20y | [X]Other schizophrenia | 0 |
| Eu20z | [X]Schizophrenia, unspecified | 0 |
| Eu21. | [X]Schizotypal disorder | 0 |
| Eu231 | [X]Acute polymorphic psychotic disorder with symptoms of schizophrenia | 0 |
| Eu232 | [X]Acute schizophrenia-like psychotic disorder | 0 |
| Eu252 | [X]Schizoaffective disorder, mixed type | 0 |
| ZV110 | [V]Personal history of schizophrenia | 0 |

---

#### CONCEPT: Skin Cancer¶

| READ\_CODE | DESCRIPTION\_SAIL | SENSITIVE\_CODE |
| --- | --- | --- |
| 14250 | H/O Malignant melanoma | 0 |
| 4M70. | Clark melanoma level 1 | 0 |
| 4M71. | Clark melanoma level 2 | 0 |
| 4M72. | Clark melanoma level 3 | 0 |
| 4M73. | Clark melanoma level 4 | 0 |
| 4M74. | Clark melanoma level 5 | 0 |
| 7G03J | Excision of melanoma | 0 |
| 7G03K | Excision malignant skin tumour | 0 |
| 7G056 | Excision biopsy of rodent ulcer | 0 |
| B3... | Malignant neoplasm of bone, connective tissue, skin and breast | 0 |
| B3104 | Malignant neoplasm of tarsus of eyelid | 0 |
| B32.. | Malignant melanoma of skin | 0 |
| B320. | Malignant melanoma of lip | 0 |
| B321. | Malignant melanoma of eyelid including canthus | 0 |
| B322. | Malignant melanoma of ear and external auricular canal | 0 |
| B3220 | Malignant melanoma of auricle (ear) | 0 |
| B3221 | Malignant melanoma of external auditory meatus | 0 |
| B322z | Malignant melanoma of ear and external auricular canal NOS | 0 |
| B323. | Malignant melanoma of other and unspecified parts of face | 0 |
| B3230 | Malignant melanoma of external surface of cheek | 0 |
| B3231 | Malignant melanoma of chin | 0 |
| B3232 | Malignant melanoma of eyebrow | 0 |
| B3233 | Malignant melanoma of forehead | 0 |
| B3234 | Malignant melanoma of external surface of nose | 0 |
| B3235 | Malignant melanoma of temple | 0 |
| B323z | Malignant melanoma of face NOS | 0 |
| B324. | Malignant melanoma of scalp and neck | 0 |
| B3240 | Malignant melanoma of scalp | 0 |
| B3241 | Malignant melanoma of neck | 0 |
| B324z | Malignant melanoma of scalp and neck NOS | 0 |
| B325. | Malignant melanoma of trunk (excluding scrotum) | 0 |
| B3250 | Malignant melanoma of axilla | 0 |
| B3251 | Malignant melanoma of breast | 0 |
| B3252 | Malignant melanoma of buttock | 0 |
| B3253 | Malignant melanoma of groin | 0 |
| B3254 | Malignant melanoma of perianal skin | 0 |
| B3255 | Malignant melanoma of perineum | 0 |
| B3256 | Malignant melanoma of umbilicus | 0 |
| B3257 | Malignant melanoma of back | 0 |
| B3258 | Malignant melanoma of chest wall | 0 |
| B325z | Malignant melanoma of trunk, excluding scrotum, NOS | 0 |
| B326. | Malignant melanoma of upper limb and shoulder | 0 |
| B3260 | Malignant melanoma of shoulder | 0 |
| B3261 | Malignant melanoma of upper arm | 0 |
| B3262 | Malignant melanoma of fore-arm | 0 |
| B3263 | Malignant melanoma of hand | 0 |
| B3264 | Malignant melanoma of finger | 0 |
| B3265 | Malignant melanoma of thumb | 0 |
| B326z | Malignant melanoma of upper limb or shoulder NOS | 0 |
| B327. | Malignant melanoma of lower limb and hip | 0 |
| B3270 | Malignant melanoma of hip | 0 |
| B3271 | Malignant melanoma of thigh | 0 |
| B3272 | Malignant melanoma of knee | 0 |
| B3273 | Malignant melanoma of popliteal fossa area | 0 |
| B3274 | Malignant melanoma of lower leg | 0 |
| B3275 | Malignant melanoma of ankle | 0 |
| B3276 | Malignant melanoma of heel | 0 |
| B3277 | Malignant melanoma of foot | 0 |
| B3278 | Malignant melanoma of toe | 0 |
| B3279 | Malignant melanoma of great toe | 0 |
| B327z | Malignant melanoma of lower limb or hip NOS | 0 |
| B328. | Malignant melanoma stage IA | 0 |
| B329. | Malignant melanoma stage IB | 0 |
| B32A. | Malignant melanoma stage IIA | 0 |
| B32B. | Malignant melanoma stage IIB | 0 |
| B32C. | Malignant melanoma stage IIC | 0 |
| B32D. | Malignant melanoma stage IIIA | 0 |
| B32E. | Malignant melanoma stage IIIB | 0 |
| B32F. | Malignant melanoma stage IIIC | 0 |
| B32G. | Malignant melanoma stage IV M1a | 0 |
| B32H. | Malignant melanoma stage IV M1b | 0 |
| B32J. | Malignant melanoma stage IV M1c | 0 |
| B32y. | Malignant melanoma of other specified skin site | 0 |
| B32y0 | Overlapping malignant melanoma of skin | 0 |
| B32z. | Malignant melanoma of skin NOS | 0 |
| B33.. | Other malignant neoplasm of skin | 0 |
| B330. | Malignant neoplasm of skin of lip | 0 |
| B331. | Malignant neoplasm of eyelid including canthus | 0 |
| B3311 | Malignant neoplasm of upper eyelid | 0 |
| B3312 | Malignant neoplasm of lower eyelid | 0 |
| B332. | Malignant neoplasm of skin of ear and external auricular canal | 0 |
| B3320 | Malignant neoplasm of skin of auricle (ear) | 0 |
| B3321 | Malignant neoplasm of skin of external auditory meatus | 0 |
| B3322 | Malignant neoplasm of pinna NEC | 0 |
| B332z | Malignant neoplasm of skin of ear and external auricular canal NOS | 0 |
| B333. | Malignant neoplasm of skin of other and unspecified parts of face | 0 |
| B3330 | Malignant neoplasm of skin of cheek, external | 0 |
| B3331 | Malignant neoplasm of skin of chin | 0 |
| B3332 | Malignant neoplasm of skin of eyebrow | 0 |
| B3333 | Malignant neoplasm of skin of forehead | 0 |
| B3334 | Malignant neoplasm of skin of nose (external) | 0 |
| B3335 | Malignant neoplasm of skin of temple | 0 |
| B333z | Malignant neoplasm of skin of other and unspecified part of face NOS | 0 |
| B334. | Malignant neoplasm of scalp and skin of neck | 0 |
| B3340 | Malignant neoplasm of scalp | 0 |
| B3341 | Malignant neoplasm of skin of neck | 0 |
| B334z | Malignant neoplasm of scalp or skin of neck NOS | 0 |
| B335. | Malignant neoplasm of skin of trunk, excluding scrotum | 0 |
| B3350 | Malignant neoplasm of skin of axillary fold | 0 |
| B3351 | Malignant neoplasm of skin of chest, excluding breast | 0 |
| B3352 | Malignant neoplasm of skin of breast | 0 |
| B3353 | Malignant neoplasm of skin of abdominal wall | 0 |
| B3354 | Malignant neoplasm of skin of umbilicus | 0 |
| B3355 | Malignant neoplasm of skin of groin | 0 |
| B3356 | Malignant neoplasm of skin of perineum | 0 |
| B3357 | Malignant neoplasm of skin of back | 0 |
| B3358 | Malignant neoplasm of skin of buttock | 0 |
| B3359 | Malignant neoplasm of perianal skin | 0 |
| B335A | Malignant neoplasm of skin of scapular region | 0 |
| B335z | Malignant neoplasm of skin of trunk, excluding scrotum, NOS | 0 |
| B336. | Malignant neoplasm of skin of upper limb and shoulder | 0 |
| B3360 | Malignant neoplasm of skin of shoulder | 0 |
| B3361 | Malignant neoplasm of skin of upper arm | 0 |
| B3362 | Malignant neoplasm of skin of fore-arm | 0 |
| B3363 | Malignant neoplasm of skin of hand | 0 |
| B3364 | Malignant neoplasm of skin of finger | 0 |
| B3365 | Malignant neoplasm of skin of thumb | 0 |
| B336z | Malignant neoplasm of skin of upper limb or shoulder NOS | 0 |
| B337. | Malignant neoplasm of skin of lower limb and hip | 0 |
| B3370 | Malignant neoplasm of skin of hip | 0 |
| B3371 | Malignant neoplasm of skin of thigh | 0 |
| B3372 | Malignant neoplasm of skin of knee | 0 |
| B3373 | Malignant neoplasm of skin of popliteal fossa area | 0 |
| B3374 | Malignant neoplasm of skin of lower leg | 0 |
| B3375 | Malignant neoplasm of skin of ankle | 0 |
| B3376 | Malignant neoplasm of skin of heel | 0 |
| B3377 | Malignant neoplasm of skin of foot | 0 |
| B3378 | Malignant neoplasm of skin of toe | 0 |
| B3379 | Malignant neoplasm of skin of great toe | 0 |
| B337z | Malignant neoplasm of skin of lower limb or hip NOS | 0 |
| B338. | Squamous cell carcinoma of skin | 0 |
| B339. | Dermatofibrosarcoma protuberans | 0 |
| B33X. | Malignant neoplasm overlapping lesion of skin | 0 |
| B33y. | Malignant neoplasm of other specified skin sites | 0 |
| B33z. | Malignant neoplasm of skin NOS | 0 |
| B33z0 | Kaposi's sarcoma of skin | 0 |
| B3y.. | Malignant neoplasm of bone, connective tissue, skin and breast otherwise specified | 0 |
| B3z.. | Malignant neoplasm of bone, connective tissue, skin and breast NOS | 0 |
| B480. | Malignant neoplasm of prepuce (foreskin) | 0 |
| B509. | Malignant melanoma of eye | 0 |
| B582. | Secondary malignant neoplasm of skin | 0 |
| B5820 | Secondary malignant neoplasm of skin of head | 0 |
| B5821 | Secondary malignant neoplasm of skin of face | 0 |
| B5822 | Secondary malignant neoplasm of skin of neck | 0 |
| B5823 | Secondary malignant neoplasm of skin of trunk | 0 |
| B5824 | Secondary malignant neoplasm of skin of shoulder and arm | 0 |
| B5825 | Secondary malignant neoplasm of skin of hip and leg | 0 |
| B5826 | Secondary malignant neoplasm of skin of breast | 0 |
| B582z | Secondary malignant neoplasm of skin NOS | 0 |
| B6286 | Cutaneous follicle centre lymphoma | 0 |
| B62E3 | Cutaneous T-cell lymphoma | 0 |
| B62xX | Other and unspecified peripheral and cutaneous T-cell lymphomas | 0 |
| B932. | Neoplasm of uncertain behaviour of skin | 0 |
| BA02. | Neoplasm of unspecified nature of bone, skin and soft tissue | 0 |
| BA022 | Neoplasm of unspecified nature of skin | 0 |
| BA02z | Neoplasm of unspecified nature of bone, skin or soft tissue NOS | 0 |
| BB2.. | [M]Papillary and squamous cell neoplasms | 0 |
| BB2A. | [M]Squamous cell carcinoma NOS | 0 |
| BB36. | [M]Metatypical carcinoma | 0 |
| BB5RA | [M]Merkel cell carcinoma | 0 |
| BB6.. | [M]Adnexal and skin appendage neoplasms | 0 |
| BB60. | [M]Skin appendage adenoma and carcinoma | 0 |
| BB601 | [M]Skin appendage carcinoma | 0 |
| BB60z | [M]Skin appendage adenoma or carcinoma NOS | 0 |
| BB611 | [M]Sweat gland tumour NOS | 0 |
| BB62. | [M]Apocrine adenoma and adenocarcinomas | 0 |
| BB621 | [M]Apocrine adenocarcinoma | 0 |
| BB62z | [M]Apocrine adenoma or adenocarcinoma NOS | 0 |
| BB69. | [M]Sebaceous adenoma and adenocarcinoma | 0 |
| BB691 | [M]Sebaceous adenocarcinoma | 0 |
| BB69z | [M]Sebaceous adenoma or adenocarcinoma NOS | 0 |
| BB6z. | [M]Adnexal and skin appendage neoplasm NOS | 0 |
| BBB5. | [M]Adenocarcinoma with apocrine metaplasia | 0 |
| BBE.. | [M]Naevi and melanomas | 0 |
| BBE1. | [M]Malignant melanoma NOS | 0 |
| BBE10 | [M]Malignant melanoma, regressing | 0 |
| BBE11 | [M]Desmoplastic melanoma, malignant | 0 |
| BBE2. | [M]Nodular melanoma | 0 |
| BBE4. | [M]Balloon cell melanoma | 0 |
| BBEA. | [M]Amelanotic melanoma | 0 |
| BBEC. | [M]Malignant melanoma in junctional naevus | 0 |
| BBEE. | [M]Malignant melanoma in precancerous melanosis | 0 |
| BBEG. | [M]Malignant melanoma in Hutchinson's melanotic freckle | 0 |
| BBEG0 | [M]Acral lentiginous melanoma, malignant | 0 |
| BBEH. | [M]Superficial spreading melanoma | 0 |
| BBEM. | [M]Malignant melanoma in giant pigmented naevus | 0 |
| BBEN. | [M]Epithelioid and spindle cell naevus | 0 |
| BBEP. | [M]Epithelioid cell melanoma | 0 |
| BBEQ. | [M]Spindle cell melanoma NOS | 0 |
| BBER. | [M]Spindle cell melanoma, type A | 0 |
| BBES. | [M]Spindle cell melanoma, type B | 0 |
| BBET. | [M]Mixed epithelioid and spindle melanoma | 0 |
| BBEz. | [M]Naevi or melanoma NOS | 0 |
| BBGM. | [M]Dermatofibrosarcoma NOS | 0 |
| BBGP. | [M]Pigmented dermatofibrosarcoma protuberans | 0 |
| BBmD. | [M] Cutaneous lymphoma | 0 |
| Byu4. | [X]Melanoma and other malignant neoplasms of skin | 0 |
| Byu40 | [X]Malignant melanoma of other and unspecified parts of face | 0 |
| Byu41 | [X]Malignant melanoma of skin, unspecified | 0 |
| Byu42 | [X]other malignant neoplasm of skin of other and unspecified parts of face | 0 |
| Byu43 | [X]Malignant neoplasm of skin, unspecified | 0 |
| Byu5A | [X]Malignant neoplasm overlapping lesion of skin | 0 |
| ByuDD | [X]Other and unspecified peripheral and cutaneous T-cell lymphomas | 0 |
| ZV10y | [V]Personal history of other specified malignant neoplasm | 0 |

---

#### CONCEPT: Stroke¶

| READ\_CODE | DESCRIPTION\_SAIL | SENSITIVE\_CODE |
| --- | --- | --- |
| 14A7. | H/O: CVA/stroke | 0 |
| 14AB. | H/O: TIA | 0 |
| 14AK. | H/O: Stroke in last year | 0 |
| 1M4.. | Central post-stroke pain | 0 |
| 662M. | Stroke monitoring | 0 |
| 662M1 | Stroke 6 month review | 0 |
| 662e. | Stroke/CVA annual review | 0 |
| 662o. | Haemorrhagic stroke monitoring | 0 |
| 70041 | Evacuation of haematoma from temporal lobe of brain | 0 |
| 70042 | Evacuation of haematoma from cerebellum | 0 |
| 70043 | Evacuation of intracerebral haematoma NEC | 0 |
| 70082 | Aspiration of haematoma of brain tissue | 0 |
| 7P242 | Delivery of rehabilitation for stroke | 0 |
| 8HBJ. | Stroke / transient ischaemic attack referral | 0 |
| 8Hd6. | Admission to stroke unit | 0 |
| 9N0p. | Seen in stroke clinic | 0 |
| Fyu55 | [X]Other transient cerebral ischaemic attacks and related syndromes | 0 |
| G6... | Cerebrovascular disease | 0 |
| G60.. | Subarachnoid haemorrhage | 0 |
| G600. | Ruptured berry aneurysm | 0 |
| G601. | Subarachnoid haemorrhage from carotid siphon and bifurcation | 0 |
| G602. | Subarachnoid haemorrhage from middle cerebral artery | 0 |
| G603. | Subarachnoid haemorrhage from anterior communicating artery | 0 |
| G604. | Subarachnoid haemorrhage from posterior communicating artery | 0 |
| G605. | Subarachnoid haemorrhage from basilar artery | 0 |
| G606. | Subarachnoid haemorrhage from vertebral artery | 0 |
| G60X. | Subarachnoid haemorrhage from intracranial artery, unspecified | 0 |
| G60z. | Subarachnoid haemorrhage NOS | 0 |
| G61.. | Intracerebral haemorrhage | 0 |
| G610. | Cortical haemorrhage | 0 |
| G611. | Internal capsule haemorrhage | 0 |
| G612. | Basal nucleus haemorrhage | 0 |
| G613. | Cerebellar haemorrhage | 0 |
| G614. | Pontine haemorrhage | 0 |
| G615. | Bulbar haemorrhage | 0 |
| G616. | External capsule haemorrhage | 0 |
| G617. | Intracerebral haemorrhage, intraventricular | 0 |
| G618. | Intracerebral haemorrhage, multiple localized | 0 |
| G619. | Lobar cerebral haemorrhage | 0 |
| G61X. | Intracerebral haemorrhage in hemisphere, unspecified | 0 |
| G61X0 | Left sided intracerebral haemorrhage, unspecified | 0 |
| G61X1 | Right sided intracerebral haemorrhage, unspecified | 0 |
| G61z. | Intracerebral haemorrhage NOS | 0 |
| G62.. | Other and unspecified intracranial haemorrhage | 0 |
| G620. | Extradural haemorrhage - nontraumatic | 0 |
| G621. | Subdural haemorrhage - nontraumatic | 0 |
| G622. | Subdural haematoma - nontraumatic | 0 |
| G623. | Subdural haemorrhage NOS | 0 |
| G62z. | Intracranial haemorrhage NOS | 0 |
| G63y0 | Cerebral infarct due to thrombosis of precerebral arteries | 0 |
| G63y1 | Cerebral infarction due to embolism of precerebral arteries | 0 |
| G64.. | Cerebral arterial occlusion | 0 |
| G6400 | Cerebral infarction due to thrombosis of cerebral arteries | 0 |
| G6410 | Cerebral infarction due to embolism of cerebral arteries | 0 |
| G64z. | Cerebral infarction NOS | 0 |
| G64z0 | Brainstem infarction | 0 |
| G64z2 | Left sided cerebral infarction | 0 |
| G64z3 | Right sided cerebral infarction | 0 |
| G64z4 | Infarction of basal ganglia | 0 |
| G65.. | Transient cerebral ischaemia | 0 |
| G650. | Basilar artery syndrome | 0 |
| G651. | Vertebral artery syndrome | 0 |
| G6510 | Vertebro-basilar artery syndrome | 0 |
| G652. | Subclavian steal syndrome | 0 |
| G653. | Carotid artery syndrome hemispheric | 0 |
| G654. | Multiple and bilateral precerebral artery syndromes | 0 |
| G656. | Vertebrobasilar insufficiency | 0 |
| G65y. | Other transient cerebral ischaemia | 0 |
| G65z. | Transient cerebral ischaemia NOS | 0 |
| G65z0 | Impending cerebral ischaemia | 0 |
| G65z1 | Intermittent cerebral ischaemia | 0 |
| G65zz | Transient cerebral ischaemia NOS | 0 |
| G66.. | Stroke and cerebrovascular accident unspecified | 0 |
| G663. | Brain stem stroke syndrome | 0 |
| G664. | Cerebellar stroke syndrome | 0 |
| G667. | Left sided CVA | 0 |
| G668. | Right sided CVA | 0 |
| G669. | Cerebral palsy, not congenital or infantile, acute | 0 |
| G6760 | Cerebral infarction due to cerebral venous thrombosis, nonpyogenic | 0 |
| G680. | Sequelae of subarachnoid haemorrhage | 0 |
| G681. | Sequelae of intracerebral haemorrhage | 0 |
| G6W.. | Cerebral infarction due to unspecified occlusion or stenosis of precerebral arteries | 0 |
| G6X.. | Cerebral infarction due to unspecified occlusion or stenosis of cerebral arteries | 0 |
| Gyu60 | [X]Subarachnoid haemorrhage from other intracranial arteries | 0 |
| Gyu61 | [X]Other subarachnoid haemorrhage | 0 |
| Gyu62 | [X]Other intracerebral haemorrhage | 0 |
| Gyu63 | [X]Cerebral infarction due to unspecified occlusion or stenosis of cerebral arteries | 0 |
| Gyu64 | [X]Other cerebral infarction | 0 |
| Gyu65 | [X]Occlusion and stenosis of other precerebral arteries | 0 |
| Gyu66 | [X]Occlusion and stenosis of other cerebral arteries | 0 |
| Gyu6E | [X]Subarachnoid haemorrhage from intracranial artery, unspecified | 0 |
| Gyu6F | [X]Intracerebral haemorrhage in hemisphere, unspecified | 0 |
| Gyu6G | [X]Cerebral infarction due to unspecified occlusion or stenosis of precerebral arteries | 0 |
| Q200. | Subdural and cerebral haemorrhage due to birth trauma | 0 |
| Q2000 | Cerebral haemorrhage unspecified, due to birth trauma | 0 |
| ZV125 | [V]Personal history of circulatory system disease | 0 |
| ZV12D | [V]Personal history of transient ischaemic attack | 0 |

---

#### CONCEPT: Systemic Lupus Erythematosus¶

| READ\_CODE | DESCRIPTION\_SAIL | SENSITIVE\_CODE |
| --- | --- | --- |
| F3710 | Polyneuropathy in disseminated lupus erythematosus | 0 |
| F3961 | Myopathy due to disseminated lupus erythematosus | 0 |
| H57y4 | Lung disease with systemic lupus erythematosus | 0 |
| K01x4 | Nephrotic syndrome in systemic lupus erythematosus | 0 |
| K0B40 | Renal tubulo-interstitial disorder in systemic lupus erythematosus | 0 |
| M154. | Lupus erythematosus | 0 |
| M1540 | Lupus erythematosus chronicus | 0 |
| M1542 | Lupus erythematosus migrans | 0 |
| M1543 | Lupus erythematosus nodularis | 0 |
| M1544 | Lupus erythematosus profundus | 0 |
| M1545 | Lupus erythematosus tumidus | 0 |
| M1546 | Lupus erythematosus unguium mutilans | 0 |
| M1547 | Subacute cutaneous lupus erythematosus | 0 |
| M154z | Lupus erythematosus NOS | 0 |
| Myu78 | [X]Other local lupus erythematosus | 0 |
| N000. | Systemic lupus erythematosus | 0 |
| N0000 | Disseminated lupus erythematosus | 0 |
| N0001 | Libman-Sacks disease | 0 |
| N0002 | Drug-induced systemic lupus erythematosus | 0 |
| N0003 | Systemic lupus erythematosus with organ or system involvement | 0 |
| N0004 | Systemic lupus erythematosus with pericarditis | 0 |
| N0006 | Cerebral lupus | 0 |
| N000z | Systemic lupus erythematosus NOS | 0 |
| Nyu43 | [X]Other forms of systemic lupus erythematosus | 0 |

---

#### CONCEPT: Systemic sclerosis¶

| READ\_CODE | DESCRIPTION\_SAIL | SENSITIVE\_CODE |
| --- | --- | --- |
| F3966 | Myopathy due to scleroderma | 0 |
| H572. | Lung disease with systemic sclerosis | 0 |
| K0H.. | Acute scleroderma renal crisis | 0 |
| K0J0. | Renal involvement in scleroderma | 0 |
| M210. | Circumscribed scleroderma | 0 |
| M2100 | Unspecified circumscribed scleroderma | 0 |
| M2104 | Linear scleroderma | 0 |
| M210z | Circumscribed scleroderma NOS | 0 |
| N001. | Scleroderma | 0 |
| N0010 | Progressive systemic sclerosis | 0 |
| N0011 | CREST syndrome | 0 |
| N0012 | Systemic sclerosis induced by drugs and chemicals | 0 |
| Nyu45 | [X]Other forms of systemic sclerosis | 0 |
| Q4711 | Scleroderma in newborn | 0 |

---

#### CONCEPT: Thalassaemia¶

| READ\_CODE | DESCRIPTION\_SAIL | SENSITIVE\_CODE |
| --- | --- | --- |
| D104. | Thalassaemia | 0 |
| D1040 | Thalassaemia major NEC | 0 |
| D1042 | Thalassaemia with haemoglobin S disease | 0 |
| D1043 | Alpha thalassaemia | 0 |
| D1046 | Beta intermedia thalassaemia | 0 |
| D1047 | Beta major thalassaemia | 0 |
| D1048 | Beta minor thalassaemia | 0 |
| D1049 | Delta-beta thalassaemia | 0 |
| D104z | Thalassaemia NOS | 0 |
| Dyu11 | [X]Other thalassaemias | 0 |

---

#### CONCEPT: TIA (Transient ischaemic attack)¶

| READ\_CODE | DESCRIPTION\_SAIL | SENSITIVE\_CODE |
| --- | --- | --- |
| 14AB. | H/O: TIA | 0 |
| 14AB0 | H/O amaurosis fugax | 0 |
| 8CRB. | Transient ischaemic attack clinical management plan | 0 |
| F4236 | Amaurosis fugax | 0 |
| Fyu55 | [X]Other transient cerebral ischaemic attacks and related syndromes | 0 |
| G65.. | Transient cerebral ischaemia | 0 |
| G657. | Carotid territory transient ischaemic attack | 0 |
| G65y. | Other transient cerebral ischaemia | 0 |
| G65z. | Transient cerebral ischaemia NOS | 0 |
| G65z0 | Impending cerebral ischaemia | 0 |
| G65z1 | Intermittent cerebral ischaemia | 0 |
| G65zz | Transient cerebral ischaemia NOS | 0 |
| R0027 | [D]Drop attack | 0 |
| ZV12D | [V]Personal history of transient ischaemic attack | 0 |

---

#### CONCEPT: Viral Hepatitis¶

| READ\_CODE | DESCRIPTION\_SAIL | SENSITIVE\_CODE |
| --- | --- | --- |
| 141E. | History of hepatitis B | 1 |
| 14i.. | H/O hepatitis C antiviral drug therapy | 1 |
| 21267 | Hepatitis C resolved | 1 |
| 2J12. | Hepatitis C non immune | 1 |
| 43B4. | Hepatitis B surface antig +ve | 1 |
| 43B6. | Hepatitis B non immune | 1 |
| 43B7. | Hepatitis C non-immune | 1 |
| 43X3. | Hepatitis C antibody test positive | 1 |
| 4JQ3. | Hepatitis C virus genotype | 1 |
| 4JQD. | Hepatitis C viral ribonucleic acid polymerase chain reaction positive | 1 |
| 4JQF. | Hepatitis C antigen positive | 1 |
| 65Q7. | Viral hepatitis carrier | 1 |
| 7Q052 | Hepatitis B treatment drugs Band 1 | 1 |
| 7Q053 | Respiratory syncytial virus treatment and Hepatitis C treatment drugs Band 1 | 1 |
| 8BB5. | 12 week virologic response to hepatitis C treatment | 1 |
| 9NgR. | On hepatitis C treatment plan | 1 |
| 9kV.. | Hepatitis C screening positive - enhanced services administration | 1 |
| 9kZ.. | Hepatitis B screening positive - enhanced services administration | 1 |
| A702. | Viral hepatitis B with coma | 1 |
| A7020 | Acute hepatitis B with delta agent (coinfection) with hepatic coma | 1 |
| A703. | Viral (serum) hepatitis B | 1 |
| A7030 | Acute hepatitis B with delta-agent (coinfection) without hepatic coma | 1 |
| A7040 | Viral hepatitis C with coma | 1 |
| A7050 | Viral hepatitis C without mention of hepatic coma | 1 |
| A7051 | Acute delta-(super)infection of hepatitis B carrier | 1 |
| A7054 | Hepatitis non A non B | 1 |
| A707. | Chronic viral hepatitis | 1 |
| A7070 | Chronic viral hepatitis B with delta-agent | 1 |
| A7071 | Chronic viral hepatitis B without delta-agent | 1 |
| A7072 | Chronic viral hepatitis C | 1 |
| A7073 | Chronic viral hepatitis B | 1 |
| A707X | Chronic viral hepatitis, unspecified | 1 |
| A70A. | Hepatitis C genotype 1 | 1 |
| A70B. | Hepatitis C genotype 2 | 1 |
| A70C. | Hepatitis C genotype 3 | 1 |
| A70D. | Hepatitis C genotype 4 | 1 |
| A70E. | Hepatitis C genotype 5 | 1 |
| A70F. | Hepatitis C genotype 6 | 1 |
| A70G. | Acute hepatitis C | 1 |
| A70z0 | Hepatitis C | 1 |
| AE23. | Sequelae of viral hepatitis | 1 |
| AyuB1 | [X]Other chronic viral hepatitis | 1 |
| AyuB2 | [X]Chronic viral hepatitis, unspecified | 1 |
| AyuJ9 | [X]Sequelae of viral hepatitis | 1 |
| Q409. | Congenital viral hepatitis | 1 |
| Q4091 | Congenital hepatitis B infection | 1 |
| Q409y | Other specified congenital viral hepatitis | 1 |
| Q409z | Congenital viral hepatitis NOS | 1 |
| ZV026 | [V]Viral hepatitis carrier | 1 |
| ZV02B | [V]Hepatitis B carrier | 1 |
| ZV02C | [V]Hepatitis C carrier | 1 |

---

#### CONCEPT: VTD (Venous thrombotic disease)¶

| READ\_CODE | DESCRIPTION\_SAIL | SENSITIVE\_CODE |
| --- | --- | --- |
| 14A81 | H/O: Deep Vein Thrombosis | 0 |
| F050. | Embolism of central nervous system venous sinus | 0 |
| F0500 | Embolism cavernous sinus | 0 |
| F0501 | Embolism superior longitudinal sinus | 0 |
| F0502 | Embolism lateral sinus | 0 |
| F0503 | Embolism transverse sinus | 0 |
| F050z | Embolism central nervous system venous sinus NOS | 0 |
| F051. | Thrombosis of central nervous system venous sinuses | 0 |
| F0510 | Thrombosis cavernous sinus | 0 |
| F0511 | Thrombosis of superior longitudinal sinus | 0 |
| F0512 | Thrombosis lateral sinus | 0 |
| F0513 | Thrombosis transverse sinus | 0 |
| F051z | Thrombosis of central nervous system venous sinus NOS | 0 |
| G676. | Nonpyogenic venous sinus thrombosis | 0 |
| G6760 | Cerebral infarction due to cerebral venous thrombosis, nonpyogenic | 0 |
| G67A. | Cerebral vein thrombosis | 0 |
| G800. | Superficial vessel phlebitis and/or thrombophlebitis of the leg | 0 |
| G801. | Deep vein phlebitis and thrombophlebitis of the leg | 0 |
| G801C | Deep vein thrombosis of leg related to air travel | 0 |
| G801D | Deep vein thrombosis of lower limb | 0 |
| G801E | Deep vein thrombosis of leg related to intravenous drug use | 0 |
| G801F | Deep vein thrombosis of peroneal vein | 0 |
| G801G | Recurrent deep vein thrombosis | 0 |
| G801H | Unprovoked deep vein thrombosis | 0 |
| G801J | Provoked deep vein thrombosis | 0 |
| G8020 | Thrombosis of vein of leg | 0 |
| G81.. | Portal vein thrombosis | 0 |
| G82.. | Other venous embolism and thrombosis | 0 |
| G820. | Budd - Chiari syndrome (hepatic vein thrombosis) | 0 |
| G822. | Embolism and thrombosis of the vena cava | 0 |
| G8220 | Thrombosis of inferior vena cava | 0 |
| G823. | Embolism and thrombosis of the renal vein | 0 |
| G824. | Axillary vein thrombosis | 0 |
| G825. | Thrombosis of subclavian vein | 0 |
| G826. | Thrombosis of internal jugular vein | 0 |
| G827. | Thrombosis of external jugular vein | 0 |
| G82z0 | Embolus of vein NOS | 0 |
| G82z1 | Thrombosis of vein NOS | 0 |
| Gyu82 | [X]Embolism and thrombosis of other specified veins | 0 |
| L413. | Antenatal deep vein thrombosis | 0 |
| L4130 | Antenatal deep vein thrombosis unspecified | 0 |
| L4131 | Antenatal deep vein thrombosis - delivered | 0 |
| L4132 | Antenatal deep vein thrombosis with antenatal complication | 0 |
| L413z | Antenatal deep vein thrombosis NOS | 0 |
| L414. | Postnatal deep vein thrombosis | 0 |
| L4140 | Postnatal deep vein thrombosis unspecified | 0 |
| L4141 | Postnatal deep vein thrombosis - delivered with postnatal complication | 0 |
| L4142 | Postnatal deep vein thrombosis with postnatal complication | 0 |
| L414z | Postnatal deep vein thrombosis NOS | 0 |
| L417. | Obstetric cerebral venous thrombosis | 0 |
| L4170 | Cerebral venous thrombosis in pregnancy | 0 |
| L4171 | Cerebral venous thrombosis in the puerperium | 0 |
| SP08a | Thrombosis of vein of transplanted kidney | 0 |
| SP122 | Post operative deep vein thrombosis | 0 |
| ZV128 | [V] Personal history deep vein thrombosis | 0 |

### **ICD10 codes**¶

---

#### CONCEPT: Addison's disease¶

| ICD10\_CODE | DESCRIPTION\_SAIL | SENSITIVE\_CODE |
| --- | --- | --- |
| A187 | Tuberculosis of adrenal glands | 0 |
| E271 | Primary adrenocortical insufficiency | 0 |
| E272 | Addisonian crisis | 0 |
| E351 | Disorders of adrenal glands in diseases classified elsewhere | 0 |
| E896 | Postprocedural adrenocortical(-medullary) hypofunction | 0 |
| G735 | Myopathy in endocrine diseases | 0 |

---

#### CONCEPT: Anaemia¶

| ICD10\_CODE | DESCRIPTION\_SAIL | SENSITIVE\_CODE |
| --- | --- | --- |
| D51 | Vitamin B12 deficiency anaemia | 0 |
| D510 | Vitamin B12 deficiency anaemia due to intrinsic factor deficiency | 0 |
| D511 | Vitamin B12 deficiency anaemia due to selective vitamin B12 malabsorption with proteinuria | 0 |
| D512 | Transcobalamin II deficiency | 0 |
| D513 | Other dietary vitamin B12 deficiency anaemia | 0 |
| D518 | Other vitamin B12 deficiency anaemias | 0 |
| D519 | Vitamin B12 deficiency anaemia, unspecified | 0 |

---

#### CONCEPT: Aneurysm¶

| ICD10\_CODE | DESCRIPTION\_SAIL | SENSITIVE\_CODE |
| --- | --- | --- |
| I71 | Aortic aneurysm and dissection | 0 |
| I710 | Dissection of aorta [any part] | 0 |
| I711 | Thoracic aortic aneurysm, ruptured | 0 |
| I712 | Thoracic aortic aneurysm, without mention of rupture | 0 |
| I713 | Abdominal aortic aneurysm, ruptured | 0 |
| I714 | Abdominal aortic aneurysm, without mention of rupture | 0 |
| I715 | Thoracoabdominal aortic aneurysm, ruptured | 0 |
| I716 | Thoracoabdominal aortic aneurysm, without mention of rupture | 0 |
| I718 | Aortic aneurysm of unspecified site, ruptured | 0 |
| I719 | Aortic aneurysm of unspecified site, without mention of rupture | 0 |

---

#### CONCEPT: Ankylosing Spondylitis¶

| ICD10\_CODE | DESCRIPTION\_SAIL | SENSITIVE\_CODE |
| --- | --- | --- |
| M081 | Juvenile ankylosing spondylitis | 0 |
| M0810 | Juvenile ankylosing spondylitis | 0 |
| M0811 | Juvenile ankylosing spondylitis | 0 |
| M0812 | Juvenile ankylosing spondylitis | 0 |
| M0813 | Juvenile ankylosing spondylitis | 0 |
| M0814 | Juvenile ankylosing spondylitis | 0 |
| M0815 | Juvenile ankylosing spondylitis | 0 |
| M0816 | Juvenile ankylosing spondylitis | 0 |
| M0817 | Juvenile ankylosing spondylitis | 0 |
| M0818 | Juvenile ankylosing spondylitis | 0 |
| M0819 | Juvenile ankylosing spondylitis | 0 |
| M45 | Ankylosing spondylitis | 0 |
| M45X0 | Ankylosing spondylitis | 0 |
| M45X1 | Ankylosing spondylitis | 0 |
| M45X2 | Ankylosing spondylitis | 0 |
| M45X3 | Ankylosing spondylitis | 0 |
| M45X4 | Ankylosing spondylitis | 0 |
| M45X5 | Ankylosing spondylitis | 0 |
| M45X6 | Ankylosing spondylitis | 0 |
| M45X7 | Ankylosing spondylitis | 0 |
| M45X8 | Ankylosing spondylitis | 0 |
| M45X9 | Ankylosing spondylitis | 0 |

---

#### CONCEPT: Anxiety¶

| ICD10\_CODE | DESCRIPTION\_SAIL | SENSITIVE\_CODE |
| --- | --- | --- |
| F064 | Organic anxiety disorder | 0 |
| F40 | Phobic anxiety disorders | 0 |
| F400 | Agoraphobia | 0 |
| F401 | Social phobias | 0 |
| F402 | Specific (isolated) phobias | 0 |
| F408 | Other phobic anxiety disorders | 0 |
| F409 | Phobic anxiety disorder, unspecified | 0 |
| F41 | Other anxiety disorders | 0 |
| F410 | Panic disorder [episodic paroxysmal anxiety] | 0 |
| F411 | Generalized anxiety disorder | 0 |
| F412 | Mixed anxiety and depressive disorder | 0 |
| F413 | Other mixed anxiety disorders | 0 |
| F418 | Other specified anxiety disorders | 0 |
| F419 | Anxiety disorder, unspecified | 0 |
| F606 | Anxious [avoidant] personality disorder | 0 |
| F930 | Separation anxiety disorder of childhood | 0 |
| F931 | Phobic anxiety disorder of childhood | 0 |
| F932 | Social anxiety disorder of childhood | 0 |

---

#### CONCEPT: Arrhythmia¶

| ICD10\_CODE | DESCRIPTION\_SAIL | SENSITIVE\_CODE |
| --- | --- | --- |
| I44 | Atrioventricular and left bundle-branch block | 0 |
| I440 | Atrioventricular block, first degree | 0 |
| I441 | Atrioventricular block, second degree | 0 |
| I442 | Atrioventricular block, complete | 0 |
| I443 | Other and unspecified atrioventricular block | 0 |
| I447 | Left bundle-branch block, unspecified | 0 |
| I451 | Other and unspecified right bundle-branch block | 0 |
| I453 | Trifascicular block | 0 |
| I455 | Other specified heart block | 0 |
| I46 | Cardiac arrest | 0 |
| I460 | Cardiac arrest with successful resuscitation | 0 |
| I461 | Sudden cardiac death, so described | 0 |
| I469 | Cardiac arrest, unspecified | 0 |
| I47 | Paroxysmal tachycardia | 0 |
| I470 | Re-entry ventricular arrhythmia | 0 |
| I471 | Supraventricular tachycardia | 0 |
| I472 | Ventricular tachycardia | 0 |
| I479 | Paroxysmal tachycardia, unspecified | 0 |
| I48 | Atrial fibrillation and flutter | 0 |
| I480 | Paroxysmal atrial fibrillation | 0 |
| I481 | Persistent atrial fibrillation | 0 |
| I482 | Chronic atrial fibrillation | 0 |
| I483 | Typical atrial flutter | 0 |
| I484 | Atypical atrial flutter | 0 |
| I489 | Atrial fibrillation and atrial flutter, unspecified | 0 |
| I49 | Other cardiac arrhythmias | 0 |
| I490 | Ventricular fibrillation and flutter | 0 |
| I491 | Atrial premature depolarization | 0 |
| I492 | Junctional premature depolarization | 0 |
| I493 | Ventricular premature depolarization | 0 |
| I494 | Other and unspecified premature depolarization | 0 |
| I495 | Sick sinus syndrome | 0 |
| I498 | Other specified cardiac arrhythmias | 0 |
| I499 | Cardiac arrhythmia, unspecified | 0 |
| P291 | Neonatal cardiac dysrhythmia | 0 |

---

#### CONCEPT: Asthma¶

| ICD10\_CODE | DESCRIPTION\_SAIL | SENSITIVE\_CODE |
| --- | --- | --- |
| J45 | Asthma | 0 |
| J450 | Predominantly allergic asthma | 0 |
| J451 | Nonallergic asthma | 0 |
| J458 | Mixed asthma | 0 |
| J459 | Asthma, unspecified | 0 |
| J46 | Status asthmaticus | 0 |

---

#### CONCEPT: Atopic Eczema¶

| ICD10\_CODE | DESCRIPTION\_SAIL | SENSITIVE\_CODE |
| --- | --- | --- |
| L20 | Atopic dermatitis | 0 |
| L200 | Besnier prurigo | 0 |
| L208 | Other atopic dermatitis | 0 |
| L209 | Atopic dermatitis, unspecified | 0 |

---

#### CONCEPT: Autism and ADHD¶

| ICD10\_CODE | DESCRIPTION\_SAIL | SENSITIVE\_CODE |
| --- | --- | --- |
| F840 | Childhood autism | 0 |
| F841 | Atypical autism | 0 |
| F845 | Asperger syndrome | 0 |
| F900 | Disturbance of activity and attention | 0 |

---

#### CONCEPT: Bipolar Disorder¶

| ICD10\_CODE | DESCRIPTION\_SAIL | SENSITIVE\_CODE |
| --- | --- | --- |
| F30 | Manic episode | 0 |
| F301 | Mania without psychotic symptoms | 0 |
| F302 | Mania with psychotic symptoms | 0 |
| F308 | Other manic episodes | 0 |
| F309 | Manic episode, unspecified | 0 |
| F31 | Bipolar affective disorder | 0 |
| F310 | Bipolar affective disorder, current episode hypomanic | 0 |
| F311 | Bipolar affective disorder, current episode manic without psychotic symptoms | 0 |
| F312 | Bipolar affective disorder, current episode manic with psychotic symptoms | 0 |
| F313 | Bipolar affective disorder, current episode mild or moderate depression | 0 |
| F314 | Bipolar affective disorder, current episode severe depression without psychotic symptoms | 0 |
| F315 | Bipolar affective disorder, current episode severe depression with psychotic symptoms | 0 |
| F316 | Bipolar affective disorder, current episode mixed | 0 |
| F317 | Bipolar affective disorder, currently in remission | 0 |
| F318 | Other bipolar affective disorders | 0 |
| F319 | Bipolar affective disorder, unspecified | 0 |
| F340 | Cyclothymia | 0 |

---

#### CONCEPT: Blindness and low vision¶

| ICD10\_CODE | DESCRIPTION\_SAIL | SENSITIVE\_CODE |
| --- | --- | --- |
| H540 | Blindness, binocular | 0 |
| H541 | Severe visual impairment, binocular | 0 |

---

#### CONCEPT: Breast Cancer¶

| ICD10\_CODE | DESCRIPTION\_SAIL | SENSITIVE\_CODE |
| --- | --- | --- |
| C50 | Malignant neoplasm of breast | 0 |
| C501 | Malignant neoplasm: Central portion of breast | 0 |
| C502 | Malignant neoplasm: Upper-inner quadrant of breast | 0 |
| C503 | Malignant neoplasm: Lower-inner quadrant of breast | 0 |
| C504 | Malignant neoplasm: Upper-outer quadrant of breast | 0 |
| C505 | Malignant neoplasm: Lower-outer quadrant of breast | 0 |
| C506 | Malignant neoplasm: Axillary tail of breast | 0 |
| C508 | Malignant neoplasm: Overlapping lesion of breast | 0 |
| C509 | Malignant neoplasm: Breast, unspecified | 0 |
| D05 | Carcinoma in situ of breast | 0 |
| D057 | Other carcinoma in situ of breast | 0 |
| D059 | Carcinoma in situ of breast, unspecified | 0 |
| D24 | Benign neoplasm of breast | 0 |
| Z853 | Personal history of malignant neoplasm of breast | 0 |

---

#### CONCEPT: Bronchiectasis¶

| ICD10\_CODE | DESCRIPTION\_SAIL | SENSITIVE\_CODE |
| --- | --- | --- |
| J47 | Bronchiectasis | 0 |
| Q334 | Congenital bronchiectasis | 0 |

---

#### CONCEPT: Chronic Liver Disease¶

| ICD10\_CODE | DESCRIPTION\_SAIL | SENSITIVE\_CODE |
| --- | --- | --- |
| B16 | Acute hepatitis B | 0 |
| B160 | Acute hepatitis B with delta-agent (coinfection) with hepatic coma | 0 |
| B161 | Acute hepatitis B with delta-agent (coinfection) without hepatic coma | 0 |
| B162 | Acute hepatitis B without delta-agent with hepatic coma | 0 |
| B169 | Acute hepatitis B without delta-agent and without hepatic coma | 0 |
| B170 | Acute delta-(super)infection of hepatitis B carrier | 0 |
| B171 | Acute hepatitis C | 1 |
| B18 | Chronic viral hepatitis | 0 |
| B180 | Chronic viral hepatitis B with delta-agent | 0 |
| B181 | Chronic viral hepatitis B without delta-agent | 0 |
| B182 | Chronic viral hepatitis C | 0 |
| B188 | Other chronic viral hepatitis | 0 |
| B189 | Chronic viral hepatitis, unspecified | 0 |
| I85 | Oesophageal varices | 0 |
| I850 | Oesophageal varices with bleeding | 0 |
| I859 | Oesophageal varices without bleeding | 0 |
| I982 | Oesophageal varices without bleeding in diseases classified elsewhere | 0 |
| I983 | Oesophageal varices with bleeding in diseases classified elsewhere | 0 |
| K70 | Alcoholic liver disease | 0 |
| K700 | Alcoholic fatty liver | 0 |
| K701 | Alcoholic hepatitis | 0 |
| K702 | Alcoholic fibrosis and sclerosis of liver | 0 |
| K703 | Alcoholic cirrhosis of liver | 0 |
| K704 | Alcoholic hepatic failure | 0 |
| K709 | Alcoholic liver disease, unspecified | 0 |
| K713 | Toxic liver disease with chronic persistent hepatitis | 0 |
| K714 | Toxic liver disease with chronic lobular hepatitis | 0 |
| K715 | Toxic liver disease with chronic active hepatitis | 0 |
| K717 | Toxic liver disease with fibrosis and cirrhosis of liver | 0 |
| K72 | Hepatic failure, not elsewhere classified | 0 |
| K721 | Chronic hepatic failure | 0 |
| K729 | Hepatic failure, unspecified | 0 |
| K73 | Chronic hepatitis, not elsewhere classified | 0 |
| K730 | Chronic persistent hepatitis, not elsewhere classified | 0 |
| K731 | Chronic lobular hepatitis, not elsewhere classified | 0 |
| K732 | Chronic active hepatitis, not elsewhere classified | 0 |
| K738 | Other chronic hepatitis, not elsewhere classified | 0 |
| K739 | Chronic hepatitis, unspecified | 0 |
| K74 | Fibrosis and cirrhosis of liver | 0 |
| K740 | Hepatic fibrosis | 0 |
| K742 | Hepatic fibrosis with hepatic sclerosis | 0 |
| K743 | Primary biliary cirrhosis | 0 |
| K744 | Secondary biliary cirrhosis | 0 |
| K745 | Biliary cirrhosis, unspecified | 0 |
| K746 | Other and unspecified cirrhosis of liver | 0 |
| K754 | Autoimmune hepatitis | 0 |
| K766 | Portal hypertension | 0 |
| K767 | Hepatorenal syndrome | 0 |

---

#### CONCEPT: Chronic Pain¶

| ICD10\_CODE | DESCRIPTION\_SAIL | SENSITIVE\_CODE |
| --- | --- | --- |
| R521 | Chronic intractable pain | 0 |
| R522 | Other chronic pain | 0 |

---

#### CONCEPT: Chronic Kidney Disease (CKD) Stage 3\_5¶

| ICD10\_CODE | DESCRIPTION\_SAIL | SENSITIVE\_CODE |
| --- | --- | --- |
| N183 | Chronic kidney disease, stage 3 | 0 |
| N184 | Chronic kidney disease, stage 4 | 0 |
| N185 | Chronic kidney disease, stage 5 | 0 |

---

#### CONCEPT: Coeliac Disease¶

| ICD10\_CODE | DESCRIPTION\_SAIL | SENSITIVE\_CODE |
| --- | --- | --- |
| K900 | Coeliac disease | 0 |

---

#### CONCEPT: Colon Cancer¶

| ICD10\_CODE | DESCRIPTION\_SAIL | SENSITIVE\_CODE |
| --- | --- | --- |
| C18 | Malignant neoplasm of colon | 0 |
| C180 | Malignant neoplasm: Caecum | 0 |
| C181 | Malignant neoplasm: Appendix | 0 |
| C182 | Malignant neoplasm: Ascending colon | 0 |
| C183 | Malignant neoplasm: Hepatic flexure | 0 |
| C184 | Malignant neoplasm: Transverse colon | 0 |
| C185 | Malignant neoplasm: Splenic flexure | 0 |
| C186 | Malignant neoplasm: Descending colon | 0 |
| C187 | Malignant neoplasm: Sigmoid colon | 0 |
| C188 | Malignant neoplasm: Overlapping lesion of colon | 0 |
| C189 | Malignant neoplasm: Colon, unspecified | 0 |
| C19 | Malignant neoplasm of rectosigmoid junction | 0 |
| C20 | Malignant neoplasm of rectum | 0 |
| C218 | Malignant neoplasm: Overlapping lesion of rectum, anus and anal canal | 0 |
| C785 | Secondary malignant neoplasm of large intestine and rectum | 0 |
| D010 | Carcinoma in situ: Colon | 0 |
| D011 | Carcinoma in situ: Rectosigmoid junction | 0 |
| D012 | Carcinoma in situ: Rectum | 0 |
| D013 | Carcinoma in situ: Anus and anal canal | 0 |
| D017 | Carcinoma in situ: Other specified digestive organs | 0 |
| D37 | Neoplasm of uncertain or unknown behaviour of oral cavity and digestive organs | 0 |
| D374 | Neoplasm of uncertain or unknown behaviour: Colon | 0 |
| D375 | Neoplasm of uncertain or unknown behaviour: Rectum | 0 |

---

#### CONCEPT: Congenital disease and chromosomal abnormalities¶

| ICD10\_CODE | DESCRIPTION\_SAIL | SENSITIVE\_CODE |
| --- | --- | --- |
| I070 | Tricuspid stenosis | 0 |
| I072 | Tricuspid stenosis with insufficiency | 0 |
| P70 | Transitory disorders of carbohydrate metabolism specific to fetus and newborn | 0 |
| P700 | Syndrome of infant of mother with gestational diabetes | 0 |
| P701 | Syndrome of infant of a diabetic mother | 0 |
| P702 | Neonatal diabetes mellitus | 0 |
| P703 | Iatrogenic neonatal hypoglycaemia | 0 |
| P704 | Other neonatal hypoglycaemia | 0 |
| P708 | Other transitory disorders of carbohydrate metabolism of fetus and newborn | 0 |
| P709 | Transitory disorder of carbohydrate metabolism of fetus and newborn, unspecified | 0 |
| P71 | Transitory neonatal disorders of calcium and magnesium metabolism | 0 |
| P72 | Other transitory neonatal endocrine disorders | 0 |
| P720 | Neonatal goitre, not elsewhere classified | 0 |
| P721 | Transitory neonatal hyperthyroidism | 0 |
| P722 | Other transitory neonatal disorders of thyroid function, not elsewhere classified | 0 |
| P728 | Other specified transitory neonatal endocrine disorders | 0 |
| P729 | Transitory neonatal endocrine disorder, unspecified | 0 |
| Q20 | Congenital malformations of cardiac chambers and connections | 0 |
| Q200 | Common arterial trunk | 0 |
| Q201 | Double outlet right ventricle | 0 |
| Q202 | Double outlet left ventricle | 0 |
| Q203 | Discordant ventriculoarterial connection | 0 |
| Q204 | Double inlet ventricle | 0 |
| Q205 | Discordant atrioventricular connection | 0 |
| Q206 | Isomerism of atrial appendages | 0 |
| Q208 | Other congenital malformations of cardiac chambers and connections | 0 |
| Q209 | Congenital malformation of cardiac chambers and connections, unspecified | 0 |
| Q21 | Congenital malformations of cardiac septa | 0 |
| Q210 | Ventricular septal defect | 0 |
| Q211 | Atrial septal defect | 1 |
| Q212 | Atrioventricular septal defect | 0 |
| Q213 | Tetralogy of Fallot | 0 |
| Q214 | Aortopulmonary septal defect | 0 |
| Q218 | Other congenital malformations of cardiac septa | 1 |
| Q219 | Congenital malformation of cardiac septum, unspecified | 1 |
| Q22 | Congenital malformations of pulmonary and tricuspid valves | 0 |
| Q220 | Pulmonary valve atresia | 0 |
| Q221 | Congenital pulmonary valve stenosis | 0 |
| Q222 | Congenital pulmonary valve insufficiency | 0 |
| Q223 | Other congenital malformations of pulmonary valve | 0 |
| Q224 | Congenital tricuspid stenosis | 0 |
| Q225 | Ebstein anomaly | 0 |
| Q226 | Hypoplastic right heart syndrome | 0 |
| Q228 | Other congenital malformations of tricuspid valve | 0 |
| Q229 | Congenital malformation of tricuspid valve, unspecified | 0 |
| Q23 | Congenital malformations of aortic and mitral valves | 0 |
| Q230 | Congenital stenosis of aortic valve | 0 |
| Q231 | Congenital insufficiency of aortic valve | 0 |
| Q232 | Congenital mitral stenosis | 0 |
| Q233 | Congenital mitral insufficiency | 0 |
| Q234 | Hypoplastic left heart syndrome | 0 |
| Q238 | Other congenital malformations of aortic and mitral valves | 0 |
| Q239 | Congenital malformation of aortic and mitral valves, unspecified | 0 |
| Q24 | Other congenital malformations of heart | 0 |
| Q240 | Dextrocardia | 0 |
| Q241 | Laevocardia | 0 |
| Q242 | Cor triatriatum | 0 |
| Q243 | Pulmonary infundibular stenosis | 0 |
| Q244 | Congenital subaortic stenosis | 0 |
| Q245 | Malformation of coronary vessels | 0 |
| Q246 | Congenital heart block | 0 |
| Q248 | Other specified congenital malformations of heart | 0 |
| Q249 | Congenital malformation of heart, unspecified | 0 |
| Q25 | Congenital malformations of great arteries | 0 |
| Q250 | Patent ductus arteriosus | 0 |
| Q251 | Coarctation of aorta | 0 |
| Q252 | Atresia of aorta | 0 |
| Q253 | Stenosis of aorta | 0 |
| Q254 | Other congenital malformations of aorta | 0 |
| Q255 | Atresia of pulmonary artery | 0 |
| Q256 | Stenosis of pulmonary artery | 0 |
| Q257 | Other congenital malformations of pulmonary artery | 0 |
| Q258 | Other congenital malformations of great arteries | 0 |
| Q259 | Congenital malformation of great arteries, unspecified | 0 |
| Q26 | Congenital malformations of great veins | 0 |
| Q260 | Congenital stenosis of vena cava | 0 |
| Q261 | Persistent left superior vena cava | 0 |
| Q262 | Total anomalous pulmonary venous connection | 0 |
| Q263 | Partial anomalous pulmonary venous connection | 0 |
| Q264 | Anomalous pulmonary venous connection, unspecified | 0 |
| Q268 | Other congenital malformations of great veins | 0 |

---

#### CONCEPT: COPD (Chronic obstructive pulmonary disease)¶

| ICD10\_CODE | DESCRIPTION\_SAIL | SENSITIVE\_CODE |
| --- | --- | --- |
| J41 | Simple and mucopurulent chronic bronchitis | 0 |
| J410 | Simple chronic bronchitis | 0 |
| J411 | Mucopurulent chronic bronchitis | 0 |
| J418 | Mixed simple and mucopurulent chronic bronchitis | 0 |
| J42 | Unspecified chronic bronchitis | 0 |
| J43 | Emphysema | 0 |
| J430 | MacLeod syndrome | 0 |
| J431 | Panlobular emphysema | 0 |
| J432 | Centrilobular emphysema | 0 |
| J438 | Other emphysema | 0 |
| J439 | Emphysema, unspecified | 0 |
| J44 | Other chronic obstructive pulmonary disease | 0 |
| J440 | Chronic obstructive pulmonary disease with acute lower respiratory infection | 0 |
| J441 | Chronic obstructive pulmonary disease with acute exacerbation, unspecified | 0 |
| J448 | Other specified chronic obstructive pulmonary disease | 0 |
| J449 | Chronic obstructive pulmonary disease, unspecified | 0 |
| J982 | Interstitial emphysema | 0 |

---

#### CONCEPT: Coronary heart disease¶

| ICD10\_CODE | DESCRIPTION\_SAIL | SENSITIVE\_CODE |
| --- | --- | --- |
| I20 | Angina pectoris | 0 |
| I200 | Unstable angina | 0 |
| I201 | Angina pectoris with documented spasm | 0 |
| I208 | Other forms of angina pectoris | 0 |
| I209 | Angina pectoris, unspecified | 0 |
| I24 | Other acute ischaemic heart diseases | 0 |
| I240 | Coronary thrombosis not resulting in myocardial infarction | 0 |
| I248 | Other forms of acute ischaemic heart disease | 0 |
| I249 | Acute ischaemic heart disease, unspecified | 0 |
| I25 | Chronic ischaemic heart disease | 0 |
| I250 | Atherosclerotic cardiovascular disease, so described | 0 |
| I251 | Atherosclerotic heart disease | 0 |
| I253 | Aneurysm of heart | 0 |
| I254 | Coronary artery aneurysm and dissection | 0 |
| I255 | Ischaemic cardiomyopathy | 0 |
| I256 | Silent myocardial ischaemia | 0 |
| I258 | Other forms of chronic ischaemic heart disease | 0 |
| I259 | Chronic ischaemic heart disease, unspecified | 0 |
| Z500 | Cardiac rehabilitation | 0 |
| Z955 | Presence of coronary angioplasty implant and graft | 0 |

---

#### CONCEPT: Cystic fibrosis¶

| ICD10\_CODE | DESCRIPTION\_SAIL | SENSITIVE\_CODE |
| --- | --- | --- |
| E84 | Cystic fibrosis | 0 |
| E840 | Cystic fibrosis with pulmonary manifestations | 0 |
| E841 | Cystic fibrosis with intestinal manifestations | 0 |
| E848 | Cystic fibrosis with other manifestations | 0 |
| E849 | Cystic fibrosis, unspecified | 0 |
| P75 | Meconium ileus in cystic fibrosis | 0 |

---

#### CONCEPT: Dementia alzheimer¶

| ICD10\_CODE | DESCRIPTION\_SAIL | SENSITIVE\_CODE |
| --- | --- | --- |
| A810 | Creutzfeldt-Jakob disease | 0 |
| F00 | Dementia in Alzheimer disease | 0 |
| F000 | Dementia in Alzheimer disease with early onset | 0 |
| F001 | Dementia in Alzheimer disease with late onset | 0 |
| F002 | Dementia in Alzheimer disease, atypical or mixed type | 0 |
| F009 | Dementia in Alzheimer disease, unspecified | 0 |
| F02 | Dementia in other diseases classified elsewhere | 0 |
| F020 | Dementia in Pick disease | 0 |
| F021 | Dementia in Creutzfeldt-Jakob disease | 0 |
| F022 | Dementia in Huntington disease | 0 |
| F023 | Dementia in Parkinson disease | 0 |
| F024 | Dementia in human immunodeficiency virus [HIV] disease | 1 |
| F028 | Dementia in other specified diseases classified elsewhere | 0 |
| F03 | Unspecified dementia | 0 |
| F051 | Delirium superimposed on dementia | 0 |
| G30 | Alzheimer disease | 0 |
| G300 | Alzheimer disease with early onset | 0 |
| G301 | Alzheimer disease with late onset | 0 |
| G308 | Other Alzheimer disease | 0 |
| G309 | Alzheimer disease, unspecified | 0 |
| G912 | Normal-pressure hydrocephalus | 0 |

---

#### CONCEPT: Depression¶

| ICD10\_CODE | DESCRIPTION\_SAIL | SENSITIVE\_CODE |
| --- | --- | --- |
| F204 | Post-schizophrenic depression | 0 |
| F251 | Schizoaffective disorder, depressive type | 0 |
| F32 | Depressive episode | 0 |
| F320 | Mild depressive episode | 0 |
| F321 | Moderate depressive episode | 0 |
| F322 | Severe depressive episode without psychotic symptoms | 0 |
| F323 | Severe depressive episode with psychotic symptoms | 0 |
| F328 | Other depressive episodes | 0 |
| F329 | Depressive episode, unspecified | 0 |
| F33 | Recurrent depressive disorder | 0 |
| F330 | Recurrent depressive disorder, current episode mild | 0 |
| F331 | Recurrent depressive disorder, current episode moderate | 0 |
| F332 | Recurrent depressive disorder, current episode severe without psychotic symptoms | 0 |
| F333 | Recurrent depressive disorder, current episode severe with psychotic symptoms | 0 |
| F338 | Other recurrent depressive disorders | 0 |
| F339 | Recurrent depressive disorder, unspecified | 0 |
| F341 | Dysthymia | 0 |
| F412 | Mixed anxiety and depressive disorder | 0 |
| F920 | Depressive conduct disorder | 0 |

---

#### CONCEPT: Diabetes Type 1¶

| ICD10\_CODE | DESCRIPTION\_SAIL | SENSITIVE\_CODE |
| --- | --- | --- |
| E10 | Type 1 diabetes mellitus | 0 |
| E100 | Type 1 diabetes mellitus | 0 |
| E101 | Type 1 diabetes mellitus | 0 |
| E102 | Type 1 diabetes mellitus | 0 |
| E103 | Type 1 diabetes mellitus | 0 |
| E104 | Type 1 diabetes mellitus | 0 |
| E105 | Type 1 diabetes mellitus | 0 |
| E106 | Type 1 diabetes mellitus | 0 |
| E107 | Type 1 diabetes mellitus | 0 |
| E108 | Type 1 diabetes mellitus | 0 |
| E109 | Type 1 diabetes mellitus | 0 |
| O240 | Diabetes mellitus in pregnancy: Pre-existing type 1 diabetes mellitus | 0 |

---

#### CONCEPT: Diabetes Type 2¶

| ICD10\_CODE | DESCRIPTION\_SAIL | SENSITIVE\_CODE |
| --- | --- | --- |
| E11 | Type 2 diabetes mellitus | 0 |
| E110 | Type 2 diabetes mellitus | 0 |
| E111 | Type 2 diabetes mellitus | 0 |
| E112 | Type 2 diabetes mellitus | 0 |
| E113 | Type 2 diabetes mellitus | 0 |
| E114 | Type 2 diabetes mellitus | 0 |
| E115 | Type 2 diabetes mellitus | 0 |
| E116 | Type 2 diabetes mellitus | 0 |
| E117 | Type 2 diabetes mellitus | 0 |
| E118 | Type 2 diabetes mellitus | 0 |
| E119 | Type 2 diabetes mellitus | 0 |
| O241 | Diabetes mellitus in pregnancy: Pre-existing type 2 diabetes mellitus | 0 |

---

#### CONCEPT: Diabetic retinopathy¶

| ICD10\_CODE | DESCRIPTION\_SAIL | SENSITIVE\_CODE |
| --- | --- | --- |
| E103 | Type 1 diabetes mellitus | 0 |
| E113 | Type 2 diabetes mellitus | 0 |
| E133 | Other specified diabetes mellitus | 0 |
| E143 | Unspecified diabetes mellitus | 0 |
| H360 | Diabetic retinopathy | 0 |

---

#### CONCEPT: Dialysis¶

| ICD10\_CODE | DESCRIPTION\_SAIL | SENSITIVE\_CODE |
| --- | --- | --- |
| Y602 | During kidney dialysis or other perfusion | 0 |
| Y612 | During kidney dialysis or other perfusion | 0 |
| Y622 | During kidney dialysis or other perfusion | 0 |
| Y841 | Kidney dialysis | 0 |
| Z49 | Care involving dialysis | 0 |
| Z490 | Preparatory care for dialysis | 0 |
| Z491 | Extracorporeal dialysis | 0 |
| Z492 | Other dialysis | 0 |
| Z992 | Dependence on renal dialysis | 0 |

---

#### CONCEPT: Diverticular Disease¶

| ICD10\_CODE | DESCRIPTION\_SAIL | SENSITIVE\_CODE |
| --- | --- | --- |
| K57 | Diverticular disease of intestine | 0 |
| K570 | Diverticular disease of small intestine with perforation and abscess | 0 |
| K571 | Diverticular disease of small intestine without perforation or abscess | 0 |
| K572 | Diverticular disease of large intestine with perforation and abscess | 0 |
| K573 | Diverticular disease of large intestine without perforation or abscess | 0 |
| K574 | Diverticular disease of both small and large intestine with perforation and abscess | 0 |
| K575 | Diverticular disease of both small and large intestine without perforation or abscess | 0 |
| K578 | Diverticular disease of intestine, part unspecified, with perforation and abscess | 0 |
| K579 | Diverticular disease of intestine, part unspecified, without perforation or abscess | 0 |

---

#### CONCEPT: Drug or alcohol misuse¶

| ICD10\_CODE | DESCRIPTION\_SAIL | SENSITIVE\_CODE |
| --- | --- | --- |
| E244 | Alcohol-induced pseudo-Cushing syndrome | 0 |
| F04 | Organic amnesic syndrome, not induced by alcohol and other psychoactive substances | 0 |
| F05 | Delirium, not induced by alcohol and other psychoactive substances | 0 |
| F10 | Mental and behavioural disorders due to use of alcohol | 0 |
| F100 | Mental and behavioural disorders due to use of alcohol | 0 |
| F101 | Mental and behavioural disorders due to use of alcohol | 0 |
| F102 | Mental and behavioural disorders due to use of alcohol | 0 |
| F103 | Mental and behavioural disorders due to use of alcohol | 0 |
| F104 | Mental and behavioural disorders due to use of alcohol | 0 |
| F105 | Mental and behavioural disorders due to use of alcohol | 0 |
| F106 | Mental and behavioural disorders due to use of alcohol | 0 |
| F107 | Mental and behavioural disorders due to use of alcohol | 0 |
| F108 | Mental and behavioural disorders due to use of alcohol | 0 |
| F109 | Mental and behavioural disorders due to use of alcohol | 0 |
| F19 | Mental and behavioural disorders due to multiple drug use and use of other psychoactive substances | 0 |
| F190 | Mental and behavioural disorders due to multiple drug use and use of other psychoactive substances | 0 |
| F191 | Mental and behavioural disorders due to multiple drug use and use of other psychoactive substances | 0 |
| F192 | Mental and behavioural disorders due to multiple drug use and use of other psychoactive substances | 0 |
| F193 | Mental and behavioural disorders due to multiple drug use and use of other psychoactive substances | 0 |
| F194 | Mental and behavioural disorders due to multiple drug use and use of other psychoactive substances | 0 |
| F195 | Mental and behavioural disorders due to multiple drug use and use of other psychoactive substances | 0 |
| F196 | Mental and behavioural disorders due to multiple drug use and use of other psychoactive substances | 0 |
| F197 | Mental and behavioural disorders due to multiple drug use and use of other psychoactive substances | 0 |
| F198 | Mental and behavioural disorders due to multiple drug use and use of other psychoactive substances | 0 |
| F199 | Mental and behavioural disorders due to multiple drug use and use of other psychoactive substances | 0 |
| F55 | Abuse of non-dependence-producing substances | 0 |
| G312 | Degeneration of nervous system due to alcohol | 0 |
| G621 | Alcoholic polyneuropathy | 0 |
| G721 | Alcoholic myopathy | 0 |
| I426 | Alcoholic cardiomyopathy | 0 |
| K292 | Alcoholic gastritis | 0 |
| K70 | Alcoholic liver disease | 0 |
| K700 | Alcoholic fatty liver | 0 |
| K701 | Alcoholic hepatitis | 0 |
| K702 | Alcoholic fibrosis and sclerosis of liver | 0 |
| K703 | Alcoholic cirrhosis of liver | 0 |
| K704 | Alcoholic hepatic failure | 0 |
| K709 | Alcoholic liver disease, unspecified | 0 |
| K852 | Alcohol-induced acute pancreatitis | 0 |
| K860 | Alcohol-induced chronic pancreatitis | 0 |
| T436 | Poisoning: Psychostimulants with abuse potential | 0 |
| Y497 | Psychostimulants with abuse potential | 0 |
| Z722 | Drug use | 0 |
| Z864 | Personal history of psychoactive substance abuse | 0 |

---

#### CONCEPT: Eating disorders¶

| ICD10\_CODE | DESCRIPTION\_SAIL | SENSITIVE\_CODE |
| --- | --- | --- |
| F50 | Eating disorders | 0 |
| F500 | Anorexia nervosa | 0 |
| F501 | Atypical anorexia nervosa | 0 |
| F502 | Bulimia nervosa | 0 |
| F503 | Atypical bulimia nervosa | 0 |
| F504 | Overeating associated with other psychological disturbances | 0 |
| F505 | Vomiting associated with other psychological disturbances | 0 |
| F508 | Other eating disorders | 0 |
| F509 | Eating disorder, unspecified | 0 |
| F983 | Pica of infancy and childhood | 0 |

---

#### CONCEPT: Epilepsy¶

| ICD10\_CODE | DESCRIPTION\_SAIL | SENSITIVE\_CODE |
| --- | --- | --- |
| F803 | Acquired aphasia with epilepsy [Landau-Kleffner] | 0 |
| G40 | Epilepsy | 0 |
| G400 | Localization-related (focal)(partial) idiopathic epilepsy and epileptic syndromes with seizures of localized onset | 0 |
| G401 | Localization-related (focal)(partial) symptomatic epilepsy and epileptic syndromes with simple partial seizures | 0 |
| G402 | Localization-related (focal)(partial) symptomatic epilepsy and epileptic syndromes with complex partial seizures | 0 |
| G403 | Generalized idiopathic epilepsy and epileptic syndromes | 0 |
| G404 | Other generalized epilepsy and epileptic syndromes | 0 |
| G405 | Special epileptic syndromes | 0 |
| G408 | Other epilepsy | 0 |
| G409 | Epilepsy, unspecified | 0 |
| G41 | Status epilepticus | 0 |
| G410 | Grand mal status epilepticus | 0 |
| G411 | Petit mal status epilepticus | 0 |
| G412 | Complex partial status epilepticus | 0 |
| G418 | Other status epilepticus | 0 |
| G419 | Status epilepticus, unspecified | 0 |

---

#### CONCEPT: Fibromyalgia¶

| ICD10\_CODE | DESCRIPTION\_SAIL | SENSITIVE\_CODE |
| --- | --- | --- |
| M797 | Fibromyalgia | 0 |
| M7970 | Fibromyalgia | 0 |
| M7971 | Fibromyalgia | 0 |
| M7972 | Fibromyalgia | 0 |
| M7973 | Fibromyalgia | 0 |
| M7974 | Fibromyalgia | 0 |
| M7975 | Fibromyalgia | 0 |
| M7976 | Fibromyalgia | 0 |
| M7977 | Fibromyalgia | 0 |
| M7978 | Fibromyalgia | 0 |
| M7979 | Fibromyalgia | 0 |

---

#### CONCEPT: Glaucoma¶

| ICD10\_CODE | DESCRIPTION\_SAIL | SENSITIVE\_CODE |
| --- | --- | --- |
| H40 | Glaucoma | 0 |
| H401 | Primary open-angle glaucoma | 0 |
| H402 | Primary angle-closure glaucoma | 0 |
| H408 | Other glaucoma | 0 |
| H409 | Glaucoma, unspecified | 0 |
| H42 | Glaucoma in diseases classified elsewhere | 0 |
| H420 | Glaucoma in endocrine, nutritional and metabolic diseases | 0 |
| H428 | Glaucoma in other diseases classified elsewhere | 0 |

---

#### CONCEPT: Gout¶

| ICD10\_CODE | DESCRIPTION\_SAIL | SENSITIVE\_CODE |
| --- | --- | --- |
| M10 | Gout | 0 |
| M100 | Idiopathic gout | 0 |
| M1000 | Idiopathic gout | 0 |
| M1001 | Idiopathic gout | 0 |
| M1002 | Idiopathic gout | 0 |
| M1003 | Idiopathic gout | 0 |
| M1004 | Idiopathic gout | 0 |
| M1005 | Idiopathic gout | 0 |
| M1006 | Idiopathic gout | 0 |
| M1007 | Idiopathic gout | 0 |
| M1008 | Idiopathic gout | 0 |
| M1009 | Idiopathic gout | 0 |
| M101 | Lead-induced gout | 0 |
| M1010 | Lead-induced gout | 0 |
| M1011 | Lead-induced gout | 0 |
| M1012 | Lead-induced gout | 0 |
| M1013 | Lead-induced gout | 0 |
| M1014 | Lead-induced gout | 0 |
| M1015 | Lead-induced gout | 0 |
| M1016 | Lead-induced gout | 0 |
| M1017 | Lead-induced gout | 0 |
| M1018 | Lead-induced gout | 0 |
| M1019 | Lead-induced gout | 0 |
| M102 | Drug-induced gout | 0 |
| M1020 | Drug-induced gout | 0 |
| M1021 | Drug-induced gout | 0 |
| M1022 | Drug-induced gout | 0 |
| M1023 | Drug-induced gout | 0 |
| M1024 | Drug-induced gout | 0 |
| M1025 | Drug-induced gout | 0 |
| M1026 | Drug-induced gout | 0 |
| M1027 | Drug-induced gout | 0 |
| M1028 | Drug-induced gout | 0 |
| M1029 | Drug-induced gout | 0 |
| M103 | Gout due to impairment of renal function | 0 |
| M1030 | Gout due to impairment of renal function | 0 |
| M1031 | Gout due to impairment of renal function | 0 |
| M1032 | Gout due to impairment of renal function | 0 |
| M1033 | Gout due to impairment of renal function | 0 |
| M1034 | Gout due to impairment of renal function | 0 |
| M1035 | Gout due to impairment of renal function | 0 |
| M1036 | Gout due to impairment of renal function | 0 |
| M1037 | Gout due to impairment of renal function | 0 |
| M1038 | Gout due to impairment of renal function | 0 |
| M1039 | Gout due to impairment of renal function | 0 |
| M104 | Other secondary gout | 0 |
| M1040 | Other secondary gout | 0 |
| M1041 | Other secondary gout | 0 |
| M1042 | Other secondary gout | 0 |
| M1043 | Other secondary gout | 0 |
| M1044 | Other secondary gout | 0 |
| M1045 | Other secondary gout | 0 |
| M1046 | Other secondary gout | 0 |
| M1047 | Other secondary gout | 0 |
| M1048 | Other secondary gout | 0 |
| M1049 | Other secondary gout | 0 |
| M109 | Gout, unspecified | 0 |
| M1090 | Gout, unspecified | 0 |
| M1091 | Gout, unspecified | 0 |
| M1092 | Gout, unspecified | 0 |
| M1093 | Gout, unspecified | 0 |
| M1094 | Gout, unspecified | 0 |
| M1095 | Gout, unspecified | 0 |
| M1096 | Gout, unspecified | 0 |
| M1097 | Gout, unspecified | 0 |
| M1098 | Gout, unspecified | 0 |
| M1099 | Gout, unspecified | 0 |
| M140 | Gouty arthropathy due to enzyme defects and other inherited disorders | 0 |

---

#### CONCEPT: Heart Valve Disorders¶

| ICD10\_CODE | DESCRIPTION\_SAIL | SENSITIVE\_CODE |
| --- | --- | --- |
| G110 | Congenital nonprogressive ataxia | 0 |
| G111 | Early-onset cerebellar ataxia | 0 |
| G112 | Late-onset cerebellar ataxia | 0 |
| G113 | Cerebellar ataxia with defective DNA repair | 0 |
| G114 | Hereditary spastic paraplegia | 0 |
| G118 | Other hereditary ataxias | 0 |
| G119 | Hereditary ataxia, unspecified | 0 |
| I05 | Rheumatic mitral valve diseases | 0 |
| I050 | Mitral stenosis | 0 |
| I051 | Rheumatic mitral insufficiency | 0 |
| I052 | Mitral stenosis with insufficiency | 0 |
| I058 | Other mitral valve diseases | 0 |
| I059 | Mitral valve disease, unspecified | 0 |
| I06 | Rheumatic aortic valve diseases | 0 |
| I060 | Rheumatic aortic stenosis | 0 |
| I062 | Rheumatic aortic stenosis with insufficiency | 0 |
| I068 | Other rheumatic aortic valve diseases | 0 |
| I069 | Rheumatic aortic valve disease, unspecified | 0 |
| I07 | Rheumatic tricuspid valve diseases | 0 |
| I070 | Tricuspid stenosis | 0 |
| I072 | Tricuspid stenosis with insufficiency | 0 |
| I078 | Other tricuspid valve diseases | 0 |
| I079 | Tricuspid valve disease, unspecified | 0 |
| I08 | Multiple valve diseases | 0 |
| I080 | Disorders of both mitral and aortic valves | 0 |
| I081 | Disorders of both mitral and tricuspid valves | 0 |
| I082 | Disorders of both aortic and tricuspid valves | 0 |
| I083 | Combined disorders of mitral, aortic and tricuspid valves | 0 |
| I088 | Other multiple valve diseases | 0 |
| I089 | Multiple valve disease, unspecified | 0 |
| I091 | Rheumatic diseases of endocardium, valve unspecified | 0 |
| I34 | Nonrheumatic mitral valve disorders | 0 |
| I340 | Mitral (valve) insufficiency | 0 |
| I341 | Mitral (valve) prolapse | 0 |
| I342 | Nonrheumatic mitral (valve) stenosis | 0 |
| I348 | Other nonrheumatic mitral valve disorders | 0 |
| I349 | Nonrheumatic mitral valve disorder, unspecified | 0 |
| I35 | Nonrheumatic aortic valve disorders | 0 |
| I350 | Aortic (valve) stenosis | 0 |
| I351 | Aortic (valve) insufficiency | 0 |
| I352 | Aortic (valve) stenosis with insufficiency | 0 |
| I358 | Other aortic valve disorders | 0 |
| I359 | Aortic valve disorder, unspecified | 0 |
| I36 | Nonrheumatic tricuspid valve disorders | 0 |
| I360 | Nonrheumatic tricuspid (valve) stenosis | 0 |
| I361 | Nonrheumatic tricuspid (valve) insufficiency | 0 |
| I362 | Nonrheumatic tricuspid (valve) stenosis with insufficiency | 0 |
| I368 | Other nonrheumatic tricuspid valve disorders | 0 |
| I369 | Nonrheumatic tricuspid valve disorder, unspecified | 0 |
| I37 | Pulmonary valve disorders | 0 |
| I370 | Pulmonary valve stenosis | 0 |
| I371 | Pulmonary valve insufficiency | 0 |
| I372 | Pulmonary valve stenosis with insufficiency | 0 |
| I378 | Other pulmonary valve disorders | 0 |
| I379 | Pulmonary valve disorder, unspecified | 0 |
| I38 | Endocarditis, valve unspecified | 0 |
| I39 | Endocarditis and heart valve disorders in diseases classified elsewhere | 0 |
| I390 | Mitral valve disorders in diseases classified elsewhere | 0 |
| I391 | Aortic valve disorders in diseases classified elsewhere | 0 |
| I392 | Tricuspid valve disorders in diseases classified elsewhere | 0 |
| I393 | Pulmonary valve disorders in diseases classified elsewhere | 0 |
| I394 | Multiple valve disorders in diseases classified elsewhere | 0 |
| I398 | Endocarditis, valve unspecified, in diseases classified elsewhere | 0 |
| Q22 | Congenital malformations of pulmonary and tricuspid valves | 0 |
| Q220 | Pulmonary valve atresia | 0 |
| Q221 | Congenital pulmonary valve stenosis | 0 |
| Q222 | Congenital pulmonary valve insufficiency | 0 |
| Q223 | Other congenital malformations of pulmonary valve | 0 |
| Q224 | Congenital tricuspid stenosis | 0 |
| Q225 | Ebstein anomaly | 0 |
| Q228 | Other congenital malformations of tricuspid valve | 0 |
| Q229 | Congenital malformation of tricuspid valve, unspecified | 0 |
| Q23 | Congenital malformations of aortic and mitral valves | 0 |
| Q230 | Congenital stenosis of aortic valve | 0 |
| Q231 | Congenital insufficiency of aortic valve | 0 |
| Q232 | Congenital mitral stenosis | 0 |
| Q233 | Congenital mitral insufficiency | 0 |
| Q238 | Other congenital malformations of aortic and mitral valves | 0 |
| Q239 | Congenital malformation of aortic and mitral valves, unspecified | 0 |
| Q243 | Pulmonary infundibular stenosis | 0 |
| Q244 | Congenital subaortic stenosis | 0 |
| Q642 | Congenital posterior urethral valves | 0 |
| T820 | Mechanical complication of heart valve prosthesis | 0 |
| T822 | Mechanical complication of coronary artery bypass and valve grafts | 0 |
| T826 | Infection and inflammatory reaction due to cardiac valve prosthesis | 0 |
| Z952 | Presence of prosthetic heart valve | 0 |
| Z953 | Presence of xenogenic heart valve | 0 |
| Z954 | Presence of other heart-valve replacement | 0 |

---

#### CONCEPT: HF (Heart Failure)¶

| ICD10\_CODE | DESCRIPTION\_SAIL | SENSITIVE\_CODE |
| --- | --- | --- |
| I11 | Hypertensive heart disease | 0 |
| I110 | Hypertensive heart disease with (congestive) heart failure | 0 |
| I119 | Hypertensive heart disease without (congestive) heart failure | 0 |
| I13 | Hypertensive heart and renal disease | 0 |
| I130 | Hypertensive heart and renal disease with (congestive) heart failure | 0 |
| I131 | Hypertensive heart and renal disease with renal failure | 0 |
| I132 | Hypertensive heart and renal disease with both (congestive) heart failure and renal failure | 0 |
| I139 | Hypertensive heart and renal disease, unspecified | 0 |
| I255 | Ischaemic cardiomyopathy | 0 |
| I260 | Pulmonary embolism with mention of acute cor pulmonale | 0 |
| I420 | Dilated cardiomyopathy | 0 |
| I421 | Obstructive hypertrophic cardiomyopathy | 0 |
| I422 | Other hypertrophic cardiomyopathy | 0 |
| I429 | Cardiomyopathy, unspecified | 0 |
| I50 | Heart failure | 0 |
| I500 | Congestive heart failure | 0 |
| I501 | Left ventricular failure | 0 |
| I509 | Heart failure, unspecified | 0 |
| P290 | Neonatal cardiac failure | 0 |
| Z994 | Dependence on artificial heart | 0 |

---

#### CONCEPT: HIV/AIDS¶

| ICD10\_CODE | DESCRIPTION\_SAIL | SENSITIVE\_CODE |
| --- | --- | --- |
| B20 | Human immunodeficiency virus [HIV] disease resulting in infectious and parasitic diseases | 1 |
| B200 | HIV disease resulting in mycobacterial infection | 1 |
| B201 | HIV disease resulting in other bacterial infections | 1 |
| B202 | HIV disease resulting in cytomegaloviral disease | 1 |
| B203 | HIV disease resulting in other viral infections | 1 |
| B204 | HIV disease resulting in candidiasis | 1 |
| B205 | HIV disease resulting in other mycoses | 1 |
| B206 | HIV disease resulting in Pneumocystis jirovecii pneumonia | 1 |
| B207 | HIV disease resulting in multiple infections | 1 |
| B208 | HIV disease resulting in other infectious and parasitic diseases | 1 |
| B209 | HIV disease resulting in unspecified infectious or parasitic disease | 1 |
| B21 | Human immunodeficiency virus [HIV] disease resulting in malignant neoplasms | 1 |
| B210 | HIV disease resulting in Kaposi sarcoma | 1 |
| B211 | HIV disease resulting in Burkitt lymphoma | 1 |
| B212 | HIV disease resulting in other types of non-Hodgkin lymphoma | 1 |
| B213 | HIV disease resulting in other malignant neoplasms of lymphoid, haematopoietic and related tissue | 1 |
| B217 | HIV disease resulting in multiple malignant neoplasms | 1 |
| B218 | HIV disease resulting in other malignant neoplasms | 1 |
| B219 | HIV disease resulting in unspecified malignant neoplasm | 1 |
| B22 | Human immunodeficiency virus [HIV] disease resulting in other specified diseases | 1 |
| B220 | HIV disease resulting in encephalopathy | 1 |
| B221 | HIV disease resulting in lymphoid interstitial pneumonitis | 1 |
| B222 | HIV disease resulting in wasting syndrome | 1 |
| B227 | HIV disease resulting in multiple diseases classified elsewhere | 1 |
| B23 | Human immunodeficiency virus [HIV] disease resulting in other conditions | 1 |
| B230 | Acute HIV infection syndrome | 1 |
| B231 | HIV disease resulting in (persistent) generalized lymphadenopathy | 1 |
| B232 | HIV disease resulting in haematological and immunological abnormalities, not elsewhere classified | 1 |
| B238 | HIV disease resulting in other specified conditions | 1 |
| B24 | Unspecified human immunodeficiency virus [HIV] disease | 1 |
| F024 | Dementia in human immunodeficiency virus [HIV] disease | 1 |
| O987 | Human immunodeficiency virus [HIV] disease complicating pregnancy, childbirth and the puerperium | 1 |
| R75 | Laboratory evidence of human immunodeficiency virus [HIV] | 1 |
| Z21 | Asymptomatic human immunodeficiency virus [HIV] infection status | 1 |

---

#### CONCEPT: Hypertension¶

| ICD10\_CODE | DESCRIPTION\_SAIL | SENSITIVE\_CODE |
| --- | --- | --- |
| I10 | Essential (primary) hypertension | 0 |
| I11 | Hypertensive heart disease | 0 |
| I110 | Hypertensive heart disease with (congestive) heart failure | 0 |
| I119 | Hypertensive heart disease without (congestive) heart failure | 0 |
| I12 | Hypertensive renal disease | 0 |
| I120 | Hypertensive renal disease with renal failure | 0 |
| I129 | Hypertensive renal disease without renal failure | 0 |
| I13 | Hypertensive heart and renal disease | 0 |
| I130 | Hypertensive heart and renal disease with (congestive) heart failure | 0 |
| I131 | Hypertensive heart and renal disease with renal failure | 0 |
| I132 | Hypertensive heart and renal disease with both (congestive) heart failure and renal failure | 0 |
| I139 | Hypertensive heart and renal disease, unspecified | 0 |
| I15 | Secondary hypertension | 0 |
| I150 | Renovascular hypertension | 0 |
| I151 | Hypertension secondary to other renal disorders | 0 |
| I152 | Hypertension secondary to endocrine disorders | 0 |
| I158 | Other secondary hypertension | 0 |
| I159 | Secondary hypertension, unspecified | 0 |
| I674 | Hypertensive encephalopathy | 0 |
| O10 | Pre-existing hypertension complicating pregnancy, childbirth and the puerperium | 0 |
| O100 | Pre-existing essential hypertension complicating pregnancy, childbirth and the puerperium | 0 |
| O101 | Pre-existing hypertensive heart disease complicating pregnancy, childbirth and the puerperium | 0 |
| O102 | Pre-existing hypertensive renal disease complicating pregnancy, childbirth and the puerperium | 0 |
| O103 | Pre-existing hypertensive heart and renal disease complicating pregnancy, childbirth and the puerperium | 0 |
| O104 | Pre-existing secondary hypertension complicating pregnancy, childbirth and the puerperium | 0 |
| Y525 | Other antihypertensive drugs, not elsewhere classified | 0 |

---

#### CONCEPT: Hyperthyroidism¶

| ICD10\_CODE | DESCRIPTION\_SAIL | SENSITIVE\_CODE |
| --- | --- | --- |
| E05 | Thyrotoxicosis [hyperthyroidism] | 0 |
| E050 | Thyrotoxicosis with diffuse goitre | 0 |
| E051 | Thyrotoxicosis with toxic single thyroid nodule | 0 |
| E052 | Thyrotoxicosis with toxic multinodular goitre | 0 |
| E053 | Thyrotoxicosis from ectopic thyroid tissue | 0 |
| E059 | Thyrotoxicosis, unspecified | 0 |

---

#### CONCEPT: Hypothyroidism¶

| ICD10\_CODE | DESCRIPTION\_SAIL | SENSITIVE\_CODE |
| --- | --- | --- |
| E001 | Congenital iodine-deficiency syndrome, myxoedematous type | 0 |
| E03 | Other hypothyroidism | 0 |
| E030 | Congenital hypothyroidism with diffuse goitre | 0 |
| E031 | Congenital hypothyroidism without goitre | 0 |
| E032 | Hypothyroidism due to medicaments and other exogenous substances | 0 |
| E033 | Postinfectious hypothyroidism | 0 |
| E034 | Atrophy of thyroid (acquired) | 0 |
| E035 | Myxoedema coma | 0 |
| E038 | Other specified hypothyroidism | 0 |
| E039 | Hypothyroidism, unspecified | 0 |
| E890 | Postprocedural hypothyroidism | 0 |
| G132 | Systemic atrophy primarily affecting central nervous system in myxoedema | 0 |

---

#### CONCEPT: IBD (Inflammatory bowel disease)¶

| ICD10\_CODE | DESCRIPTION\_SAIL | SENSITIVE\_CODE |
| --- | --- | --- |
| K50 | Crohn disease [regional enteritis] | 0 |
| K500 | Crohn disease of small intestine | 0 |
| K501 | Crohn disease of large intestine | 0 |
| K508 | Other Crohn disease | 0 |
| K509 | Crohn disease, unspecified | 0 |
| K51 | Ulcerative colitis | 0 |
| K510 | Ulcerative (chronic) pancolitis | 0 |
| K512 | Ulcerative (chronic) proctitis | 0 |
| K513 | Ulcerative (chronic) rectosigmoiditis | 0 |
| K518 | Other ulcerative colitis | 0 |
| K519 | Ulcerative colitis, unspecified | 0 |
| M074 | Arthropathy in Crohn disease [regional enteritis] | 0 |
| M0740 | Arthropathy in Crohn disease [regional enteritis] | 0 |
| M0741 | Arthropathy in Crohn disease [regional enteritis] | 0 |
| M0742 | Arthropathy in Crohn disease [regional enteritis] | 0 |
| M0743 | Arthropathy in Crohn disease [regional enteritis] | 0 |
| M0744 | Arthropathy in Crohn disease [regional enteritis] | 0 |
| M0745 | Arthropathy in Crohn disease [regional enteritis] | 0 |
| M0746 | Arthropathy in Crohn disease [regional enteritis] | 0 |
| M0747 | Arthropathy in Crohn disease [regional enteritis] | 0 |
| M0748 | Arthropathy in Crohn disease [regional enteritis] | 0 |
| M0749 | Arthropathy in Crohn disease [regional enteritis] | 0 |
| M075 | Arthropathy in ulcerative colitis | 0 |
| M0750 | Arthropathy in ulcerative colitis | 0 |
| M0751 | Arthropathy in ulcerative colitis | 0 |
| M0752 | Arthropathy in ulcerative colitis | 0 |
| M0753 | Arthropathy in ulcerative colitis | 0 |
| M0754 | Arthropathy in ulcerative colitis | 0 |
| M0755 | Arthropathy in ulcerative colitis | 0 |
| M0756 | Arthropathy in ulcerative colitis | 0 |
| M0757 | Arthropathy in ulcerative colitis | 0 |
| M0758 | Arthropathy in ulcerative colitis | 0 |
| M0759 | Arthropathy in ulcerative colitis | 0 |
| M091 | Juvenile arthritis in Crohn disease [regional enteritis] | 0 |
| M0910 | Juvenile arthritis in Crohn disease [regional enteritis] | 0 |
| M0911 | Juvenile arthritis in Crohn disease [regional enteritis] | 0 |
| M0912 | Juvenile arthritis in Crohn disease [regional enteritis] | 0 |
| M0913 | Juvenile arthritis in Crohn disease [regional enteritis] | 0 |
| M0914 | Juvenile arthritis in Crohn disease [regional enteritis] | 0 |
| M0915 | Juvenile arthritis in Crohn disease [regional enteritis] | 0 |
| M0916 | Juvenile arthritis in Crohn disease [regional enteritis] | 0 |
| M0917 | Juvenile arthritis in Crohn disease [regional enteritis] | 0 |
| M0918 | Juvenile arthritis in Crohn disease [regional enteritis] | 0 |
| M0919 | Juvenile arthritis in Crohn disease [regional enteritis] | 0 |
| M092 | Juvenile arthritis in ulcerative colitis | 0 |
| M0920 | Juvenile arthritis in ulcerative colitis | 0 |
| M0921 | Juvenile arthritis in ulcerative colitis | 0 |
| M0922 | Juvenile arthritis in ulcerative colitis | 0 |
| M0923 | Juvenile arthritis in ulcerative colitis | 0 |
| M0924 | Juvenile arthritis in ulcerative colitis | 0 |
| M0925 | Juvenile arthritis in ulcerative colitis | 0 |
| M0926 | Juvenile arthritis in ulcerative colitis | 0 |
| M0927 | Juvenile arthritis in ulcerative colitis | 0 |
| M0928 | Juvenile arthritis in ulcerative colitis | 0 |
| M0929 | Juvenile arthritis in ulcerative colitis | 0 |

---

#### CONCEPT: ILD (Inflammatory Lung Disease)¶

| ICD10\_CODE | DESCRIPTION\_SAIL | SENSITIVE\_CODE |
| --- | --- | --- |
| J703 | Chronic drug-induced interstitial lung disorders | 0 |
| J704 | Drug-induced interstitial lung disorders, unspecified | 0 |
| J841 | Other interstitial pulmonary diseases with fibrosis | 0 |

---

#### CONCEPT: Leukaemia¶

| ICD10\_CODE | DESCRIPTION\_SAIL | SENSITIVE\_CODE |
| --- | --- | --- |
| B213 | HIV disease resulting in other malignant neoplasms of lymphoid, haematopoietic and related tissue | 1 |
| C91 | Lymphoid leukaemia | 0 |
| C910 | Acute lymphoblastic leukaemia [ALL] | 0 |
| C911 | Chronic lymphocytic leukaemia of B-cell type | 0 |
| C913 | Prolymphocytic leukaemia of B-cell type | 0 |
| C914 | Hairy-cell leukaemia | 0 |
| C915 | Adult T-cell lymphoma/leukaemia [HTLV-1-associated] | 0 |
| C916 | Prolymphocytic leukaemia of T-cell type | 0 |
| C917 | Other lymphoid leukaemia | 0 |
| C918 | Mature B-cell leukaemia Burkitt-type | 0 |
| C919 | Lymphoid leukaemia, unspecified | 0 |
| C92 | Myeloid leukaemia | 0 |
| C920 | Acute myeloblastic leukaemia [AML] | 0 |
| C921 | Chronic myeloid leukaemia [CML], BCR/ABL-positive | 0 |
| C922 | Atypical chronic myeloid leukaemia, BCR/ABL-negative | 0 |
| C923 | Myeloid sarcoma | 0 |
| C924 | Acute promyelocytic leukaemia [PML] | 0 |
| C925 | Acute myelomonocytic leukaemia | 0 |
| C926 | Acute myeloid leukaemia with 11q23-abnormality | 0 |
| C927 | Other myeloid leukaemia | 0 |
| C928 | Acute myeloid leukaemia with multilineage dysplasia | 0 |
| C929 | Myeloid leukaemia, unspecified | 0 |
| C93 | Monocytic leukaemia | 0 |
| C930 | Acute monoblastic/monocytic leukaemia | 0 |
| C931 | Chronic myelomonocytic leukaemia | 0 |
| C933 | Juvenile myelomonocytic leukaemia | 0 |
| C937 | Other monocytic leukaemia | 0 |
| C939 | Monocytic leukaemia, unspecified | 0 |
| C94 | Other leukaemias of specified cell type | 0 |
| C940 | Acute erythroid leukaemia | 0 |
| C942 | Acute megakaryoblastic leukaemia | 0 |
| C943 | Mast cell leukaemia | 0 |
| C944 | Acute panmyelosis with myelofibrosis | 0 |
| C946 | Myelodysplastic and myeloproliferative disease, not elsewhere classified | 0 |
| C947 | Other specified leukaemias | 0 |
| C95 | Leukaemia of unspecified cell type | 0 |
| C950 | Acute leukaemia of unspecified cell type | 0 |
| C951 | Chronic leukaemia of unspecified cell type | 0 |
| C957 | Other leukaemia of unspecified cell type | 0 |
| C959 | Leukaemia, unspecified | 0 |
| C96 | Other and unspecified malignant neoplasms of lymphoid, haematopoietic and related tissue | 0 |
| C967 | Other specified malignant neoplasms of lymphoid, haematopoietic and related tissue | 0 |
| C969 | Malignant neoplasm of lymphoid, haematopoietic and related tissue, unspecified | 0 |
| D474 | Osteomyelofibrosis | 0 |
| D475 | Chronic eosinophilic leukaemia [hypereosinophilic syndrome] | 0 |
| Z856 | Personal history of leukaemia | 0 |
| Z857 | Personal history of other malignant neoplasms of lymphoid, haematopoietic and related tissues | 0 |

---

#### CONCEPT: Metastatic cancers¶

| ICD10\_CODE | DESCRIPTION\_SAIL | SENSITIVE\_CODE |
| --- | --- | --- |
| C77 | Secondary and unspecified malignant neoplasm of lymph nodes | 0 |
| C770 | Secondary and unspecified malignant neoplasm: Lymph nodes of head, face and neck | 0 |
| C771 | Secondary and unspecified malignant neoplasm: Intrathoracic lymph nodes | 0 |
| C772 | Secondary and unspecified malignant neoplasm: Intra-abdominal lymph nodes | 0 |
| C773 | Secondary and unspecified malignant neoplasm: Axillary and upper limb lymph nodes | 0 |
| C774 | Secondary and unspecified malignant neoplasm: Inguinal and lower limb lymph nodes | 0 |
| C775 | Secondary and unspecified malignant neoplasm: Intrapelvic lymph nodes | 0 |
| C778 | Secondary and unspecified malignant neoplasm: Lymph nodes of multiple regions | 0 |
| C779 | Secondary and unspecified malignant neoplasm: Lymph node, unspecified | 0 |
| C78 | Secondary malignant neoplasm of respiratory and digestive organs | 0 |
| C780 | Secondary malignant neoplasm of lung | 0 |
| C781 | Secondary malignant neoplasm of mediastinum | 0 |
| C782 | Secondary malignant neoplasm of pleura | 0 |
| C783 | Secondary malignant neoplasm of other and unspecified respiratory organs | 0 |
| C784 | Secondary malignant neoplasm of small intestine | 0 |
| C785 | Secondary malignant neoplasm of large intestine and rectum | 0 |
| C786 | Secondary malignant neoplasm of retroperitoneum and peritoneum | 0 |
| C787 | Secondary malignant neoplasm of liver and intrahepatic bile duct | 0 |
| C788 | Secondary malignant neoplasm of other and unspecified digestive organs | 0 |
| C79 | Secondary malignant neoplasm of other and unspecified sites | 0 |
| C790 | Secondary malignant neoplasm of kidney and renal pelvis | 0 |
| C791 | Secondary malignant neoplasm of bladder and other and unspecified urinary organs | 0 |
| C792 | Secondary malignant neoplasm of skin | 0 |
| C793 | Secondary malignant neoplasm of brain and cerebral meninges | 0 |
| C794 | Secondary malignant neoplasm of other and unspecified parts of nervous system | 0 |
| C795 | Secondary malignant neoplasm of bone and bone marrow | 0 |
| C796 | Secondary malignant neoplasm of ovary | 0 |
| C797 | Secondary malignant neoplasm of adrenal gland | 0 |
| C798 | Secondary malignant neoplasm of other specified sites | 0 |
| C799 | Secondary malignant neoplasm, unspecified site | 0 |

---

#### CONCEPT: Multiple sclerosis¶

| ICD10\_CODE | DESCRIPTION\_SAIL | SENSITIVE\_CODE |
| --- | --- | --- |
| G35 | Multiple sclerosis | 0 |

---

#### CONCEPT: Osteoarthritis¶

| ICD10\_CODE | DESCRIPTION\_SAIL | SENSITIVE\_CODE |
| --- | --- | --- |
| G552 | Nerve root and plexus compressions in spondylosis | 0 |
| M15 | Polyarthrosis | 0 |
| M150 | Primary generalized (osteo)arthrosis | 0 |
| M153 | Secondary multiple arthrosis | 0 |
| M154 | Erosive (osteo)arthrosis | 0 |
| M158 | Other polyarthrosis | 0 |
| M159 | Polyarthrosis, unspecified | 0 |
| M16 | Coxarthrosis [arthrosis of hip] | 0 |
| M160 | Primary coxarthrosis, bilateral | 0 |
| M161 | Other primary coxarthrosis | 0 |
| M164 | Post-traumatic coxarthrosis, bilateral | 0 |
| M165 | Other post-traumatic coxarthrosis | 0 |
| M166 | Other secondary coxarthrosis, bilateral | 0 |
| M167 | Other secondary coxarthrosis | 0 |
| M169 | Coxarthrosis, unspecified | 0 |
| M17 | Gonarthrosis [arthrosis of knee] | 0 |
| M170 | Primary gonarthrosis, bilateral | 0 |
| M171 | Other primary gonarthrosis | 0 |
| M172 | Post-traumatic gonarthrosis, bilateral | 0 |
| M173 | Other post-traumatic gonarthrosis | 0 |
| M174 | Other secondary gonarthrosis, bilateral | 0 |
| M175 | Other secondary gonarthrosis | 0 |
| M179 | Gonarthrosis, unspecified | 0 |
| M18 | Arthrosis of first carpometacarpal joint | 0 |
| M180 | Primary arthrosis of first carpometacarpal joints, bilateral | 0 |
| M181 | Other primary arthrosis of first carpometacarpal joint | 0 |
| M182 | Post-traumatic arthrosis of first carpometacarpal joints, bilateral | 0 |
| M183 | Other post-traumatic arthrosis of first carpometacarpal joint | 0 |
| M184 | Other secondary arthrosis of first carpometacarpal joints, bilateral | 0 |
| M185 | Other secondary arthrosis of first carpometacarpal joint | 0 |
| M189 | Arthrosis of first carpometacarpal joint, unspecified | 0 |
| M19 | Other arthrosis | 0 |
| M190 | Primary arthrosis of other joints | 0 |
| M1900 | Primary arthrosis of other joints | 0 |
| M1901 | Primary arthrosis of other joints | 0 |
| M1902 | Primary arthrosis of other joints | 0 |
| M1903 | Primary arthrosis of other joints | 0 |
| M1904 | Primary arthrosis of other joints | 0 |
| M1905 | Primary arthrosis of other joints | 0 |
| M1907 | Primary arthrosis of other joints | 0 |
| M1908 | Primary arthrosis of other joints | 0 |
| M1909 | Primary arthrosis of other joints | 0 |
| M191 | Post-traumatic arthrosis of other joints | 0 |
| M1910 | Post-traumatic arthrosis of other joints | 0 |
| M1911 | Post-traumatic arthrosis of other joints | 0 |
| M1912 | Post-traumatic arthrosis of other joints | 0 |
| M1913 | Post-traumatic arthrosis of other joints | 0 |
| M1914 | Post-traumatic arthrosis of other joints | 0 |
| M1915 | Post-traumatic arthrosis of other joints | 0 |
| M1917 | Post-traumatic arthrosis of other joints | 0 |
| M1918 | Post-traumatic arthrosis of other joints | 0 |
| M1919 | Post-traumatic arthrosis of other joints | 0 |
| M192 | Other secondary arthrosis | 0 |
| M1920 | Other secondary arthrosis | 0 |
| M1921 | Other secondary arthrosis | 0 |
| M1922 | Other secondary arthrosis | 0 |
| M1923 | Other secondary arthrosis | 0 |
| M1924 | Other secondary arthrosis | 0 |
| M1925 | Other secondary arthrosis | 0 |
| M1927 | Other secondary arthrosis | 0 |
| M1928 | Other secondary arthrosis | 0 |
| M1929 | Other secondary arthrosis | 0 |
| M198 | Other specified arthrosis | 0 |
| M1980 | Other specified arthrosis | 0 |
| M1981 | Other specified arthrosis | 0 |
| M1982 | Other specified arthrosis | 0 |
| M1983 | Other specified arthrosis | 0 |
| M1984 | Other specified arthrosis | 0 |
| M1985 | Other specified arthrosis | 0 |
| M1986 | Other specified arthrosis | 0 |
| M1987 | Other specified arthrosis | 0 |
| M1988 | Other specified arthrosis | 0 |
| M1989 | Other specified arthrosis | 0 |
| M199 | Arthrosis, unspecified | 0 |
| M1990 | Arthrosis, unspecified | 0 |
| M1991 | Arthrosis, unspecified | 0 |
| M1992 | Arthrosis, unspecified | 0 |
| M1993 | Arthrosis, unspecified | 0 |
| M1994 | Arthrosis, unspecified | 0 |
| M1995 | Arthrosis, unspecified | 0 |
| M1997 | Arthrosis, unspecified | 0 |
| M1998 | Arthrosis, unspecified | 0 |
| M1999 | Arthrosis, unspecified | 0 |
| M47 | Spondylosis | 0 |
| M471 | Other spondylosis with myelopathy | 0 |
| M4710 | Other spondylosis with myelopathy | 0 |
| M4711 | Other spondylosis with myelopathy | 0 |
| M4712 | Other spondylosis with myelopathy | 0 |
| M4713 | Other spondylosis with myelopathy | 0 |
| M4714 | Other spondylosis with myelopathy | 0 |
| M4715 | Other spondylosis with myelopathy | 0 |
| M4716 | Other spondylosis with myelopathy | 0 |
| M4717 | Other spondylosis with myelopathy | 0 |
| M4718 | Other spondylosis with myelopathy | 0 |
| M4719 | Other spondylosis with myelopathy | 0 |
| M472 | Other spondylosis with radiculopathy | 0 |
| M4720 | Other spondylosis with radiculopathy | 0 |
| M4721 | Other spondylosis with radiculopathy | 0 |
| M4722 | Other spondylosis with radiculopathy | 0 |
| M4723 | Other spondylosis with radiculopathy | 0 |
| M4724 | Other spondylosis with radiculopathy | 0 |
| M4725 | Other spondylosis with radiculopathy | 0 |
| M4726 | Other spondylosis with radiculopathy | 0 |
| M4727 | Other spondylosis with radiculopathy | 0 |
| M4728 | Other spondylosis with radiculopathy | 0 |
| M4729 | Other spondylosis with radiculopathy | 0 |
| M478 | Other spondylosis | 0 |
| M4780 | Other spondylosis | 0 |
| M4781 | Other spondylosis | 0 |
| M4782 | Other spondylosis | 0 |
| M4783 | Other spondylosis | 0 |
| M4784 | Other spondylosis | 0 |
| M4785 | Other spondylosis | 0 |
| M4786 | Other spondylosis | 0 |
| M4787 | Other spondylosis | 0 |
| M4788 | Other spondylosis | 0 |
| M4789 | Other spondylosis | 0 |
| M479 | Spondylosis, unspecified | 0 |
| M4790 | Spondylosis, unspecified | 0 |
| M4791 | Spondylosis, unspecified | 0 |
| M4792 | Spondylosis, unspecified | 0 |
| M4793 | Spondylosis, unspecified | 0 |
| M4794 | Spondylosis, unspecified | 0 |
| M4795 | Spondylosis, unspecified | 0 |
| M4796 | Spondylosis, unspecified | 0 |
| M4797 | Spondylosis, unspecified | 0 |
| M4798 | Spondylosis, unspecified | 0 |
| M4799 | Spondylosis, unspecified | 0 |

---

#### CONCEPT: Osteoporosis¶

| ICD10\_CODE | DESCRIPTION\_SAIL | SENSITIVE\_CODE |
| --- | --- | --- |
| M80 | Osteoporosis with pathological fracture | 0 |
| M800 | Postmenopausal osteoporosis with pathological fracture | 0 |
| M8000 | Postmenopausal osteoporosis with pathological fracture | 0 |
| M8001 | Postmenopausal osteoporosis with pathological fracture | 0 |
| M8002 | Postmenopausal osteoporosis with pathological fracture | 0 |
| M8003 | Postmenopausal osteoporosis with pathological fracture | 0 |
| M8004 | Postmenopausal osteoporosis with pathological fracture | 0 |
| M8005 | Postmenopausal osteoporosis with pathological fracture | 0 |
| M8006 | Postmenopausal osteoporosis with pathological fracture | 0 |
| M8007 | Postmenopausal osteoporosis with pathological fracture | 0 |
| M8008 | Postmenopausal osteoporosis with pathological fracture | 0 |
| M8009 | Postmenopausal osteoporosis with pathological fracture | 0 |
| M801 | Postoophorectomy osteoporosis with pathological fracture | 0 |
| M8010 | Postoophorectomy osteoporosis with pathological fracture | 0 |
| M8011 | Postoophorectomy osteoporosis with pathological fracture | 0 |
| M8012 | Postoophorectomy osteoporosis with pathological fracture | 0 |
| M8013 | Postoophorectomy osteoporosis with pathological fracture | 0 |
| M8014 | Postoophorectomy osteoporosis with pathological fracture | 0 |
| M8015 | Postoophorectomy osteoporosis with pathological fracture | 0 |
| M8016 | Postoophorectomy osteoporosis with pathological fracture | 0 |
| M8017 | Postoophorectomy osteoporosis with pathological fracture | 0 |
| M8018 | Postoophorectomy osteoporosis with pathological fracture | 0 |
| M8019 | Postoophorectomy osteoporosis with pathological fracture | 0 |
| M802 | Osteoporosis of disuse with pathological fracture | 0 |
| M8020 | Osteoporosis of disuse with pathological fracture | 0 |
| M8021 | Osteoporosis of disuse with pathological fracture | 0 |
| M8022 | Osteoporosis of disuse with pathological fracture | 0 |
| M8023 | Osteoporosis of disuse with pathological fracture | 0 |
| M8024 | Osteoporosis of disuse with pathological fracture | 0 |
| M8025 | Osteoporosis of disuse with pathological fracture | 0 |
| M8026 | Osteoporosis of disuse with pathological fracture | 0 |
| M8027 | Osteoporosis of disuse with pathological fracture | 0 |
| M8028 | Osteoporosis of disuse with pathological fracture | 0 |
| M8029 | Osteoporosis of disuse with pathological fracture | 0 |
| M803 | Postsurgical malabsorption osteoporosis with pathological fracture | 0 |
| M8030 | Postsurgical malabsorption osteoporosis with pathological fracture | 0 |
| M8031 | Postsurgical malabsorption osteoporosis with pathological fracture | 0 |
| M8032 | Postsurgical malabsorption osteoporosis with pathological fracture | 0 |
| M8033 | Postsurgical malabsorption osteoporosis with pathological fracture | 0 |
| M8034 | Postsurgical malabsorption osteoporosis with pathological fracture | 0 |
| M8035 | Postsurgical malabsorption osteoporosis with pathological fracture | 0 |
| M8036 | Postsurgical malabsorption osteoporosis with pathological fracture | 0 |
| M8037 | Postsurgical malabsorption osteoporosis with pathological fracture | 0 |
| M8038 | Postsurgical malabsorption osteoporosis with pathological fracture | 0 |
| M8039 | Postsurgical malabsorption osteoporosis with pathological fracture | 0 |
| M804 | Drug-induced osteoporosis with pathological fracture | 0 |
| M8040 | Drug-induced osteoporosis with pathological fracture | 0 |
| M8041 | Drug-induced osteoporosis with pathological fracture | 0 |
| M8042 | Drug-induced osteoporosis with pathological fracture | 0 |
| M8043 | Drug-induced osteoporosis with pathological fracture | 0 |
| M8044 | Drug-induced osteoporosis with pathological fracture | 0 |
| M8045 | Drug-induced osteoporosis with pathological fracture | 0 |
| M8046 | Drug-induced osteoporosis with pathological fracture | 0 |
| M8047 | Drug-induced osteoporosis with pathological fracture | 0 |
| M8048 | Drug-induced osteoporosis with pathological fracture | 0 |
| M8049 | Drug-induced osteoporosis with pathological fracture | 0 |
| M805 | Idiopathic osteoporosis with pathological fracture | 0 |
| M8050 | Idiopathic osteoporosis with pathological fracture | 0 |
| M8051 | Idiopathic osteoporosis with pathological fracture | 0 |
| M8052 | Idiopathic osteoporosis with pathological fracture | 0 |
| M8053 | Idiopathic osteoporosis with pathological fracture | 0 |
| M8054 | Idiopathic osteoporosis with pathological fracture | 0 |
| M8055 | Idiopathic osteoporosis with pathological fracture | 0 |
| M8056 | Idiopathic osteoporosis with pathological fracture | 0 |
| M8057 | Idiopathic osteoporosis with pathological fracture | 0 |
| M8058 | Idiopathic osteoporosis with pathological fracture | 0 |
| M8059 | Idiopathic osteoporosis with pathological fracture | 0 |
| M808 | Other osteoporosis with pathological fracture | 0 |
| M8080 | Other osteoporosis with pathological fracture | 0 |
| M8081 | Other osteoporosis with pathological fracture | 0 |
| M8082 | Other osteoporosis with pathological fracture | 0 |
| M8083 | Other osteoporosis with pathological fracture | 0 |
| M8084 | Other osteoporosis with pathological fracture | 0 |
| M8085 | Other osteoporosis with pathological fracture | 0 |
| M8086 | Other osteoporosis with pathological fracture | 0 |
| M8087 | Other osteoporosis with pathological fracture | 0 |
| M8088 | Other osteoporosis with pathological fracture | 0 |
| M8089 | Other osteoporosis with pathological fracture | 0 |
| M809 | Unspecified osteoporosis with pathological fracture | 0 |
| M8090 | Unspecified osteoporosis with pathological fracture | 0 |
| M8091 | Unspecified osteoporosis with pathological fracture | 0 |
| M8092 | Unspecified osteoporosis with pathological fracture | 0 |
| M8093 | Unspecified osteoporosis with pathological fracture | 0 |
| M8094 | Unspecified osteoporosis with pathological fracture | 0 |
| M8095 | Unspecified osteoporosis with pathological fracture | 0 |
| M8096 | Unspecified osteoporosis with pathological fracture | 0 |
| M8097 | Unspecified osteoporosis with pathological fracture | 0 |
| M8098 | Unspecified osteoporosis with pathological fracture | 0 |
| M8099 | Unspecified osteoporosis with pathological fracture | 0 |
| M81 | Osteoporosis without pathological fracture | 0 |
| M810 | Postmenopausal osteoporosis | 0 |
| M8100 | Postmenopausal osteoporosis | 0 |
| M8101 | Postmenopausal osteoporosis | 0 |
| M8102 | Postmenopausal osteoporosis | 0 |
| M8103 | Postmenopausal osteoporosis | 0 |
| M8104 | Postmenopausal osteoporosis | 0 |
| M8105 | Postmenopausal osteoporosis | 0 |
| M8106 | Postmenopausal osteoporosis | 0 |
| M8107 | Postmenopausal osteoporosis | 0 |
| M8108 | Postmenopausal osteoporosis | 0 |
| M8109 | Postmenopausal osteoporosis | 0 |
| M811 | Postoophorectomy osteoporosis | 0 |
| M8110 | Postoophorectomy osteoporosis | 0 |
| M8111 | Postoophorectomy osteoporosis | 0 |
| M8112 | Postoophorectomy osteoporosis | 0 |
| M8113 | Postoophorectomy osteoporosis | 0 |
| M8114 | Postoophorectomy osteoporosis | 0 |
| M8115 | Postoophorectomy osteoporosis | 0 |
| M8116 | Postoophorectomy osteoporosis | 0 |
| M8117 | Postoophorectomy osteoporosis | 0 |
| M8118 | Postoophorectomy osteoporosis | 0 |
| M8119 | Postoophorectomy osteoporosis | 0 |
| M812 | Osteoporosis of disuse | 0 |
| M8120 | Osteoporosis of disuse | 0 |
| M8121 | Osteoporosis of disuse | 0 |
| M8122 | Osteoporosis of disuse | 0 |
| M8123 | Osteoporosis of disuse | 0 |
| M8124 | Osteoporosis of disuse | 0 |
| M8125 | Osteoporosis of disuse | 0 |
| M8126 | Osteoporosis of disuse | 0 |
| M8127 | Osteoporosis of disuse | 0 |
| M8128 | Osteoporosis of disuse | 0 |
| M8129 | Osteoporosis of disuse | 0 |
| M813 | Postsurgical malabsorption osteoporosis | 0 |
| M8130 | Postsurgical malabsorption osteoporosis | 0 |
| M8131 | Postsurgical malabsorption osteoporosis | 0 |
| M8132 | Postsurgical malabsorption osteoporosis | 0 |
| M8133 | Postsurgical malabsorption osteoporosis | 0 |
| M8134 | Postsurgical malabsorption osteoporosis | 0 |
| M8135 | Postsurgical malabsorption osteoporosis | 0 |
| M8136 | Postsurgical malabsorption osteoporosis | 0 |
| M8137 | Postsurgical malabsorption osteoporosis | 0 |
| M8138 | Postsurgical malabsorption osteoporosis | 0 |
| M8139 | Postsurgical malabsorption osteoporosis | 0 |
| M814 | Drug-induced osteoporosis | 0 |
| M8140 | Drug-induced osteoporosis | 0 |
| M8141 | Drug-induced osteoporosis | 0 |
| M8142 | Drug-induced osteoporosis | 0 |
| M8143 | Drug-induced osteoporosis | 0 |
| M8144 | Drug-induced osteoporosis | 0 |
| M8145 | Drug-induced osteoporosis | 0 |
| M8146 | Drug-induced osteoporosis | 0 |
| M8147 | Drug-induced osteoporosis | 0 |
| M8148 | Drug-induced osteoporosis | 0 |
| M8149 | Drug-induced osteoporosis | 0 |
| M815 | Idiopathic osteoporosis | 0 |
| M8150 | Idiopathic osteoporosis | 0 |
| M8151 | Idiopathic osteoporosis | 0 |
| M8152 | Idiopathic osteoporosis | 0 |
| M8153 | Idiopathic osteoporosis | 0 |
| M8154 | Idiopathic osteoporosis | 0 |
| M8155 | Idiopathic osteoporosis | 0 |
| M8156 | Idiopathic osteoporosis | 0 |
| M8157 | Idiopathic osteoporosis | 0 |
| M8158 | Idiopathic osteoporosis | 0 |
| M8159 | Idiopathic osteoporosis | 0 |
| M816 | Localized osteoporosis [Lequesne] | 0 |
| M8160 | Localized osteoporosis [Lequesne] | 0 |
| M8161 | Localized osteoporosis [Lequesne] | 0 |
| M8162 | Localized osteoporosis [Lequesne] | 0 |
| M8163 | Localized osteoporosis [Lequesne] | 0 |
| M8164 | Localized osteoporosis [Lequesne] | 0 |
| M8165 | Localized osteoporosis [Lequesne] | 0 |
| M8166 | Localized osteoporosis [Lequesne] | 0 |
| M8167 | Localized osteoporosis [Lequesne] | 0 |
| M8168 | Localized osteoporosis [Lequesne] | 0 |
| M8169 | Localized osteoporosis [Lequesne] | 0 |
| M818 | Other osteoporosis | 0 |
| M8180 | Other osteoporosis | 0 |
| M8181 | Other osteoporosis | 0 |
| M8182 | Other osteoporosis | 0 |
| M8183 | Other osteoporosis | 0 |
| M8184 | Other osteoporosis | 0 |
| M8185 | Other osteoporosis | 0 |
| M8186 | Other osteoporosis | 0 |
| M8187 | Other osteoporosis | 0 |
| M8188 | Other osteoporosis | 0 |
| M8189 | Other osteoporosis | 0 |
| M819 | Osteoporosis, unspecified | 0 |
| M8190 | Osteoporosis, unspecified | 0 |
| M8191 | Osteoporosis, unspecified | 0 |
| M8192 | Osteoporosis, unspecified | 0 |
| M8193 | Osteoporosis, unspecified | 0 |
| M8194 | Osteoporosis, unspecified | 0 |
| M8195 | Osteoporosis, unspecified | 0 |
| M8196 | Osteoporosis, unspecified | 0 |
| M8197 | Osteoporosis, unspecified | 0 |
| M8198 | Osteoporosis, unspecified | 0 |
| M8199 | Osteoporosis, unspecified | 0 |
| M82 | Osteoporosis in diseases classified elsewhere | 0 |
| M820 | Osteoporosis in multiple myelomatosis | 0 |
| M8200 | Osteoporosis in multiple myelomatosis | 0 |
| M8201 | Osteoporosis in multiple myelomatosis | 0 |
| M8202 | Osteoporosis in multiple myelomatosis | 0 |
| M8203 | Osteoporosis in multiple myelomatosis | 0 |
| M8204 | Osteoporosis in multiple myelomatosis | 0 |
| M8205 | Osteoporosis in multiple myelomatosis | 0 |
| M8206 | Osteoporosis in multiple myelomatosis | 0 |
| M8207 | Osteoporosis in multiple myelomatosis | 0 |
| M8208 | Osteoporosis in multiple myelomatosis | 0 |
| M8209 | Osteoporosis in multiple myelomatosis | 0 |
| M821 | Osteoporosis in endocrine disorders | 0 |
| M8210 | Osteoporosis in endocrine disorders | 0 |
| M8211 | Osteoporosis in endocrine disorders | 0 |
| M8212 | Osteoporosis in endocrine disorders | 0 |
| M8213 | Osteoporosis in endocrine disorders | 0 |
| M8214 | Osteoporosis in endocrine disorders | 0 |
| M8215 | Osteoporosis in endocrine disorders | 0 |
| M8216 | Osteoporosis in endocrine disorders | 0 |
| M8217 | Osteoporosis in endocrine disorders | 0 |
| M8218 | Osteoporosis in endocrine disorders | 0 |
| M8219 | Osteoporosis in endocrine disorders | 0 |
| M828 | Osteoporosis in other diseases classified elsewhere | 0 |
| M8280 | Osteoporosis in other diseases classified elsewhere | 0 |
| M8281 | Osteoporosis in other diseases classified elsewhere | 0 |
| M8282 | Osteoporosis in other diseases classified elsewhere | 0 |
| M8283 | Osteoporosis in other diseases classified elsewhere | 0 |
| M8284 | Osteoporosis in other diseases classified elsewhere | 0 |
| M8285 | Osteoporosis in other diseases classified elsewhere | 0 |
| M8286 | Osteoporosis in other diseases classified elsewhere | 0 |
| M8287 | Osteoporosis in other diseases classified elsewhere | 0 |
| M8288 | Osteoporosis in other diseases classified elsewhere | 0 |
| M8289 | Osteoporosis in other diseases classified elsewhere | 0 |

---

#### CONCEPT: Pancreatic disease¶

| ICD10\_CODE | DESCRIPTION\_SAIL | SENSITIVE\_CODE |
| --- | --- | --- |
| K860 | Alcohol-induced chronic pancreatitis | 0 |
| K861 | Other chronic pancreatitis | 0 |

---

#### CONCEPT: Paralysis¶

| ICD10\_CODE | DESCRIPTION\_SAIL | SENSITIVE\_CODE |
| --- | --- | --- |
| G041 | Tropical spastic paraplegia | 0 |
| G114 | Hereditary spastic paraplegia | 0 |
| G800 | Spastic quadriplegic cerebral palsy | 0 |
| G802 | Spastic hemiplegic cerebral palsy | 0 |
| G81 | Hemiplegia | 0 |
| G810 | Flaccid hemiplegia | 0 |
| G811 | Spastic hemiplegia | 0 |
| G819 | Hemiplegia, unspecified | 0 |
| G82 | Paraplegia and tetraplegia | 0 |
| G820 | Flaccid paraplegia | 0 |
| G821 | Spastic paraplegia | 0 |
| G822 | Paraplegia, unspecified | 0 |
| G823 | Flaccid tetraplegia | 0 |
| G824 | Spastic tetraplegia | 0 |
| G825 | Tetraplegia, unspecified | 0 |

---

#### CONCEPT: Parkinson's disease¶

| ICD10\_CODE | DESCRIPTION\_SAIL | SENSITIVE\_CODE |
| --- | --- | --- |
| F023 | Dementia in Parkinson disease | 0 |
| G20 | Parkinson disease | 0 |

---

#### CONCEPT: Plasmacell¶

| ICD10\_CODE | DESCRIPTION\_SAIL | SENSITIVE\_CODE |
| --- | --- | --- |
| C882 | Other heavy chain disease | 0 |
| C90 | Multiple myeloma and malignant plasma cell neoplasms | 0 |
| C900 | Multiple myeloma | 0 |
| C901 | Plasma cell leukaemia | 0 |
| C902 | Extramedullary plasmacytoma | 0 |
| C903 | Solitary plasmacytoma | 0 |
| M820 | Osteoporosis in multiple myelomatosis | 0 |
| M8200 | Osteoporosis in multiple myelomatosis | 0 |
| M8201 | Osteoporosis in multiple myelomatosis | 0 |
| M8202 | Osteoporosis in multiple myelomatosis | 0 |
| M8203 | Osteoporosis in multiple myelomatosis | 0 |
| M8204 | Osteoporosis in multiple myelomatosis | 0 |
| M8205 | Osteoporosis in multiple myelomatosis | 0 |
| M8206 | Osteoporosis in multiple myelomatosis | 0 |
| M8207 | Osteoporosis in multiple myelomatosis | 0 |
| M8208 | Osteoporosis in multiple myelomatosis | 0 |
| M8209 | Osteoporosis in multiple myelomatosis | 0 |

---

#### CONCEPT: PMR (Polymyalgia reheumatica)¶

| ICD10\_CODE | DESCRIPTION\_SAIL | SENSITIVE\_CODE |
| --- | --- | --- |
| M315 | Giant cell arteritis with polymyalgia rheumatica | 0 |
| M353 | Polymyalgia rheumatica | 0 |

---

#### CONCEPT: Primary Lung Cancer¶

| ICD10\_CODE | DESCRIPTION\_SAIL | SENSITIVE\_CODE |
| --- | --- | --- |
| C33 | Malignant neoplasm of trachea | 0 |
| C34 | Malignant neoplasm of bronchus and lung | 0 |
| C39 | Malignant neoplasm of other and ill-defined sites in the respiratory system and intrathoracic organs | 0 |

---

#### CONCEPT: Prostate Cancer¶

| ICD10\_CODE | DESCRIPTION\_SAIL | SENSITIVE\_CODE |
| --- | --- | --- |
| C61 | Malignant neoplasm of prostate | 0 |

---

#### CONCEPT: Psoriasis¶

| ICD10\_CODE | DESCRIPTION\_SAIL | SENSITIVE\_CODE |
| --- | --- | --- |
| L40 | Psoriasis | 0 |
| L400 | Psoriasis vulgaris | 0 |
| L401 | Generalized pustular psoriasis | 0 |
| L404 | Guttate psoriasis | 0 |
| L405 | Arthropathic psoriasis | 0 |
| L408 | Other psoriasis | 0 |
| L409 | Psoriasis, unspecified | 0 |

---

#### CONCEPT: Psoriasis Arthritis¶

| ICD10\_CODE | DESCRIPTION\_SAIL | SENSITIVE\_CODE |
| --- | --- | --- |
| L405 | Arthropathic psoriasis | 0 |
| M07 | Psoriatic and enteropathic arthropathies | 0 |
| M070 | Distal interphalangeal psoriatic arthropathy | 0 |
| M0700 | Distal interphalangeal psoriatic arthropathy | 0 |
| M0704 | Distal interphalangeal psoriatic arthropathy | 0 |
| M0707 | Distal interphalangeal psoriatic arthropathy | 0 |
| M0709 | Distal interphalangeal psoriatic arthropathy | 0 |
| M071 | Arthritis mutilans | 0 |
| M0710 | Arthritis mutilans | 0 |
| M0711 | Arthritis mutilans | 0 |
| M0712 | Arthritis mutilans | 0 |
| M0713 | Arthritis mutilans | 0 |
| M0714 | Arthritis mutilans | 0 |
| M0715 | Arthritis mutilans | 0 |
| M0716 | Arthritis mutilans | 0 |
| M0717 | Arthritis mutilans | 0 |
| M0718 | Arthritis mutilans | 0 |
| M0719 | Arthritis mutilans | 0 |
| M073 | Other psoriatic arthropathies | 0 |
| M0730 | Other psoriatic arthropathies | 0 |
| M0731 | Other psoriatic arthropathies | 0 |
| M0732 | Other psoriatic arthropathies | 0 |
| M0733 | Other psoriatic arthropathies | 0 |
| M0734 | Other psoriatic arthropathies | 0 |
| M0735 | Other psoriatic arthropathies | 0 |
| M0736 | Other psoriatic arthropathies | 0 |
| M0737 | Other psoriatic arthropathies | 0 |
| M0738 | Other psoriatic arthropathies | 0 |
| M0739 | Other psoriatic arthropathies | 0 |
| M090 | Juvenile arthritis in psoriasis | 0 |
| M0900 | Juvenile arthritis in psoriasis | 0 |
| M0901 | Juvenile arthritis in psoriasis | 0 |
| M0902 | Juvenile arthritis in psoriasis | 0 |
| M0903 | Juvenile arthritis in psoriasis | 0 |
| M0904 | Juvenile arthritis in psoriasis | 0 |
| M0905 | Juvenile arthritis in psoriasis | 0 |
| M0906 | Juvenile arthritis in psoriasis | 0 |
| M0907 | Juvenile arthritis in psoriasis | 0 |
| M0908 | Juvenile arthritis in psoriasis | 0 |
| M0909 | Juvenile arthritis in psoriasis | 0 |

---

#### CONCEPT: PTSD (Post-traumatic stress disorders)¶

| ICD10\_CODE | DESCRIPTION\_SAIL | SENSITIVE\_CODE |
| --- | --- | --- |
| F431 | Post-traumatic stress disorder | 0 |

---

#### CONCEPT: Pheripheral vascular disease¶

| ICD10\_CODE | DESCRIPTION\_SAIL | SENSITIVE\_CODE |
| --- | --- | --- |
| I70 | Atherosclerosis | 0 |
| I700 | Atherosclerosis of aorta | 0 |
| I7000 | Atherosclerosis of aorta | 0 |
| I7001 | Atherosclerosis of aorta | 0 |
| I701 | Atherosclerosis of renal artery | 0 |
| I7010 | Atherosclerosis of renal artery | 0 |
| I7011 | Atherosclerosis of renal artery | 0 |
| I702 | Atherosclerosis of arteries of extremities | 0 |
| I7020 | Atherosclerosis of arteries of extremities | 0 |
| I7021 | Atherosclerosis of arteries of extremities | 0 |
| I708 | Atherosclerosis of other arteries | 0 |
| I7080 | Atherosclerosis of other arteries | 0 |
| I7081 | Atherosclerosis of other arteries | 0 |
| I709 | Generalized and unspecified atherosclerosis | 0 |
| I7090 | Generalized and unspecified atherosclerosis | 0 |
| I7091 | Generalized and unspecified atherosclerosis | 0 |
| I73 | Other peripheral vascular diseases | 0 |
| I730 | Raynaud syndrome | 0 |
| I731 | Thromboangiitis obliterans [Buerger] | 0 |
| I738 | Other specified peripheral vascular diseases | 0 |
| I739 | Peripheral vascular disease, unspecified | 0 |
| I792 | Peripheral angiopathy in diseases classified elsewhere | 0 |

---

#### CONCEPT: Renal transplant¶

| ICD10\_CODE | DESCRIPTION\_SAIL | SENSITIVE\_CODE |
| --- | --- | --- |
| T861 | Kidney transplant failure and rejection | 0 |
| Z940 | Kidney transplant status | 0 |

---

#### CONCEPT: Rheumatoid Arthritis¶

| ICD10\_CODE | DESCRIPTION\_SAIL | SENSITIVE\_CODE |
| --- | --- | --- |
| J990 | Rheumatoid lung disease | 0 |
| M05 | Seropositive rheumatoid arthritis | 0 |
| M050 | Felty syndrome | 0 |
| M0500 | Felty syndrome | 0 |
| M0501 | Felty syndrome | 0 |
| M0502 | Felty syndrome | 0 |
| M0503 | Felty syndrome | 0 |
| M0504 | Felty syndrome | 0 |
| M0505 | Felty syndrome | 0 |
| M0506 | Felty syndrome | 0 |
| M0507 | Felty syndrome | 0 |
| M0508 | Felty syndrome | 0 |
| M0509 | Felty syndrome | 0 |
| M051 | Rheumatoid lung disease | 0 |
| M0510 | Rheumatoid lung disease | 0 |
| M0511 | Rheumatoid lung disease | 0 |
| M0512 | Rheumatoid lung disease | 0 |
| M0513 | Rheumatoid lung disease | 0 |
| M0514 | Rheumatoid lung disease | 0 |
| M0515 | Rheumatoid lung disease | 0 |
| M0516 | Rheumatoid lung disease | 0 |
| M0517 | Rheumatoid lung disease | 0 |
| M0518 | Rheumatoid lung disease | 0 |
| M0519 | Rheumatoid lung disease | 0 |
| M052 | Rheumatoid vasculitis | 0 |
| M0520 | Rheumatoid vasculitis | 0 |
| M0521 | Rheumatoid vasculitis | 0 |
| M0522 | Rheumatoid vasculitis | 0 |
| M0523 | Rheumatoid vasculitis | 0 |
| M0524 | Rheumatoid vasculitis | 0 |
| M0525 | Rheumatoid vasculitis | 0 |
| M0526 | Rheumatoid vasculitis | 0 |
| M0527 | Rheumatoid vasculitis | 0 |
| M0528 | Rheumatoid vasculitis | 0 |
| M0529 | Rheumatoid vasculitis | 0 |
| M053 | Rheumatoid arthritis with involvement of other organs and systems | 0 |
| M0530 | Rheumatoid arthritis with involvement of other organs and systems | 0 |
| M0531 | Rheumatoid arthritis with involvement of other organs and systems | 0 |
| M0532 | Rheumatoid arthritis with involvement of other organs and systems | 0 |
| M0533 | Rheumatoid arthritis with involvement of other organs and systems | 0 |
| M0534 | Rheumatoid arthritis with involvement of other organs and systems | 0 |
| M0535 | Rheumatoid arthritis with involvement of other organs and systems | 0 |
| M0536 | Rheumatoid arthritis with involvement of other organs and systems | 0 |
| M0537 | Rheumatoid arthritis with involvement of other organs and systems | 0 |
| M0538 | Rheumatoid arthritis with involvement of other organs and systems | 0 |
| M0539 | Rheumatoid arthritis with involvement of other organs and systems | 0 |
| M058 | Other seropositive rheumatoid arthritis | 0 |
| M0580 | Other seropositive rheumatoid arthritis | 0 |
| M0581 | Other seropositive rheumatoid arthritis | 0 |
| M0582 | Other seropositive rheumatoid arthritis | 0 |
| M0583 | Other seropositive rheumatoid arthritis | 0 |
| M0584 | Other seropositive rheumatoid arthritis | 0 |
| M0585 | Other seropositive rheumatoid arthritis | 0 |
| M0586 | Other seropositive rheumatoid arthritis | 0 |
| M0587 | Other seropositive rheumatoid arthritis | 0 |
| M0588 | Other seropositive rheumatoid arthritis | 0 |
| M0589 | Other seropositive rheumatoid arthritis | 0 |
| M059 | Seropositive rheumatoid arthritis, unspecified | 0 |
| M0590 | Seropositive rheumatoid arthritis, unspecified | 0 |
| M0591 | Seropositive rheumatoid arthritis, unspecified | 0 |
| M0592 | Seropositive rheumatoid arthritis, unspecified | 0 |
| M0593 | Seropositive rheumatoid arthritis, unspecified | 0 |
| M0594 | Seropositive rheumatoid arthritis, unspecified | 0 |
| M0595 | Seropositive rheumatoid arthritis, unspecified | 0 |
| M0596 | Seropositive rheumatoid arthritis, unspecified | 0 |
| M0597 | Seropositive rheumatoid arthritis, unspecified | 0 |
| M0598 | Seropositive rheumatoid arthritis, unspecified | 0 |
| M0599 | Seropositive rheumatoid arthritis, unspecified | 0 |
| M06 | Other rheumatoid arthritis | 0 |
| M060 | Seronegative rheumatoid arthritis | 0 |
| M0600 | Seronegative rheumatoid arthritis | 0 |
| M0601 | Seronegative rheumatoid arthritis | 0 |
| M0602 | Seronegative rheumatoid arthritis | 0 |
| M0603 | Seronegative rheumatoid arthritis | 0 |
| M0604 | Seronegative rheumatoid arthritis | 0 |
| M0605 | Seronegative rheumatoid arthritis | 0 |
| M0606 | Seronegative rheumatoid arthritis | 0 |
| M0607 | Seronegative rheumatoid arthritis | 0 |
| M0608 | Seronegative rheumatoid arthritis | 0 |
| M0609 | Seronegative rheumatoid arthritis | 0 |
| M062 | Rheumatoid bursitis | 0 |
| M0620 | Rheumatoid bursitis | 0 |
| M0621 | Rheumatoid bursitis | 0 |
| M0622 | Rheumatoid bursitis | 0 |
| M0623 | Rheumatoid bursitis | 0 |
| M0624 | Rheumatoid bursitis | 0 |
| M0625 | Rheumatoid bursitis | 0 |
| M0626 | Rheumatoid bursitis | 0 |
| M0627 | Rheumatoid bursitis | 0 |
| M0628 | Rheumatoid bursitis | 0 |
| M0629 | Rheumatoid bursitis | 0 |
| M063 | Rheumatoid nodule | 0 |
| M0630 | Rheumatoid nodule | 0 |
| M0631 | Rheumatoid nodule | 0 |
| M0632 | Rheumatoid nodule | 0 |
| M0633 | Rheumatoid nodule | 0 |
| M0634 | Rheumatoid nodule | 0 |
| M0635 | Rheumatoid nodule | 0 |
| M0636 | Rheumatoid nodule | 0 |
| M0637 | Rheumatoid nodule | 0 |
| M0638 | Rheumatoid nodule | 0 |
| M0639 | Rheumatoid nodule | 0 |
| M068 | Other specified rheumatoid arthritis | 0 |
| M0680 | Other specified rheumatoid arthritis | 0 |
| M0681 | Other specified rheumatoid arthritis | 0 |
| M0682 | Other specified rheumatoid arthritis | 0 |
| M0683 | Other specified rheumatoid arthritis | 0 |
| M0684 | Other specified rheumatoid arthritis | 0 |
| M0685 | Other specified rheumatoid arthritis | 0 |
| M0686 | Other specified rheumatoid arthritis | 0 |
| M0687 | Other specified rheumatoid arthritis | 0 |
| M0688 | Other specified rheumatoid arthritis | 0 |
| M0689 | Other specified rheumatoid arthritis | 0 |
| M069 | Rheumatoid arthritis, unspecified | 0 |
| M0690 | Rheumatoid arthritis, unspecified | 0 |
| M0691 | Rheumatoid arthritis, unspecified | 0 |
| M0692 | Rheumatoid arthritis, unspecified | 0 |
| M0693 | Rheumatoid arthritis, unspecified | 0 |
| M0694 | Rheumatoid arthritis, unspecified | 0 |
| M0695 | Rheumatoid arthritis, unspecified | 0 |
| M0696 | Rheumatoid arthritis, unspecified | 0 |
| M0697 | Rheumatoid arthritis, unspecified | 0 |
| M0698 | Rheumatoid arthritis, unspecified | 0 |
| M0699 | Rheumatoid arthritis, unspecified | 0 |

---

#### CONCEPT: Schizophrenia¶

| ICD10\_CODE | DESCRIPTION\_SAIL | SENSITIVE\_CODE |
| --- | --- | --- |
| F062 | Organic delusional [schizophrenia-like] disorder | 0 |
| F20 | Schizophrenia | 0 |
| F200 | Paranoid schizophrenia | 0 |
| F201 | Hebephrenic schizophrenia | 0 |
| F202 | Catatonic schizophrenia | 0 |
| F203 | Undifferentiated schizophrenia | 0 |
| F204 | Post-schizophrenic depression | 0 |
| F205 | Residual schizophrenia | 0 |
| F206 | Simple schizophrenia | 0 |
| F208 | Other schizophrenia | 0 |
| F209 | Schizophrenia, unspecified | 0 |
| F230 | Acute polymorphic psychotic disorder without symptoms of schizophrenia | 0 |
| F231 | Acute polymorphic psychotic disorder with symptoms of schizophrenia | 0 |
| F232 | Acute schizophrenia-like psychotic disorder | 0 |

---

#### CONCEPT: Skin Cancer¶

| ICD10\_CODE | DESCRIPTION\_SAIL | SENSITIVE\_CODE |
| --- | --- | --- |
| C43 | Malignant melanoma of skin | 0 |
| C430 | Malignant neoplasm: Malignant melanoma of lip | 0 |
| C431 | Malignant neoplasm: Malignant melanoma of eyelid, including canthus | 0 |
| C432 | Malignant neoplasm: Malignant melanoma of ear and external auricular canal | 0 |
| C433 | Malignant neoplasm: Malignant melanoma of other and unspecified parts of face | 0 |
| C434 | Malignant neoplasm: Malignant melanoma of scalp and neck | 0 |
| C435 | Malignant neoplasm: Malignant melanoma of trunk | 0 |
| C436 | Malignant neoplasm: Malignant melanoma of upper limb, including shoulder | 0 |
| C437 | Malignant neoplasm: Malignant melanoma of lower limb, including hip | 0 |
| C438 | Malignant neoplasm: Overlapping malignant melanoma of skin | 0 |
| C439 | Malignant neoplasm: Malignant melanoma of skin, unspecified | 0 |
| C44 | Other malignant neoplasms of skin | 0 |
| C440 | Malignant neoplasm: Skin of lip | 0 |
| C441 | Malignant neoplasm: Skin of eyelid, including canthus | 0 |
| C442 | Malignant neoplasm: Skin of ear and external auricular canal | 0 |
| C443 | Malignant neoplasm: Skin of other and unspecified parts of face | 0 |
| C444 | Malignant neoplasm: Skin of scalp and neck | 0 |
| C445 | Malignant neoplasm: Skin of trunk | 0 |
| C446 | Malignant neoplasm: Skin of upper limb, including shoulder | 0 |
| C447 | Malignant neoplasm: Skin of lower limb, including hip | 0 |
| C448 | Malignant neoplasm: Overlapping lesion of skin | 0 |
| C449 | Malignant neoplasm: Malignant neoplasm of skin, unspecified | 0 |
| C460 | Kaposi sarcoma of skin | 0 |
| C792 | Secondary malignant neoplasm of skin | 0 |
| C826 | Cutaneous follicle centre lymphoma | 0 |
| C848 | Cutaneous T-cell lymphoma, unspecified | 0 |
| D485 | Neoplasm of uncertain or unknown behaviour: Skin | 0 |

---

#### CONCEPT: Stroke¶

| ICD10\_CODE | DESCRIPTION\_SAIL | SENSITIVE\_CODE |
| --- | --- | --- |
| G463 | Brain stem stroke syndrome | 0 |
| G464 | Cerebellar stroke syndrome | 0 |
| I60 | Subarachnoid haemorrhage | 0 |
| I600 | Subarachnoid haemorrhage from carotid siphon and bifurcation | 0 |
| I601 | Subarachnoid haemorrhage from middle cerebral artery | 0 |
| I602 | Subarachnoid haemorrhage from anterior communicating artery | 0 |
| I603 | Subarachnoid haemorrhage from posterior communicating artery | 0 |
| I604 | Subarachnoid haemorrhage from basilar artery | 0 |
| I605 | Subarachnoid haemorrhage from vertebral artery | 0 |
| I606 | Subarachnoid haemorrhage from other intracranial arteries | 0 |
| I607 | Subarachnoid haemorrhage from intracranial artery, unspecified | 0 |
| I608 | Other subarachnoid haemorrhage | 0 |
| I609 | Subarachnoid haemorrhage, unspecified | 0 |
| I61 | Intracerebral haemorrhage | 0 |
| I610 | Intracerebral haemorrhage in hemisphere, subcortical | 0 |
| I611 | Intracerebral haemorrhage in hemisphere, cortical | 0 |
| I612 | Intracerebral haemorrhage in hemisphere, unspecified | 0 |
| I613 | Intracerebral haemorrhage in brain stem | 0 |
| I614 | Intracerebral haemorrhage in cerebellum | 0 |
| I615 | Intracerebral haemorrhage, intraventricular | 0 |
| I616 | Intracerebral haemorrhage, multiple localized | 0 |
| I618 | Other intracerebral haemorrhage | 0 |
| I619 | Intracerebral haemorrhage, unspecified | 0 |
| I64 | Stroke, not specified as haemorrhage or infarction | 0 |
| I672 | Cerebral atherosclerosis | 0 |
| I679 | Cerebrovascular disease, unspecified | 0 |
| I691 | Sequelae of intracerebral haemorrhage | 0 |

---

#### CONCEPT: Systemic Lupus Erythematosus¶

| ICD10\_CODE | DESCRIPTION\_SAIL | SENSITIVE\_CODE |
| --- | --- | --- |
| L93 | Lupus erythematosus | 0 |
| L931 | Subacute cutaneous lupus erythematosus | 0 |
| L932 | Other local lupus erythematosus | 0 |
| M32 | Systemic lupus erythematosus | 0 |
| M320 | Drug-induced systemic lupus erythematosus | 0 |
| M321 | Systemic lupus erythematosus with organ or system involvement | 0 |
| M328 | Other forms of systemic lupus erythematosus | 0 |
| M329 | Systemic lupus erythematosus, unspecified | 0 |

---

#### CONCEPT: Systemic sclerosis¶

| ICD10\_CODE | DESCRIPTION\_SAIL | SENSITIVE\_CODE |
| --- | --- | --- |
| L940 | Localized scleroderma [morphea] | 0 |
| L941 | Linear scleroderma | 0 |
| M34 | Systemic sclerosis | 0 |
| M340 | Progressive systemic sclerosis | 0 |
| M342 | Systemic sclerosis induced by drugs and chemicals | 0 |
| M348 | Other forms of systemic sclerosis | 0 |
| M349 | Systemic sclerosis, unspecified | 0 |

---

#### CONCEPT: Thalassaemia¶

| ICD10\_CODE | DESCRIPTION\_SAIL | SENSITIVE\_CODE |
| --- | --- | --- |
| D56 | Thalassaemia | 0 |
| D560 | Alpha thalassaemia | 0 |
| D561 | Beta thalassaemia | 0 |
| D562 | Delta-beta thalassaemia | 0 |
| D563 | Thalassaemia trait | 0 |
| D564 | Hereditary persistence of fetal haemoglobin [HPFH] | 0 |
| D568 | Other thalassaemias | 0 |
| D569 | Thalassaemia, unspecified | 0 |

---

#### CONCEPT: TIA (Transient ischaemic attack)¶

| ICD10\_CODE | DESCRIPTION\_SAIL | SENSITIVE\_CODE |
| --- | --- | --- |
| G45 | Transient cerebral ischaemic attacks and related syndromes | 0 |
| G453 | Amaurosis fugax | 0 |
| G454 | Transient global amnesia | 0 |
| G458 | Other transient cerebral ischaemic attacks and related syndromes | 0 |
| G459 | Transient cerebral ischaemic attack, unspecified | 0 |

---

#### CONCEPT: Viral Hepatitis¶

| ICD10\_CODE | DESCRIPTION\_SAIL | SENSITIVE\_CODE |
| --- | --- | --- |
| B16 | Acute hepatitis B | 1 |
| B160 | Acute hepatitis B with delta-agent (coinfection) with hepatic coma | 1 |
| B161 | Acute hepatitis B with delta-agent (coinfection) without hepatic coma | 1 |
| B162 | Acute hepatitis B without delta-agent with hepatic coma | 1 |
| B169 | Acute hepatitis B without delta-agent and without hepatic coma | 1 |
| B170 | Acute delta-(super)infection of hepatitis B carrier | 1 |
| B171 | Acute hepatitis C | 1 |
| B180 | Chronic viral hepatitis B with delta-agent | 1 |
| B181 | Chronic viral hepatitis B without delta-agent | 1 |
| B182 | Chronic viral hepatitis C | 1 |

---

#### CONCEPT: VTD (Venous thrombotic disease)¶

| ICD10\_CODE | DESCRIPTION\_SAIL | SENSITIVE\_CODE |
| --- | --- | --- |
| I802 | Phlebitis and thrombophlebitis of other deep vessels of lower extremities | 0 |
| I822 | Embolism and thrombosis of vena cava | 0 |
| I828 | Embolism and thrombosis of other specified veins | 0 |
| I870 | Postthrombotic syndrome | 0 |
| O223 | Deep phlebothrombosis in pregnancy | 0 |
| O225 | Cerebral venous thrombosis in pregnancy | 0 |
| O871 | Deep phlebothrombosis in the puerperium | 0 |
| O873 | Cerebral venous thrombosis in the puerperium | 0 |
